# THRESHER: A tool for evidence-based, phylogeny-corrected SNP thresholds for whole genome molecular epidemiology

**DOI:** 10.64898/2026.10.07.26364951

**Authors:** Qianxuan She, Andries Feder, Erin Theiller, Joseph P. Zackular, Ahmed M. Moustafa, Paul J. Planet

## Abstract

Whole-genome sequencing (WGS) is the gold standard for bacterial pathogen surveillance, where transmission is commonly inferred by calculating pairwise single nucleotide polymorphisms (SNPs) and applying a fixed threshold below which two genomes are considered the “same” strain. Such cutoffs are largely arbitrary. The relationship between genetic distance and epidemiological linkage varies with sample size, sampling duration, setting, and patterns of infection and carriage. A threshold calibrated in one study may not transfer to another. To address this, we present THRESHER, a pipeline that infers data-driven phylothresholds directly from each dataset’s genomic structure. THRESHER implements four endpoint methods, *plateau, public, peak*, and *discrepancy*, with each reflecting a distinct hypothesis about what constitutes the “sameness” of a strain. Across 55 published datasets spanning *Staphylococcus aureus, Clostridioides difficile*, and *Klebsiella pneumoniae*, THRESHER reproduced the originally reported strain compositions in smaller studies. In larger studies, where the two diverged, SNP distances and phylogenetic topology supported THRESHER’s assignments. In simulated populations with known strain structure, THRESHER recovered transmission clusters with substantially higher sensitivity than fixed cutoffs while retaining high specificity.

## INTRODUCTION

Whole-genome sequencing (WGS) has become the gold standard for bacterial pathogen surveillance and outbreak investigation, where it is widely used to track pathogen dissemination and to infer transmission^1^. Putative transmission is anchored on defining whether strains are the same and different. Commonly, strain “sameness” is determined by calculation of pairwise single-nucleotide polymorphisms (SNPs) and applying a fixed threshold below which genomes are considered linked^2-4^.

Several challenges remain unresolved for this approach. Foremost, a fixed threshold is largely arbitrary. It imposes a single value on a relationship that is not itself fixed because genomic sequences diversify and change over time. What constitutes the same strain is context-dependent, relying critically on the question being asked and on the structure of the data at hand. The primary goal of WGS approaches is to resolve isolates into informative units for infection prevention and control (IP&C) teams to target and reduce local transmission^4-7^. The correspondence between genetic distance and epidemiological linkage also varies with sample size, sampling duration, setting, and the underlying patterns of infection and carriage, so a threshold calibrated in one study may not transfer to another^8-10^. Thus, single fixed cutoffs may over- or under-split strain compositions across studies, which limits both the accuracy and the comparability of strain and transmission inference.

Another challenge emerges as datasets grow. SNP distances are conventionally computed by aligning every genome to a single reference, and a single phylogeny is used to represent the genomic relationships among them^11-14^. For small, closely related collections this approach is adequate, but as the number and diversity of genomes increase, no single reference aligns well to all of them and no single tree can provide sufficient resolution. The choice of reference then biases the resulting distances and topology, so that the results to which a fixed cutoff is applied become less reliable in large datasets.

Here we present THRESHER, an end-to-end command line pipeline that implements a framework to replace fixed SNP cutoffs with data-driven phylothresholds: SNP thresholds corrected for phylogenetic topology and inferred directly from the genomic structure of each dataset. THRESHER implements four endpoint methods (*plateau, public, peak*, and *discrepancy*), each reflecting a distinct hypothesis about what constitutes the same strain. THRESHER identifies the phylothreshold that best captures the underlying phylogenetic structure rather than imposing a predefined value. To remain robust as dataset size grows, THRESHER partitions genomes into groups by silhouette-score-based hierarchical clustering when necessary, selecting an optimal reference for each group to ensure high-quality alignments and reliable phylogenies, and computes genetic distances through comprehensive all-versus-all pairwise comparison so that each distance is reference-free. Because thresholds are derived for each dataset, the approach adapts across species, settings, and sampling regimes without manual recalibration. Given the complexity of the framework and the range of its potential users, we provide a locally deployable chatbot that offers interactive usage guidance across levels of expertise.

To validate THRESHER, we simulated *S. aureus* outbreaks in which the true transmission cluster composition is known and defined at a granularity matching the informative unit described above. Comparing three of THRESHER’s endpoint methods with fixed cutoffs, we found that THRESHER outperformed fixed cutoffs across the sensitivity–specificity trade-off landscape. We then tested THRESHER on 55 published datasets spanning *Staphylococcus aureus, Clostridioides difficile*, and *Klebsiella pneumoniae*, comparing THRESHER-inferred strain compositions with those reported in the original studies. Concordance was high for smaller datasets, where a single phylogeny adequately represents the underlying genomic structure. For larger datasets, THRESHER recovered strain structure that fixed cutoffs had missed, and assessment using both phylogeny and SNP distances provided strong evidence that THRESHER yields more accurate strain compositions than the original fixed-threshold assignments.

## RESULTS

### Published WGS studies vary considerably in SNP cutoffs, data structure, diversity, and timeframe

To characterize how cutoffs have been chosen in the published literature, we assembled 55 published studies (*S. aureus*, n = 22; *K. pneumoniae*, n = 16; *C. difficile*, n = 17, Supplementary Table 1) with publicly available metadata together with sequencing reads or genome assemblies, summarizing each by predominant genomic background, sample size, sampling duration, and applied SNP cutoff (Figure 1a–c. *S. aureus*, n = 22; *C. difficile*, n = 17; *K. pneumoniae*, n = 16). These studies span a range of geographic locations and settings and encompass diverse genomic backgrounds, described by clonal complex (*S. aureus*), sequence type (*C. difficile*), and sequence type or clonal group (*K. pneumoniae*). In a subset of smaller studies, a single phylogeny clearly delineated monophyletic strains allowing for manual demarcation directly from tree topology and subsequent calculation of a range of within-strain SNP distances (S6, S18, S32, S38, S50, S52, C7, C13, C15, C17, C18, K3, K12, K18). In many larger studies, however, phylogenetic complexity and scale precluded manual delineation, and strains were instead assigned using a fixed cutoff applied across diverse lineages and populations. Across the full collection of studies, we found no consistent relationship between the chosen cutoff and the characteristics that should affect it, despite pronounced differences in sample size, sampling duration, and genetic background. This pattern suggests that many researchers adopt cutoffs used in earlier studies, even though those values might not fit the genomic structure of their own datasets. These observations motivated an approach that infers the threshold from the genomic structure of each dataset rather than basing it on previously published and unrelated datasets, and this is the principle underlying THRESHER.

**Figure 1.**
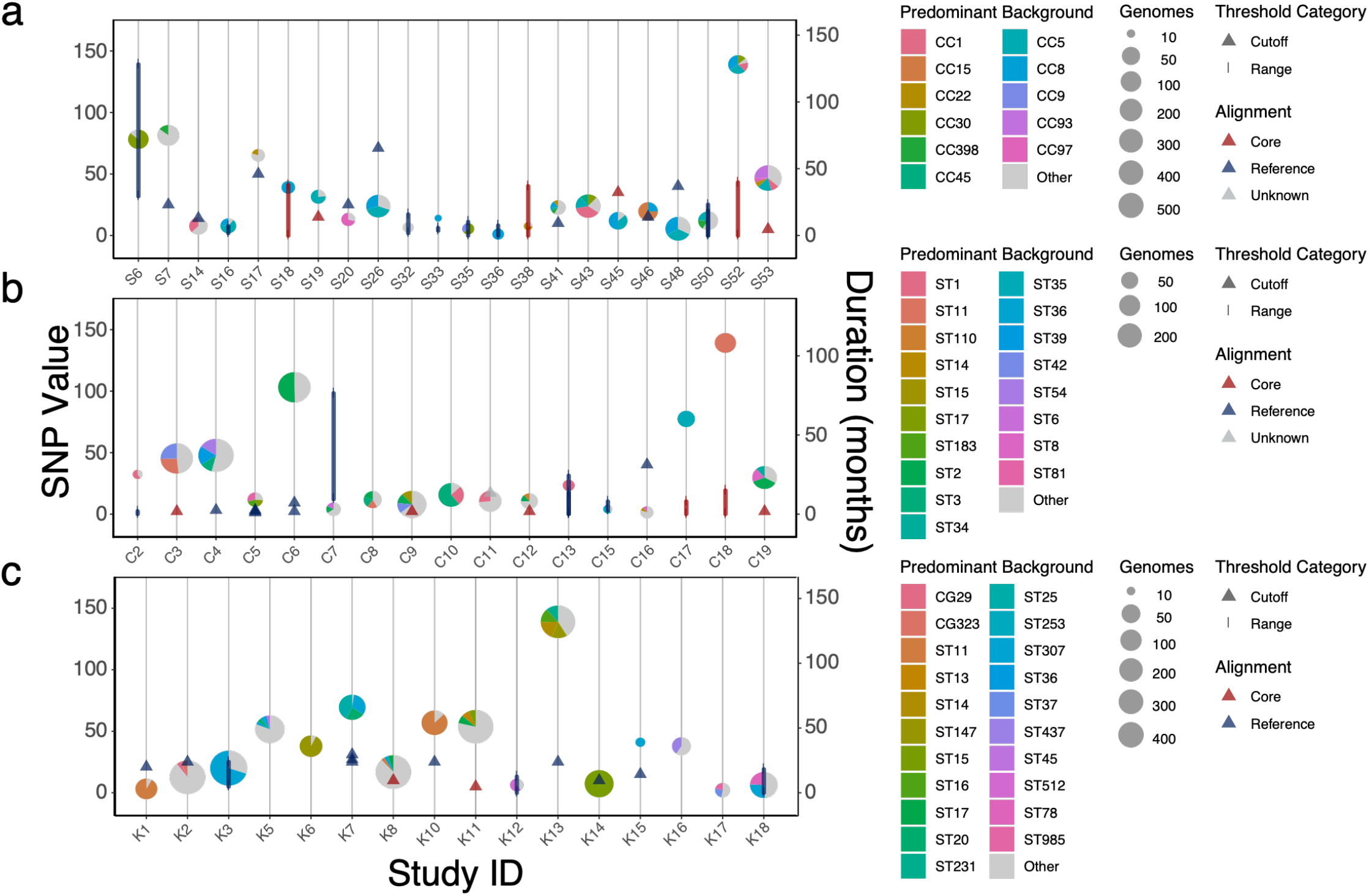
SNP thresholds, sampling characteristics, and genomic backgrounds across published WGS studies of three bacterial pathogens. Each panel presents the studies for one species: (a) *S. aureus* (n = 22), (b) *C. difficile* (n = 17), and (c) *K. pneumoniae* (n = 16), with every study occupying one position along the x-axis (Study ID). Two quantities share the vertical space and are read against different axes: the applied SNP threshold against the left axis (SNP Value) and sampling duration against the right axis (Duration, months). For each study, the SNP threshold is drawn as a marker placed at its value on the left axis and colored by the alignment strategy used for variant calling (Alignment: Reference, reference-based; Core, core-genome; Unknown, not reported). A single reported cutoff appears as a triangle and a reported range as a vertical line spanning the interval, and studies reporting several discrete cutoffs show multiple triangles (Threshold Category). The pie at each study is centered vertically at that study’s sampling duration on the right axis and conveys two further attributes: its size increases with the number of genomes (Genomes), and its slices give the proportional composition of the predominant genomic backgrounds. Backgrounds are summarized as clonal complexes (*S. aureus*), sequence types (*C. difficile*), and sequence types or clonal groups (*K. pneumoniae*). Minor backgrounds are pooled as “Other” and unassigned isolates as “Unknown.” Because the predominant backgrounds are species-specific, the Predominant Background key is shown separately for each panel.

### THRESHER: inferring phylothresholds from the genomic structure of each dataset

THRESHER infers SNP thresholds directly from the genomic diversity of each dataset and corrects them for phylogenetic topology. We refer to such a threshold as a *phylothreshold*. The overall workflow is summarized in Figure 2, with each endpoint’s algorithm and graphical criterion elaborated in Figure 3. THRESHER takes two inputs: genome assemblies of the study isolates and a metadata table recording, for each isolate, its name, file path, GenBank accession (if applicable), and optionally patient identifier and collection date for downstream epidemiological analysis. Users additionally specify the species under study. *S. aureus, K. pneumoniae, C. difficile*, and *S. epidermidis* are currently supported, but infrastructure is readily available to expand to any bacterial species.

**Figure 2.**
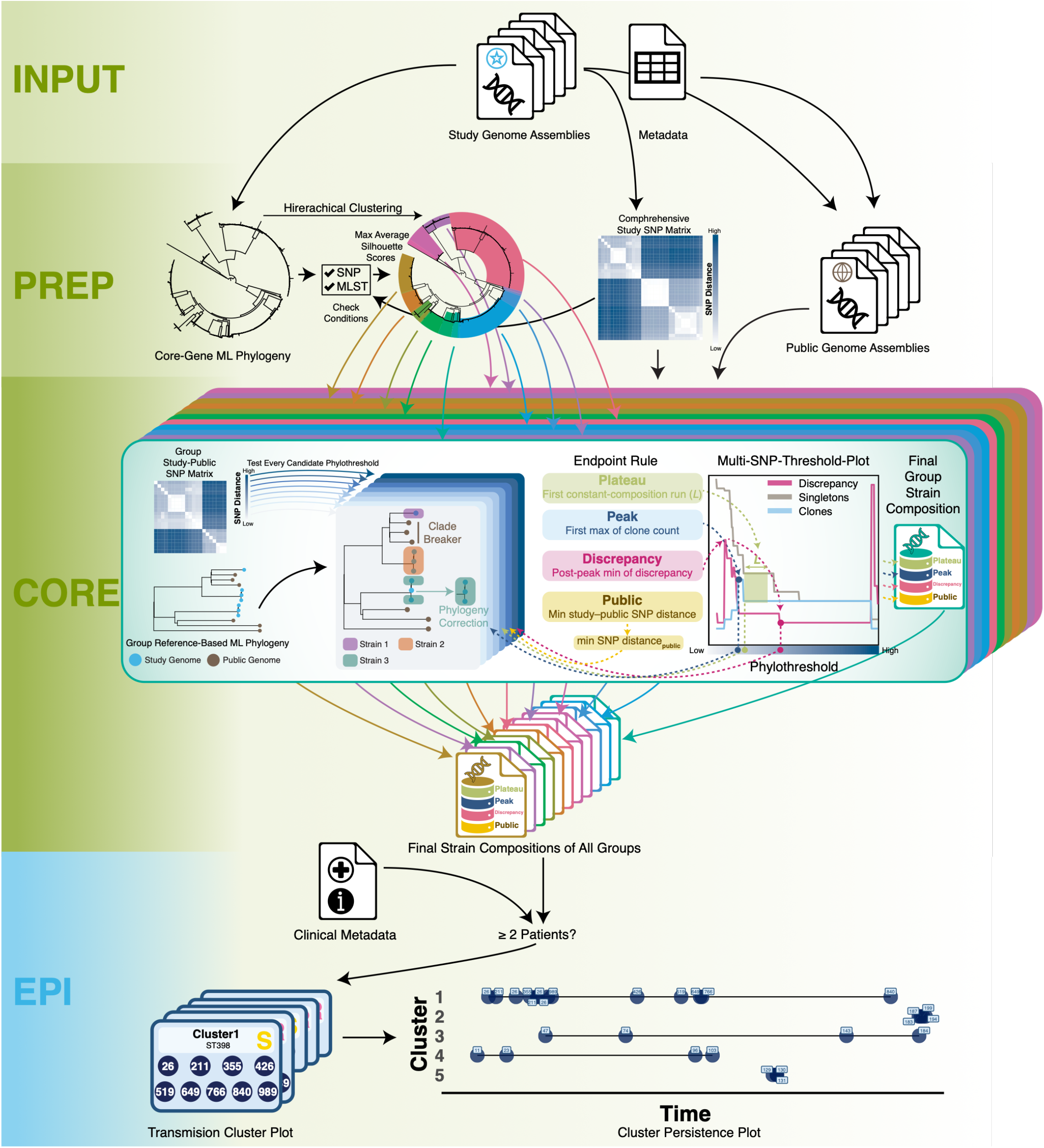
Overview of the overall THRESHER workflow. THRESHER proceeds through four stages INPUT, PREP, CORE, and the optional EPI shown in colored sections, from top to bottom. **INPUT**. THRESHER takes the genome assemblies of the study isolates together with a metadata table recording isolate names, file paths, GenBank accessions (where applicable), and optional patient identifiers and collection dates. **PREP**. Three analyses run in parallel. A core-gene alignment yields a core-gene maximum-likelihood phylogeny representing the overall genomic diversity of the dataset (left). A comprehensive all-versus-all SNP distance matrix is computed (center). Each study genome is matched against a curated database of public genomes to retrieve its closest publicly available match (right). From the phylogeny, the SNP distances, and the multi-locus sequence typing (MLST) scheme, THRESHER determines whether the dataset can be analyzed as a whole or should be partitioned into several groups. When partitioning is warranted, genomes are divided by hierarchical clustering at the partition maximizing the average silhouette score (circular tree, colored by group). **CORE**. The core algorithm is applied independently to each hierarchical clustering group (stacked cards). Within a group, a reference-based ML phylogeny is inferred from the study genomes together with their closest public genomes (blue: study, grey: public), and a Group Study-Public SNP submatrix is assembled from the corresponding block of the comprehensive study SNP matrix together with the study-public SNP distances. Distances between study genomes and unrelated public genomes, and all public–public pairs, are neither required nor computed, leaving the study–public block sparse. Strain composition is then evaluated at every candidate phylothreshold across the user-configurable distance range, where single-linkage clustering on SNP distances is mapped onto the phylogeny and corrected to restore monophyly (Phylogeny Correction), with CladeBreaker optionally preventing strains from harboring public genomes as the “same strain” (Clade Breaker). The compositions obtained across all candidate phylothresholds are summarized and visualized in the Multi-SNP-Threshold Plot, which tracks the number of clones and singletons and the discrepancy metric as the phylothreshold increases. Each endpoint method then selects the group’s final phylothreshold by its own rule (*Plateau*, the first constant-composition run of length L; *Peak*, the first maximum of the clone count; *Discrepancy*, the post-peak minimum of the discrepancy; *Public*, the smallest study-to-public SNP distance), and the strain composition at the corresponding phylothreshold is retrieved as the group’s final composition. Because each rule responds to the population structure of the particular dataset, the endpoints are ordered consistently across different datasets. For instance, the *peak* endpoint is not always the most conserved. The schematic illustrates the rules rather than fixed ordering. Strain compositions from all groups are then combined. **EPI**. When patient identifiers and collection dates are provided, an optional epidemiological step identifies transmission clusters (clones detected in more than one patient) and then visualizes their membership (Transmission Cluster Plot) and persistence over time (Cluster Persistence Plot).

**Figure 3.**
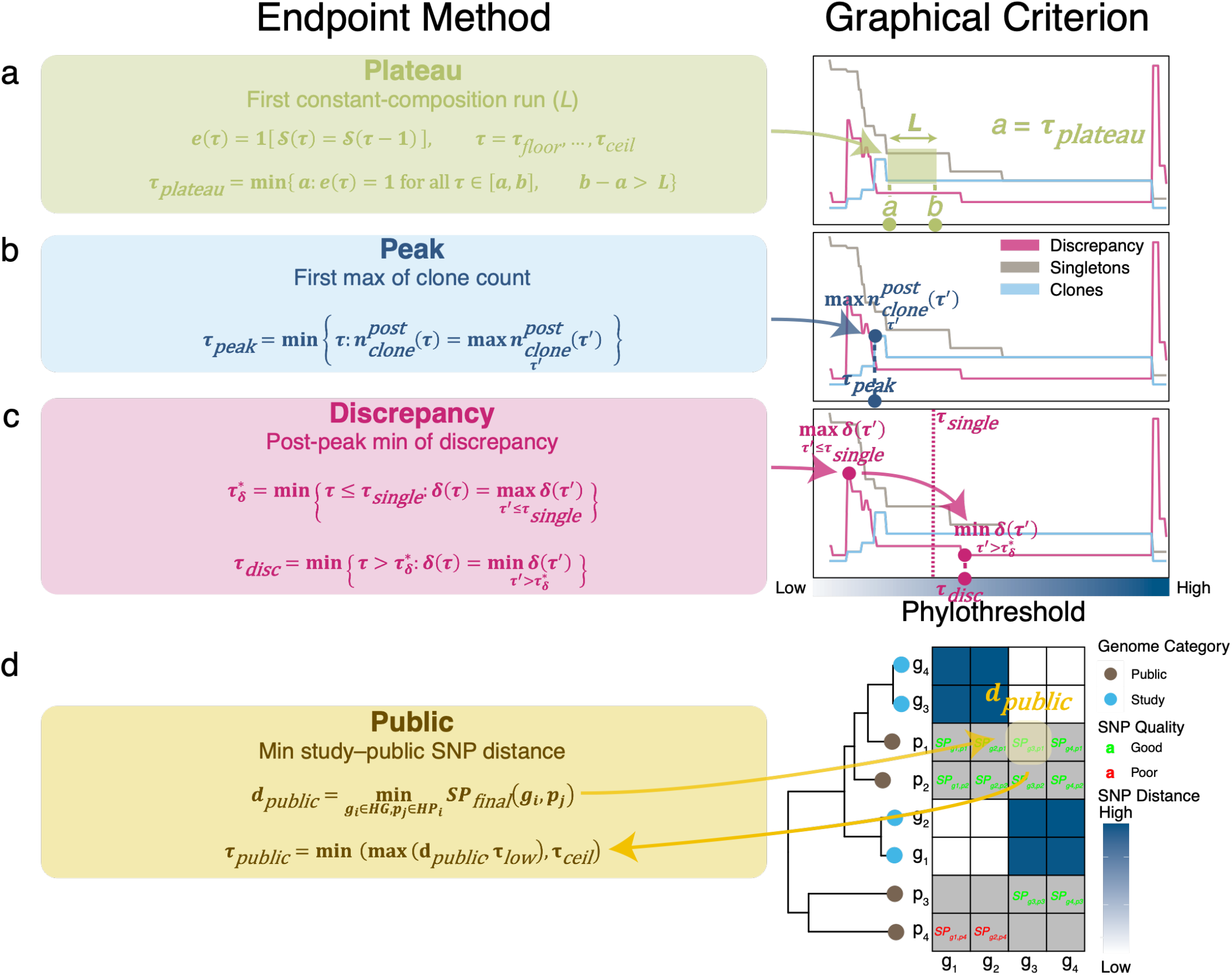
Definition and graphical criterion for each endpoint method. The four endpoint methods introduced in Figure 2 (CORE) are expanded here: (a) *Plateau*, (b) *Peak*, (c) *Discrepancy*, and (d) *Public*. In each panel, the left panel (Endpoint Method) provides the formal definition, and the right panel (Graphical Criterion) illustrates where the resulting phylothreshold locates. For *Plateau, Peak*, and *Discrepancy* the criterion can be visualized on the Multi-SNP-Threshold Plot (MSTP), whose x-axis shows the phylothreshold candidates (increasing left to right) and whose three curves track the clone count (light blue), singleton count (grey), and discrepancy metric (red). The final phylothreshold using Public method is instead read from the group’s phylogeny and its study–public SNP submatrix. τ_*floor*_ and τ_*ceil*_ denote the lower and upper bounds of the tested phylothreshold candidates. **(a)*Plateau*** S(τ) is the strain composition at phylothreshold τ and e(τ) = 1 indicates strain composition is unchanged from the previous tested phylothreshold τ− 1. τ_plateau_ is the start (position *a*) of the first run of constant composition spanning more than *L* consecutive values (default *L* = 15; shaded a to b on the MSTP). **(b)*Peak*** 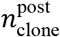 (τ) is the clone count after phylogeny correction and τ_peak_ is the first phylothreshold at which it reaches its maximum. **(c)*Discrepancy*** δ(τ) is the discrepancy metric before phylogeny correction at phylothreshold candidate τ. 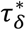 is the first phylothreshold at which δ reaches its maximum within the singleton threshold τ_*single*_, and τ_disc_ is the first phylothreshold beyond 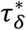 at which δ reaches its minimum. If every δ beyond singleton threshold τ_*single*_ exceeds δ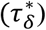, the search instead returns the phylothreshold candidate generating the smallest δ, whether it precedes 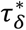. **(d)*Public*** *SP*_final_(*g*_*i*_, *p*_*j*_) is the SNP distance between study genome *g*_*i*_ and public genome *p*_*j*_. *HG* and *HP*_*i*_ are the group’s study and public genome sets, and *d*_public_ is the smallest good-quality *SP*_final_ over all pairs. The final phylothreshold τ_public_ equals the value of *d*_public_ within the interval [τ_low_, τ_ceil_]. Phylogeny tips are colored by genome category (study, blue; public, brown), with study and public tips labeled g and p. Only study–study cells are shaded by SNP distance (dark, high; light, low). Study–public cells are grey. Computed pairs are annotated with *SP*_final_, and the label *SP*_final_ is colored by SNP quality (green, good; red, poor). Pairs neither required nor computed (Figure 2, Core) are left blank. Poor-quality study–public pairs are excluded when determining *d*_public_. The positions shown are illustrative because each rule responds to a dataset’s population structure, the endpoints need not fall in the order drawn here.

Once the input passes validation, three preparatory analyses run in parallel. First, a comprehensive all-versus-all SNP distance matrix is computed, in which every distance is reference-free and thus captures the genomic difference between two genomes without dependence on a common reference. Second, a core-gene alignment is derived from pan-genome analysis and used to infer a core-gene phylogeny representing the overall genomic diversity of the dataset. Third, each study genome is compared against a curated database of conspecific GenBank genomes to identify its closest publicly available matches, which are required for the later CladeBreaker step. Because a single threshold is unlikely to be appropriate across divergent genetic backgrounds THRESHER then uses the core-gene phylogeny, the SNP distances, and the MLST scheme to assess population structure and determine whether the dataset is homogeneous enough for one threshold to suffice or should instead be partitioned for multiple independent analyses. When partitioning is warranted, partitions ranging from 2 to *n*−1 groups (*n* = number of study genomes) are evaluated by silhouette-score-based^15^ hierarchical clustering, and the cluster count that maximizes with the mean silhouette width is retained, ensuring high-quality reference-based alignments and reliable phylogenies within each group (see Methods 1.2.3, Population-structure assessment and Hierarchical Clustering).

The core algorithm is then applied to each group. Genomes separated from all others by more than a customizable singleton threshold (default 100 SNPs, Method 1.3.1) are set aside as singletons. This filter removes genomes on very long branches, which would otherwise be integrated into a strain by the monophyly correction and produce misleading strain compositions. Once excluded, these singletons are never added back during the subsequent phylothreshold search. If genomes remain, a group-specific phylogeny is inferred from the study genomes together with their closest public genomes and used for phylogeny correction. For every threshold between a customizable floor (default 5 SNPs) and ceiling (default 500 SNPs), strain composition is first determined by single-linkage clustering on SNP distances alone and then mapped onto the group phylogeny. Any strain that is not monophyletic is corrected by adding the genomes inside the clade required to restore monophyly. The singleton threshold and the phylothreshold search serve different purposes. The phylothreshold is the value in the search range that satisfies an endpoint’s definition of a strain (Method 1.3.6). A phylothreshold can exceed the singleton threshold, but this reflects links accumulating among the remaining genomes but not the excluded singletons.

Users may enable the CladeBreaker setting, which is appropriate when transmission is assumed to be hyperlocal, such that study genomes should not be grouped with publicly deposited genomes of the same species. CladeBreaker constrains the phylogenetic correction using the matched public genomes: any clade whose most recent common ancestor (MRCA) contains a public genome is split, until the largest MRCA clades encompass all study genomes while excluding every public genome. (GenBank accessions allow THRESHER to recognize study isolates already deposited in NCBI).

For each group, strain compositions across all tested phylothresholds are stored in the intermediate database and visualized in the Multi-SNP-Threshold Plot (MSTP). An MSTP for each group of all 55 tested studies is provided in the Supplementary Data 1 at /thresher/output/MSTP within each study’s THRESHER output directory. From this database, each “endpoint” method selects the group’s final phylothreshold. Four endpoints are currently available, each encoding a different hypothesis about what defines a strain. The *plateau* method assumes that a natural gap in divergence, finer than species-level structure, separates strains, as observed across published datasets (see plots/SNP_Distance.pdf within each study’s output directory in the Supplementary Data1), and in our prior work^16^. The *plateau* method sets the phylothreshold at the start of the first contiguous window of tested phylothresholds (default length 15) within which the number and composition of strains (singletons and clones; a clone being a strain of at least two genomes) remain unchanged (Figure 3a, *plateau*). A composition that persists across such a window indicates that these phylothresholds all fall within the divergence gap, so the resulting strains are internally cohesive yet mutually separated, with the user-customizable window length setting how wide a gap the method requires. The *peak* method sets the phylothreshold at the maximum number of clones across all tested thresholds (Figure 3b, *peak*). This is the finest resolution at which the largest number of distinct clones are simultaneously recovered. The *discrepancy* method (Figure 3c, *discrepancy*) places the phylothreshold at the post-peak minimum of the discrepancy metric, a measurement of strain monophyly before phylogeny correction (Method 1.3.3). This metric penalizes strains whose SNP-only composition is not monophyletic on the phylogeny. Minimal discrepancy identifies strains that are simultaneously closely related in SNP distance and consistent with the tree topology. The *public* method sets the phylothreshold at the smallest good-quality SNP distance between a study genome in the group and a public genome (Figure 3d, *public*). This approach grounds the strain boundary in external evidence, using the closest publicly available genomes to calibrate a data-driven phylothreshold rather than imposing a fixed cutoff. In our simulations, these four endpoints separated along the sensitivity–specificity spectrum, with the *peak* endpoint toward the specificity end and *discrepancy* toward the sensitivity end, while the *plateau* method offers an intermediate, tunable option whose window length adjusts the balance between the two. We characterize these behaviors systematically below using a simulated outbreak with known transmission cluster structure.

Finally, the strain composition at each group’s selected phylothreshold is retrieved and merged across groups into a single genome-wide strain composition. If patient study numbers and collection dates are provided, an optional epidemiological step identifies transmission clusters (clones detected in more than one patient) and quantifies their persistence over time.

### Simulated outbreaks with known transmission clusters

To rigorously validate THRESHER with known phylogenetic scenarios, we sought a highly realistic simulation approach that closely aligned with the specific purpose and targets of molecular epidemiology. THRESHER is designed to capture hyperlocal transmission circulating within a clinical or community setting over a short window of a few years, rather than global dissemination at long time scales. Existing approaches for simulating bacterial evolution over short timescales rely largely on parametric assumptions that can introduce bias and oversimplification, and few exploit the thousands of genomes now banked in public databases to ground their models^17-19^. We generated our validation populations with SILOSim, an integrated framework that learns directly from real, site-by-site genomic variation^20^. From a single user-supplied genome assembly, SILOSim by default assembles a panel of closely related public genomes, aligns them to the that assembly, and derives a per-base map of mutational entropy together with the mobile genetic elements detected across the genome. Users may also supply their own panel of query genomes, in which case the public-database retrieval is skipped. It then simulates forward along a user-defined, time-scaled phylogeny specified in years, modeling substitution weighted by per-site entropy, within-sample and external homologous recombination, and gene gain and loss drawn from the empirically detected genetic mobile element pool, with all rates and parameters under user control. SILOSim outputs whole-genome sequences for every descendant in the population, matching THRESHER’s input. The profiling and simulation are described in full in the Methods Section^20^.

Although SILOSim is validated across species, we used *S. aureus* as the model organism here. We deduplicated 68,299 high-quality publicly available *S. aureus* genomes collected up until April 2024, at a 99.5% average nucleotide identity threshold within each sequence type, yielding 1,830 representative genomes. From these, five were selected as ancestral seeds, chosen to span the phylogeny, and each also represented a large number of genomes, providing the depth needed for accurate entropy and mobile-element inference (Methods 2.1, PROFILING panel, Figure 4 and Supplementary Figure 3). We then simulated evolution along three time-scaled phylogenies (44 genomes each including internal nodes and tips, spanning 5 years; speciation and extinction rates λ = 1 and μ = 100; SIMULATION panel, Figure 4 and Supplementary Figure 1). The same three phylogenies were applied to every seed, holding the evolutionary trajectory constant while allowing only the seed-specific entropy and mobile-element pools to differ, for a total of 660 simulated genomes (132 per seed).

**Figure 4.**
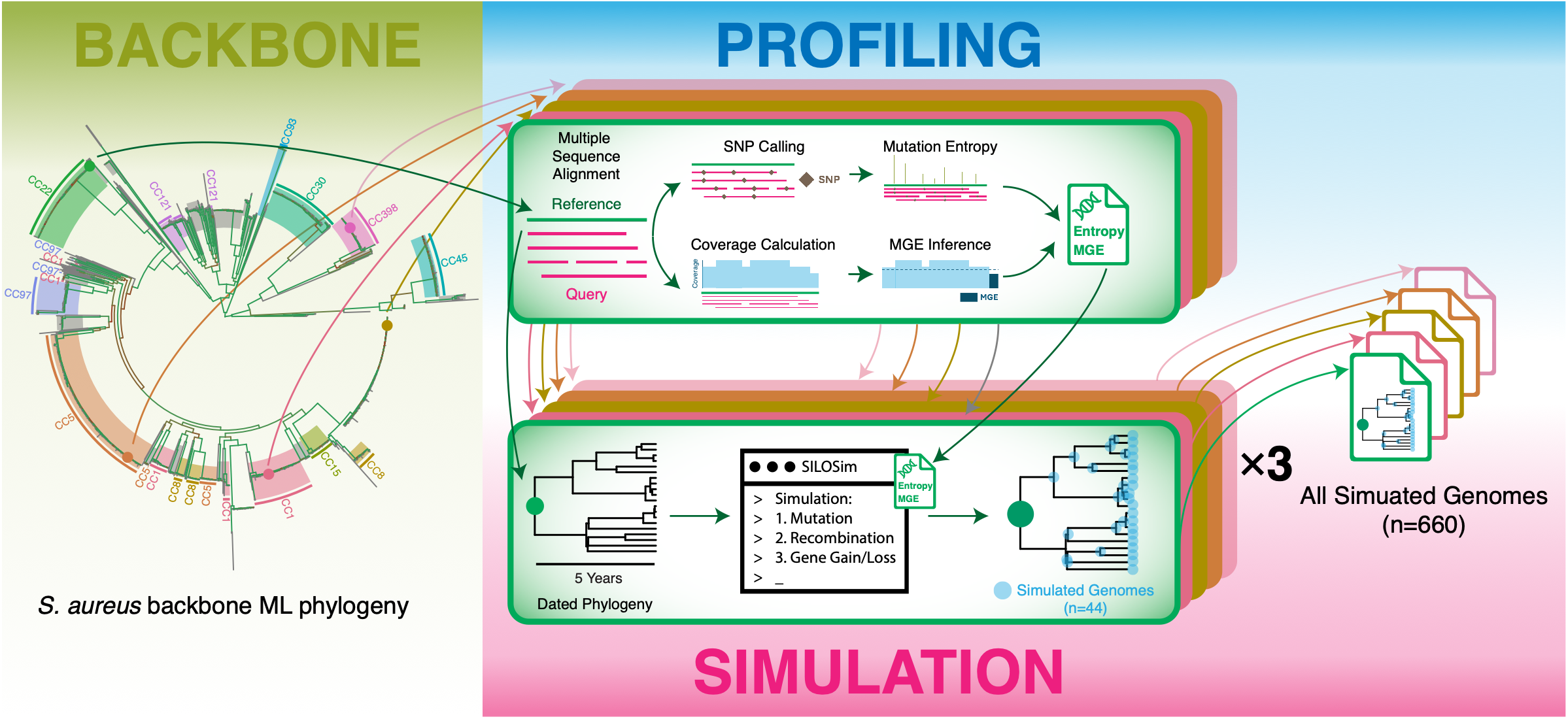
Generation of simulated *S. aureus* outbreak populations with SILOSim algorithm. The procedure comprises three stages: *S. aureus* backbone ML phylogeny and placement of ancestral seed genomes, profiling of each ancestral seed genome, and time-forward simulation (colored bands). **BACKBONE**. A core-gene maximum-likelihood phylogeny of 1,830 representative *S. aureus* genomes, obtained by deduplicating 68,299 publicly available genomes at 99.5% average nucleotide identity within each sequence type, summarizes the genomic diversity of the species. Major clonal complexes are highlighted. The five representatives subsuming the largest numbers of genomes (colored points) were selected as ancestral seeds for simulation. **PROFILING**. For each ancestral seed, a panel of closely related public genomes is aligned to the seed (Reference and Query). From this alignment, SILOSim calls SNPs and estimates per-site mutational entropy across the genome (Mutation Entropy), and from reductions in alignment coverage it infers mobile genetic elements (MGE Inference). The result is a per-genome profile of mutational entropy and detected MGEs. **SIMULATION**. Beginning from a time-scaled phylogeny spanning 5 years (Dated Phylogeny), the evolution simulator generates sequences along the tree under the seed’s entropy and MGE profile, modeling substitution, recombination, and gene gain and loss. Each simulation yields whole-genome sequences for all 44 tips and internal nodes of the tree (Simulated Genomes, n = 44). The same three time-scale phylogenies were applied to each of the five seeds, holding the evolutionary trajectory constant while allowing only the seed-specific entropy and MGE content to vary, for 660 simulated genomes in total.

Because our interest lies in defining local transmission over a relatively short interval, the informative units in the phylogeny are sub-lineages that would be inferred to be diverging very recently. Therefore, we treat each of these recent clades as independent transmission clusters for the purpose of validation (Supplementary Figure 1). To achieve this, we placed a boundary at year 3 of each 5-year time-scale phylogeny and identified the most ancestral internal nodes after this boundary. Each such node defines the most recent common ancestor of a distinct transmission cluster, comprising that node and all of its descendants, while genomes whose most recent coalescence predates Year 3 are designated singletons (Supplementary Figure 1). Internal nodes before Year 3 define no transmission cluster of interest and do not enter the calculation of sensitivity or specificity.

### In simulated outbreaks, THRESHER outperforms fixed arbitrary cutoffs across the sensitivity–specificity trade-off

After simulation, we compared the THRESHER approach against the a priori, fixed threshold (FIXED) approach. THRESHER assigns genomes to strains in two steps: single-linkage clustering on SNP distances followed by correction using the phylogeny. The accuracy depends on how both the SNP distances and the phylogeny represent the true diversity of the population. Because surveillance sampling is never complete and inevitably misses isolates, we built incomplete sampling directly into the validation: from the 660 simulated genomes we drew random subsamples at 20%, 40%, 60%, 80%, and 100% (ideal complete sampling), with 10 replicates per rate, for 50 subsampled populations in total.

THRESHER partitions genomes from divergent backgrounds before the CORE step (CORE panel in Figure 2), so genomes from different *S. aureus* clonal complexes can be assigned to separate hierarchical groups, which is consistent with the placement of the five seed genomes across the backbone phylogeny (BACKBONE panel, Figure 4). The FIXED approach also depends on a reference that covers the query genomes well. To compare the two methods under identical partitions, we split each subsample by sequence type of its ancestral seed. Sequence type was not re-typed from the simulated sequences but inherited from the seed, so grouping by sequence type is equivalent to grouping by seed of origin. Genomes sharing a seed sequence but originating from different time-scaled phylogenies were also pooled, yielding 250 per-ST groups of varying size in total (5 sequence types × 5 sampling rates × 10 replicates), so that both THRESHER and FIXED approaches were compared under identical, well-referenced partitions and any difference reflected the threshold method itself rather than the partitioning.

Published studies overwhelmingly apply cutoffs between 0 and 50 SNPs (Figure 1), so we evaluated fixed cutoffs of 0, 10, 20, 30, 40, and 50. For THRESHER we tested the *plateau, peak*, and *discrepancy* endpoints; the public endpoint does not apply over these short timescales, where every simulated genome’s closest match is its own ancestral seed. For *plateau* we used window lengths of 5 and 15 (the parameter governing the assumed width of the natural gap separating strains), giving four THRESHER configurations in total. For each of the 250 groups we visualized assignments by all methods alongside the simulated ground truth for defining true negatives, true positives, false negatives, and false positives with a matrix of method-specific cluster identifiers, combining the group phylogeny, and a SNP-distance heatmap (Supplementary Data 2).

Comparing each method against the simulated ground truth revealed several consistent patterns (Figure 5). FIXED cutoffs maintained near-perfect specificity across all six values and all sampling rates (n = 1,500; mean 99.86%; median 100%; IQR 99.87–100%), but sensitivity collapsed at conservative cutoffs: it reached at most 2.24% at a cutoff of 0 and remained below 20% at a cutoff of 10 (mean 14.39%; median 14.84%; IQR 12.95–16.05%). A cutoff of 0 can only link genomes between which no simulated substitution occurred, and across the 217,470 unique pairwise comparisons among the 660 simulated genomes only 22 (0.01%) were identical, all separated by less than two months of simulated time. This outcome challenges the common assumption that conservative cutoffs are the more “rigorous” choice; while they rarely raise false alarms, they fragment true transmission clusters, underestimating their size while overestimating their number, and thus risk misinforming clinical decisions (Figure 5).

**Figure 5.**
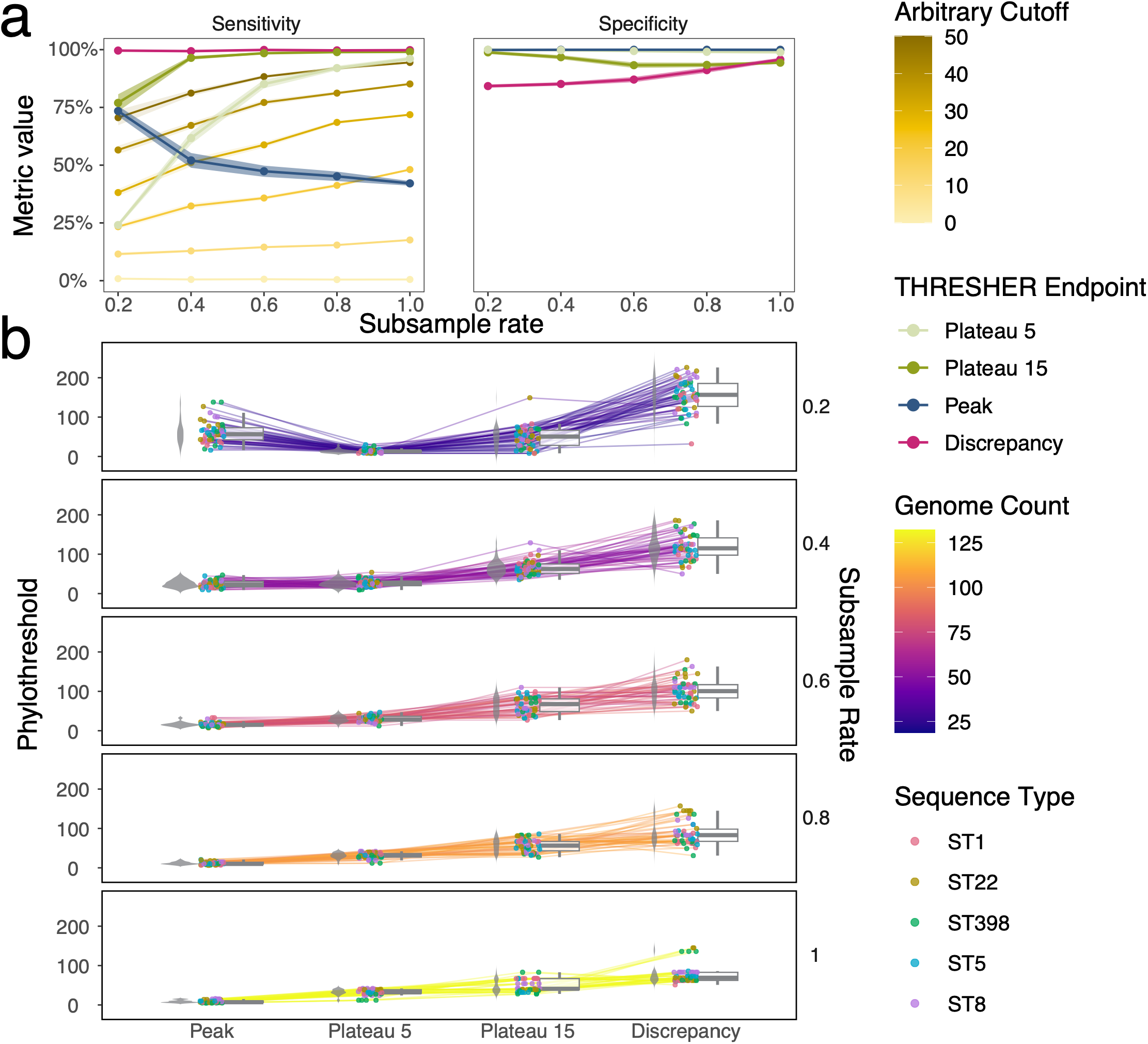
THRESHER endpoints versus arbitrary cutoffs across subsampling rates in simulated outbreaks. Both panels summarize the 250 per-ST groups (five seed sequence types × five subsampling rates × ten replicates) compared against the simulated ground truth. **(a)**Sensitivity (left) and specificity (right) across subsample rates, comparing THRESHER’s four endpoints (discrete colors; *Plateau* 5, *Plateau* 15, *Peak, Discrepancy*) with fixed arbitrary cutoffs of 0–50 SNPs (gold gradient, light to dark with increasing cutoff). Lines and points denote the mean across replicates and seeds at each rate. Shaded bands give the mean ± standard error. Conservative fixed cutoffs combine near-perfect specificity with very low sensitivity, and even permissive cutoffs remain strongly sensitivity-limited at low sampling, while THRESHER’s endpoints separate along the sensitivity–specificity trade-off, with *discrepancy* favoring sensitivity and *peak* favoring specificity. **(b)**Final phylothreshold selected by each endpoint, with one vertical panel per subsample rate (0.2–1.0, right labels). Within each panel, the four endpoints (*Peak, Plateau* 5, *Plateau* 15, *Discrepancy*) are shown as grey violin plots with overlaid boxplots (median, interquartile range, and 1.5×IQR whiskers with outliers omitted). Individual points are single groups, colored by seed sequence type (ST). Each thin line connects one group (a single ST × replicate) across the four endpoints, colored by the number of genomes in that group (Genome Count). These connectors show that phylothresholds rise from *Peak* through *Discrepancy* and that lower sampling rates require larger thresholds to link genomes. Sequence type had no significant effect on the phylothreshold within any endpoint–rate stratum except discrepancy at 80% sampling (Kruskal–Wallis with Benjamini–Hochberg correction across the 16 strata at rates 0.2–0.8).

Sensitivity rose with the cutoff value but remained strongly dependent on sampling completeness. Even at a cutoff of 50, mean sensitivity climbed from 70.56% at 20% sampling to 94.46% at complete sampling (Figure 5), confirming that single-linkage clustering on SNP distances alone, the prevailing standard, is inherently limited when sampling is sparse.

THRESHER’s phylogeny correction substantially mitigated this limitation. At the lowest sampling rate (20%), three of the four endpoints already achieved high sensitivity (*peak* 73.44%; *plateau* 15 76.95%; *discrepancy* 99.61%); the exception, plateau 5, rose sharply from 24.06% to 85.13% by 60% sampling. Critically, the endpoints occupied distinct positions along the sensitivity–specificity trade-off, with the plateau length being a critical factor for the *plateau* endpoint.

The *discrepancy* endpoint favored sensitivity (mean 99.66%; median 100%; IQR 99.58–100%) over specificity (mean 88.65%; median 87.67%; IQR 84.05–91.70%), achieved through the highest phylothresholds of any endpoint (mean 108.41; median 100; IQR 73–136) (Figure 5). Sequence type had no significant effect on phylothresholds within any subsample rate for any endpoint except *discrepancy* at 80% sub-sampling (*p*_FDR_ = 0.013). Across the remaining strata, median differences among the five sequence types spanned from 3.0 to 46.5 SNPs. For every endpoint, phylothresholds shifted further with sampling completeness than with genetic background (Supplementary Table 6). These thresholds rose as sampling worsened, from a median of 69 at complete sampling (mean 76.92; IQR 62–83) to 156.5 at 20% sampling (mean 156.28; IQR 127–185.5). With sparser sampling, more genomes during the simulated evolution are not sampled, which leaves larger SNP distance gaps between the sampled genomes. Grouping genomes from the same strain therefore requires a higher phylothreshold. A lower fixed cutoff would split these genomes into separate strains, contrary to the simulated ground truth.

The *peak* endpoint showed similar sampling dependence but leaned toward specificity, producing the most stringent phylothresholds (mean 24.60; median 15; IQR 10.00–29.75). Its sensitivity declined as sampling improved. The *peak* endpoint sets the phylothreshold at the value that maximizes the number of clones, yielding the finest resolution. The *peak* endpoint thus minimizes false alarms at the cost of underestimating cluster size.

The *plateau* endpoint balanced sensitivity and specificity, with its plateau window length setting the operating point, a length of 15 assumes a 15-SNP natural gap between strains, whereas a shorter window reaches the plateau at lower phylothresholds (plateau 5: mean 26.50, median 27, IQR 18.25–34.00; plateau 15: mean 56.61, median 56, IQR 40–69). As with the other methods, the larger window raised sensitivity, most markedly under poor sampling, where a wider distance span is needed to recover linkages: at 20% and 40% sampling, plateau 5 reached only 24.06% and 61.73% sensitivity, against 76.95% and 96.46% for plateau 15.

Together, these results show that although raising a FIXED cutoff improves sensitivity while preserving specificity, it remains heavily compromised by incomplete sampling. THRESHER overcomes this problem by correcting SNP-based strain assignments against the group phylogeny, allowing it to outperform fixed cutoffs while deriving its phylothresholds from the data rather than from preset values. Each endpoint encodes a different hypothesis about what constitutes a strain; given that sampling in real clinical settings is often incomplete, with many cases and sources unsampled^21^, plateau 15 offers a well-balanced default for accurate transmission-cluster inference under realistic sampling. We therefore set it as THRESHER’s default, with a plateau window length of 15.

### THRESHER reproduces strain composition in small studies and reveals under- and overestimation in large ones

Using plateau 15, we applied THRESHER to the 55 published studies introduced in Figure 1 (per-study THRESHER outputs are provided in Supplementary Data 1). The studies span *S. aureus, K. pneumoniae*, and *C. difficile* and were conducted worldwide, predominantly in clinical settings but also in community, zoonotic, and hybrid contexts. This breadth of geography and predominant genetic background was chosen to probe generalizability and limit location- or lineage-specific bias (Figure 6, Supplementary Table 1). Study size varied widely (mean 125.38, median 76, IQR 34.5–148.5), ranging from 9 (S33, K15) to 598 (S53) genomes, spanning the sampling depths and durations summarized in Figure 1.

**Figure 6.**
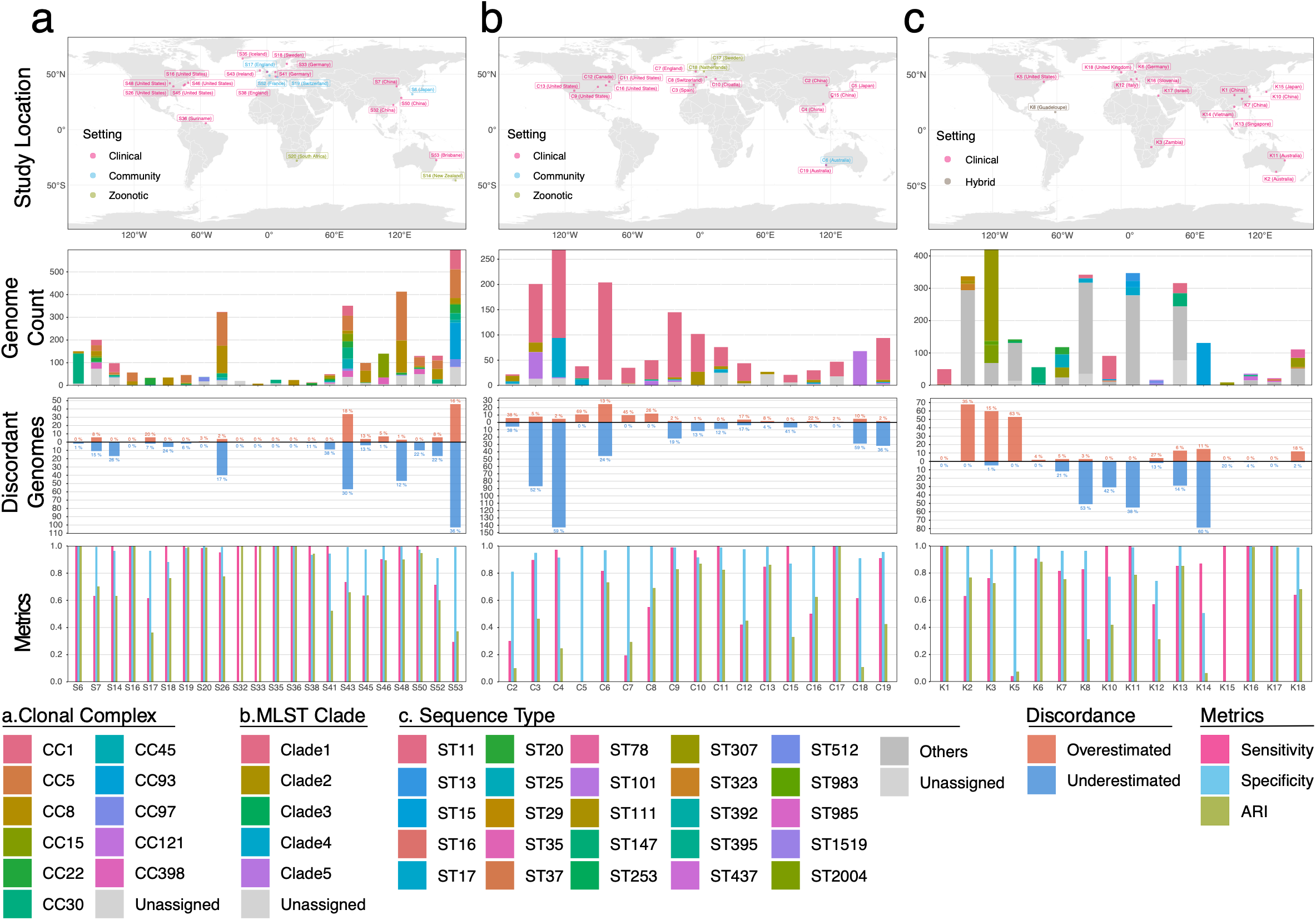
Concordance between THRESHER and originally reported strain assignments across 55 published studies. Results are shown for three species in separate columns. **(a)** *S. aureus* (n=22 studies), **(b)** *C. difficile* (n=17 studies), and **(c)** *K. pneumoniae* (n=16 studies). THRESHER was run using the *plateau* endpoint with 15 plateau length. Within each column, four rows describe the same set of studies (Study ID on the x-axis). Row 1 (study location). Geographic origin of each study, with points colored by setting (Clinical, Community, Zoonotic, or Hybrid where a study spans more than one setting). Row 2 (predominant genomic background). Composition of each study by predominant genomic background—clonal complex (*S. aureus*), MLST clade (*C. difficile*), or sequence type (*K. pneumoniae*), as stacked genome counts. Minor or untyped backgrounds are pooled as “Unassigned.” Row 3 (discordant genomes). Genomes whose original assignment placed them in a smaller strain than THRESHER (underestimated, below the axis) or a larger strain (overestimated, above the axis). Bars and the y-axis indicate genome counts, and the annotation above each bar shows the corresponding percentage of the genomes. Row 4 (concordance metrics). Per-study sensitivity, specificity, and adjusted Rand index (ARI) of the original assignment relative to THRESHER. In all rows, sensitivity, specificity, ARI, and estimation are measures of agreement with the THRESHER assignment and quantify concordance rather than accuracy; see Methods.

Taking the median (76 genomes) as the cut point, we designated studies with ≤76 genomes as small and the remainder as large. Because of the lack of known ground truth in real-life data, the metrics below quantify concordance between THRESHER and the originally reported assignments rather than accuracy; “overestimation” and “underestimation” denote genomes that the original assignment placed in a larger or smaller strain, respectively, compared to that inferred by THRESHER. For small studies (n = 28; *S. aureus* 11, *K. pneumoniae* 6, *C. difficile* 11), concordance was high. Few genomes were overestimated (mean 10.49%, median 0%, IQR 0–18.04%) or underestimated (mean 10.41%, median 2.08%, IQR 0–14.35%), with high sensitivity (mean 80.35%, median 100%, IQR 60.37–100%), specificity (mean 92.26%, median 99.39%, IQR 93.60–100%), and Adjusted Rand Index (ARI) (mean 67.98%, median 84.22%, IQR 35.47–99.81%).

Large studies (n = 27; *S. aureus* 11, *K. pneumoniae* 10, *C. difficile* 6) retained comparable overall concordance, with sensitivity (mean 80.89%, median 86.98%, IQR 72.37–98.06%), specificity (mean 94.89%, median 97.41%, IQR 95.90– 99.35%), and ARI (mean 63.37%, median 70.17%, IQR 44.40–80.72%; Methods 4.2), but underestimation rose markedly (mean 23.39%, median 21.43%, IQR 12.25–36.18%), while overestimation remained similar (mean 9.42%, median 5.19%, IQR 1.40–13.10%) (Figure 6).

To understand this difference between large and small studies, we mapped both THRESHER and the originally reported strain assignments onto the group phylogeny of every hierarchical group in every study (Supplementary Data 3), revealing several typical recurring scenarios. In small datasets of limited genomic diversity, a single phylogeny captures the full genomic diversity and strains can be read and determined directly from the tree. Here, THRESHER and the original assignments were identical or nearly so (over- and underestimation ≤5%; e.g., S16, S20, S32, S33, S35, S36, C17, K1, K6, K17). The same held for larger datasets dominated by a single genomic background (e.g., S6, S45, S46).

As genomic diversity increased, multiple and often confounding factors degraded the original assignments. Underestimation typically arose from reliance on SNP distances alone, as observed in the simulations. In S26, for example, despite a permissive ≤71 SNP cutoff, several originally reported strains (Strains 1, 7, 12, 13, 17, 36 in Group 1) were not monophyletic in the group phylogeny, while THRESHER’s phylogeny correction restored monophyly by recovering the missing members. Within the same group, original strains 8 and 9 fell within a single clade, and strains 37 and 47 shared an unrecognized link. Both were corrected by THRESHER (all four example original strains were drawn from Group 1 of S26, visualization provided as S26/Group1.pdf in Supplementary Data 3).

Overestimation, by contrast, traced to reference-dependent distance estimation. In K2, THRESHER partitioned the population into three hierarchical groups, indicating substantial background diversity, whereas the original study computed SNP distances and inferred phylogenies against a single reference. Within Group 3 of K2, the divergences were miscalculated in the original study, such that lineages 45, 46, and 47, hundreds of SNPs apart by reference-free, all-versus-all comparison in THRESHER, were grouped under a single strain despite the “strict” ≤25-SNP cutoff. This discrepancy in SNP distance reflects the two approaches counting over different sets of base positions. Reference-based mapping resolves only sites covered across all genomes, so divergent genomes may lose the variation before the distance is computed, whereas all-versus-all comparison used in THRESHER recovers the full divergence between each pair (the example plot for Group 3 of K2 is provided as K2/Group3.pdf in Supplementary Data 3).

Versions of these patterns were detected in multiple studies. Rather than describe every study individually, we provide the full data in Supplementary Data 3, organized as one folder per study. Each folder contains the visualization per hierarchical clustering group, annotated with the THRESHER and originally reported strain assignments on the group phylogeny, along with a corresponding file storing the study’s concordance metrics.

Across all 524 hierarchical groups from 55 re-analyzed studies (*S. aureus*, 212 groups from 22 studies; *C. difficile*, 161 from 17; *K. pneumoniae*, 151 from 16), the final phylothresholds fell largely within the range of cutoffs used in the published literature (Supplementary Table 1) but corrected for phylogenetic structure (Supplementary Figure 2; per-endpoint values in Supplementary Table 2). Pairwise ARI among the *peak, plateau*, and *discrepancy* endpoints was uniformly high. *Public*, however, was excluded because its quality depends on the number of genomes deposited at NCBI across species. The ARI distribution indicates that although the endpoints encode different definitions of a strain, their assignments largely agree on real data (Supplementary Figure 4).

Taken together, our systematic review and re-analysis of 55 published studies across three bacterial species revealed a shared limitation in the field, which is fixed SNP cutoffs adopted without biological justification. In small, or larger ones dominated by one genomic background, a single phylogeny captured the full diversity and phylothresholds implemented by THRESHER reproduced the published compositions. Such agreement is necessary but not sufficient as THRESHER is most needed where a single phylogeny no longer captures the full diversity. As genomic diversity increased, the published assignments became less accurate, with underestimation when SNP distances were used without phylogenetic context and overestimation when distances depended on the choice of reference genome. By correcting for phylogenetic topology and inferring reference-free pairwise SNP distances, THRESHER addresses both sources of error, replacing fixed cutoffs with data-driven phylothresholds grounded in each dataset’s genomic structure. The resulting strain compositions provide a more robust and principled basis for downstream epidemiology.

## DISCUSSION

Whole-genome sequencing is now a global standard for high-resolution tracking of bacterial pathogen dissemination, and fixed SNP cutoffs are deeply embedded as the conventional method to define transmission clusters^22, 23^, yet no consensus value has been established^8-10, 24^, and cutoffs are most often inherited from earlier work rather than derived from the data at hand. Because genetic distance and epidemiological linkage covary with sample size, sampling duration, setting, and patterns of transmission, colonization and infection, a cutoff calibrated in one study may not transfer to another, and the reliance on fixed, arbitrary cutoffs limits the accuracy and comparability of strain and transmission inference. A growing body of work has recognized that a single universal cutoff is unlikely to suit the full diversity of genomic backgrounds encountered across pathogens, settings, and sampling scenarios^8, 9, 22, 24^. However, these efforts have largely stopped short of a generalizable, reusable solution: they have not provided a systematic procedure that transfers across studies, or a ready-to-use implementation that researchers can apply directly to their own data^8-10^. THRESHER addresses this gap by inferring a phylogeny-corrected threshold—the phylothreshold—from the genomic structure of each dataset, packaged as a command-line tool with an accompanying locally deployable assistant to lower the barrier to adoption across levels of expertise.

Here we distinguish what THRESHER establishes from what it leaves to the users. Phylothresholds inferred by THRESHER can be considered an attribute of a dataset’s own genomic structure. The corresponding strain composition is the partition obtained at that phylothreshold under the chosen endpoint method, with each method encoding a distinct hypothesis of what evidence supports a “strain”. This output is distinct from a further question that genomic data alone cannot settle: whether that structure, and the partition derived from it, reflect the biological or epidemiological reality of a given setting and serve the purpose at hand. Answering that question rests on the user’s knowledge of their own sampling and context. The two are separable because “strain” is not a fixed concept but a context- and purpose-dependent one: a phylothreshold inferred faithfully from the data may still produce a partition that lacks the sensitivity or specificity to delineate the units a particular investigation requires, even as the same groups remain valid strains under a different definition. For this reason, THRESHER does not present its partitions as a verdict, but presents the full genomic structure it summarizes through extensive visualizations and all intermediate data, so that users can bring their knowledge of sampling, setting, and purpose to bear on whether a given phylothreshold and its strains are meaningful for their question.

A few limitations remain, and each points to ongoing work. The first concerns CladeBreaker, which is enabled by default and encodes a specific assumption: that the strains constituted of study genomes are hyperlocal and should not be grouped with publicly available genomes already deposited at GenBank. This assumption is reasonable in many surveillance contexts, but it cannot be verified automatically. Isolates collected at one location are sometimes deposited across separate BioProjects, and an investigator may not know whether genomes related to their own have already been submitted; in such cases CladeBreaker cannot itself determine whether a matched public genome should be excluded, or whether it should be disabled for correcting strain compositions altogether. Correct use therefore depends on the user’s prior knowledge, and mishandling can lead to misidentification. For the present study, whenever close matches arose from the public-genome database, we manually examined the geographic origin and, where available, the collection date of the matched genomes to decide whether CladeBreaker should be active for a given run. These settings are recorded in Supplementary Table 3. As with any distinguishing quality, separating genomes based on origin should also be adjudicated with caution. For instance, among the *C. difficile* datasets, C17 (Sweden) and C18 (the Netherlands) each turned up among the other’s closest public genomes through samples collected from pigs in both datasets. That two independent studies in neighboring countries, sampling the same host, should retrieve each other’s genomes is unlikely to be coincidental. It might point to a real biological link that CladeBreaker, by design, would treat as a reason to keep the strains apart. Public-genome matching can therefore both constrain strain expansion and also point to relationships that reach beyond a single study. However, interpreting it correctly takes judgment that the tool currently leaves to the user. Making this assessment automatic is a direction for future work.

A second limitation concerns the *plateau* endpoint. Although our simulation results identify Plateau 15 as a well-balanced default, the plateau length itself remains a fixed parameter rather than an evidence-based, data-driven quantity. The window length encodes an assumption about the width of the natural gap separating strains, and while we show that adjusting it tunes the sensitivity–specificity balance predictably, we do not yet infer the appropriate width from the data. Deriving the plateau length directly from each dataset’s divergence structure is a natural extension of the phylothreshold principle and a focus of continuing work.

The *plateau* concept itself rests on an empirical observation that did not hold uniformly across species. The motivation for using a natural gap to capture strain-level structure came from the distribution of pairwise SNP distances, in which a distinct cluster of small distances reflects close clonal relationships. We observed this pattern frequently in *S. aureus* and *K. pneumoniae* but far less so in *C. difficile*. We lack direct evidence for the cause, but we speculate that it may relate to a biological feature distinguishing *C. difficile* from the other two species: its capacity to form spores during colonization, transmission, and environmental persistence under stress. Sporulation may alter the rate or pattern of molecular change in ways that are not captured by substitution-rate models calibrated on non-sporulating organisms, and this is a factor worth considering in future efforts to model the evolution of spore-forming pathogens.

Finally, THRESHER’s emphasis on accuracy comes at a computational cost. To maximize precision, several steps are deliberately exhaustive: SNP distances are computed by all-versus-all pairwise comparison, and core-gene phylogenies are inferred using all study genomes. Both of these approaches scale unfavorably with the number of genomes, and we anticipate that very large datasets, on the order of a thousand genomes or more, will require substantially longer run times than the datasets analyzed in this study. Benchmark and resource-usage data are provided in Supplementary Table 4. We regard this exhaustiveness as a deliberate trade-off in favor of accuracy. In the interim, a practical way to reduce run time on large datasets is to partition the genomes before analysis using prior knowledge to split them along established typing groups, such as clonal complexes in *S. aureus*, MLST clades in *C. difficile* or other typing methods, and then run THRESHER separately on each subset. Because genomes from different typing groups are less likely to belong to the same strain, this rarely separates genomes that THRESHER would otherwise have grouped, while substantially lowering the cost of the two limiting steps, as each run builds a smaller all-versus-all SNP matrix and a smaller core-gene phylogeny. A future version will automate this partitioning; validating and generalizing such an approach, ideally without sacrificing accuracy, will require additional datasets.

Taken together, these results show that the threshold separating one strain from another need not be an *a priori* convention applied uniformly across different datasets, but can instead be inferred as an attribute of each dataset’s own genomic structure. By replacing fixed cutoffs with phylogeny-corrected phylothresholds, THRESHER recovers transmission clusters that strict cutoffs miss while remaining robust to the incomplete sampling that characterizes real surveillance. It makes the choice of threshold explicit, reproducible, and tied to evidence rather than precedent. As genomic surveillance continues to scale and to span an ever-wider range of pathogens and settings, methods that adapt to the data rather than imposing a single rule upon it will be increasingly necessary for strain and transmission inference that is both accurate and comparable across studies. THRESHER is a step toward that goal, and an openly available foundation on which such inference can be built.

## METHODS

### 1. THRESHER

THRESHER is organized into four stages (Figure 2): INPUT (Section 1.1), which ingests and validates the user’s data; PREP (Section 1.2), which characterizes overall genomic structure and assembles the inputs for CORE analysis (Section 1.3); CORE, which infers the phylothreshold and resolves strain composition for each group; and the optional EPI (Section 1.4), which links strains to patients to identify transmission clusters. Each stage is described in detail below.

#### 1.1 INPUT (Input Validation)

Before any analysis is launched, THRESHER validates the user’s inputs to fail early and informatively rather than partway through the workflow. The metadata table is checked for the required columns (genome name, accession, file path, and, for transmission-cluster analysis, patient identifier and collection date), and each genome name is screened for uniqueness, sanitized of characters that would interfere with downstream parsing. The corresponding assembly files are confirmed to exist and to be free of duplicate paths. A minimum of four genomes is required, and the accession field must be filled for every isolate. Genomes already deposited in GenBank carry their accession, while those not yet deposited must be marked with the explicit placeholder “new”. We require this marker deliberately rather than allowing the field to be left blank, so that designating a genome as undeposited is an affirmative choice the user makes rather than an omission. The requested species is checked against those supported (*S. aureus, S. epidermidis, C. difficile*, and *K. pneumoniae*), and all numerical and categorical parameters, including the threshold floor and ceiling, singleton threshold, plateau length, bootstrap settings, endpoint method, and thread count, are validated against their permitted ranges, with out-of-range or missing values reset to documented defaults. When patient identifiers or collection dates are incomplete, THRESHER automatically falls back to strain identification without transmission-cluster inference. THRESHER additionally verifies that the host system meets the operating-system and memory requirements of its dependencies before launching the workflow.

#### 1.2 PREP (Preparatory Step)

##### 1.2.1 Core-Gene Maximum-Likelihood Phylogeny

Genome assemblies were annotated with Bakta v1.11.4 with full database v6.0^25^, and the resulting GFF3 files were used for core-genome alignment in Panaroo v1.5.1^26^. Panaroo was run in strict cleaning mode with invalid genes removed, a family threshold of 0.7, a refind proportion-match of 0.5, and a search radius of 5,000 bp. Core genes were aligned with MAFFT^27^, with the fraction of isolates required for a gene to be considered core set by a user-configurable core threshold (default 0.95). The resulting core-gene alignment was used to infer a maximum-likelihood phylogeny in IQ-TREE v2.3.6^28^ under the general time-reversible model (GTR+R)^29^, with the number of threads selected automatically. Branch support was assessed by one of two user-configurable methods: ultrafast bootstrap^30^ (UFBoot, the default) or nonparametric bootstrap, each additionally accompanied by an SH-aLRT test using the same number of replicates; UFBoot defaults to 1,000 replicates and the nonparametric option to 100. In both cases, UFBoot’s reduced susceptibility to model violation was enabled (the --bnni optimization).

##### 1.2.2 Comprehensive Study SNP Matrix

Pairwise SNP distances were computed with the dnadiff() wrapper in MUMmer v4.0.0rc1^31^ taking the gSNP count from the dnadiff() report as the raw distance between two genomes.

The study SNP matrix quantifies the genomic distance between every pair of study genomes. Let *SG* = {*sg*_1_, *sg*_2_, …, *sg*_*n*_} be the set of *n* study genomes. For each ordered pair (*sg*_*i*_, *sg*_*j*_) with *i* ≠ *j*, MUMmer performs a whole-genome alignment with *sg*_*i*_ as reference and *sg*_*j*_ as query, from which it reports the number of SNPs in the alignment and the proportion of the query covered by alignments compared to reference. Constructing the matrix therefore requires all *n*(*n* − 1) directed comparisons, yielding an asymmetric raw distance matrix whose entries are

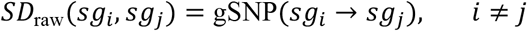

Whole-genome alignment is not perfectly symmetric as the assignment of reference and query affects which regions align, so the two directed distances for a pair are averaged to give a single symmetric distance:

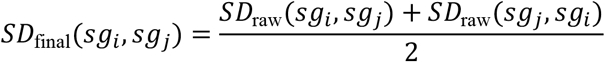

Each directed alignment also yields an alignment coverage, the fraction of the reference genome covered by alignments to the query:

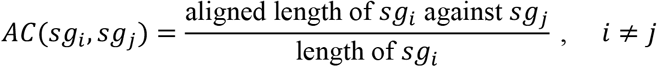

A SNP distance is meaningful only when coverage is high enough that it reflects divergence across the whole genome rather than a small aligned fraction, which would understate the true difference between two assemblies. For each pair, both directed coverages are therefore evaluated against a configurable alignment-coverage threshold (default 0.80). The pair is labeled “good” quality only if both *AC*(*sg*_*i*_, *sg*_*j*_) and *AC*(*sg*_*j*_, *sg*_*i*_) meet or exceed the threshold, and “poor” otherwise.

Poor-quality pairs are retained but flagged, and are highlighted in red fonts in the SNP-distance heatmap accompanying the strain-composition visualization so that low-coverage comparisons remain visible to the user. As a safeguard, if a study genome is of poor quality against all other study genomes in its group, it is automatically treated as a singleton and excluded from the subsequent analysis of that group. This is expected to be exceedingly rare, since THRESHER requires stringent assembly quality at input. In practice, it arises when a genome of a different species is included, as only such a genome would fail to align well against every other member of the group. The singleton fallback therefore also acts as a check against this kind of input error.

Distances were computed in parallel across available cores, and redundant directional comparisons were collapsed so that each unordered pair contributes a single averaged distance and a single quality label.

##### 1.2.3 Population-structure assessment and Hierarchical Clustering

Because a single phylothreshold is unlikely to be appropriate across highly divergent genomic backgrounds, THRESHER next decides whether the dataset can be analyzed as a whole or should be partitioned into groups for independent parallel downstream analysis. The core-gene ML phylogeny is midpoint-rooted using midpoint_root() with R package phytools^32^. For every internal node *v*, the descendant tip set *L*(*v*) and the node’s bootstrap support values are recorded.

The partitioning decision rests on two conditions, both of which must hold for Hierarchical Clustering to be skipped.

Condition 1: Single typing group spanning the tree. Let *T* = {*t*_1_, …, *t*_*m*_} be the set of typing labels (*S. aureus:* Clonal Complex; *K. pneumoniae*: Sequence Type; *C. difficile*: MLST Clade) assigned to the study genomes after excluding the “Unassigned” label, and let *G* ⊆ *SG* be the genomes carrying an assigned label. Condition 1 holds when exactly one typing label remains,

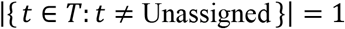

and the smallest clade containing all labeled genomes is the root of the tree.

In practice, the smallest spanning clade is identified by selecting, among all nodes whose descendant tip set contains *G*, the node with the fewest descendants. Condition 1 is satisfied only if that node is the root. Intuitively, this holds when the dataset comprises a single genomic background that occupies the entire tree, so that no partitioning into distinct backgrounds is needed.

Condition 2: No discontinuity in the pairwise SNP distribution. Condition 1 alone is insufficient because a single typing label does not guarantee that the genomes form one closely related group as typing groups are not always monophyletic. Among the *S. aureus* representative genomes, for example, CC1, CC5 and CC8 are paraphyletic (BACKBONE panel, Figure 4; Supplementary Figure 3), and a single clonal complex can encompass multiple sequence types separated by thousands of SNPs. Treating such a typing group as a single group would therefore be inappropriate, so a second condition tests directly for natural discontinuities in the genomic distances rather than relying on typing alone.

Condition 2 is evaluated only when Condition 1 holds. The distribution of the symmetric pairwise SNP distances among the study genomes, *d*_*k*_ = *SD*_final_(*sg*_*i*_, *sg*_*j*_) over all 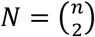 pairs, is summarized by kernel density estimation using the density() function of the R stats package with a Gaussian kernel and a fixed bandwidth *h* = 1000 bp. The kernel is one whose statistical properties are determined by 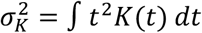, which equals 1 for the kernels used by density(), so that the bandwidth corresponds to the standard deviation of the kernel. The number of modes in the estimated density is then taken as one more than the number of interior local minima, with the local minima identified numerically as sign changes in the first difference of the density. Condition 2 holds when the density is unimodal: a single mode indicates that the genomes form one continuous cluster, whereas additional modes indicate natural discontinuities in divergence that warrant separation.

When both conditions hold, hierarchical clustering is skipped and all study genomes are assigned to a single group. Otherwise, the genomes are partitioned as follows. A pair-wise distance matrix *D* is obtained using cophenetic.phylo() in R package ape^33^, from the core-gene ML phylogeny, where

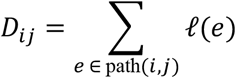

is the sum of branch lengths *l*(*e*) along the path between paired tips *i* and *j*. Hierarchical clustering with complete linkage is applied to *D* using hclust() in built-in R package stats^34^. For each candidate cluster count *k* ∈ {2, …, *n* − 1}, the genomes are cut into *k* clusters and the mean silhouette width is computed,

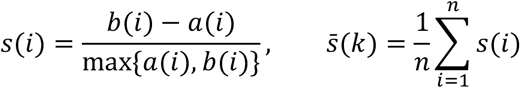

where *a*(*i*) is the mean distance from tip *i* to the other tips of its cluster and *b*(*i*) is the lowest mean distance from *i* to any other cluster. The dataset is partitioned at the cluster count that maximizes the mean silhouette width across the tested range,

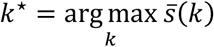

The single-group outcome is decided upstream by Conditions 1 and 2, so the silhouette search considers only genuine partitions (*k* ≥ 2).

Because cuts of a cophenetic tree may not necessarily correspond to monophyletic clades, each resulting group is then reconciled with tree topology. If a group’s genomes do not form an exact clade, the smallest clade containing all of them is identified and its additional descendant genomes are absorbed into the group. Finally, within each multi-genome group, any genome whose minimum pairwise SNP distance to the remaining group members is at least the singleton threshold (a user-configurable parameter, default 100 bp. The same threshold applied later in the core algorithm) is separated from the group as a singleton. If no within-group pair falls below this threshold, the entire group is treated as singletons.

##### 1.2.4 Identification of closely related public genomes

The closely related public genomes identified for each study genome serve two purposes in the downstream core algorithm. First, they act as guardrails on strain composition. Because the core algorithm tests every candidate phylothreshold and strain clades expand on the tree as the phylothreshold increases, the public genomes mark how far that expansion may go. When CladeBreaker is enabled, a strain clade is not permitted to grow to encompass a public genome, constraining each strain to the study population. When it is disabled, the public genomes remain in the phylogeny but impose no such constraint, so the phylogeny correction could expand a strain clade past the public genomes. However, in either case, public genomes are never assigned to the same strain as study genomes. Second, they define the public endpoint, at which the phylothreshold is set to the smallest SNP distance between any study genome and its closest public genome. To support both uses, every study genome *sg*_*i*_ is queried against a species-specific WhatsGNU database of publicly deposited GenBank genomes^35^. WhatsGNU ranks database genomes by the number of exact-match proteins shared with the query and returns the top *k*(configurable; default *k* = 20) most closely related public genomes for each *sg*_*i*_. The public reference set is the union of these per-genome matches across all study genomes.

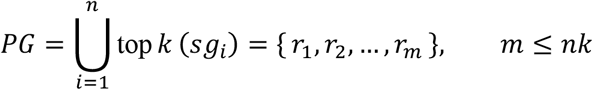

##### 1.2.5 Multi-locus sequence typing

Each study genome is assigned a sequence type with mlst v2.23.0^36^, which scans the assembly against the PubMLST database and reports the sequence type^37^. A genome whose allelic profile matches no defined sequence type is labelled as Unassigned. These assignments are used in two places: by the population-structure assessment (Section 1.2.3), where the presence of a single typing label across the dataset is one of the two conditions under which the dataset is treated as one group (Condition 1), and by the optional epidemiological step (Section 1.4), where each transmission cluster is annotated with the sequence type.

#### 1.3 CORE (Core Algorithm)

Sections 1.2.1–1.2.4 prepare, for each group, the all-versus-all SNP distances, and the matched public genomes that the core algorithm consumes. CORE then runs independently on each hierarchical clustering group *H*_*k*_ returned by the population-structure step, or on the whole dataset when partitioning was bypassed.

Within a group, CORE first builds two inputs in parallel, a reference-based ML phylogeny and a Study-Public SNP matrix. Let *HG* = {*g*_1_, …, *g*_*n*_} be the study genomes assigned to group *H*_*k*_ and *HP* = {*p*_1_, …, *p*_*m*_} the closely related public genomes matched to them, so that *HG* ∪ *HP* is the combined set of study and public genomes in the group *H*_*k*_. When the group contains at least four genomes in total (∣ *HG* ∪ *HP* ∣≥ 4), a multiple-sequence alignment is generated with Snippy v4.6.0^38^ from all study genomes and their matched public genomes, using the study assembly with the highest N50 as the reference. IQ-TREE v2.3.6^28^ then infers a reference-based maximum-likelihood phylogeny from this alignment, under the same substitution model and branch-support settings as the core-gene ML phylogeny (Section 1.2.1). Groups with fewer than four genomes are not given a tree.

In parallel, the group-specific Study-Public SNP matrix is assembled. With *HG* and *HP* as defined above, let *d*(*g*_*i*_, *g*_*j*_)=*SD*_final_(*g*_*i*_, *g*_*j*_) be the SNP distance between two study genomes, averaged over both alignment directions, taken from the comprehensive study SNP matrix (Section 1.2.2). Study genomes are not compared against every public genome in the group but each study genome *g*_*i*_ is compared only with its closely related public genomes, the subset *HP*_*i*_ ⊆ *HP* identified for *g*_*i*_ by WhatsGNU (Section 1.2.4), so that 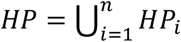. For each such pair, a study genome *g*_*i*_ and one of its matched public genomes *p*_*j*_ ∈ *HP*_*i*_, the distance is obtained by the same dnadiff (MUMmer v4.0.0rc1^31^) procedure used for the study matrix (Section 1.2.2), aligning the two genomes in both directions and averaging:

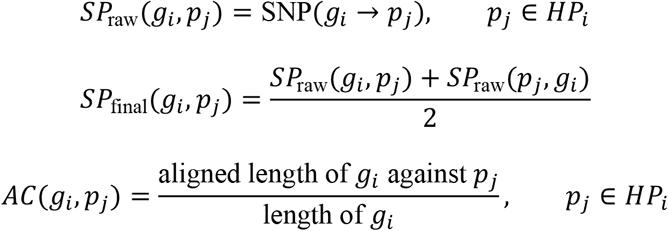

A study–public distance is flagged good when the alignment coverage in both directions meets a user-configurable threshold (default 80%) and poor otherwise. CladeBreaker later uses this flag to discard unreliable public genomes for strain composition correction (Section 1.3.4). Collecting the matched pairs into

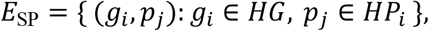

the Study-Public SNP matrix over the combined genome set *HG* ∪ *HP* is constituted from the extracted study block 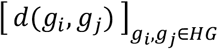, taken directly from the comprehensive study SNP matrix (Section 1.2.2), together with the study–public block 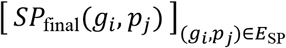. Pairs outside *E*_SP_, which are study genomes with unrelated public genomes, and all public–public pairs, are not required by the algorithm and are not computed, leaving the study–public block sparse.

With the Study-Public SNP matrix and the group reference-based phylogeny if applicable, CORE scans the full range of candidate SNP thresholds and, at each one, assigns the study genomes to strains in up to three steps, which are single-linkage clustering on SNP distances, correction against the group phylogeny, and CladeBreaker. Then the resulting strain composition is recorded for each tested threshold. We refer to each tested threshold as a *candidate phylothreshold*. The value ultimately retained for the group is selected by each of the four currently supported endpoint methods (Section 1.3.6). Correction using the phylogeny is applied only when the group tree exists (∣ *HG* ∪ *HP* ∣≥ 4). Otherwise, strains are assigned with SNP distances alone. The scan is bounded by the observed data: the floor is the larger of τ_floor_(default 5) and the smallest study–study SNP distance in the matrix, and the ceiling is the smaller of τ_ceil_ (default 500) and the largest study–study SNP distance, so no cutoff below the closest study pair or above the most distant study pair is tested, shown below.

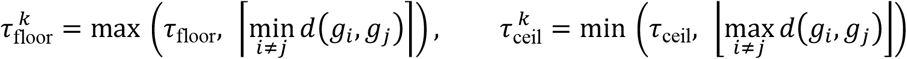

The superscript *k* in 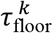 and 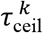 indexes the hierarchical clustering group *H*_*k*_, since both bounds are derived from the SNP distances within that group and therefore differ between groups. The singleton threshold τ_single_ (default 100) and the minimum bootstrap support *β*_min_required to accept a phylogenetic correction are likewise user-configurable.

The remainder of this section details each component of the core algorithm in turn: the genomes set aside as singletons before the scan and the handling of all-singleton groups (Section 1.3.1), single-linkage clustering on SNP distances at each candidate phylothreshold (Section 1.3.2), correction of the resulting clusters against the group phylogeny (Section 3.3), the optional CladeBreaker constraint imposed by public genomes (Section 1.3.4), the strain compositions recorded across the full range of candidate phylothresholds (Section 1.3.5), and the endpoint methods that select each group’s final phylothreshold from that record (Section 1.3.6).

##### 1.3.1 Overlimit exclusion and singleton handling

Before the scan, study genomes that the population-structure step flagged as exceeding the within-group overlimit distance λ (Section 1.2.3) are set aside as the overlimit set *HG*_over_ ⊆ *HG*. These genomes are too divergent from the remainder of the group to belong to any strain and are reported as singletons at every candidate phylothreshold. The scan therefore operates on the retained genomes, denoted as *HG*^′^. The lower end of the scan is the larger of the user floor τ_floor_ (default 5. The superscript *k* in 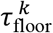 indexes the hierarchical clustering group *H*_*k*_.) and the smallest SNP distance among the retained genomes, rounded up to remain an integer,

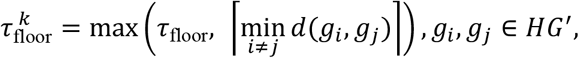

so that no cutoff below the closest retained pair is tested. Two cases resolve the group without a scan.

Firstly, if the overlimit step removed every study genome (*HG*^′^ = ∅), each genome in *HG*_over_ is reported as a singleton and no cutoff is recorded.

Secondly, If the closest retained pair is itself more divergent than the singleton threshold,

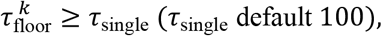

then no link can form at any tested phylothreshold, every study genome in *HG* = *HG*^′^ ∪ *HG*_over_ is reported as a singleton, and the cutoff is recorded as ⌈min_*i*≠*j*_ *d*(*g*_*i*_, *g*_*j*_)⌉ over the retained genomes.

In both cases the group yields no phylogeny-corrected strains, so the correction-dependent summaries, discrepancy, corrected-clone count, and strain bootstrap support, are returned as missing.

##### 1.3.2 SNP-based single-linkage clustering

At each candidate phylothreshold 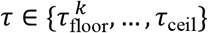 within hierarchical clustering group *H*_*k*_, two retained genomes are linked when their SNP distance does not exceed τ,

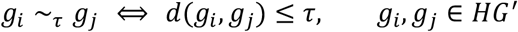

defining a graph *g*_τ_ = (*HG*^′^, *E*_τ_) with edge set *E*_τ_ = { (*g*_*i*_, *g*_*j*_): *g*_*i*_ ~_τ_ *g*_*j*_, *i* ≠ *j*}. The single-linkage strains at τ are the connected components of *g*_τ_, obtained by iterative label propagation in which each genome is assigned the smallest identifier,

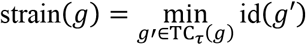

A component of at least two genomes is a *clone*; a genome with no link at τ is a *singleton*. The overlimit genomes *HG*_over_ set aside in Section 1.3.1 are appended as singletons, so that every study genome in the group receives an assignment. This yields the pre-phylogeny-correction counts

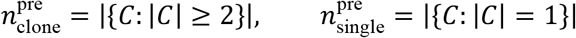

where each *C* ranges over the strains assigned at τ.

##### 1.3.3 Phylogenetic correction

When a group phylogeny *T*_*k*_ is available (∣ *HG* ∪ *HP* ∣≥ 4), each clone is checked and corrected using tree topology, because pairwise SNP distances can fail to link all genomes that share the most recent common ancestor (MRCA), such as when under incomplete alignment coverage or recombination. Let *L*(*v*) be the set of tips descending from an internal node *v* and *b*(*v*) for its bootstrap support, the minimum clade for a clone *C* is

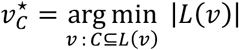

and its after-phylogeny-correction composition under the tree is 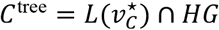, the study genomes descending from 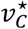(public genomes excluded). The genomes the tree adds to a clone beyond single linkage form the discrepancy set Δ_C_ = *C*^tree^ \ *C*, and the group’s discrepancy at τ is

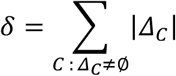

A phylogeny correction is accepted only when the clone clade *C*^tree^ is well supported 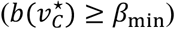, or when it is the root of the group tree 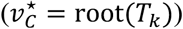. A clone after accepted correction has its membership expanded to *C*^tree^.

##### 1.3.4 CladeBreaker

By default, CladeBreaker constrains the size of corrected clones using the matched public genomes, under the assumption that the study genomes represent hyperlocal strains that should not be grouped with public genomes already deposited in public databases. Throughout this section *g* denotes a study genome (*g* ∈ *HG*) and *p* denotes a matched public genome (*p* ∈ *HP*). CladeBreaker acts on each corrected clone accepted in Section 1.3.3 whose clade contains at least one public genome. The public genomes appearing in that clade are 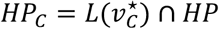. SNP quality is taken from the alignment coverage of Section 1.2.2, where a pairwise distance is good when its coverage meets a user-configurable threshold *c* (default 0.80) in both alignment directions and poor otherwise,

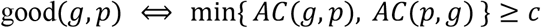

Two safeguards precede clade breaking. First, a study genome whose distances to every other study genome in the clade are poor is removed from the clade and reported as a singleton. This is a safety check that rarely triggers, because genomes are required to pass the user’s quality filtering before entering the analysis. Let *HG*_*good*_ ⊆ *C*^tree^ be the study genomes that survive this check, a public genome is retained for breaking only if it has no poor alignment to any surviving study genome, yielding the valid public set

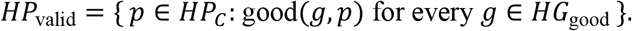

The clade is then partitioned into its largest study-only subclades. A node *u* defines the largest pure-study clade when its own clade contains no valid public genome, but its parent’s clade does,

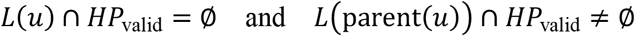

The clade is treated as pure when it carries no valid public genome. The study genomes within each such subclade constitute one separate strain, and any study genome not captured by a pure-study subclade is reported as a singleton.

CladeBreaker should be disabled manually when the user has evidence that the matched public genomes belong to the same strains as the study genomes. In that case no clade breaking is performed. Thus, the corrected clone keeps every study genome of the clade and drops only the public genomes, returning *C*^tree^.

Because correction and clade breaking are applied per clone, two corrected strains can come to share genomes. Overlaps are resolved by single-linkage merging, where among strains that share any genome, the larger absorbs the smaller through a union of their genomes, *S*_large_ ← *S*_large_ ∪ *S*_small_, and the pass is repeated until all strains are disjoint, so that every study genome is assigned to exactly one strain. The reconciled assignment gives the post-correction counts 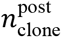 and 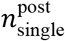, together with the mean and median bootstrap support taken over the clone strains.

##### 1.3.5 Strain composition across candidate phylothresholds

Sections 1.3.2–1.3.4 produce, at every candidate phylothreshold τ, a complete assignment of the group’s study genomes to strains. CORE records this assignment, together with a set of summary statistics, for every τ in the full scan range from τ_floor_ to τ_cell_. We refer to the complete assignment at a single τ as the complete strain composition *S*(τ).

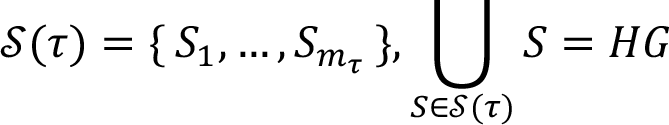

Each strain *S* ∈ *S*(τ) carries three attributes: its category, clone or singleton as defined in Section 1.3.2; a correction flag recording whether its composition was modified by phylogenetic correction or CladeBreaker (Sections 1.3.3, 1.3.4) rather than set by SNP single-linkage alone, and a bootstrap support *b*(*S*), taken as the support of the strain’s clade for a clone and set to zero for a singleton. When no group phylogeny is available, the correction flag and bootstrap support are recorded as missing.

The complete per-threshold record collects the composition together with the quantities defined in Sections 1.3.2–1.3.4,

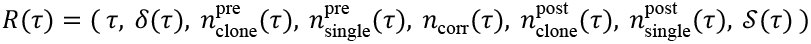

where *δ* is the discrepancy and *n*_corr_ the number of accepted corrections from Section 3.3, and the pre- and post-correction counts are those of Sections 1.3.2 and 1.3.4. Scanning over the full range gives the group’s intermediate database,

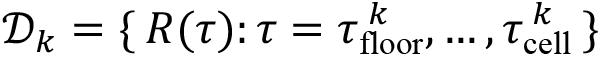

the strain composition of the group at every candidate phylothreshold together with its summaries. A group resolved without a scan (Section 1.3.1) contributes a single all-singleton record. For each group, *D*_*k*_ is visualized in a Multi-SNP-Threshold Plot (MSTP; see thresher/output/MSTP/Group#_MSTP.pdf within each study’s output directory in Supplementary Data 1), which traces the clone count, the singleton count, and the discrepancy across the candidate phylothresholds, with mean and median of bootstrap support of all clones in the group, shown in the upper panel.

From *D*_*k*_, each of the four currently supported endpoint methods selects the group’s final phylothreshold, and with it the strain composition reported for the group. These methods are described in Section 1.3.6.

##### 1.3.6 Endpoints

A single phylothreshold must be chosen so that a hierarchical clustering group contributes its strain compositions to the dataset-wide result from the intermediate database *D*_*k*_ (Section 1.3.5). An endpoint method makes this choice, analyzing *D*_*k*_ and returning the group’s final phylothreshold together with its composition *S*(τ). Four endpoint methods are currently supported, each encoding a different hypothesis about what defines one strain from the others and, with it, a different position on the sensitivity–specificity trade-off. The *plateau* method takes the phylothreshold at which the compositions remain unchanged over a contiguous run of candidate phylothresholds (Section 1.3.6.1); the public method takes the phylothreshold at which a clone first absorbs a public genome (Section 1.3.6.2); the *peak* method takes the phylothreshold that maximizes the number of clones (Section 1.3.6.3); and the *discrepancy* method takes the phylothreshold that minimizes the discrepancy beyond its initial *peak* (Section 1.3.6.4). Figure 3 presents the formal definition and graphical criterion for each method. The methods run independently. The composition at a group’s selected phylothreshold is written out as a strain table in which each strain receives the identifier ⟨group⟩_⟨index⟩ and is listed against its constituent genomes, concatenating these tables over all groups yields the final dataset-wide strain composition.

###### 1.3.6.1 Plateau

Let *S*(τ) be the strain compositions of a group at candidate phylothreshold τ (Section 1.3.5). For each τ in the scanned range [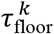, τ_ceil_], define an indicator of whether the composition is unchanged from the preceding phylothreshold,

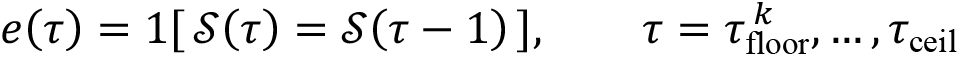

where the two complete strain compositions at the consecutive candidate phylothresholds are compared as partitions of the study genomes *HG*, independent of strain labelling or ordering. A *plateau* is a run of consecutive phylothresholds over which the composition does not change, that is, an integer interval [*a, b*] with *e*(τ) = 1 for all τ ∈ [*a, b*]. The length is *b* − *a*. With *L* the required plateau length (default 15), the *plateau* endpoint is the earliest run that reaches this length,

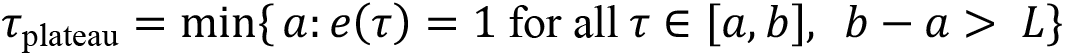

and the group’s reported composition is *S*(τ_plateau_), as the compositions are shared across [*a, b*].

Two outcomes are reported in words rather than as a phylothreshold. A group that was resolved without a scan, or whose smallest retained SNP distance already exceeds the singleton threshold τ_single_ (default 100), contains no clones and is labelled *Singletons*. When a qualifying plateau exists but its shared composition consists entirely of singletons, the *Singletons* label is also used, as no clone is present to anchor a phylothreshold. When no run reaches length *L*, the group is labelled *No Plateau Found* and the composition at the largest tested phylothreshold is reported.

The selected or labelled outcome for every group is recorded in a per-group plateau table, and the corresponding compositions are concatenated into the dataset-wide strain table described in Section 1.3.6.

###### 1.3.6.2 Public

Let *SP*_final_(*g*_*i*_, *p*_*j*_)(Section 1.3) be the symmetric SNP distance between a study genome *g*_*i*_ ∈ *HG* and one of its matched public genomes *p*_*j*_ ∈ *HP*_*i*_ (Section 1.3.4). The analysis below is restricted to pairs with good-quality study-public SNPs. Only a good-quality study-public SNP distance yields a reliable estimate of the genetic relatedness between a study genome and its public match, whereas a distance derived from a poor-quality call does not correspond to a biologically meaningful phylothreshold and is therefore excluded.

Over the good-quality study-public pairs, Let

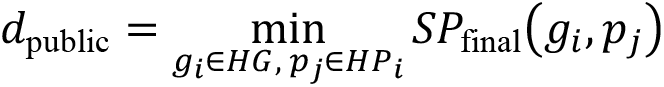

be the smallest study-to-public distance in the group. The public endpoint is this distance, rounded to the nearest integer and constrained to the scanned range,

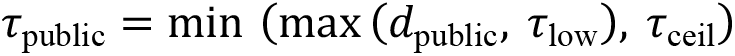

where τ_lo_ = min_τ_ is the smallest phylothreshold tested in the group, and τ_ceil_ (default 500) is the scan ceiling. The group’s reported composition is *S*(τ_public_), the composition at the largest tested phylothreshold not exceeding τ_public_ when no candidate equals it exactly.

The rationale is that the smallest study-to-public distance marks the point at which the group’s strains stop being exclusive to the study: at any phylothreshold below it, no public genome links to a study genome, whereas at τ_public_ a public genome first becomes eligible to be merged with a study strain. Setting the phylothreshold here leverages the boundary against publicly deposited, same-species genomes as the limit of strain composition. Public endpoint also serves as the complement for the CladeBreaker’s assumption (Section 1.3.4) that study strains are hyperlocal and should not be the “same” strain with public genomes. Because strain composition is determined jointly by SNP distance and tree topology (Sections 1.3.2–1.3.4), the phylothreshold at which a strain first incorporates a public genome need not coincide with *d*_pub_ itself but the public endpoint locates the SNP distance at which this incorporation becomes possible.

Note that the phylothreshold is capped at τ_ceil_, so that a distant nearest public genome does not push it past the scan. When the group yields all-singleton compositions, in which its smallest study–study SNP distance already exceeds the singleton threshold (Section 1.3.1), this group’s phylothreshold is reported directly. A group with no clone at any tested phylothreshold has no public boundary to locate and is reported as its all-singleton composition too.

The selected phylothreshold for every group is recorded in a per-group public table, and the corresponding compositions are concatenated into the dataset-wide strain table described in Section 1.3.6.

###### 1.3.6.3 Peak

Let 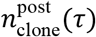 be the number of clones in the post-phylogeny-correction at candidate phylothreshold τ (Section 1.3.5). The *peak* endpoint is the phylothreshold attaining the maximum clone count,

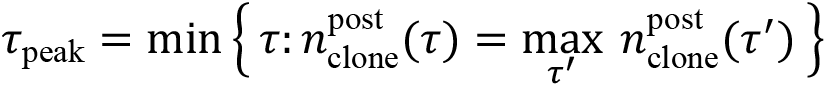

and the group’s reported composition is *S*(τ_peak_). When several candidate phylothresholds tie at the maximum clone count, the smallest is taken. This phylothreshold is the location of the clone-count peak traced in the Multi-SNP-Threshold Plot (Section 1.3.5).

When the group forms no clone at any tested phylothreshold, whether it was resolved without a scan (Section 1.3.1) or its composition consists entirely of singletons throughout the scan, the clone count is zero. In this case, no *peak* phylothreshold is defined, and the all-singleton composition is reported.

The selected phylothreshold for every group is recorded in a per-group *peak* table, and the corresponding compositions are concatenated into the dataset-wide strain table described in Section 1.3.6.

###### 1.3.6.4 Discrepancy

Let *δ*(τ) be the discrepancy at candidate phylothreshold τ, computed before phylogeny correction (Sections 1.3.3, 1.3.5), and let τ_single_ be the singleton threshold (default 100 SNPs). This endpoint places the phylothreshold where the phylogeny and the SNP distances are in closest agreement, where δ is minimized. *δ* can be low at very conserved phylothresholds, at which the strains are still too finely split for any disagreement between SNP and phylogeny to appear. Thus, the minimum is taken only after the initial discrepancy peak. The method proceeds in two phases.

First, the discrepancy peak is located within the conserved range, at phylothresholds no greater than the singleton threshold (Section 1.3.1),

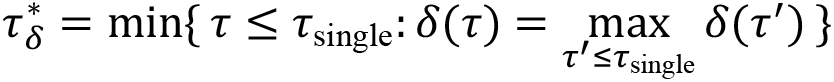

Second, the endpoint is the phylothreshold minimizing *δ* after 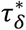,

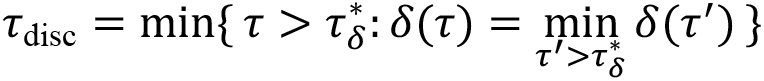

and the group’s reported strain compositions are *S*(τ_disc_). In both phases ties are resolved to the smallest qualifying phylothreshold: the discrepancy peak is the most conserved phylothreshold with the maximum discrepancy counts, and the endpoint is the smallest post-peak phylothreshold attaining the minimum, so that the strains are no larger than the concordance minimum requires. Both quantities can be read directly from the discrepancy curve traced in the Multi-SNP-Threshold Plot (Section 1.3.5, Figure 3c, *Discrepancy*).

The two-phase structure is designed based on how discrepancy behaves as the phylothreshold increases. At conserved phylothresholds, single-linkage clustering on SNP distances groups genomes (Method 1.3.2), which the phylogeny does not resolve as monophyletic. Reconciling the two requires merging these genomes into a common clade, producing high discrepancy that decreases quickly as the phylothreshold rises and the corrected strains regain monophyly. Across many groups in the published datasets, this peak occurs at highly conserved phylothresholds (MSTP in Supplementary Data 1). Low discrepancy also appears sporadically before the peak, caused by the scenarios where the strains are still split too finely for the disagreement to have emerged. The endpoint is therefore drawn only from the post-peak descent, where the minimum reflects genuine agreement between the phylogeny and the SNP distances.

When the smallest study–study SNP distance already exceeds the singleton threshold (Section 1.3.1) in the group, no phylothreshold range is scanned and the group is reported to have all-singleton composition. Otherwise, the selected phylothreshold for every group is recorded in a per-group discrepancy table, and the corresponding compositions are concatenated into the dataset-wide strain table described in Section 1.3.6.

###### 1.3.6.5 Endpoint quality control

Each endpoint encodes a different hypothesis about what separates one strain from the other, but all pursue the same balance: a phylothreshold that is neither too sensitive, merging distinct strains, nor too specific, splitting a single strain. THRESHER reaches this partition in two layers. The hierarchical clustering partitions all genomes into groups of related genomic backgrounds (Section 1.2.4), then, within each group, the strain composition is defined by the endpoint at its selected phylothreshold (Sections 1.3.6.1–1.3.6.4). To make both layers visually inspectable, THRESHER produces quality-control plot that places each pair of identified strains in two distance spaces at once, so that the separation between strains can be judged directly rather than inferred from the phylothreshold alone.

For an endpoint’s final strain composition, every pair of strains is compared by two between-strain distances, each the mean over all cross-strain genome pairs. For strains *S*_*a*_ and *S*_*b*_, the average SNP distance is

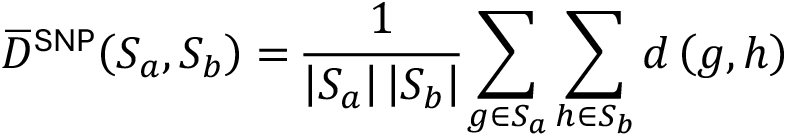

where *d*(*g, h*) is the symmetric SNP distance (Section 1.2.2), and the average phylogenetic distance is

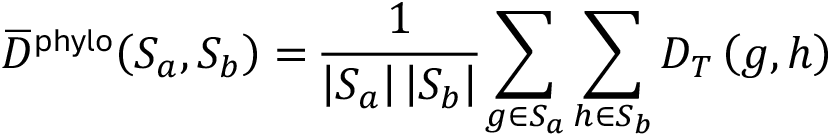

where *D*_*T*_(*g, h*) is the sum of branch lengths on the path between *g* and *h*, taken from the midpoint-rooted core-gene phylogeny (Section 1.2.1).

The two distances are plotted against each other, one point per strain pair, colored by whether the pair is within the same hierarchical clustering group, with the marginal density of each distance shown alongside. The plot shows both partition layers in a single view, and a well-resolved dataset shows a characteristic pattern. Across-group pairs (the population-structure layer) should sit at large distances, well separated from within-group pairs, confirming that hierarchical clustering placed divergent backgrounds in different groups. Within-group pairs (the endpoint layer) should be close, since they share a background, but not arbitrarily close. When within-group pairs cluster near zero distance, it flags that the endpoint has merged genomes that should have been kept in separate strains.

The plot and its underlying table are generated for all four endpoints. When an endpoint resolves the entire dataset into a single strain, no between-strain pair exists, and the comparison is reported as a message in place of the plot. For all 55 tested published datasets, the QC plots for each of the four endpoints are provided as thresher/output/QC/{endpoint}_qc_plot.pdf, with the raw tables underlying the plots in the corresponding thresher/output/QC/{endpoint}_qc_table.csv, within each study’s output directory in Supplementary Data 1.

##### 1.3.7 Methicillin-resistance status (*S. aureus*)

When the species of study is *S. aureus*, an optional step predicts the methicillin-resistance status of each genome, and of each strain, from carriage of the *mec* gene. Each study genome assembly is searched with blastX (BLAST+ ^39^) against a built-in database of thirteen reference proteins, including the penicillin-binding protein PBP2a and its homologs encoded by mecA (n = 10 variants; RefSeq WP_057521704.1, WP_063852670.1, WP_063852677.1, WP_063852683.1, WP_000721309.1, WP_000721306.1, WP_063852626.1, WP_063852617.1, WP_000721310.1, and WP_063851348.1), mecB (n = 2 variants; WP_012655867.1 and WP_063852710.1), and mecC (n = 1 variant; WP_000725529.1). Those genes encode the proteins that confer β-lactam resistance in staphylococci by substituting for the native penicillin-binding proteins^40-42^. For each genome, the best hit (highest bitscore) to each reference protein is retained, and the coverage of a retained hit is the aligned length as a fraction of the reference protein amino acid length. A genome is called MRSA when at least one retained hit passes 70% coverage and 70% identity thresholds.

#### 1.4 EPI (epidemiological step)

When the metadata table supplies a patient identifier and a collection date for every genome, THRESHER runs an optional epidemiological step that links the strains identified by CORE to the patients they were sampled from and reports the strains shared across patients as transmission clusters. EPI utilizes the single dataset-wide strain composition produced by the selected endpoint method (Section 1.3.6. *plateau* endpoint, plateau length: 15 by default), as well as the metadata and, where available, the MLST assignments and, for *S. aureus*, methicillin-resistance status. When patient identifiers or collection dates are incomplete, this step is skipped and THRESHER fallbacks to the strain composition alone (Section 1.1). Each stage is described below.

##### 1.4.1 Transmission cluster identification

A clone is reported as a transmission cluster when its genomes collected from more than one patient. Its persistence is summarized by the first and last collection dates among its genomes. When no strain spans more than one patient, no transmission cluster is reported.

##### 1.4.2 Transmission Cluster Plot

The members of each transmission cluster is drawn as a separate panel. A rounded card carrying one marker per patient, labelled with the patient identifier, above a header that gives the cluster identifier and the sequence type. For *S. aureus*, a resistance badge marks the cluster as resistant (R, MRSA) or susceptible (S, MSSA) to methicillin based on the blastX results (Section 1.3.7). The plot visualizes which patients share the transmission cluster, and because it shows patients rather than genomes, a patient who contributed several genomes to a cluster appears only once. Plots were generated with R package ggplot2^43^.

##### 1.4.3 Cluster Persistence Plot

The persistence of the clusters is drawn as a single timeline, with collection month on the horizontal axis and cluster on the vertical axis. Each genome is plotted as a point at its collection date and labelled with its patient identifier, and the points belonging to one cluster are joined along that cluster’s row, so the horizontal extent of each row is the cluster’s persistence and the points along it show how its sampling was distributed in time. In contrast to the Transmission Cluster Plot, a patient who contributed genomes on several dates appears as several points along the row. When no transmission cluster is present, both plots are replaced by a single panel reading *No Transmission Cluster Found*. Plots were generated with R package ggplot2^43^.

Both plots require the optional EPI stage, which is enabled only when patient identifiers and collection dates are supplied. Among the 55 re-analyzed studies, 16 deposited metadata from which patients and collection dates could be identified or inferred. For these studies, the transmission cluster plots are provided in Supplementary Data 3 at <StudyID>/ClusterPlots/Cluster_{#}.pdf, one file per cluster, and the persistence plot at <StudyID>/ClusterPersistencePlot/PersistencePlot.pdf. Each is accompanied by the corresponding RDS object so users can regenerate or modify the figure locally. A more complete demonstration of both plots is given in our previous study of *S. aureus* transmission in a neonatal intensive care unit^16^.

#### 1.5 Modes of THRESHER

THRESHER provides five modes that share the same underlying algorithm but vary in the input and output. So that the THRESHER results can be reused, re-examined, extended, or re-parameterized without re-running the whole analysis. Full Pipeline (Section 1.5.1) is the standard entry point that runs through the complete workflow described in Sections 1.1–1.4. The remaining four modes each take the output of a completed Full Pipeline (or New Full) run as their starting point: Cladebreaker OFF (Section 1.5.2) recomputes strain compositions with the CladeBreaker constraint disabled; Redo Endpoint (Section 1.5.3) requires full metadata including the complete patient identifiers and collection dates, and reanalyzes the cluster composition with the new endpoint method; New SNPs (Section 1.5.4) places additional genomes into the existing strains and transmission clusters under the phylothresholds already determined; and New Full (Section 1.5.5) adds genomes and re-derives the phylothresholds from the combined dataset. The modes differ principally in whether they hold the inferred phylothresholds fixed or recompute them, and in how much of the upstream pipeline they repeat.

##### 1.5.1 Full Pipeline

Full Pipeline runs the complete analysis from genome assemblies with no prior THRESHER output (Sections 1.1–1.4). Full mode determines strains and their visualizations, adding transmission clusters when patient identifiers and collection dates are supplied, while lite mode determines strains only. For every group it records the strain composition at each candidate phylothreshold and selects a final phylothreshold under all four endpoint methods, so the *plateau, peak, discrepancy*, and *public* compositions are produced in a single run and reused by the four modes below.

##### 1.5.2 Cladebreaker OFF

CladeBreaker (Section 1.3.4) assumes the study genomes are hyperlocal strains absent from public databases, so a public genome should not be within the “same” strain with a study genome, which restricts strain size. When the user instead has evidence that the matched public genomes belong to the same strains, Cladebreaker OFF re-runs identification with the CladeBreaker step disabled, allowing public genomes to cluster with study genomes and avoiding the error of inflating the number of strains. It reuses the SNP distances and phylogenies from the original run, re-executing only the phylothreshold scan and endpoint selection.

##### 1.5.3 Redo Endpoint

Redo Endpoint revisits the endpoint choice without rerunning the core algorithm. Using the intermediate files from a completed run together with the original complete metadata, it reanalyzes the transmission cluster under a different endpoint method without repeating any earlier stage, so the cluster analysis can be moved from one endpoint method to another at minimal cost.

##### 1.5.4 New SNPs

New SNPs adds genomes by reusing the previously inferred phylothresholds. The user provides the metadata of the new genomes, which THRESHER combines with the original metadata and the prior output to compute the pairwise SNP distances between the new and existing genomes. Each new genome is then assigned to the existing strains by single-linkage clustering. The existing strains are considered in turn. For each, the phylothreshold of that strain’s hierarchical clustering group is applied, and a new genome joins the strain when its SNP distance to a member of that strain falls at or below that phylothreshold. When that strain’s group has no defined phylothreshold, the phylothreshold of the nearest group that has one is used instead. After all existing strains have been considered, the new genomes that have joined none of them are grouped among themselves by SNP-based single-linkage clustering (Section 1.3.2) under the same rule, forming a new strain wherever a pair of new genomes falls at or below the applicable phylothreshold. When patient identifiers and collection dates are provided, transmission-cluster identification (Section 1.4.1) is rerun to update the transmission clusters. Because strain compositions are updated without rebuilding the phylogenies, repeating hierarchical clustering, or re-inferring the phylothresholds, the mode is fast and suited to adding a few genomes that are unlikely to alter the dataset’s population structure. Its output records, for each genome, whether it is original or new and the strain and cluster it joins.

##### 1.5.5 New Full

New Full also adds genomes but re-derives the phylothresholds from the combined dataset. It annotates the new genomes, regenerates the core-gene alignment through pan-genome analysis, re-infers the core-gene ML phylogeny, repeats hierarchical clustering, and re-runs the CORE algorithm and endpoint selection on all genomes together, so both the phylothresholds and the strain compositions reflect the full set. It is slower than New SNPs but appropriate when many genomes are added or the population genomic structure may change substantially. Its outputs match the Full Pipeline format and are themselves a valid starting point for any downstream mode, including further New Full runs, with an additional table summarizing how strain and cluster composition changed.

#### 1.6 THRESHER-Chat

Running THRESHER, interpreting its outputs, and troubleshooting, optimizing, or extending the analysis as well as the underlying code and algorithm, which may later be maintained by other students or researchers, all depend on a combined understanding of the algorithm, the parameters governing each analytical step, the underlying biological hypotheses, and the structure of the tool and its code, which is a demand that varies with the user’s background and is challenging and time-consuming for most. In addition, THRESHER is aimed largely at clinical settings investigating the transmission of bacterial pathogens, where much of the data is patient-sensitive and uploading externally may violate regulations such as HIPAA. To support users across levels of expertise without any data leaving their system, we provide THRESHER-Chat^44^, a retrieval-augmented assistant that runs entirely locally. It indexes THRESHER’s documentation and source code into a vector store and answers natural-language questions by retrieving the most relevant passages and supplying them, together with the question, to a locally served language model. The language and embedding models (served through Ollama [https://github.com/ollama/ollama]), the vector store (ChromaDB [https://github.com/chroma-core/chroma]), the retrieval pipeline (LangChain [https://github.com/langchain-ai/langchain]), and the conversational memory (SQLite [https://www.sqlite.org/]) all run on the user’s own system, so neither uploaded files nor patient-associated metadata leaves the local system.

THRESHER-Chat installs in a single step and is operated through a terminal interface and a user-friendly local web interface. Its workflow is visualized in Supplementary Figure, which comprises three components: knowledge base construction (Section 1.6.1), retrieval-augmented question answering (Section 1.6.2), and personalization through the user profile and memory (Section 1.6.3). A screenshot of the web interface is shown in Supplementary Figure 6.

##### 1.6.1 Knowledge base construction

THRESHER-Chat indexes the THRESHER code repository together with a set of curated guides shipped with the assistant. Source files (Python, R, and Snakemake) and Markdown documentation are loaded and split into chunks. Each chunk is embedded with an embedding model (EmbeddingGemma^45^ by default) and stored in a persistent ChromaDB vector store, which is built once and reused across conversation sessions. The curated guides hold only rationale and interpretation guidance, while the repository documentation supplies command syntax, inputs, and outputs, keeping the two complementary.

##### 1.6.2 Retrieval-augmented question answering

For each question, the most similar chunks are retrieved from the vector store (Section 1.6.1) and supplied as context to a locally served chat model (Gemma 4^46^ E4B by default, user-selectable). A system prompt constrains the model to answer only from the retrieved documentation, to cite the source file, to decline questions outside THRESHER’s scope, and to ask for clarification rather than guess when a question is ambiguous, which guards against prompt injection. Because the assistant routinely returns commands and code, system prompt directs to copy commands, paths, and code exactly as written. The chat model is run deterministically with temperature zero to maximize the answer fidelity. Both the language model and the embedding model are selected by the user, and the quality of the responses depend on these choices. The answers should be verified against the primary documentation.

##### 1.6.3 User profile and memory

Two approaches adapt responses to the user. First, the user may select an audience profile such as graduate student, principal investigator, clinician, bioinformatician, statistician, or infection-prevention-and-control team. Each contributing a short prompt that sets the depth and framing of the answers. In addition, profiles are defined in an editable file, so users can add their own. Second, a single-user memory backed by SQLite and stored alongside the vector store persists across sessions, retaining the conversation history and a compact record of the user, which contains short profile tags inferred from the dialogue (which the user can correct or reject), stated analytical objectives, and brief summaries of past sessions, all generated by the same local model. The file storing user’s profile and memory is portable: the user can copy or move it to another machine and resume with the existing memory. The selected profile and the current memory are added to the model’s input for answering, while document retrieval and the stored history use the user’s original question, which keeps retrieval focused and prevents the injected context from re-entering memory.

### 2. Simulation

The simulated *S. aureus* populations with a known evolutionary history was generated with SILOSim^20^, an integrated framework that learns a per-site map of mutation entropy and mobile genetic elements from real, closely related genomes and then simulates forward along a time-scaled phylogeny. The procedure comprised three stages: assembling a species-wide backbone phylogeny and selecting ancestral seed genomes (Section 2.1), profiling each seed (Section 2.2), and simulating evolution forward along dated phylogenies (Section 2.3). The backbone was constructed for this study only. The SILOSim profiler and simulator, including the command-line options, algorithmic detail, and usage, are described in full in the publication introducing SILOSim^20^. Here we summarize the steps relevant to generating the validation populations (Figure 4).

#### 2.1 Backbone

By April 2024, 91,980 publicly available *S. aureus* genome assemblies were downloaded from NCBI. Each was screened with CheckM v1.2.2^47^, MLST^36^, and Mash version 2.3^48^ and retained only if it was placed within the genus Staphylococcus (CheckM lineage marker set UID301) with completeness ≥ 95% and contamination ≤ 5%, had genome size and GC content each within two standard deviations of the dataset mean (2,861,903 ± 121,283 bp and 32.71 ± 0.19%, respectively), passed the Mash screen, and was not typed as ST2250 or ST1223. Both sequence types were originally grouped with S. aureus but are now recognized as *S. argenteus*. In total, 68,299 assemblies passed quality control and were carried forward to dereplication.

Within each sequence type (ST), genomes were dereplicated with dRep v3.4.5^49^ (--ignoreGenomeQuality --multiround_primary_clustering --greedy_secondary_clustering --S_algorithm fastANI -pa 0.975 -sa 0.995), retaining representatives at a 99.5% ANI threshold and yielding 1,830 representatives across 65 STs.

Representatives were annotated with Bakta v1.11.4 (full database v6.0)^50^, and a core-gene alignment was generated with Panaroo v1.5.1^51^ (--remove-invalid-genes --clean-mode strict --alignment core --core_threshold 0.95 --aligner prank --family_threshold 0.7 --refind_prop_match 0.5 --search_radius 5000). A maximum-likelihood phylogeny was inferred from this alignment with IQ-TREE v2.3.6^28^ using the GTR+R model (-m GTR+R --ufboot 1000 --bnni -alrt 1000; 1,000 ultrafast bootstrap and 1,000 SH-aLRT replicates; Supplementary Figure 3).

Five genomes were selected as ancestral seeds for the simulations. Seeds were chosen to span the phylogeny rather than concentrate within few clades, representing the diversity of the species, while each retained sufficient depth (the number of genomes being represented; the n_genomes value in Supplementary Table 5) for accurate and informative entropy and mobile genetic element (MGE) inference. The five seeds with their STs and depths were GCA_010603155.1 (ST8, depth 12,034), GCA_010599565.1 (ST5, depth 10,199), GCA_003970295.1 (ST22, depth 7,323), GCA_027082625.1 (ST398, depth 3,763), and GCA_024213975.1 (ST1, depth 3,053). Of the 1,830 representative genomes, only 64 carried a depth greater than one, which represented additional genomes assigned to the same secondary cluster, while the remaining 1,766 represented only themselves. These 64 representatives, together with their secondary_cluster, ST, and n_genomes (the number of genomes each represented), are summarized in Supplementary Table 5. The phylogeny and the positions of the five seeds are shown in Supplementary Figure 3. ST assignments and the metadata of all 1,830 representatives are provided in Supplementary Table 6.

#### 2.2 Profiling

For each of the five seeds, the genomes it represents in the backbone dereplication served as the closely related query genomes. Because secondary clustering applied a 99.5% ANI threshold (Section 2.1), every query genome is at least 99.5% identical to its seed, ensuring sufficient relatedness for reliable profiling (PROFILING panel, Figure 4). Each draft seed assembly was first concatenated into a single sequence, retaining a map to the original contigs, and every query genome was aligned to the seed with nucmer in MUMmer v4.0.0rc1^31^. Alignment coordinates were obtained with show-coords and substitutions with show-snps, the latter restricted to uniquely aligned regions and to SNPs at least 20 bp from the nearest mismatch (the buffer-distance criterion applied by MUMmer’s dnadiff), so that poorly aligned SNP calls do not enter the analysis.

For a seed, let *i* index the positions of its concatenated sequence. The coverage depth *c*_*i*_ is the number of query genomes whose alignment covers position *i*, and serves as the denominator for the per-site entropy. Let *r*_*i*_ be the seed allele at *i*. A query genome with no SNP called at position *i* carries *r*_*i*_. These genomes carrying *r*_*i*_, together with the seed itself (which contributes one observation of its own allele), form the reference-allele count, while each alternate allele *a* is counted over the query genomes that carry it. Let reference-allele count be *y*_*i,r*_*i*:

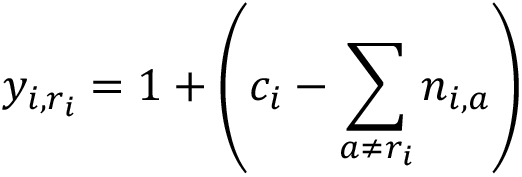

Let alternate-allele counts be *y*_*i,a*_:

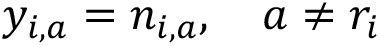

From these counts, indexing the alleles observed at position *i* by *k* (the seed allele together with any alternates), the maximum-likelihood allele frequencies were shrunk toward a uniform target *t*_*k*_ over the alleles observed at the site *i* using the James–Stein-type shrinkage estimator of Hausser and Strimmer, as implemented in the entropy() function of the R package entropy^52^, with a data-driven intensity *λ*_*i*_:

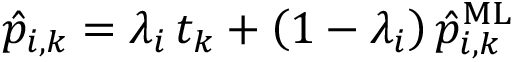

The per-site mutational entropy *H*_*i*_ is the Shannon entropy of the shrunken frequencies,

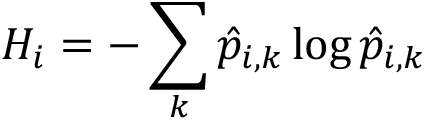

Positions with no observed substitution were assigned zero entropy. The resulting profile {*H*_*i*_} scores every position of the seed’s concatenated sequence by its variability across the query genomes and weights the placement of substitutions during simulation (Section 2.3).

Mobile and accessory regions were inferred from localized drops in coverage. Coverage was first normalized to its genome-wide maximum, taken over all positions *j* of the concatenated sequence,

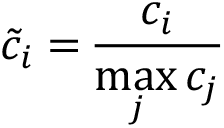

and a mobile genetic element (MGE) was defined as a maximal run, spanning positions *s* to *e*, of at least 100 bp and confined to a single original contig, over which normalized coverage falls below a cutoff *κ* (default 0.7):

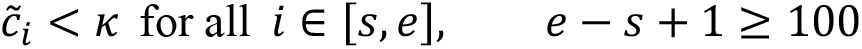

Each element was annotated with the coding sequences it overlaps, taken from the seed’s Bakta annotation (Section 2.1), and its nucleotide sequence was extracted from the concatenated assembly. Across a seed, these elements constitute the pool from which gene gain is drawn during simulation (Section 2.3). Per-site coverage and entropy were additionally summarized over bins defined by the seed’s annotated coding sequences and the intervening intergenic regions (Supplementary Data 4).

#### 2.3 Simulation

The profiling outputs of the five seed genomes (Section 2.2) provided the inputs to the simulator. Evolution was simulated forward in time along a time-scale phylogeny with a tool adapted from CoreSimul^53^, modified and extended in three aspects: substitutions are placed according to the per-site entropy profile rather than uniformly, candidates of gene gain and loss simulation draws from the seed’s empirically detected MGE pool, and recombination admits both within-sample and external donors (SIMULATION panel, Figure 4).

For each seed, a time-scale phylogeny was generated under a birth–death process with TreeSim^54^, with the number of tips, the time span in years, and the speciation and extinction rates λ and μ set by the user (5 years; λ = 1; μ = 100). Because the random seed fixes the topology and branch lengths, the same three phylogenies were generated once and reused across all five seeds, holding the evolutionary trajectory constant while only the seed-specific entropy and MGE profiles varied (Supplementary Figure 1).

The unit of branch lengths is years, and each tree was partitioned into successive time intervals defined by the set of lineages co-occurring over a span Δ*t*. Within an interval, every active sub-lineage independently accumulated substitutions, recombination events, and events of gene gains and losses. The number of each kind of event was drawn from a Poisson distribution whose mean is the product of the corresponding per-year rate *r*_event_ and Δ*t*, and for substitutions the mean additionally scales with the lineage’s valid time-scale length *L*,

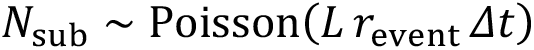

Because gene gain, gene loss, and recombination change a descendant’s sequence length relative to its ancestor, inserted and deleted regions were represented as gap characters across all lineages so that the evolving sequences remained mutually aligned (Section 2.3, below). All events generated across an interval were then applied to those sequences in random order.

Each substitution was placed and resolved in two steps. First, its position was drawn across the genome with probability proportional to the per-site entropy *H*_*i*_ (Section 2.2),

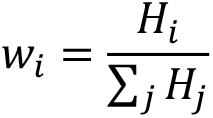

so that hypervariable sites change more often than conserved ones. Sites with non-positive entropy were assigned the smallest positive entropy value so that every site retains a non-zero chance of mutation, and positions already removed by a deletion were skipped. The replacement base was then drawn under the chosen nucleotide model, including JC69, K2P, K3P, and GTR, with each change weighted by the model’s exchangeability *q* for that pair,

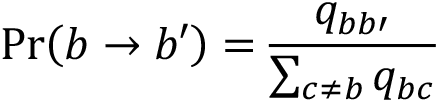

Here the GTR model was used, with the six exchangeabilities fixed to values estimated from a previously published *S. aureus* dataset from our group^16^. The entropy profile therefore governs where substitutions are calculated to locate, and the substitution model governs which base substitutes with.

Recombination replaced a contiguous segment of the recipient. The number of events *N*_recom_ per sublineage followed the same Poisson form,

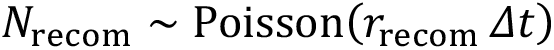

and each segment’s length was drawn from an exponential distribution with a user-set mean and floored at a minimum size (default 1bp),

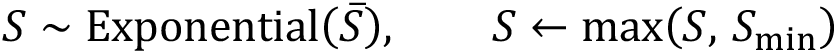

Each event was independently internal or external with equal probability. An internal event, applicable only where more than one sub-lineage co-occurs in the interval, drew a donor from the other co-occurring sub-lineages and copied its segment directly, so the substitutions introduced were exactly the donor–recipient differences over that segment. This represents the exchange of DNA between co-circulating sub-lineages in close proximity. An external event represented an unsampled donor more distantly related to the simulated sub-lineages. Typical external recombination often presents segment of the same length but with substitutions. To reproduce the elevated divergence such distantly related donor segments carry, the de novo user-defined density ? (SNPs per site) is introduced to allocate the substitutions across the recombinants. Here *v* sets the per-site rate at which substitutions are placed within the recombinant segment, governing how many SNPs the external donor introduces over its valid (non-gap) length *S*_*valid*_.

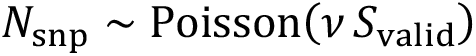

Gene gain and loss acted on the bins defined during profiling (Section 2.2), with counts again Poisson-distributed,

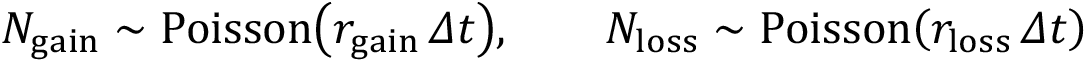

A gain inserted a sequence drawn at random from the seed’s MGE pool into a randomly chosen intergenic bin, and a loss removed an MGE-overlapping bin in full. To keep all descendants mutually aligned, an insertion was recorded as gap characters of equal length in every other lineage and a loss as gap characters spanning the removed bin, so the output remains a multiple sequence alignment of equal-length sequences. As the gene gain and loss proceeded, the per-site entropy table was updated in step with each insertion.

All rates and model parameters were taken from a previously published *S. aureus* genomic dataset of neonatal intensive-care genomes^16^. The substitution rate (8.5 × 10^−6^ substitutions per site per year) was the median per-clone molecular-clock rate estimated with BEAST v2.7.6^55^ across longitudinally persistent clones with at least four genomes each, under an HKY model and a strict clock with a constant-population coalescent prior (Supplementary Figure 7). The six GTR exchangeabilities were estimated from the same dataset and normalized to sum to one. In the order expected by the simulator (A↔G, A↔C, A↔T, G↔C, G↔T, C↔T), these were 0.289, 0.081, 0.082, 0.046, 0.069, and 0.433, respectively. Gene gain and loss rates (0.934 and 0.863 events per year) were obtained by tracking 7,352 variably present genes across 92 clones from the same dataset mentioned above. Each gene’s gain or loss was timed from the inferred emergence of the clone to the first genome in which the gene appeared or disappeared, and these waiting times were fit to an exponential distribution whose rate set the Poisson rate (Supplementary Figure 8). The recombination rate (4.34 events per year), mean segment length (76.34 bp), and SNP density (*v* = 0.03) were estimated with ClonalFrameML^56^ on the same clones, as the slope of recombination events against persistence, the mean of an exponential fit to inferred segment lengths, and the mean per-clone *v*, respectively (Supplementary Figure 9).

Each run produced whole-genome sequences for every tip and internal node of its time-scale phylogeny, together with logs of all substitution, recombination, and gain and loss events. The three phylogenies applied to each of the five seeds yielded 660 simulated genomes in total.

### 3 Benchmarking on simulated populations

The simulated populations of Section 2 carry a known evolutionary trajectory, providing a defined answer that can validate inferred strain structure. We used them to benchmark THRESHER against fixed SNP cutoffs under incomplete sampling, defining the ground-truth transmission clusters (Section 3.1), the subsampling and comparison design (Section 3.2), and the metrics (Section 3.3).

#### 3.1 Defining a transmission cluster

We placed a boundary at Year 3 of each 5-year time-scaled phylogeny and, on each sub-lineage, took the first internal node at or beyond that boundary, whose parent node still precedes Year 3. Each such node is the most recent common ancestor of a distinct transmission cluster, comprising that node and all of its descendants, whether internal nodes or tips. A genome whose most recent coalescence predates Year 3 was designated a singleton (Supplementary Figure 1). Internal-node genomes older than Year 3 define no transmission cluster of interest and were excluded from the sensitivity and specificity calculations (Section 3.3).

#### 3.2 Subsampling and comparison design

From the 660 simulated genomes we drew random subsamples at 20%, 40%, 60%, 80%, and 100%, with 10 replicates per rate, for 50 subsampled populations in total. We subsequently split each subsampled population by sequence type, where we pooled genomes that share an ST but could originate from different time-scaled phylogenies. This pooling yielded 250 per-ST groups.

We evaluated fixed cutoffs of 0, 10, 20, 30, 40, and 50. For THRESHER we tested the *plateau, peak*, and *discrepancy* endpoints. The *public* endpoint does not apply over these short timescales, where each simulated genome’s closest match is its own ancestral seed. For the *plateau* endpoint we used window lengths of 5 and 15, giving four THRESHER method configurations in total. For each of the 250 groups we visualized the assignments from every method alongside the simulated ground truth, pairing a matrix of method-specific cluster identifiers with the group maximum-likelihood phylogeny and a SNP-distance heatmap (Supplementary Data 1).

To test whether the inferred phylothreshold depended on genetic background, we compared the five sequence types within each endpoint method at each subsample rate, giving 20 strata (4 endpoint methods × 5 sampling rates), and each stratum contains 50 observations (5 sequence types × 10 replicates). Within each stratum we applied a Kruskal–Wallis rank sum test. The 100% subsample rate was excluded from this analysis. Benjamini–Hochberg correction was applied across the 16 informative strata (4 endpoint methods at 100% subsample rate excluded) at rates 0.2–0.8. Results for all 20 strata, together with per-sequence-type medians and the magnitude of the sequence-type effect relative to the sampling-rate effect, are reported in Supplementary Table 6.

#### 3.3 Sensitivity and specificity

Each simulated genome carries its ground-truth, which is its transmission-cluster identity or singleton from Section 3.1. For every method, a method-specific label given by the fixed cutoff or the THRESHER endpoint under evaluation. Before computing metrics we restricted each comparison to an informative set of genomes: we excluded genomes called singletons by both the ground truth and the test method, and we excluded internal-node genomes predating Year 3, which define no transmission cluster (Section 3.1).

Let the retained set contain *n* genomes, and let *g*_*i*_ and *g*_*j*_ denote two distinct genomes within it. Let *R* be the ground-truth label and let *M* be the label produced by the test method, so that *R*(*g*) and *M*(*g*) are the ground-truth and test labels of a genome *g*. For each unordered pair, let 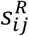 equal 1 if *R*(*g*_*i*_) = *R*(*g*_*j*_) and 0 otherwise, and 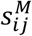 equal 1 if *M*(*g*_*i*_) = *M*(*g*_*j*_) and 0 otherwise. We treated every pair as a binary classification, where a true positive (TP) is a pair with 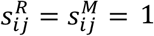(assigned the same strain by both); a true negative (TN), a pair with 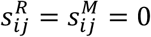(different strains in both); a false positive (FP), a pair the test groups but the ground truth separates 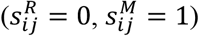; and a false negative (FN), a pair the ground truth groups but the test separates 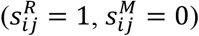. Sensitivity was calculated as,

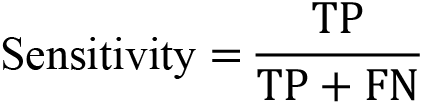

while specificity was calculated as,

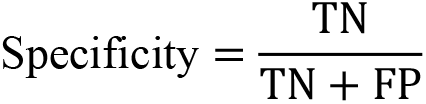

### 4 Concordance with published datasets

We next compared THRESHER to the assignments reported in the original publications of the 55 published studies introduced in Figure 1, using the *plateau* endpoint at window length 15 throughout.

#### 4.1 CladeBreaker application

We enabled or disabled CladeBreaker (Section 1.5.2) per study according to whether its matched public genomes were judged to represent the same strains as the study genomes. Of the 55 studies, 38 were analyzed with CladeBreaker enabled and 17 with it disabled; CladeBreaker was disabled for S19, S35, S48, and S53 (*S. aureus*); K10 (*K. pneumoniae*); and C2–C7, C9, C11, and C16–C19 (*C. difficile*) (Supplementary Table 3).

#### 4.2 Quantifying concordance with originally reported assignments

Unlike the simulated populations, the published studies provide no ground truth, so we quantified concordance between THRESHER and the originally reported strain assignments. The metrics mirror those of Section 3.3, with two adaptations. First, the reference labels are the strains reported in each original study, and genomes the original study left unassigned were each treated as a distinct singleton strain. Second, no Year-3 exclusion applies. However, genomes called singletons by both the original study and THRESHER were still removed before computing metrics.

On the retained genomes we applied the identical pairwise framework: a true positive is a genome pair assigned the same strain by both the original study and THRESHER, a true negative a pair assigned different strains by both, a false positive a pair THRESHER groups but the original study separates, and a false negative a pair the original study groups but THRESHER separates. Adjusted Rand index were computed with the original assignment in place of the ground-truth partition. Let *n*_*ij*_ be the number of genomes shared by originally reported strain *i* and THRESHER strain *j, a*_*i*_ and *b*_*j*_ be the corresponding strain sizes, and *n* be the number of retained genomes, and *E* be the index expected by chance.

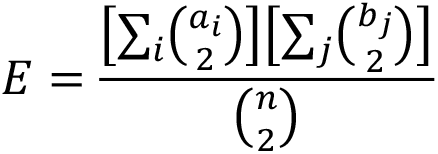

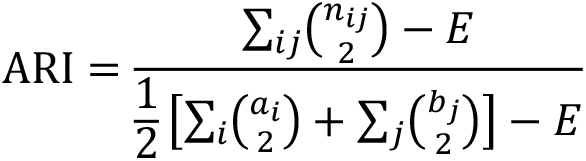

Genome-level disagreement was likewise summarized as over- and underestimation. Within each THRESHER strain, genomes not in its majority study strain were underestimated, placed by the original study in a smaller strain than by THRESHER. Within each study strain, genomes not in its majority THRESHER strain were overestimated. High values therefore indicate that the original study agreed with THRESHER and low values that it diverged. However, note that they do not establish that either assignment is ground truth.

## DATA AVAILABILITY

Raw sequencing reads and/or genome assemblies for the 55 published studies were downloaded from NCBI. The corresponding BioProject accessions are listed in the Accession(s) column of Supplementary Table 1 along with the study metadata. The THRESHER input and output files supporting validation and replication of the results for all 55 studies are provided in Supplementary Data 1, with each study in its own folder. The SILOSim Profile output files used as input for the subsequent simulations are provided in Supplementary Data 4, and the 660 simulated genomes are provided in Supplementary Data 5. The three time-scaled phylogenies are intermediate outputs of the evolution simulation. Those phylogenies and the 50 subsampled populations can be regenerated exactly using the random seeds and parameter sets included in the analysis code (see Code Availability).

## CODE AVAILABILITY

THRESHER is available at https://github.com/microbialARC/THRESHER and SILOSim at https://github.com/microbialARC/SILOSim. THRESHER-Chat, the accompanying locally deployed retrieval-augmented assistant is publicly available at https://github.com/microbialARC/THRESHER-Chat.

## ACKNOWLEDGEMENTS

This work was funded and supported by The Center for Microbial Medicine at the Children’s Hospital of Philadelphia. P.J.P, A.M.M., and J.P.Z. were supported by NIH grant 1R01AI185544. A.M.M. and J.P.Z. were supported by NIH grant U19AI174998. J.P.Z. was supported by Burroughs Wellcome Fund Investigators in the Pathogenesis of Infectious Disease Award 1356736 and NIH grant R35GM138369. Q.S. was supported by the Chappell Culpeper Family Foundation Fellowship in the Center for Microbial Medicine at the Children’s Hospital of Philadelphia.

## AUTHOR CONTRIBUTIONS

Q.S. performed formal analysis, developed conceptualization, methodology and analytical methodology (code and pipelines), created visualizations, and wrote the original draft. All authors reviewed and approved the final manuscript. E.T., and A.A., contributed to conceptualization, validation and methodology. J.P.Z., A.M.M., and P.J.P. contributed to conceptualization, methodology, formal analysis, supervision, funding acquisition, validation, and writing (original draft and review & editing).

