## Supplementary Figure 1 for "THRESHER: A tool for evidence-based, phylogeny-corrected SNP thresholds for whole genome molecular epidemiology"

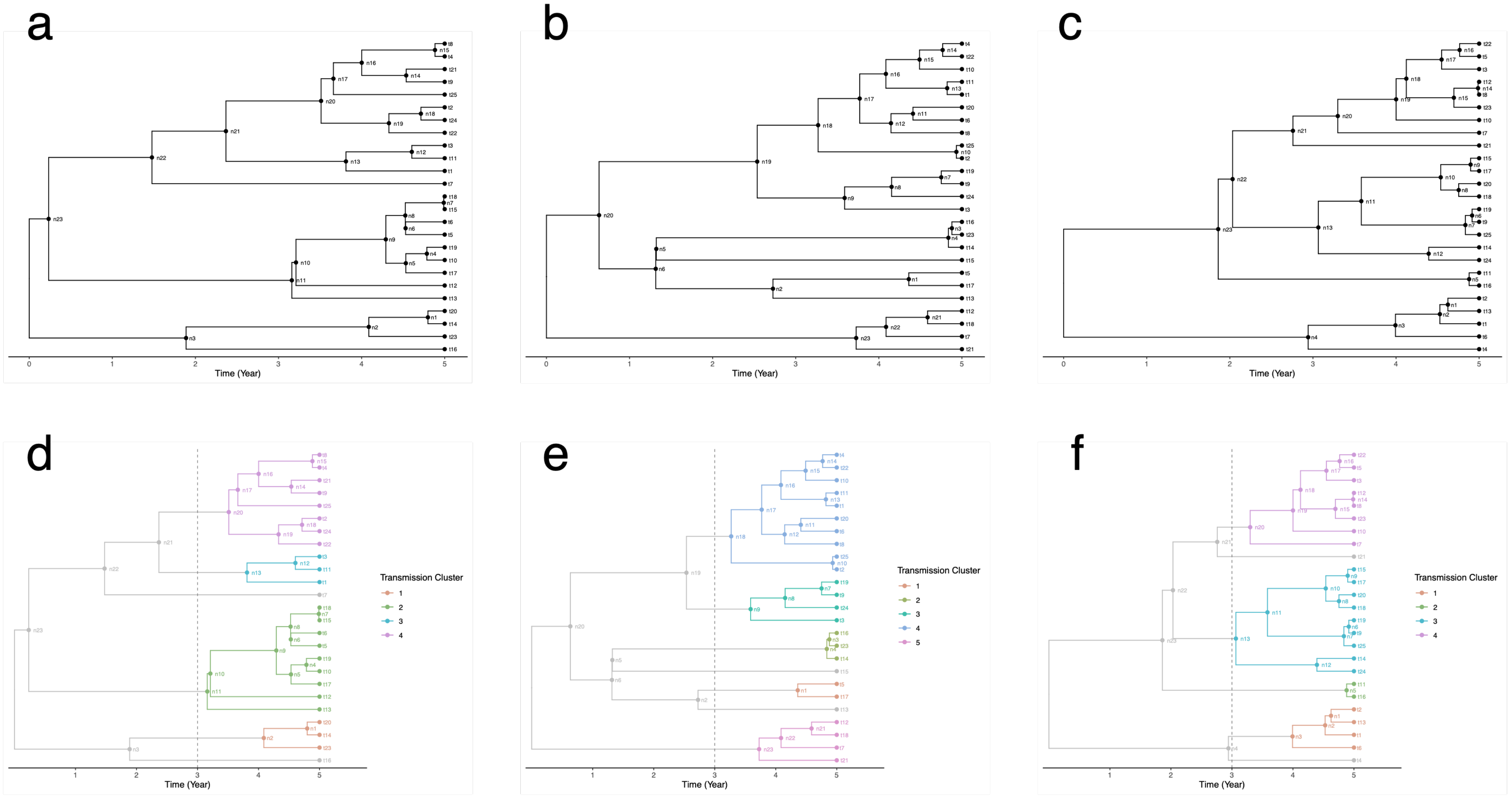


**Supplementary Figure 1. Ground-truth transmission clusters in the simulated phylogenies.**

(a–c) Three time-scaled phylogenies labeled with internal nodes (n#) and tips (t#). Each tree spanned 5 years, along which genome evolution is simulated (SIMULATION panel, Figure 4; Methods). (d–f) Ground-truth transmission clusters for the phylogenies in a–c, respectively. A boundary is placed at Year 3 (dashed line), and each of the most ancestral internal nodes after this boundary defines a distinct transmission cluster comprising that node and all of its descendants (colored). Genomes whose most recent coalescence predates Year 3 are designated singletons (gray). Clusters are numbered independently within each phylogeny.
