## Supplementary Figure 2 for "THRESHER: A tool for evidence-based, phylogeny-corrected SNP thresholds for whole genome molecular epidemiology"

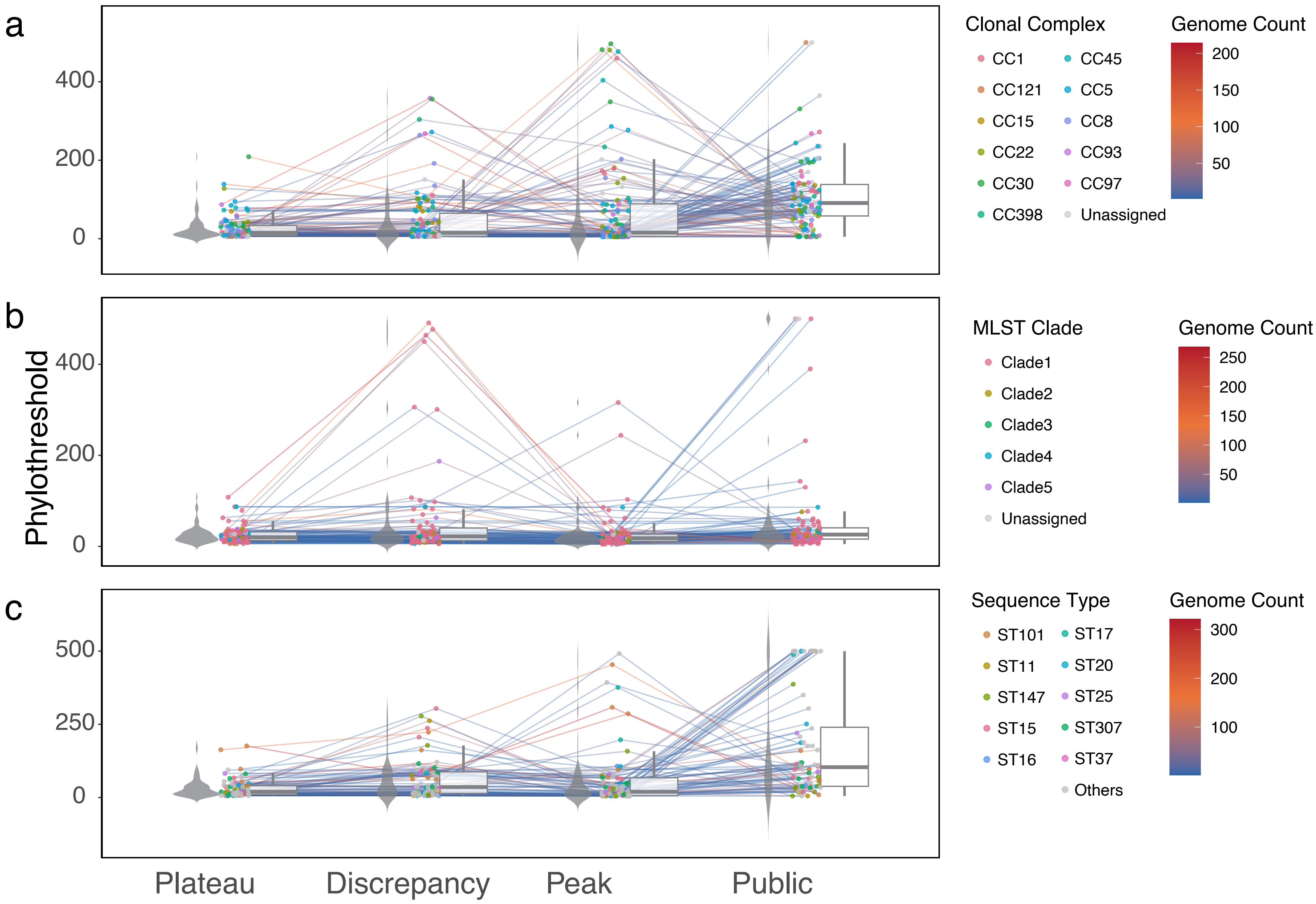


**Supplementary Figure 2. Phylothresholds by endpoint method across the re-analyzed published studies**.

Phylothresholds inferred by the *plateau*, *discrepancy*, *peak*, and *public* endpoints for hierarchical groups from studies of (a): *S. aureus* (n = 212 groups), (b): *C. difficile* (n = 161), and (c): *K. pneumoniae* (n = 151). For each study, phylothresholds are taken from the run with the CladeBreaker setting used for that study, and only groups with a phylothreshold at all four endpoints are shown. Each point is one group, jittered horizontally and colored by the most common genetic background among its genomes. Clonal Complex (CC) for *S. aureus*, MLST clade for *C. difficile*, and Sequence Type (ST) for *K. pneumoniae*, with the ten most frequent STs shown and the rest grouped as Others. Groups without an assigned background are labeled Unassigned. Lines connect each group's phylothresholds across endpoints and are colored by the number of genomes in the group. Violins (left of each endpoint) show the distribution of phylothresholds; box plots (right) show the median (center line) and interquartile range (box), with whiskers extending to the most extreme values within 1.5× the interquartile range.
