## Supplementary Figure 3 for "THRESHER: A tool for evidence-based, phylogeny-corrected SNP thresholds for whole genome molecular epidemiology"

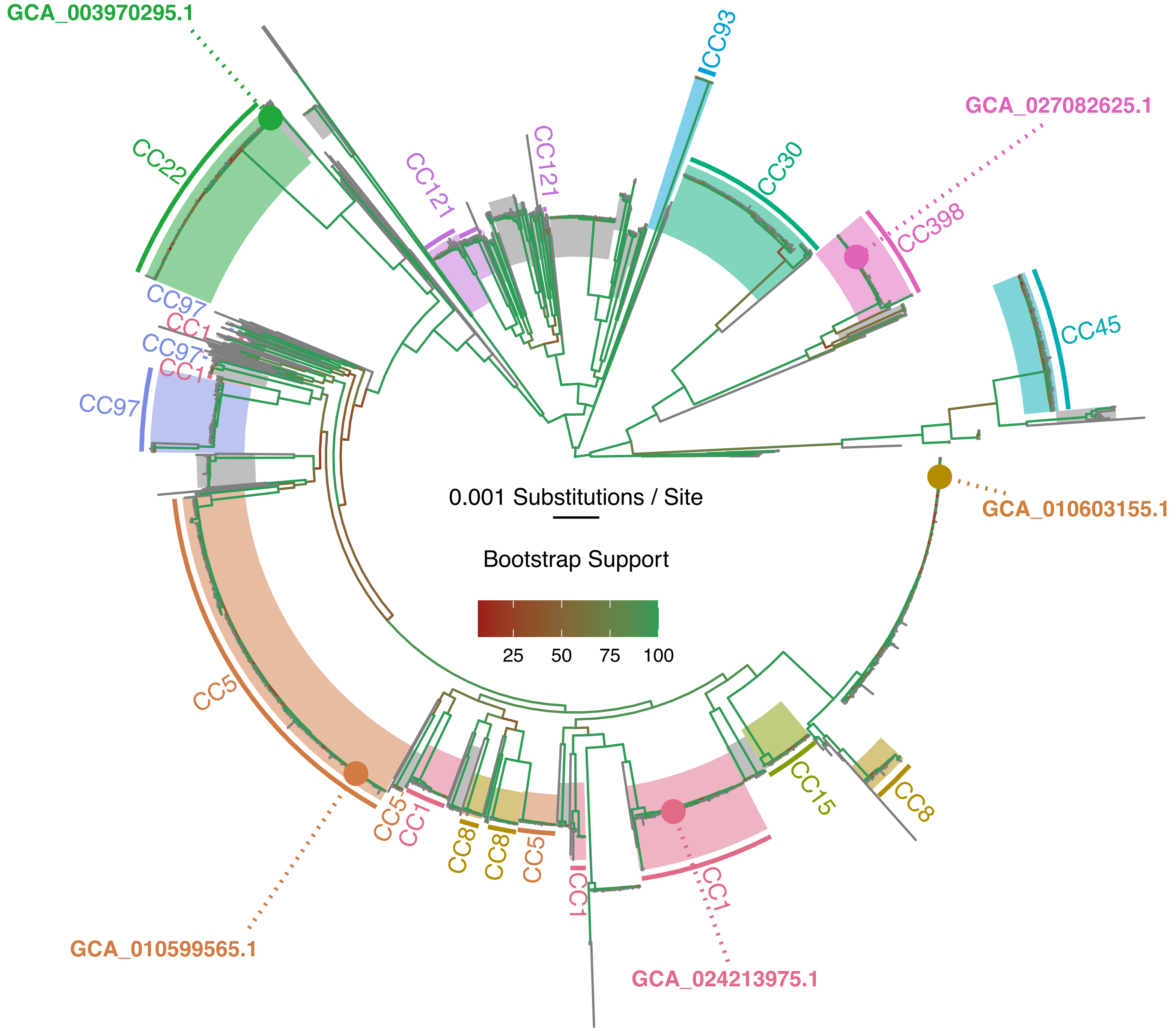


**Supplementary Figure 3. Core-gene maximum likelihood phylogeny of the *S. aureus* representative genomes and the five ancestral seeds**. Maximum-likelihood phylogeny of 1,830 representative genomes, filtered from high-quality public *S. aureus* genomes by deduplication at 99.5% average nucleotide identity within each sequence type (Methods 2.1). Branches are colored by ultrafast bootstrap support values. Terminal branches that have no support values are gray. Shaded clades and outer arcs mark clonal complexes (CCs), and gray shading marks genomes without an assigned CC. The five ancestral seeds, chosen to span the phylogeny, are marked with filled circles and labeled with their GenBank accessions. ST assignments and metadata for all 1,830 representatives are provided in Supplementary Table 6.
