## Supplementary Figure 4 for "THRESHER: A tool for evidence-based, phylogeny-corrected SNP thresholds for whole genome molecular epidemiology"

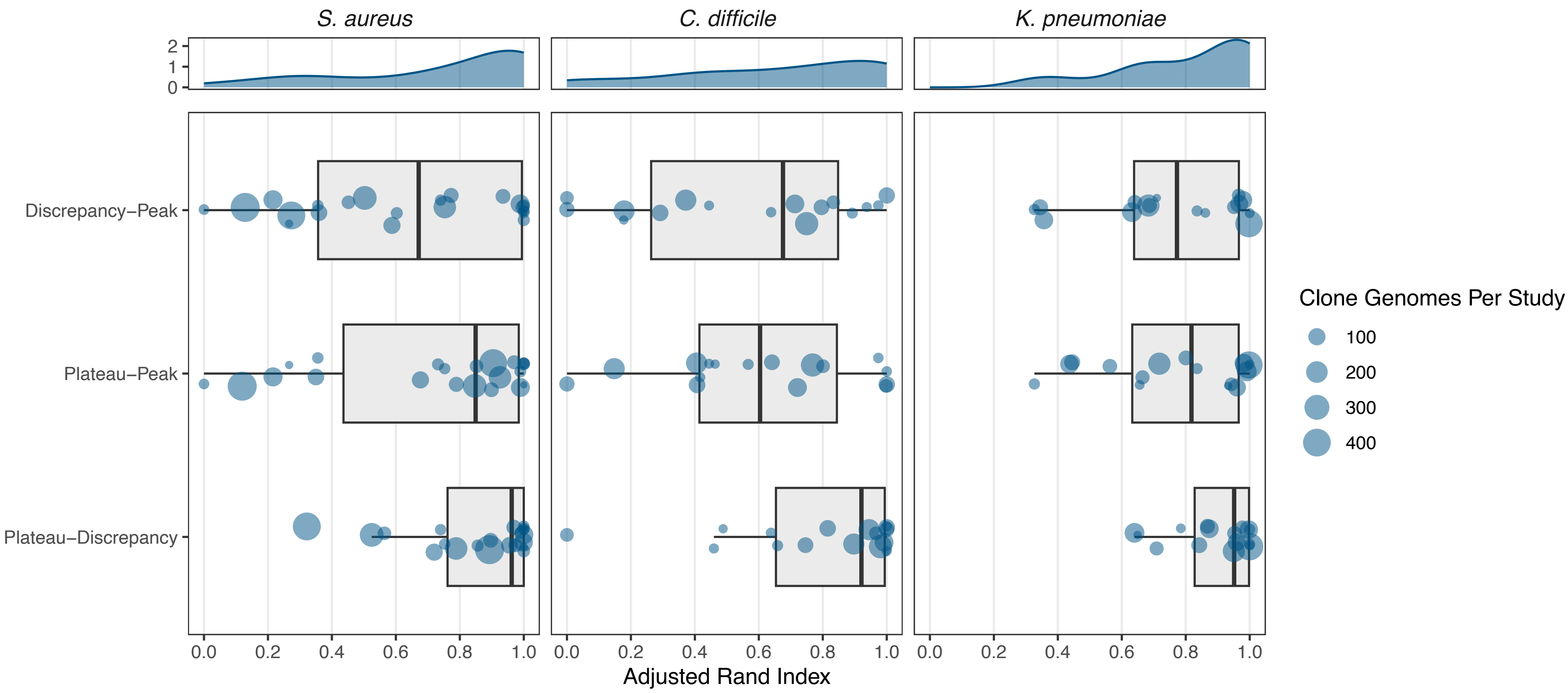


**Supplementary Figure 4. Agreement in strain assignments among the *plateau*, *discrepancy*, and *peak* endpoint methods.** Adjusted Rand index (ARI) between the strain compositions inferred by each pair of endpoints, computed per study for *S. aureus*, *C. difficile*, and *K. pneumoniae* (n = 22, 17, and 16 studies, respectively). An ARI of 1 indicates identical assignments, and 0 indicates agreement expected by chance. For each study, ARI is computed over genomes with a strain assignment at all three endpoints, excluding genomes that are singletons under every endpoint. Each point is one study, jittered along the endpoint-pair axis and sized by the number of genomes retained, that is, genomes assigned to a clone by at least one endpoint. Box plots show the median (center line) and interquartile range (box), with whiskers extending to the most extreme values within 1.5× the interquartile range. Top, density of ARI values for each species, pooled across endpoint pairs.
