## Supplementary Figure 5 for "THRESHER: A tool for evidence-based, phylogeny-corrected SNP thresholds for whole genome molecular epidemiology"

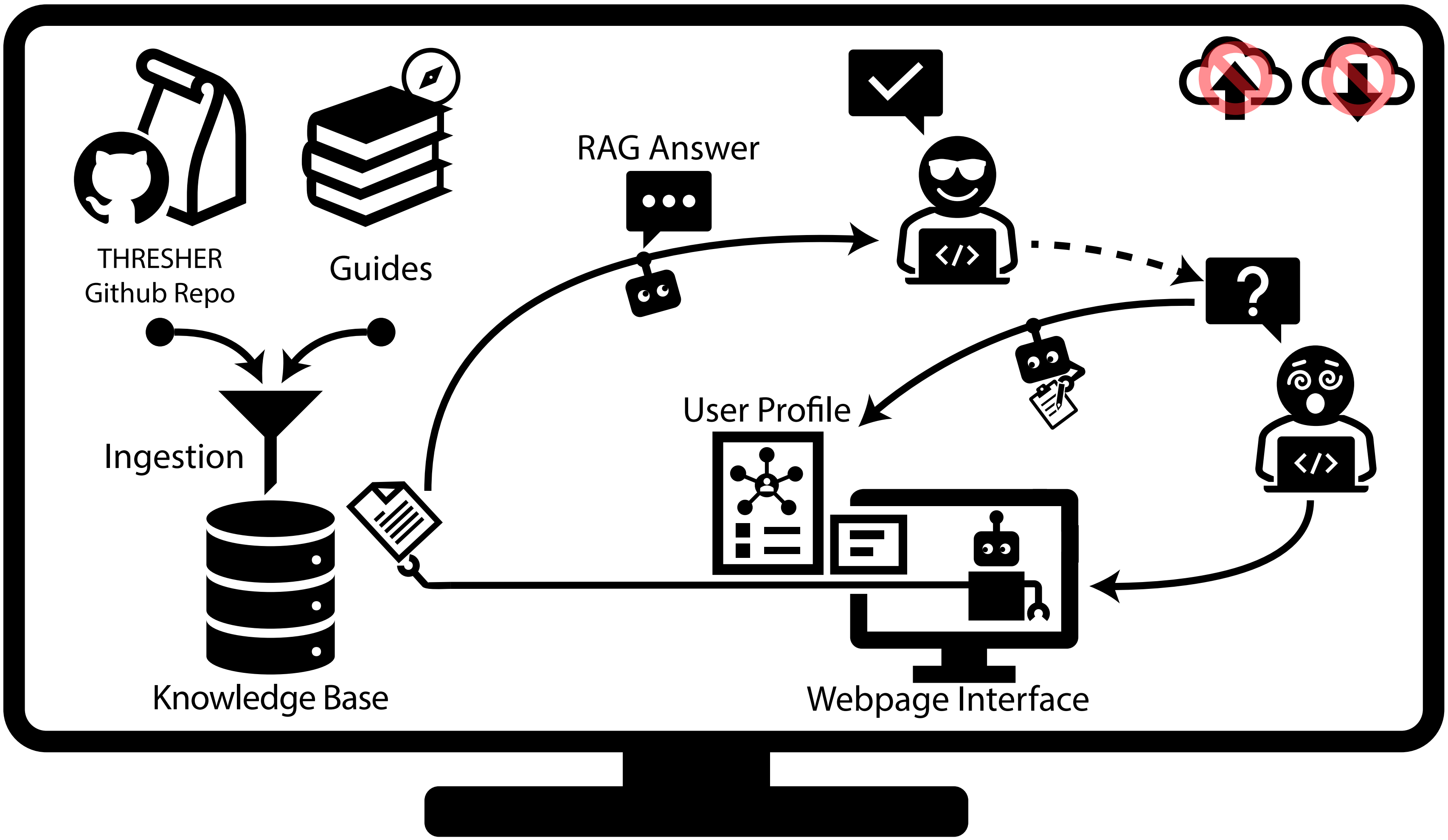


**Supplementary Figure 5. Workflow of THRESHER-Chat, a local offline assistant for THRESHER.** All components run on the user's own system (outer monitor). The crossed-out cloud upload and download icons indicate that THRESHER-Chat does not rely on cloud services, so questions, uploaded files, and patient-associated metadata never leave the local system. During ingestion, a local copy of the THRESHER GitHub repository (Python, R, and Snakemake source code and Markdown documentation) and a set of curated guides shipped with THRESHER-Chat are split into chunks. These chunks are embedded with a locally served embedding model and stored in a persistent ChromaDB vector store (the knowledge base), which is built once and reused across sessions. The guides hold only rationale and interpretation guidance, while the repository documentation supplies command syntax, inputs, and outputs. For each question submitted through the local web interface, the most similar chunks (document icon) are retrieved from the knowledge base. They are supplied, together with the question, to a locally served, user-selectable language model (robot icons). A system prompt constrains the model to answer only from the retrieved passages and to cite their source files, and the resulting retrieval-augmented generation (RAG) answer is returned to the user. The user profile tailors the depth and framing of answers. It combines an audience profile selected in the web interface (e.g., graduate student, principal investigator, clinician, bioinformatician, statistician, or infection prevention and control team) with a persistent local memory (SQLite). The memory retains the conversation history and a compact user record (robot holding the clipboard). This record holds profile tags inferred from the dialogue, stated analytical objectives, and summaries of past sessions, all generated by the same local model. The profile and memory are added to the model input for answering, whereas retrieval uses the original question alone. The dashed arrow denotes follow-up questions, within or across sessions, which repeat the cycle with the updated profile and memory. Details are provided in Methods Section 1.6.
