## Supplementary Figure 6 for "THRESHER: A tool for evidence-based, phylogeny-corrected SNP thresholds for whole genome molecular epidemiology"

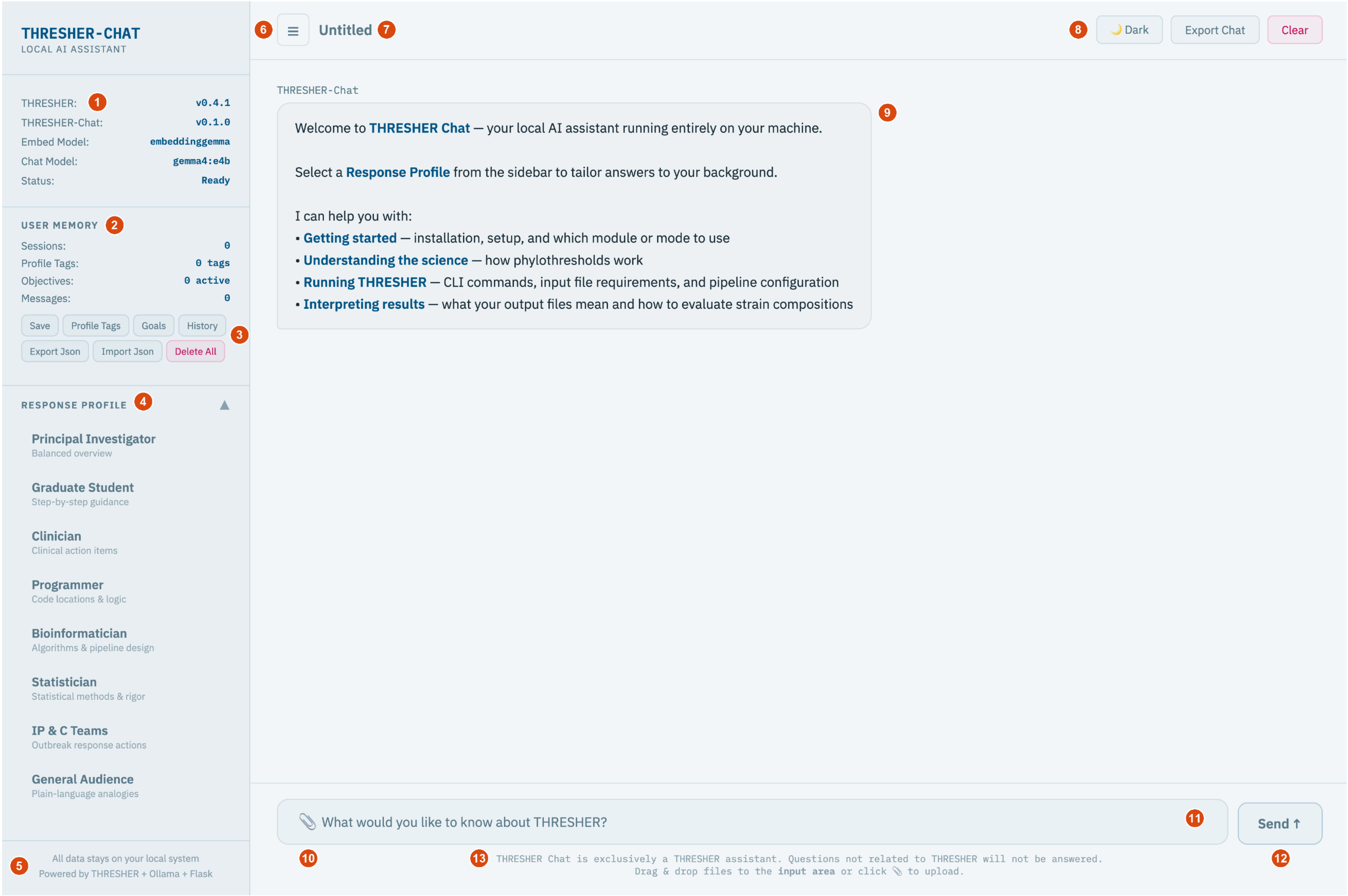


**Supplementary Figure 6. THRESHER-Chat web interface.** THRESHER-Chat is a local, offline chatbot assistant that answers questions about the functions, concepts, and usage of THRESHER. For each question, relevant contexts are retrieved from the THRESHER source repository and curated guides, and a language model running locally provides an answer that cites the source files used. The interface is shown at launch (thresher-chat server), before any conversation and with no stored memory, with the Response Profile list expanded.

(1) Status panel: versions of the ingested THRESHER repository (v0.4.1) and of THRESHER-Chat (v0.1.0), the embedding (embeddinggemma) and chat (gemma4:e4b) models, and the server connection status.

(2) User Memory counters: numbers of saved sessions, inferred profile tags, active research objectives, and stored messages. Memory is stored locally and supplied as context to tailor later answers.

(3) Memory controls: Save button summarizes the current session and extracts profile tags and research objectives. Profile Tags, Goals, and History buttons open lists for review and correction. Export Json and Import Json back up and restore memory. Delete All permanently erases it.

(4) Response Profile: eight profiles (Principal Investigator, Graduate Student, Clinician, Programmer, Bioinformatician, Statistician, Infection Prevention and Control [IP & C] Teams, and General Audience) that adjust the depth and focus of answers without changing their sources.

(5) Reminder that all data stay on the local system.

(6) Sidebar toggle.

(7) Conversation title, generated automatically from the first question.

(8) Header controls: switch between light and dark themes, export the conversation as a text file, and clear the conversation (the session is saved to memory before a new one starts).

(9) Conversation area, showing the welcome message; each answer is followed by the names of the source files it drew on.

(10) File upload: text files such as metadata tables or logs (up to 50 MB; the first 15,000 characters are read) can be attached here or by drag and drop, so that questions can refer to them.

(11) Question box.

(12) Send button: questions can also be sent with Enter.

(13) Scope notice: questions unrelated to THRESHER are declined.
