## Supplementary Figure 7 for "THRESHER: A tool for evidence-based, phylogeny-corrected SNP thresholds for whole genome molecular epidemiology"

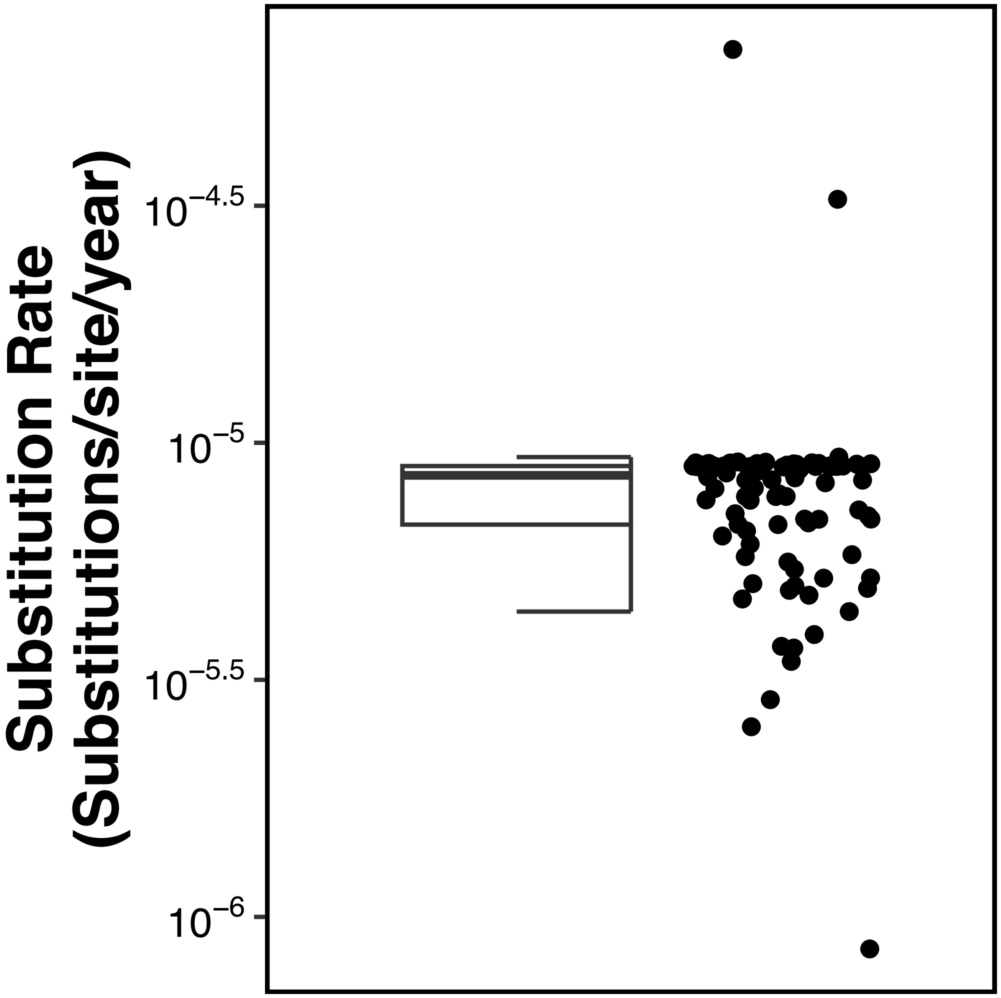


**Supplementary Figure 7**. **Per-clone substitution rates in a published NICU *S. aureus* dataset**. Substitution rates for longitudinally persistent clones with at least four genomes each (n = 93) from a previously published neonatal intensive care unit *S. aureus* dataset[16]. Rates are estimated with BEAST v2.7.6 under an HKY substitution model and a strict clock with a constant-population coalescent prior. Each point is the posterior mean rate of one clone, jittered horizontally. The box plot, computed on log10-transformed rates, shows the median (center line) and interquartile range (box), with whiskers extending to the most extreme values within 1.5× the interquartile range. The median rate, $8.5 \times 10^{-6}$ substitutions per site per year (IQR, $6.7 \times 10^{-6}$ – $8.9 \times 10^{-6}$), is used as the substitution rate in the simulations.
