## Supplementary Figure 8 for "THRESHER: A tool for evidence-based, phylogeny-corrected SNP thresholds for whole genome molecular epidemiology"

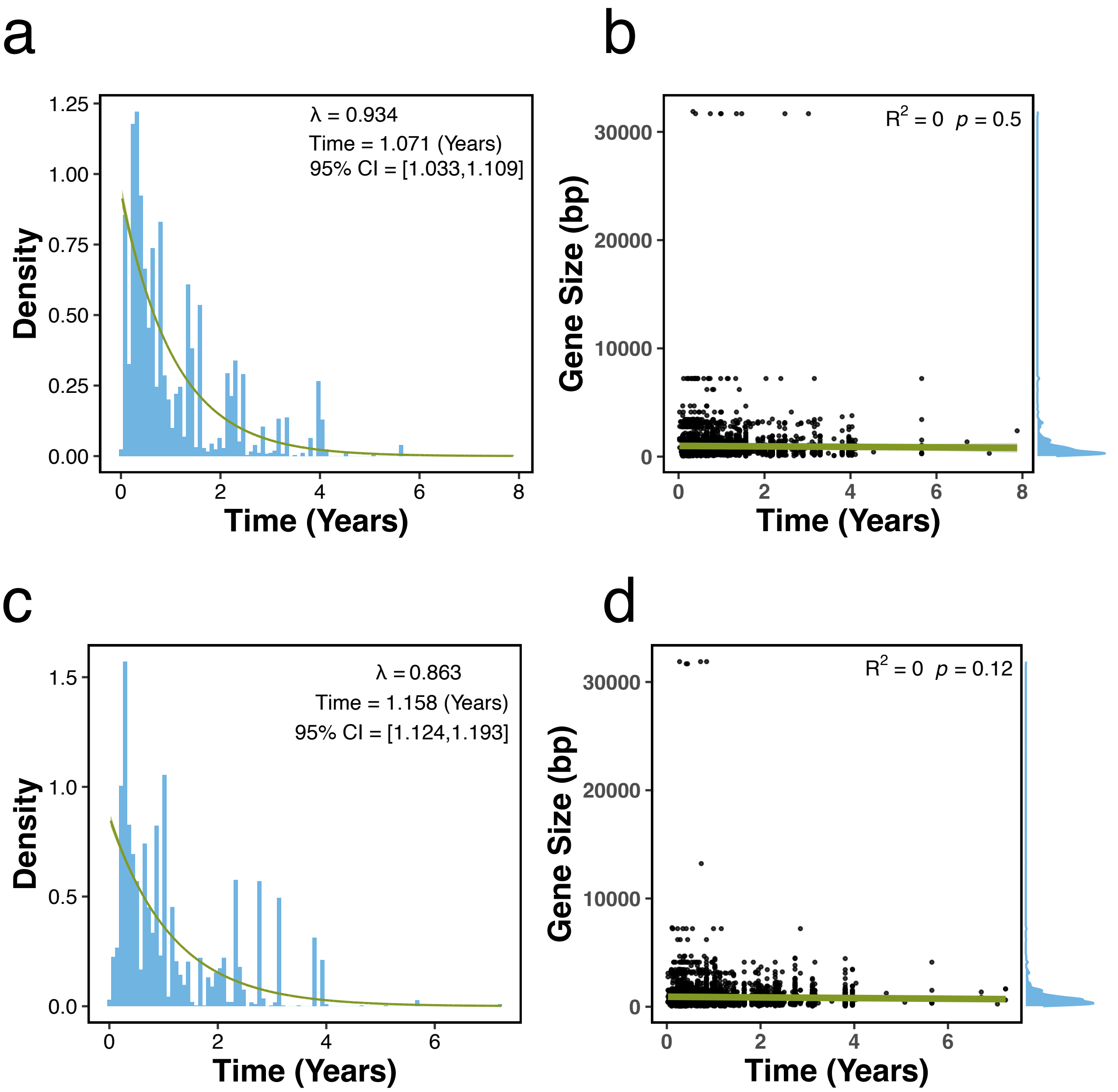


**Supplementary Figure 8. Time for gene gain and loss in NICU *S. aureus* dataset.** Gain and loss of 7,352 variably present genes across 92 clones from a previously published neonatal intensive care unit (NICU) *S. aureus* dataset[16]. For each gene within a clone, the time in years is measured on the time-scaled phylogeny of a clone, from the inferred emergence of the clone to the first genome in which the gene appeared or disappeared. A gene present in the earliest genome is assumed to have been present in the most recent common ancestor and is scored as a loss, otherwise as a gain.

(a) Distribution of times to gene gain (bars) with the fitted exponential distribution (line) and its 95% bootstrap confidence interval from 10,000 replicates (shaded). The fitted rate $\lambda$, the corresponding mean time $1/\lambda$, and its 95% confidence interval are shown above the panel.

(b) Gene size against the time to gain, with a linear regression line and 95% confidence interval. The adjusted $R^2$ and $p$ value for the slope are shown above the panel, and the marginal density of gene size is shown at right.

(c, d) As in (a) and (b), for gene loss. Gene size shows no relationship with the time of either event, so the fitted rates are applied independently of gene size in the simulations.
