## Supplementary Figure 9 for "THRESHER: A tool for evidence-based, phylogeny-corrected SNP thresholds for whole genome molecular epidemiology"

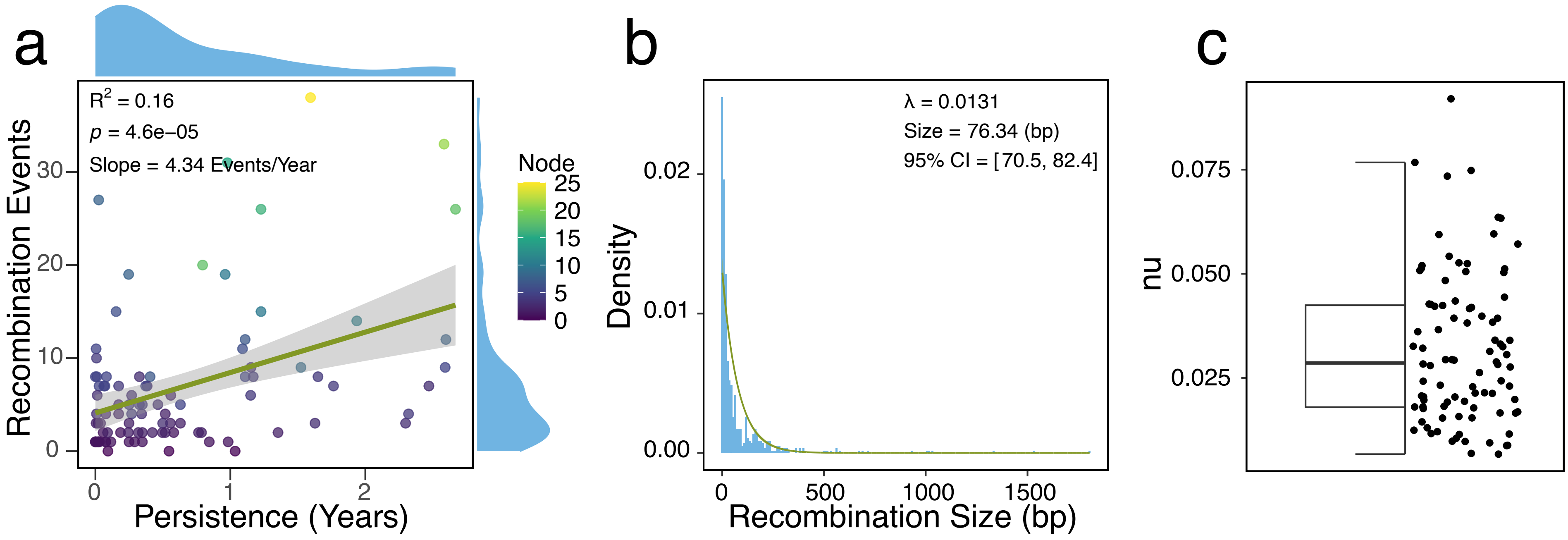


**Supplementary Figure 9**. **Recombination in NICU *S. aureus* dataset**.

Recombination inferred with ClonalFrameML for the clones in a previously published neonatal intensive care unit (NICU) *S. aureus* dataset[16] (n = 92 clones).

(a) Number of recombination events against the persistence per clone. The persistence was measured as the time interval in years between the earliest and latest genome collection dates. Each point is one clone, colored by the number of phylogeny nodes at which recombination was inferred. The line is a linear regression with its 95% confidence interval (shaded). The adjusted $R^2$, $p$ value, and slope are shown in the panel, and the slope gives the recombination rate of 4.34 events per year used in the simulations. Marginal densities of both variables are shown at the top and right of the panel.

(b) Distribution of inferred recombination segment lengths across all clones (bars) with the fitted exponential distribution (line) and its 95% bootstrap confidence interval from 10,000 replicates (shaded). The fitted rate λ, the corresponding mean segment length 1/λ, and its 95% confidence interval are shown in the panel.

(c) Posterior mean ν, the SNP density of recombined segments relative to the clonal background, for each clone. Points are individual clones, jittered horizontally. The box plot shows the median (center line) and interquartile range (box), with whiskers extending to the most extreme values within 1.5× the interquartile range. The mean across clones, 0.03, is the value used in the simulations.
