## Supplementary Data 2 for "THRESHER: A tool for evidence-based, phylogeny-corrected SNP thresholds for whole genome molecular epidemiology": ST1_20_iter_1.pdf

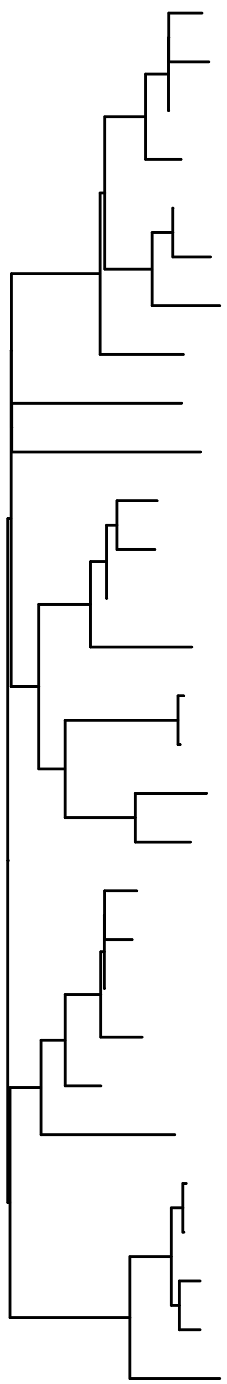

| | Simulation | Plateau 5 | Plateau 15 | Peak | Discrepancy | Cutoff 0 | Cutoff 10 | Cutoff 20 | Cutoff 30 | Cutoff 40 | Cutoff 50 |
|---|---|---|---|---|---|---|---|---|---|---|---|
| ST1_3_t9 | 33 | 13 | 5 | 5 | 1 | 39 | 38 | 38 | 36 | 19 | 19 |
| ST1_3_n9 | 33 | 23 | 5 | 5 | 1 | 88 | 83 | 80 | 75 | 19 | 19 |
| ST1_3_n11 | 33 | 7 | 5 | 5 | 1 | 21 | 20 | 20 | 20 | 19 | 19 |
| ST1_3_n12 | 33 | 6 | 5 | 8 | 1 | 10 | 10 | 10 | 10 | 10 | 10 |
| ST1_3_n19 | 34 | 8 | 6 | 6 | 1 | 24 | 23 | 23 | 3 | 3 | 3 |
| ST1_3_t3 | 34 | 3 | 6 | 6 | 1 | 3 | 3 | 3 | 3 | 3 | 3 |
| ST1_3_t7 | 34 | 25 | 6 | 18 | 1 | 107 | 99 | 95 | 88 | 75 | 69 |
| ST1_3_t16 | 32 | 5 | 7 | 7 | 1 | 9 | 9 | 9 | 9 | 9 | 9 |
| ST1_3_n1 | 31 | 12 | 9 | 11 | 1 | 37 | 36 | 36 | 34 | 33 | 32 |
| ST1_1_t23 | 11 | 15 | 11 | 13 | 1 | 53 | 50 | 49 | 47 | 44 | 42 |
| ST1_2_t4 | 24 | 19 | 3 | 3 | 1 | 72 | 68 | 65 | 61 | 53 | 50 |
| ST1_2_t20 | 24 | 18 | 3 | 3 | 1 | 69 | 65 | 62 | 58 | 53 | 50 |
| ST1_2_n18 | 24 | 27 | 3 | 3 | 1 | 121 | 113 | 105 | 95 | 53 | 50 |
| ST1_2_t3 | 23 | 21 | 14 | 17 | 1 | 82 | 77 | 74 | 70 | 63 | 59 |
| ST1_2_t23 | 22 | 2 | 4 | 4 | 1 | 55 | 15 | 15 | 15 | 15 | 15 |
| ST1_2_t16 | 22 | 2 | 4 | 4 | 1 | 16 | 15 | 15 | 15 | 15 | 15 |
| ST1_2_t13 | 26 | 17 | 13 | 15 | 1 | 63 | 59 | 57 | 55 | 50 | 48 |
| ST1_2_n1 | 21 | 16 | 12 | 14 | 1 | 56 | 52 | 51 | 49 | 46 | 44 |
| ST1_1_t25 | 14 | 22 | 1 | 1 | 1 | 85 | 80 | 77 | 4 | 4 | 4 |
| ST1_1_n14 | 14 | 11 | 1 | 1 | 1 | 36 | 35 | 35 | 4 | 4 | 4 |
| ST1_1_n17 | 14 | 4 | 1 | 1 | 1 | 4 | 4 | 4 | 4 | 4 | 4 |
| ST1_1_n18 | 14 | 24 | 1 | 1 | 1 | 104 | 96 | 92 | 85 | 4 | 4 |
| ST1_1_n13 | 13 | 9 | 1 | 9 | 1 | 28 | 27 | 27 | 26 | 25 | 25 |
| ST1_1_t7 | 16 | 14 | 10 | 12 | 1 | 50 | 48 | 48 | 46 | 43 | 41 |
| ST1_1_t19 | 12 | 1 | 2 | 2 | 1 | 123 | 56 | 54 | 52 | 48 | 46 |
| ST1_1_t10 | 12 | 1 | 2 | 2 | 1 | 60 | 56 | 54 | 52 | 48 | 46 |
| ST1_1_t6 | 12 | 20 | 2 | 2 | 1 | 77 | 73 | 70 | 66 | 48 | 46 |
| ST1_1_n7 | 12 | 26 | 2 | 2 | 1 | 113 | 105 | 100 | 91 | 48 | 46 |
| ST1_1_t13 | 12 | 10 | 8 | 10 | 1 | 34 | 33 | 33 | 32 | 31 | 30 |

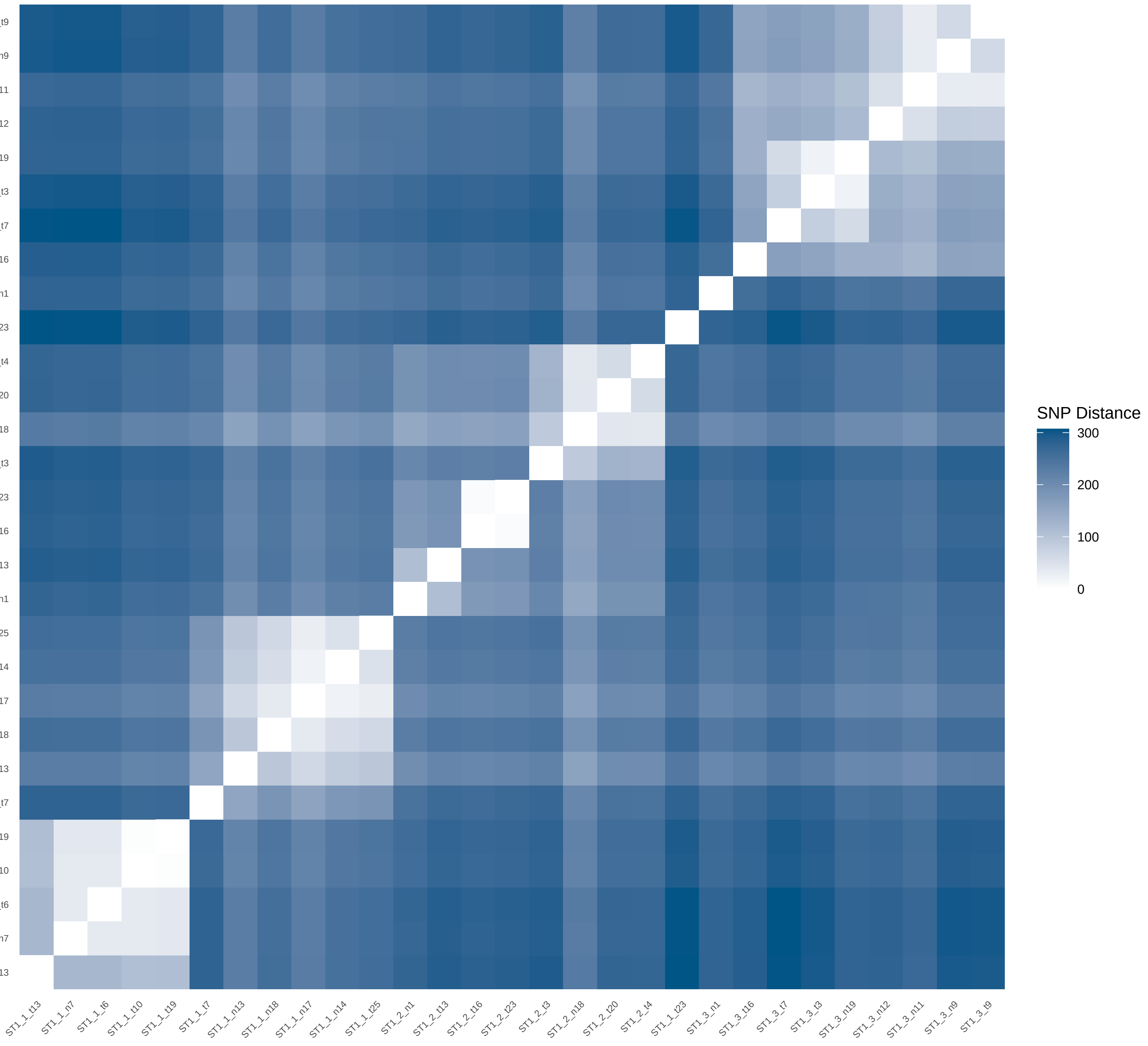
