## Supplementary Data 2 for "THRESHER: A tool for evidence-based, phylogeny-corrected SNP thresholds for whole genome molecular epidemiology": ST1_20_iter_2.pdf

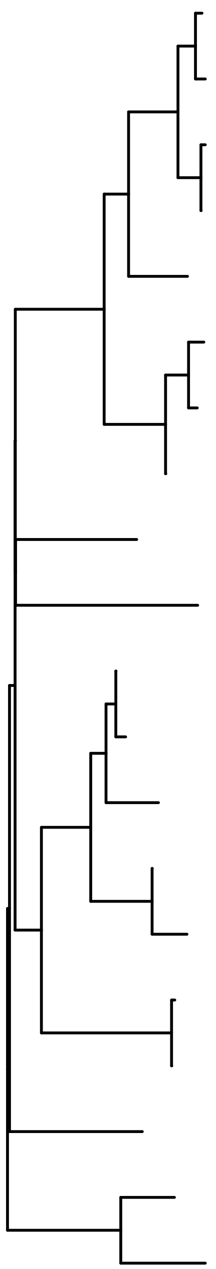

| | Simulation | Plateau 5 | Plateau 15 | Peak | Discrepancy | Cutoff 0 | Cutoff 10 | Cutoff 20 | Cutoff 30 | Cutoff 40 | Cutoff 50 |
|---|---|---|---|---|---|---|---|---|---|---|---|
| ST1_3_t23 | 34 | 3 | 4 | 5 | 2 | 128 | 113 | 56 | 55 | 50 | 33 |
| ST1_3_n14 | 34 | 3 | 4 | 5 | 2 | 60 | 57 | 56 | 55 | 50 | 33 |
| ST1_3_t22 | 34 | 4 | 4 | 6 | 2 | 87 | 39 | 39 | 38 | 36 | 33 |
| ST1_3_n16 | 34 | 4 | 4 | 6 | 2 | 42 | 39 | 39 | 38 | 36 | 33 |
| ST1_3_t21 | 36 | 9 | 7 | 9 | 2 | 57 | 54 | 53 | 52 | 48 | 43 |
| ST1_3_n9 | 33 | 10 | 3 | 4 | 2 | 64 | 61 | 60 | 49 | 46 | 42 |
| ST1_3_n8 | 33 | 11 | 3 | 4 | 2 | 78 | 73 | 71 | 49 | 46 | 42 |
| ST1_3_n11 | 33 | 8 | 3 | 4 | 2 | 54 | 51 | 50 | 49 | 46 | 42 |
| ST1_3_n3 | 31 | 14 | 9 | 12 | 2 | 105 | 95 | 92 | 80 | 73 | 65 |
| ST1_1_t20 | 11 | 6 | 5 | 7 | 2 | 47 | 44 | 44 | 43 | 41 | 38 |
| ST1_2_n17 | 24 | 1 | 1 | 1 | 2 | 74 | 62 | 61 | 59 | 54 | 48 |
| ST1_2_n12 | 24 | 1 | 1 | 1 | 2 | 65 | 62 | 61 | 59 | 54 | 48 |
| ST1_2_n10 | 24 | 13 | 1 | 11 | 2 | 98 | 89 | 86 | 74 | 67 | 60 |
| ST1_2_n9 | 23 | 15 | 1 | 3 | 2 | 108 | 98 | 95 | 82 | 8 | 7 |
| ST1_2_t3 | 23 | 5 | 1 | 3 | 2 | 8 | 8 | 8 | 8 | 8 | 7 |
| ST1_2_t16 | 22 | 2 | 2 | 2 | 2 | 117 | 36 | 36 | 35 | 33 | 31 |
| ST1_2_n4 | 22 | 2 | 2 | 2 | 2 | 39 | 36 | 36 | 35 | 33 | 31 |
| ST1_1_t3 | 13 | 7 | 6 | 8 | 2 | 52 | 49 | 48 | 47 | 45 | 41 |
| ST1_1_t19 | 12 | 16 | 10 | 13 | 1 | 120 | 107 | 103 | 90 | 81 | 70 |
| ST1_1_t13 | 12 | 12 | 8 | 10 | 1 | 85 | 80 | 78 | 68 | 62 | 55 |

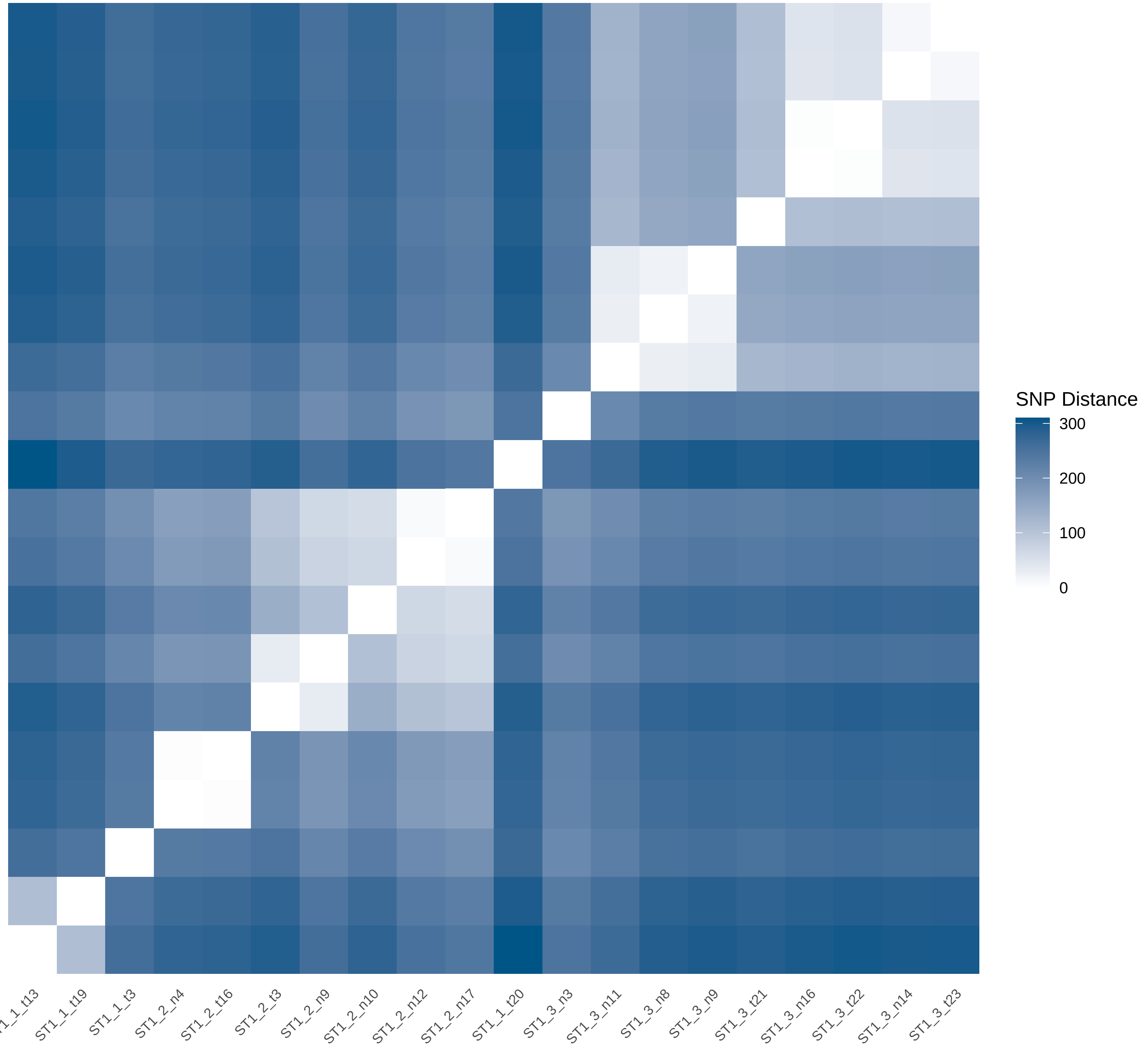
