## Supplementary Data 2 for "THRESHER: A tool for evidence-based, phylogeny-corrected SNP thresholds for whole genome molecular epidemiology": ST1_20_iter_3.pdf

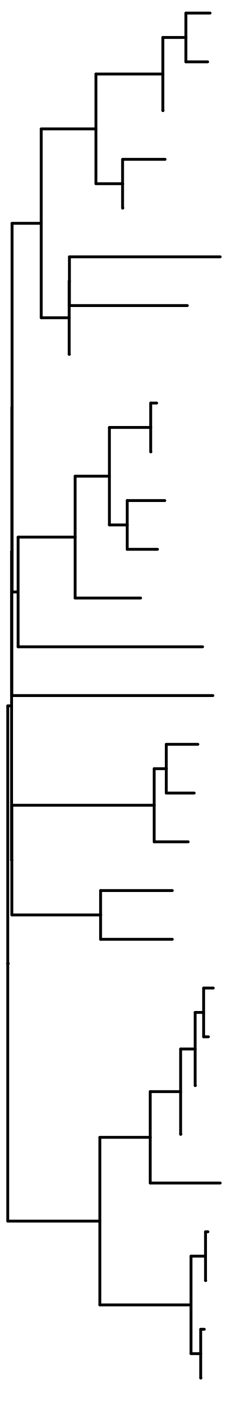

| | Simulation | Plateau 5 | Plateau 15 | Peak | Discrepancy | Cutoff 0 | Cutoff 10 | Cutoff 20 | Cutoff 30 | Cutoff 40 | Cutoff 50 |
|---|---|---|---|---|---|---|---|---|---|---|---|
| ST1_2_t24 | 23 | 10 | 2 | 4 | 1 | 44 | 44 | 44 | 41 | 34 | 33 |
| ST1_2_t19 | 23 | 18 | 2 | 4 | 1 | 72 | 70 | 69 | 65 | 34 | 33 |
| ST1_2_n9 | 23 | 9 | 2 | 4 | 1 | 38 | 38 | 38 | 37 | 34 | 33 |
| ST1_2_t4 | 24 | 7 | 2 | 3 | 1 | 22 | 22 | 22 | 22 | 22 | 21 |
| ST1_2_n17 | 24 | 8 | 2 | 3 | 1 | 34 | 34 | 34 | 22 | 22 | 21 |
| ST1_2_t5 | 21 | 17 | 2 | 13 | 1 | 60 | 60 | 60 | 56 | 51 | 50 |
| ST1_2_n4 | 22 | 13 | 2 | 12 | 1 | 48 | 48 | 48 | 45 | 40 | 39 |
| ST1_2_n5 | 211 | 6 | 2 | 9 | 1 | 6 | 6 | 6 | 6 | 6 | 6 |
| ST1_1_t2 | 14 | 1 | 1 | 2 | 1 | 35 | 35 | 35 | 34 | 33 | 32 |
| ST1_1_n18 | 14 | 1 | 1 | 2 | 1 | 65 | 35 | 35 | 34 | 33 | 32 |
| ST1_1_t9 | 14 | 23 | 1 | 1 | 1 | 129 | 115 | 104 | 93 | 85 | 42 |
| ST1_1_n15 | 14 | 15 | 1 | 1 | 1 | 51 | 51 | 51 | 48 | 43 | 42 |
| ST1_1_t1 | 13 | 12 | 8 | 11 | 1 | 46 | 46 | 46 | 43 | 38 | 37 |
| ST1_1_t10 | 12 | 19 | 11 | 14 | 1 | 100 | 93 | 87 | 78 | 72 | 68 |
| ST1_1_t14 | 11 | 5 | 6 | 8 | 1 | 1 | 1 | 1 | 1 | 1 | 1 |
| ST1_2_t7 | 25 | 16 | 3 | 5 | 1 | 57 | 57 | 57 | 53 | 48 | 47 |
| ST1_2_t18 | 25 | 14 | 3 | 5 | 1 | 50 | 50 | 50 | 47 | 42 | 41 |
| ST1_2_t21 | 25 | 20 | 3 | 15 | 1 | 104 | 96 | 89 | 80 | 74 | 69 |
| ST1_3_t6 | 31 | 11 | 7 | 10 | 1 | 45 | 45 | 45 | 42 | 37 | 36 |
| ST1_3_t4 | 35 | 22 | 12 | 17 | 1 | 121 | 108 | 100 | 90 | 83 | 78 |
| ST1_3_t5 | 34 | 2 | 4 | 6 | 3 | 107 | 64 | 26 | 26 | 26 | 25 |
| ST1_3_t22 | 34 | 2 | 4 | 6 | 3 | 64 | 64 | 26 | 26 | 26 | 25 |
| ST1_3_n17 | 34 | 2 | 4 | 6 | 3 | 26 | 26 | 26 | 26 | 26 | 25 |
| ST1_3_n18 | 34 | 2 | 4 | 6 | 3 | 127 | 113 | 26 | 26 | 26 | 25 |
| ST1_3_t7 | 34 | 21 | 4 | 16 | 3 | 105 | 97 | 90 | 81 | 75 | 70 |
| ST1_3_t15 | 33 | 4 | 5 | 7 | 3 | 8 | 8 | 8 | 8 | 8 | 8 |
| ST1_3_n9 | 33 | 4 | 5 | 7 | 3 | 110 | 8 | 8 | 8 | 8 | 8 |
| ST1_3_t20 | 33 | 3 | 5 | 7 | 3 | 73 | 56 | 56 | 8 | 8 | 8 |
| ST1_3_n8 | 33 | 3 | 5 | 7 | 3 | 56 | 56 | 56 | 8 | 8 | 8 |

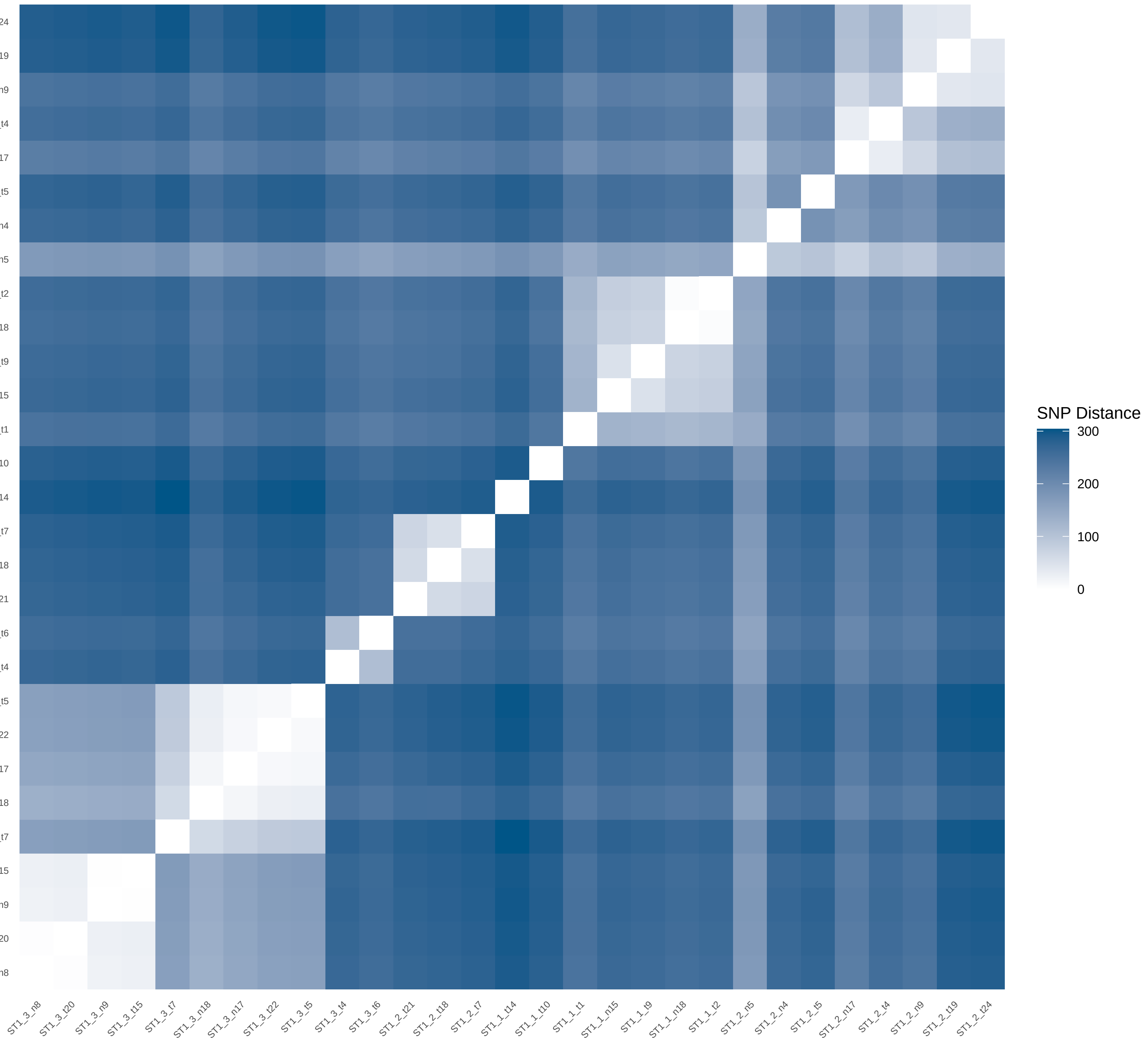
