## Supplementary Data 2 for "THRESHER: A tool for evidence-based, phylogeny-corrected SNP thresholds for whole genome molecular epidemiology": ST1_20_iter_4.pdf

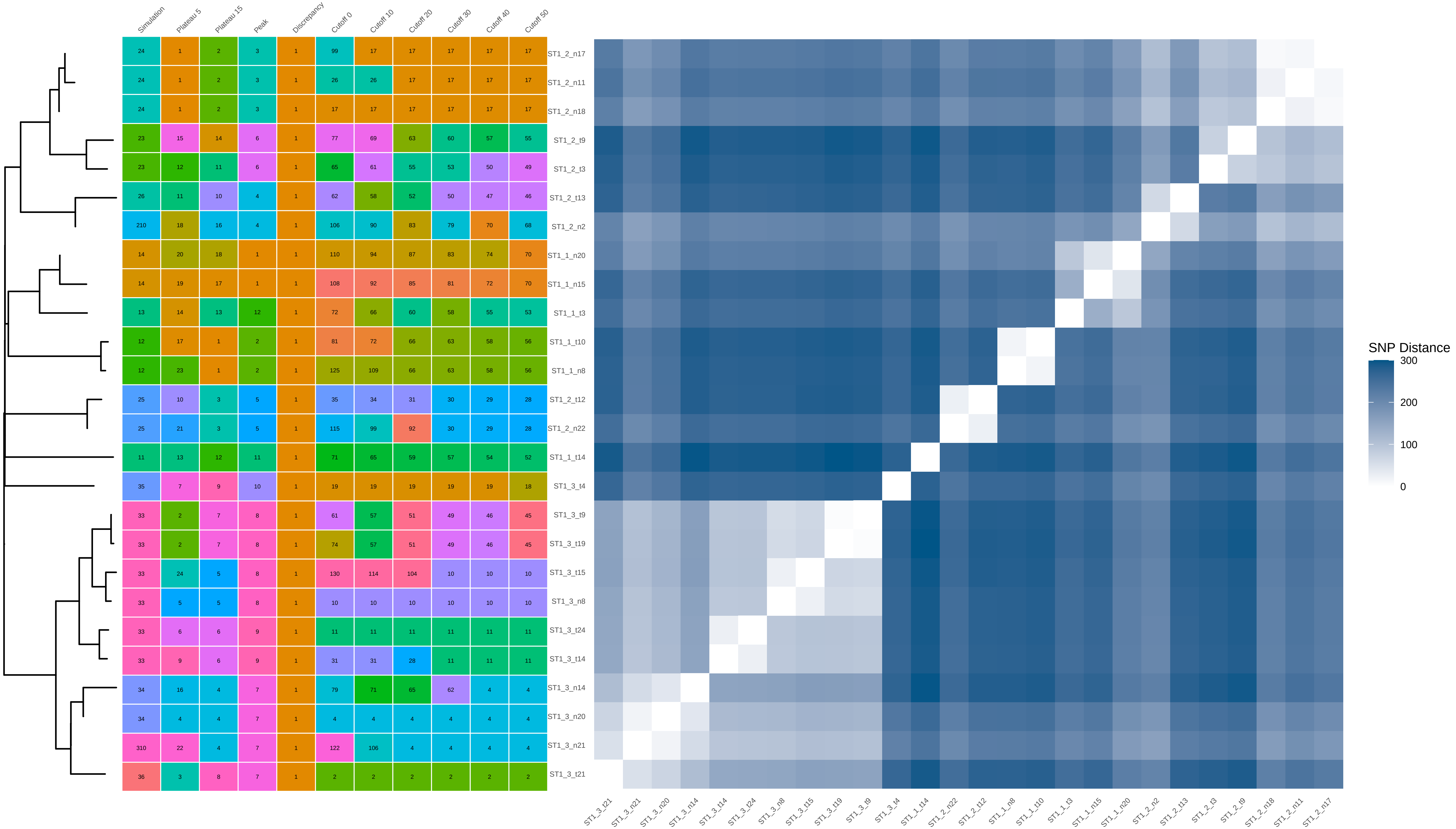


| | Simulation | Plateau 5 | Plateau 15 | Peak | Discrepancy | Cutoff 0 | Cutoff 10 | Cutoff 20 | Cutoff 30 | Cutoff 40 | Cutoff 50 |
|---|---|---|---|---|---|---|---|---|---|---|---|
| ST1_2_n17 | 24 | 1 | 2 | 3 | 1 | 99 | 17 | 17 | 17 | 17 | 17 |
| ST1_2_n11 | 24 | 1 | 2 | 3 | 1 | 26 | 26 | 17 | 17 | 17 | 17 |
| ST1_2_n18 | 24 | 1 | 2 | 3 | 1 | 17 | 17 | 17 | 17 | 17 | 17 |
| ST1_2_t9 | 23 | 15 | 14 | 6 | 1 | 77 | 69 | 63 | 60 | 57 | 55 |
| ST1_2_t3 | 23 | 12 | 11 | 6 | 1 | 65 | 61 | 55 | 53 | 50 | 49 |
| ST1_2_t13 | 26 | 11 | 10 | 4 | 1 | 62 | 58 | 52 | 50 | 47 | 46 |
| ST1_2_n2 | 210 | 18 | 16 | 4 | 1 | 106 | 90 | 83 | 79 | 70 | 68 |
| ST1_1_n20 | 14 | 20 | 18 | 1 | 1 | 110 | 94 | 87 | 83 | 74 | 70 |
| ST1_1_n15 | 14 | 19 | 17 | 1 | 1 | 108 | 92 | 85 | 81 | 72 | 70 |
| ST1_1_t3 | 13 | 14 | 13 | 12 | 1 | 72 | 66 | 60 | 58 | 55 | 53 |
| ST1_1_t10 | 12 | 17 | 1 | 2 | 1 | 81 | 72 | 66 | 63 | 58 | 56 |
| ST1_1_n8 | 12 | 23 | 1 | 2 | 1 | 125 | 109 | 66 | 63 | 58 | 56 |
| ST1_2_t12 | 25 | 10 | 3 | 5 | 1 | 35 | 34 | 31 | 30 | 29 | 28 |
| ST1_2_n22 | 25 | 21 | 3 | 5 | 1 | 115 | 99 | 92 | 30 | 29 | 28 |
| ST1_1_t14 | 11 | 13 | 12 | 11 | 1 | 71 | 65 | 59 | 57 | 54 | 52 |
| ST1_3_t4 | 35 | 7 | 9 | 10 | 1 | 19 | 19 | 19 | 19 | 19 | 18 |
| ST1_3_t9 | 33 | 2 | 7 | 8 | 1 | 61 | 57 | 51 | 49 | 46 | 45 |
| ST1_3_t19 | 33 | 2 | 7 | 8 | 1 | 74 | 57 | 51 | 49 | 46 | 45 |
| ST1_3_t15 | 33 | 24 | 5 | 8 | 1 | 130 | 114 | 104 | 10 | 10 | 10 |
| ST1_3_n8 | 33 | 5 | 5 | 8 | 1 | 10 | 10 | 10 | 10 | 10 | 10 |
| ST1_3_t24 | 33 | 6 | 6 | 9 | 1 | 11 | 11 | 11 | 11 | 11 | 11 |
| ST1_3_t14 | 33 | 9 | 6 | 9 | 1 | 31 | 31 | 28 | 11 | 11 | 11 |
| ST1_3_n14 | 34 | 16 | 4 | 7 | 1 | 79 | 71 | 65 | 62 | 4 | 4 |
| ST1_3_n20 | 34 | 4 | 4 | 7 | 1 | 4 | 4 | 4 | 4 | 4 | 4 |
| ST1_3_n21 | 310 | 22 | 4 | 7 | 1 | 122 | 106 | 4 | 4 | 4 | 4 |
| ST1_3_t21 | 36 | 3 | 8 | 7 | 1 | 2 | 2 | 2 | 2 | 2 | 2 |
