## Supplementary Data 2 for "THRESHER: A tool for evidence-based, phylogeny-corrected SNP thresholds for whole genome molecular epidemiology": ST1_20_iter_6.pdf

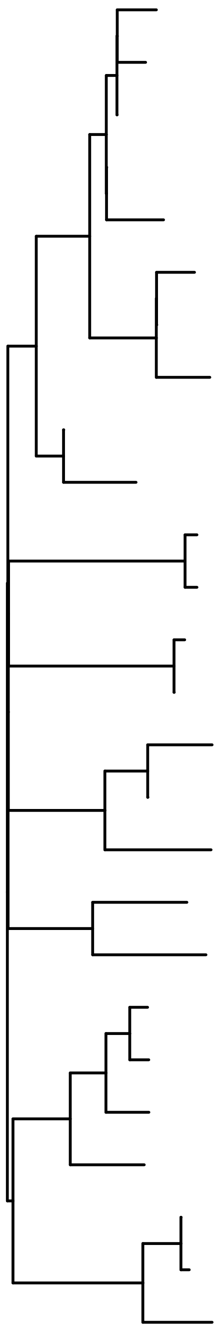

| | Simulation | Plateau 5 | Plateau 15 | Peak | Discrepancy | Cutoff 0 | Cutoff 10 | Cutoff 20 | Cutoff 30 | Cutoff 40 | Cutoff 50 |
|---|---|---|---|---|---|---|---|---|---|---|---|
| ST1_2_t6 | 24 | 2 | 3 | 3 | 1 | 80 | 77 | 68 | 7 | 7 | 7 |
| ST1_2_n15 | 24 | 2 | 3 | 3 | 1 | 23 | 23 | 7 | 7 | 7 | 7 |
| ST1_2_n17 | 24 | 2 | 3 | 3 | 1 | 7 | 7 | 7 | 7 | 7 | 7 |
| ST1_2_n18 | 24 | 2 | 3 | 3 | 1 | 104 | 7 | 7 | 7 | 7 | 7 |
| ST1_2_n10 | 24 | 2 | 3 | 3 | 1 | 126 | 113 | 102 | 91 | 80 | 7 |
| ST1_2_t3 | 23 | 10 | 3 | 6 | 1 | 35 | 35 | 34 | 33 | 16 | 16 |
| ST1_2_n9 | 23 | 5 | 3 | 6 | 1 | 16 | 16 | 16 | 16 | 16 | 16 |
| ST1_2_t24 | 23 | 20 | 3 | 6 | 1 | 107 | 98 | 88 | 78 | 70 | 16 |
| ST1_2_n6 | 29 | 8 | 3 | 4 | 1 | 32 | 32 | 31 | 30 | 30 | 29 |
| ST1_2_n2 | 210 | 11 | 3 | 4 | 1 | 38 | 38 | 37 | 36 | 30 | 29 |
| ST1_3_t2 | 31 | 14 | 6 | 8 | 1 | 53 | 53 | 19 | 19 | 19 | 19 |
| ST1_3_t13 | 31 | 6 | 6 | 8 | 1 | 19 | 19 | 19 | 19 | 19 | 19 |
| ST1_2_t12 | 25 | 3 | 4 | 5 | 1 | 89 | 39 | 38 | 37 | 35 | 33 |
| ST1_2_n21 | 25 | 3 | 4 | 5 | 1 | 39 | 39 | 38 | 37 | 35 | 33 |
| ST1_3_t19 | 33 | 21 | 5 | 7 | 1 | 112 | 102 | 92 | 81 | 73 | 64 |
| ST1_3_n13 | 33 | 16 | 5 | 7 | 1 | 71 | 68 | 60 | 56 | 54 | 51 |
| ST1_3_t23 | 34 | 12 | 9 | 11 | 1 | 45 | 45 | 44 | 43 | 41 | 39 |
| ST1_1_t16 | 15 | 9 | 8 | 10 | 1 | 34 | 34 | 33 | 32 | 32 | 30 |
| ST1_1_t14 | 11 | 19 | 10 | 13 | 1 | 100 | 94 | 85 | 75 | 67 | 61 |
| ST1_1_t22 | 14 | 15 | 1 | 1 | 1 | 63 | 61 | 55 | 51 | 49 | 47 |
| ST1_1_n18 | 14 | 18 | 1 | 1 | 1 | 96 | 90 | 81 | 51 | 49 | 47 |
| ST1_1_t21 | 14 | 13 | 1 | 12 | 1 | 46 | 46 | 45 | 44 | 42 | 40 |
| ST1_1_n12 | 13 | 4 | 7 | 9 | 1 | 14 | 14 | 14 | 14 | 14 | 14 |
| ST1_1_n9 | 12 | 1 | 2 | 2 | 1 | 36 | 36 | 35 | 34 | 33 | 31 |
| ST1_1_n6 | 12 | 1 | 2 | 2 | 1 | 85 | 36 | 35 | 34 | 33 | 31 |
| ST1_1_t12 | 12 | 22 | 11 | 14 | 1 | 122 | 111 | 100 | 89 | 78 | 68 |

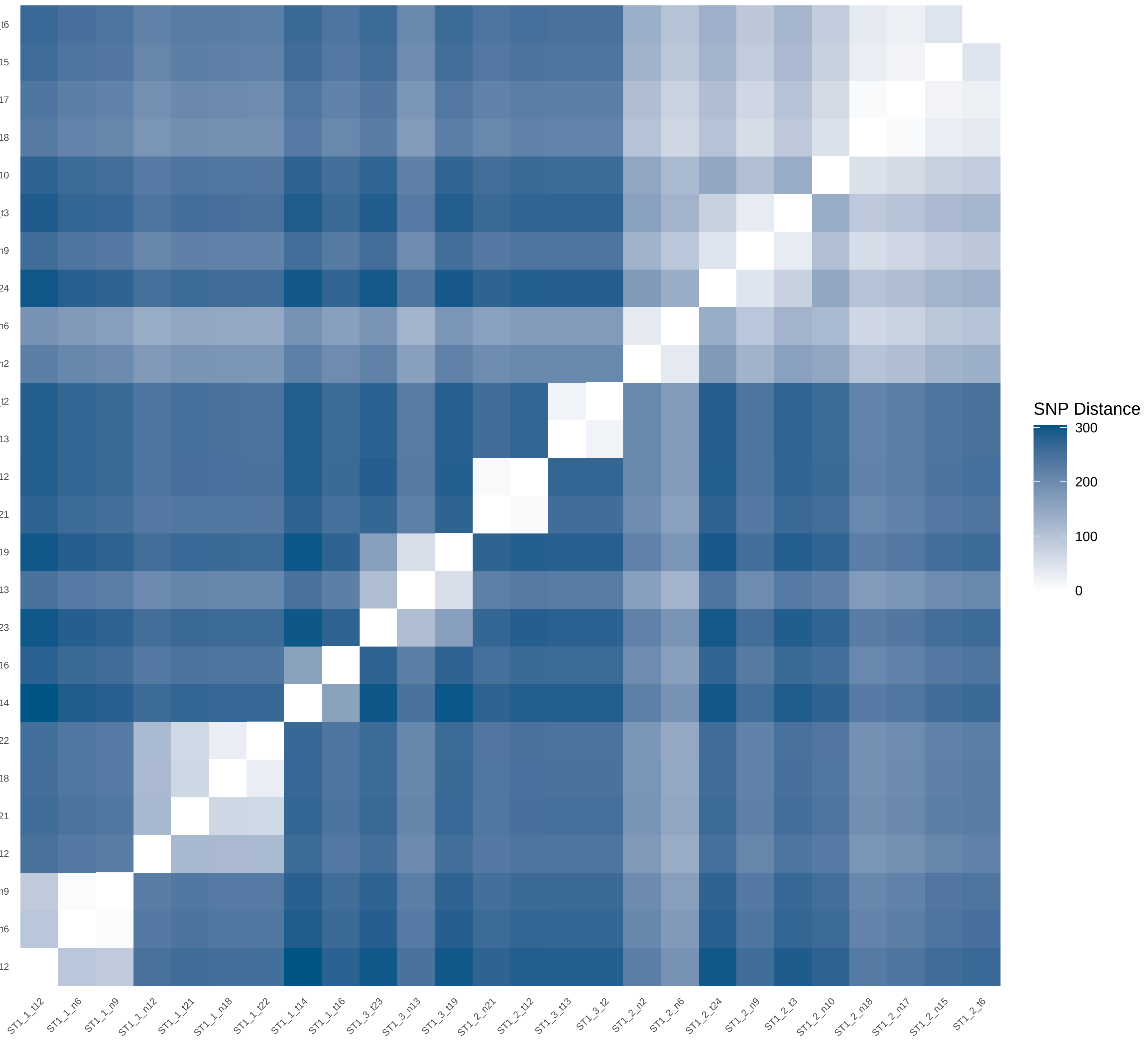
