## Supplementary Data 2 for "THRESHER: A tool for evidence-based, phylogeny-corrected SNP thresholds for whole genome molecular epidemiology": ST1_40_iter_7.pdf

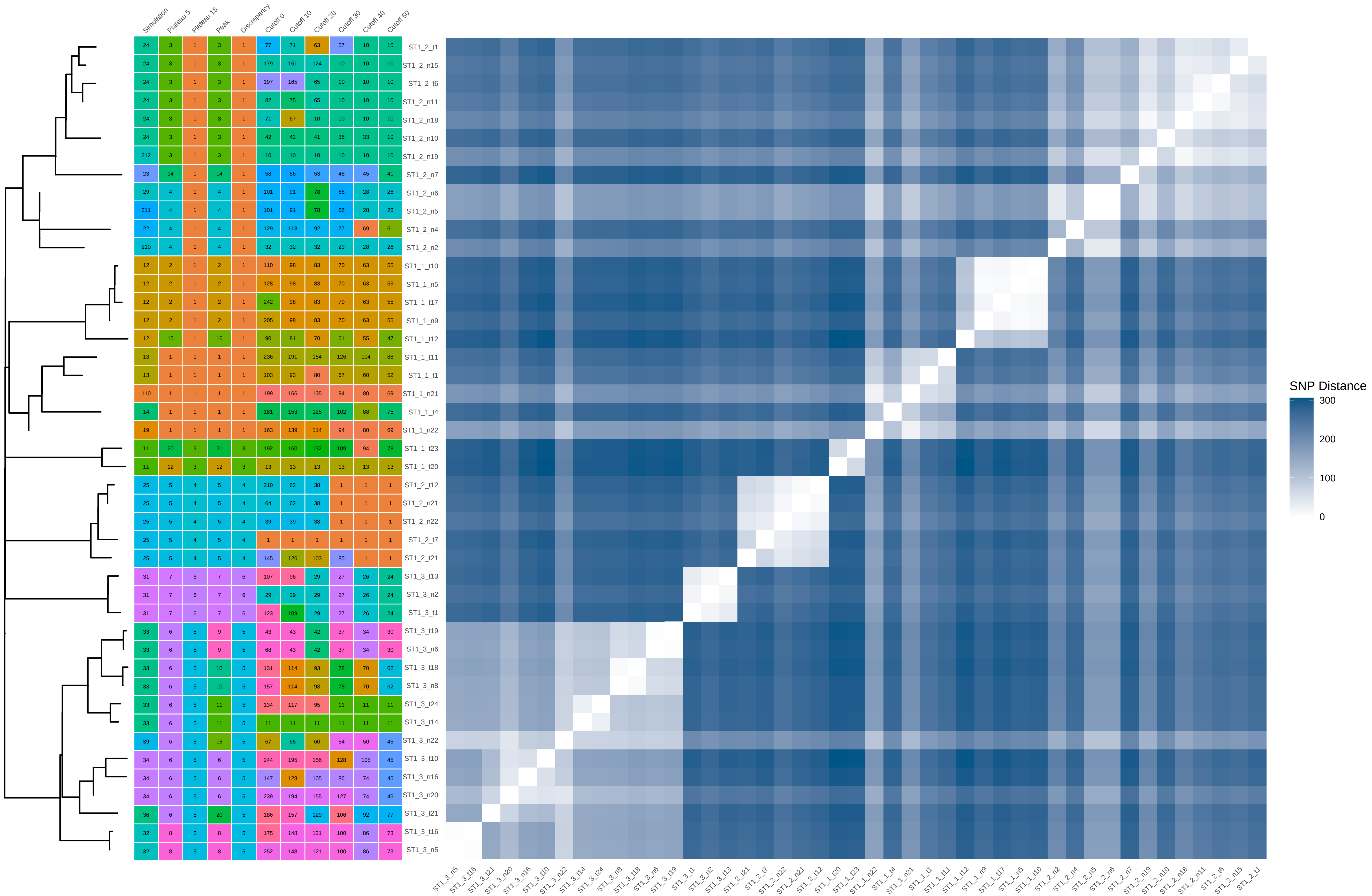


| Simulation | Plateau 5 | Plateau 15 | Peak | Discrepancy | Cutoff 0 | Cutoff 10 | Cutoff 20 | Cutoff 30 | Cutoff 40 | Cutoff 50 | |
|---|---|---|---|---|---|---|---|---|---|---|---|
| 24 | 3 | 1 | 3 | 1 | 77 | 71 | 63 | 57 | 10 | 10 | ST1_2_t1 |
| 24 | 3 | 1 | 3 | 1 | 179 | 151 | 124 | 10 | 10 | 10 | ST1_2_n15 |
| 24 | 3 | 1 | 3 | 1 | 197 | 165 | 65 | 10 | 10 | 10 | ST1_2_t6 |
| 24 | 3 | 1 | 3 | 1 | 82 | 75 | 65 | 10 | 10 | 10 | ST1_2_n11 |
| 24 | 3 | 1 | 3 | 1 | 71 | 67 | 10 | 10 | 10 | 10 | ST1_2_n18 |
| 24 | 3 | 1 | 3 | 1 | 42 | 42 | 41 | 36 | 33 | 10 | ST1_2_n10 |
| 212 | 3 | 1 | 3 | 1 | 10 | 10 | 10 | 10 | 10 | 10 | ST1_2_n19 |
| 23 | 14 | 1 | 14 | 1 | 58 | 56 | 53 | 48 | 45 | 41 | ST1_2_n7 |
| 29 | 4 | 1 | 4 | 1 | 101 | 91 | 78 | 66 | 28 | 26 | ST1_2_n6 |
| 211 | 4 | 1 | 4 | 1 | 101 | 91 | 78 | 66 | 28 | 26 | ST1_2_n5 |
| 22 | 4 | 1 | 4 | 1 | 129 | 113 | 92 | 77 | 69 | 61 | ST1_2_n4 |
| 210 | 4 | 1 | 4 | 1 | 32 | 32 | 32 | 29 | 28 | 26 | ST1_2_n2 |
| 12 | 2 | 1 | 2 | 1 | 110 | 98 | 83 | 70 | 63 | 55 | ST1_1_t10 |
| 12 | 2 | 1 | 2 | 1 | 128 | 98 | 83 | 70 | 63 | 55 | ST1_1_n5 |
| 12 | 2 | 1 | 2 | 1 | 242 | 98 | 83 | 70 | 63 | 55 | ST1_1_t17 |
| 12 | 2 | 1 | 2 | 1 | 205 | 98 | 83 | 70 | 63 | 55 | ST1_1_n9 |
| 12 | 15 | 1 | 16 | 1 | 90 | 81 | 70 | 61 | 55 | 47 | ST1_1_t12 |
| 13 | 1 | 1 | 1 | 1 | 236 | 191 | 154 | 126 | 104 | 88 | ST1_1_t11 |
| 13 | 1 | 1 | 1 | 1 | 103 | 93 | 80 | 67 | 60 | 52 | ST1_1_t1 |
| 110 | 1 | 1 | 1 | 1 | 199 | 166 | 135 | 94 | 80 | 69 | ST1_1_n21 |
| 14 | 1 | 1 | 1 | 1 | 181 | 153 | 125 | 102 | 88 | 75 | ST1_1_t4 |
| 19 | 1 | 1 | 1 | 1 | 163 | 139 | 114 | 94 | 80 | 69 | ST1_1_n22 |
| 11 | 20 | 3 | 21 | 3 | 192 | 160 | 132 | 109 | 94 | 78 | ST1_1_t23 |
| 11 | 12 | 3 | 12 | 3 | 13 | 13 | 13 | 13 | 13 | 13 | ST1_1_t20 |
| 25 | 5 | 4 | 5 | 4 | 210 | 62 | 38 | 1 | 1 | 1 | ST1_2_t12 |
| 25 | 5 | 4 | 5 | 4 | 64 | 62 | 38 | 1 | 1 | 1 | ST1_2_n21 |
| 25 | 5 | 4 | 5 | 4 | 39 | 39 | 38 | 1 | 1 | 1 | ST1_2_n22 |
| 25 | 5 | 4 | 5 | 4 | 1 | 1 | 1 | 1 | 1 | 1 | ST1_2_t7 |
| 25 | 5 | 4 | 5 | 4 | 145 | 126 | 103 | 85 | 1 | 1 | ST1_2_t21 |
| 31 | 7 | 6 | 7 | 6 | 107 | 96 | 29 | 27 | 26 | 24 | ST1_3_t13 |
| 31 | 7 | 6 | 7 | 6 | 29 | 29 | 29 | 27 | 26 | 24 | ST1_3_n2 |
| 31 | 7 | 6 | 7 | 6 | 123 | 109 | 29 | 27 | 26 | 24 | ST1_3_t1 |
| 33 | 6 | 5 | 9 | 5 | 43 | 43 | 42 | 37 | 34 | 30 | ST1_3_t19 |
| 33 | 6 | 5 | 9 | 5 | 68 | 43 | 42 | 37 | 34 | 30 | ST1_3_n6 |
| 33 | 6 | 5 | 10 | 5 | 131 | 114 | 93 | 78 | 70 | 62 | ST1_3_t18 |
| 33 | 6 | 5 | 10 | 5 | 157 | 114 | 93 | 78 | 70 | 62 | ST1_3_n8 |
| 33 | 6 | 5 | 11 | 5 | 134 | 117 | 95 | 11 | 11 | 11 | ST1_3_t24 |
| 33 | 6 | 5 | 11 | 5 | 11 | 11 | 11 | 11 | 11 | 11 | ST1_3_t14 |
| 39 | 6 | 5 | 15 | 5 | 67 | 65 | 60 | 54 | 50 | 45 | ST1_3_n22 |
| 34 | 6 | 5 | 6 | 5 | 244 | 195 | 156 | 128 | 105 | 45 | ST1_3_t10 |
| 34 | 6 | 5 | 6 | 5 | 147 | 128 | 105 | 86 | 74 | 45 | ST1_3_n16 |
| 34 | 6 | 5 | 6 | 5 | 239 | 194 | 155 | 127 | 74 | 45 | ST1_3_n20 |
| 36 | 6 | 5 | 20 | 5 | 186 | 157 | 129 | 106 | 92 | 77 | ST1_3_t21 |
| 32 | 8 | 5 | 8 | 5 | 175 | 148 | 121 | 100 | 86 | 73 | ST1_3_t16 |
| 32 | 8 | 5 | 8 | 5 | 252 | 148 | 121 | 100 | 86 | 73 | ST1_3_n5 |
