## Supplementary Data 2 for "THRESHER: A tool for evidence-based, phylogeny-corrected SNP thresholds for whole genome molecular epidemiology": ST1_60_iter_10.pdf

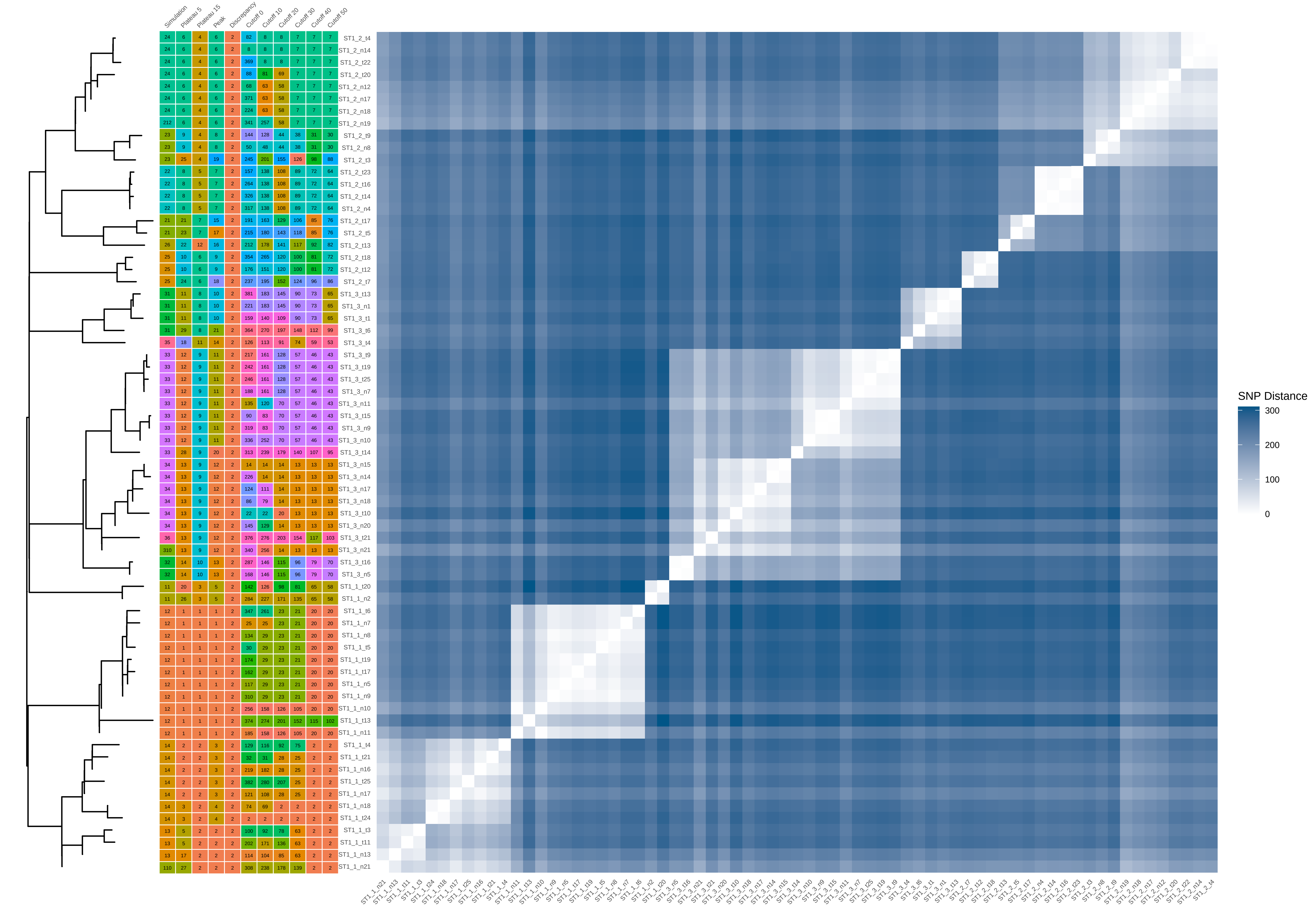


| Sample | Simulation | Plateau 5 | Plateau 15 | Peak | Discrepancy | Cutoff 0 | Cutoff 10 | Cutoff 20 | Cutoff 30 | Cutoff 40 | Cutoff 50 |
|---|---|---|---|---|---|---|---|---|---|---|---|
| ST1_2_t4 | 24 | 6 | 4 | 6 | 2 | 82 | 8 | 8 | 7 | 7 | 7 |
| ST1_2_n14 | 24 | 6 | 4 | 6 | 2 | 8 | 8 | 8 | 7 | 7 | 7 |
| ST1_2_t22 | 24 | 6 | 4 | 6 | 2 | 369 | 8 | 8 | 7 | 7 | 7 |
| ST1_2_t20 | 24 | 6 | 4 | 6 | 2 | 88 | 81 | 69 | 7 | 7 | 7 |
| ST1_2_n12 | 24 | 6 | 4 | 6 | 2 | 68 | 63 | 58 | 7 | 7 | 7 |
| ST1_2_n17 | 24 | 6 | 4 | 6 | 2 | 371 | 63 | 58 | 7 | 7 | 7 |
| ST1_2_n18 | 24 | 6 | 4 | 6 | 2 | 224 | 63 | 58 | 7 | 7 | 7 |
| ST1_2_n19 | 212 | 6 | 4 | 6 | 2 | 341 | 257 | 58 | 7 | 7 | 7 |
| ST1_2_t9 | 23 | 9 | 4 | 8 | 2 | 144 | 128 | 44 | 38 | 31 | 30 |
| ST1_2_n8 | 23 | 9 | 4 | 8 | 2 | 50 | 48 | 44 | 38 | 31 | 30 |
| ST1_2_t3 | 23 | 25 | 4 | 19 | 2 | 245 | 201 | 155 | 126 | 98 | 88 |
| ST1_2_t23 | 22 | 8 | 5 | 7 | 2 | 157 | 138 | 108 | 89 | 72 | 64 |
| ST1_2_t16 | 22 | 8 | 5 | 7 | 2 | 264 | 138 | 108 | 89 | 72 | 64 |
| ST1_2_t14 | 22 | 8 | 5 | 7 | 2 | 326 | 138 | 108 | 89 | 72 | 64 |
| ST1_2_n4 | 22 | 8 | 5 | 7 | 2 | 317 | 138 | 108 | 89 | 72 | 64 |
| ST1_2_t17 | 21 | 21 | 7 | 15 | 2 | 191 | 163 | 129 | 106 | 85 | 76 |
| ST1_2_t5 | 21 | 23 | 7 | 17 | 2 | 215 | 180 | 143 | 118 | 85 | 76 |
| ST1_2_t13 | 26 | 22 | 12 | 16 | 2 | 212 | 178 | 141 | 117 | 92 | 82 |
| ST1_2_t18 | 25 | 10 | 6 | 9 | 2 | 354 | 265 | 120 | 100 | 81 | 72 |
| ST1_2_t12 | 25 | 10 | 6 | 9 | 2 | 176 | 151 | 120 | 100 | 81 | 72 |
| ST1_2_t7 | 25 | 24 | 6 | 18 | 2 | 237 | 195 | 152 | 124 | 96 | 86 |
| ST1_3_t13 | 31 | 11 | 8 | 10 | 2 | 381 | 183 | 145 | 90 | 73 | 65 |
| ST1_3_n1 | 31 | 11 | 8 | 10 | 2 | 221 | 183 | 145 | 90 | 73 | 65 |
| ST1_3_t1 | 31 | 11 | 8 | 10 | 2 | 159 | 140 | 109 | 90 | 73 | 65 |
| ST1_3_t6 | 31 | 29 | 8 | 21 | 2 | 364 | 270 | 197 | 148 | 112 | 99 |
| ST1_3_t4 | 35 | 18 | 11 | 14 | 2 | 126 | 113 | 91 | 74 | 59 | 53 |
| ST1_3_t9 | 33 | 12 | 9 | 11 | 2 | 217 | 161 | 128 | 57 | 46 | 43 |
| ST1_3_t19 | 33 | 12 | 9 | 11 | 2 | 242 | 161 | 128 | 57 | 46 | 43 |
| ST1_3_t25 | 33 | 12 | 9 | 11 | 2 | 246 | 161 | 128 | 57 | 46 | 43 |
| ST1_3_n7 | 33 | 12 | 9 | 11 | 2 | 188 | 161 | 128 | 57 | 46 | 43 |
| ST1_3_n11 | 33 | 12 | 9 | 11 | 2 | 135 | 120 | 70 | 57 | 46 | 43 |
| ST1_3_t15 | 33 | 12 | 9 | 11 | 2 | 90 | 83 | 70 | 57 | 46 | 43 |
| ST1_3_n9 | 33 | 12 | 9 | 11 | 2 | 319 | 83 | 70 | 57 | 46 | 43 |
| ST1_3_n10 | 33 | 12 | 9 | 11 | 2 | 336 | 252 | 70 | 57 | 46 | 43 |
| ST1_3_t14 | 33 | 28 | 9 | 20 | 2 | 313 | 239 | 179 | 140 | 107 | 95 |
| ST1_3_n15 | 34 | 13 | 9 | 12 | 2 | 14 | 14 | 14 | 13 | 13 | 13 |
| ST1_3_n14 | 34 | 13 | 9 | 12 | 2 | 226 | 14 | 14 | 13 | 13 | 13 |
| ST1_3_n17 | 34 | 13 | 9 | 12 | 2 | 124 | 111 | 14 | 13 | 13 | 13 |
| ST1_3_n18 | 34 | 13 | 9 | 12 | 2 | 86 | 79 | 14 | 13 | 13 | 13 |
| ST1_3_t10 | 34 | 13 | 9 | 12 | 2 | 22 | 22 | 20 | 13 | 13 | 13 |
| ST1_3_n20 | 34 | 13 | 9 | 12 | 2 | 145 | 129 | 14 | 13 | 13 | 13 |
| ST1_3_t21 | 36 | 13 | 9 | 12 | 2 | 376 | 276 | 203 | 154 | 117 | 103 |
| ST1_3_n21 | 310 | 13 | 9 | 12 | 2 | 340 | 256 | 14 | 13 | 13 | 13 |
| ST1_3_t16 | 32 | 14 | 10 | 13 | 2 | 287 | 146 | 115 | 96 | 79 | 70 |
| ST1_3_n5 | 32 | 14 | 10 | 13 | 2 | 168 | 146 | 115 | 96 | 79 | 70 |
| ST1_1_t20 | 11 | 20 | 3 | 5 | 2 | 142 | 126 | 98 | 81 | 65 | 58 |
| ST1_1_n2 | 11 | 26 | 3 | 5 | 2 | 284 | 227 | 171 | 135 | 65 | 58 |
| ST1_1_t6 | 12 | 1 | 1 | 1 | 2 | 347 | 261 | 23 | 21 | 20 | 20 |
| ST1_1_n7 | 12 | 1 | 1 | 1 | 2 | 25 | 25 | 23 | 21 | 20 | 20 |
| ST1_1_n8 | 12 | 1 | 1 | 1 | 2 | 134 | 29 | 23 | 21 | 20 | 20 |
| ST1_1_t5 | 12 | 1 | 1 | 1 | 2 | 30 | 29 | 23 | 21 | 20 | 20 |
| ST1_1_t19 | 12 | 1 | 1 | 1 | 2 | 174 | 29 | 23 | 21 | 20 | 20 |
| ST1_1_t17 | 12 | 1 | 1 | 1 | 2 | 162 | 29 | 23 | 21 | 20 | 20 |
| ST1_1_n5 | 12 | 1 | 1 | 1 | 2 | 117 | 29 | 23 | 21 | 20 | 20 |
| ST1_1_n9 | 12 | 1 | 1 | 1 | 2 | 310 | 29 | 23 | 21 | 20 | 20 |
| ST1_1_n10 | 12 | 1 | 1 | 1 | 2 | 256 | 158 | 126 | 105 | 20 | 20 |
| ST1_1_t13 | 12 | 1 | 1 | 1 | 2 | 374 | 274 | 201 | 152 | 115 | 102 |
| ST1_1_n11 | 12 | 1 | 1 | 1 | 2 | 185 | 158 | 126 | 105 | 20 | 20 |
| ST1_1_t4 | 14 | 2 | 2 | 3 | 2 | 129 | 116 | 92 | 75 | 2 | 2 |
| ST1_1_t21 | 14 | 2 | 2 | 3 | 2 | 32 | 31 | 28 | 25 | 2 | 2 |
| ST1_1_n16 | 14 | 2 | 2 | 3 | 2 | 219 | 182 | 28 | 25 | 2 | 2 |
| ST1_1_t25 | 14 | 2 | 2 | 3 | 2 | 382 | 280 | 207 | 25 | 2 | 2 |
| ST1_1_n17 | 14 | 2 | 2 | 3 | 2 | 121 | 108 | 28 | 25 | 2 | 2 |
| ST1_1_n18 | 14 | 3 | 2 | 4 | 2 | 74 | 69 | 2 | 2 | 2 | 2 |
| ST1_1_t24 | 14 | 3 | 2 | 4 | 2 | 2 | 2 | 2 | 2 | 2 | 2 |
| ST1_1_t3 | 13 | 5 | 2 | 2 | 2 | 100 | 92 | 78 | 63 | 2 | 2 |
| ST1_1_t11 | 13 | 5 | 2 | 2 | 2 | 202 | 171 | 136 | 63 | 2 | 2 |
| ST1_1_n13 | 13 | 17 | 2 | 2 | 2 | 114 | 104 | 85 | 63 | 2 | 2 |
| ST1_1_n21 | 110 | 27 | 2 | 2 | 2 | 308 | 238 | 178 | 139 | 2 | 2 |
