## Supplementary Data 2 for "THRESHER: A tool for evidence-based, phylogeny-corrected SNP thresholds for whole genome molecular epidemiology": ST5_20_iter_3.pdf

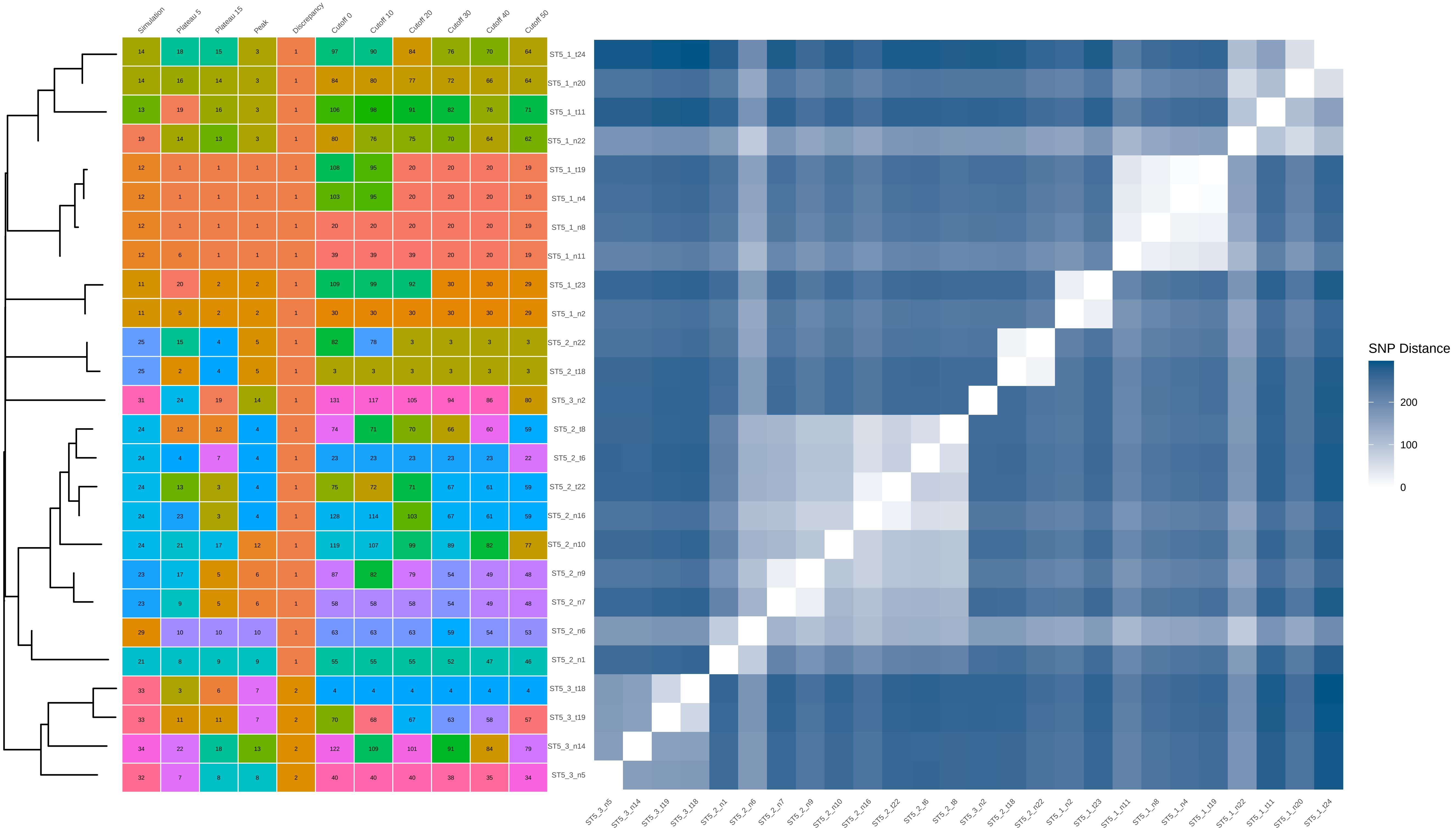


| | Simulation | Plateau 5 | Plateau 15 | Peak | Discrepancy | Cutoff 0 | Cutoff 10 | Cutoff 20 | Cutoff 30 | Cutoff 40 | Cutoff 50 |
|---|---|---|---|---|---|---|---|---|---|---|---|
| ST5_1_t24 | 14 | 18 | 15 | 3 | 1 | 97 | 90 | 84 | 76 | 70 | 64 |
| ST5_1_n20 | 14 | 16 | 14 | 3 | 1 | 84 | 80 | 77 | 72 | 66 | 64 |
| ST5_1_t11 | 13 | 19 | 16 | 3 | 1 | 106 | 98 | 91 | 82 | 76 | 71 |
| ST5_1_n22 | 19 | 14 | 13 | 3 | 1 | 80 | 76 | 75 | 70 | 64 | 62 |
| ST5_1_t19 | 12 | 1 | 1 | 1 | 1 | 108 | 95 | 20 | 20 | 20 | 19 |
| ST5_1_n4 | 12 | 1 | 1 | 1 | 1 | 103 | 95 | 20 | 20 | 20 | 19 |
| ST5_1_n8 | 12 | 1 | 1 | 1 | 1 | 20 | 20 | 20 | 20 | 20 | 19 |
| ST5_1_n11 | 12 | 6 | 1 | 1 | 1 | 39 | 39 | 39 | 20 | 20 | 19 |
| ST5_1_t23 | 11 | 20 | 2 | 2 | 1 | 109 | 99 | 92 | 30 | 30 | 29 |
| ST5_1_n2 | 11 | 5 | 2 | 2 | 1 | 30 | 30 | 30 | 30 | 30 | 29 |
| ST5_2_n22 | 25 | 15 | 4 | 5 | 1 | 82 | 78 | 3 | 3 | 3 | 3 |
| ST5_2_t18 | 25 | 2 | 4 | 5 | 1 | 3 | 3 | 3 | 3 | 3 | 3 |
| ST5_3_n2 | 31 | 24 | 19 | 14 | 1 | 131 | 117 | 105 | 94 | 86 | 80 |
| ST5_2_t8 | 24 | 12 | 12 | 4 | 1 | 74 | 71 | 70 | 66 | 60 | 59 |
| ST5_2_t6 | 24 | 4 | 7 | 4 | 1 | 23 | 23 | 23 | 23 | 23 | 22 |
| ST5_2_t22 | 24 | 13 | 3 | 4 | 1 | 75 | 72 | 71 | 67 | 61 | 59 |
| ST5_2_n16 | 24 | 23 | 3 | 4 | 1 | 128 | 114 | 103 | 67 | 61 | 59 |
| ST5_2_n10 | 24 | 21 | 17 | 12 | 1 | 119 | 107 | 99 | 89 | 82 | 77 |
| ST5_2_n9 | 23 | 17 | 5 | 6 | 1 | 87 | 82 | 79 | 54 | 49 | 48 |
| ST5_2_n7 | 23 | 9 | 5 | 6 | 1 | 58 | 58 | 58 | 54 | 49 | 48 |
| ST5_2_n6 | 29 | 10 | 10 | 10 | 1 | 63 | 63 | 63 | 59 | 54 | 53 |
| ST5_2_n1 | 21 | 8 | 9 | 9 | 1 | 55 | 55 | 55 | 52 | 47 | 46 |
| ST5_3_t18 | 33 | 3 | 6 | 7 | 2 | 4 | 4 | 4 | 4 | 4 | 4 |
| ST5_3_t19 | 33 | 11 | 11 | 7 | 2 | 70 | 68 | 67 | 63 | 58 | 57 |
| ST5_3_n14 | 34 | 22 | 18 | 13 | 2 | 122 | 109 | 101 | 91 | 84 | 79 |
| ST5_3_n5 | 32 | 7 | 8 | 8 | 2 | 40 | 40 | 40 | 38 | 35 | 34 |
