## Supplementary Data 2 for "THRESHER: A tool for evidence-based, phylogeny-corrected SNP thresholds for whole genome molecular epidemiology": ST5_20_iter_4.pdf

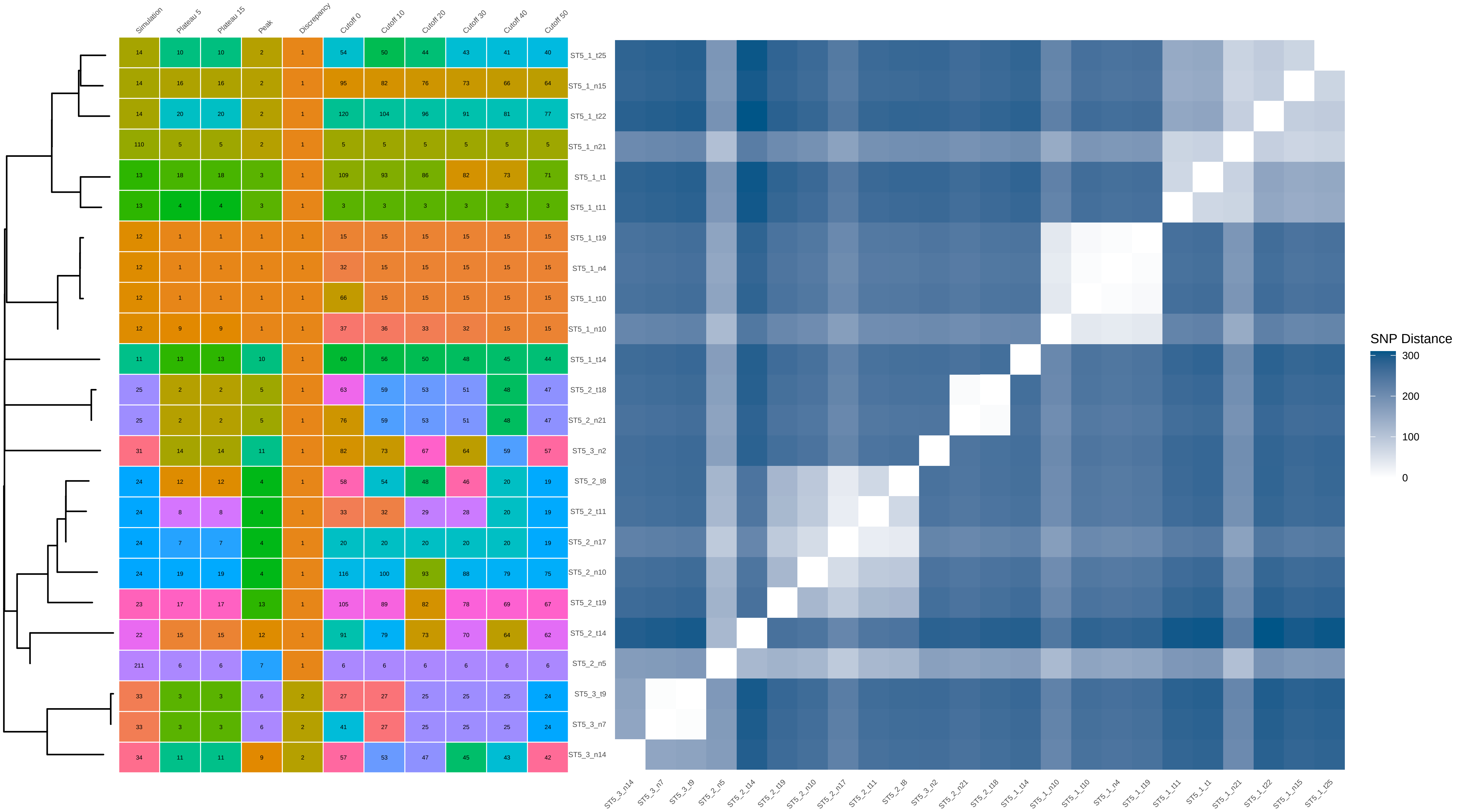


| | Simulation | Plateau 5 | Plateau 15 | Peak | Discrepancy | Cutoff 0 | Cutoff 10 | Cutoff 20 | Cutoff 30 | Cutoff 40 | Cutoff 50 |
|---|---|---|---|---|---|---|---|---|---|---|---|
| ST5_1_t25 | 14 | 10 | 10 | 2 | 1 | 54 | 50 | 44 | 43 | 41 | 40 |
| ST5_1_n15 | 14 | 16 | 16 | 2 | 1 | 95 | 82 | 76 | 73 | 66 | 64 |
| ST5_1_t22 | 14 | 20 | 20 | 2 | 1 | 120 | 104 | 96 | 91 | 81 | 77 |
| ST5_1_n21 | 110 | 5 | 5 | 2 | 1 | 5 | 5 | 5 | 5 | 5 | 5 |
| ST5_1_t1 | 13 | 18 | 18 | 3 | 1 | 109 | 93 | 86 | 82 | 73 | 71 |
| ST5_1_t11 | 13 | 4 | 4 | 3 | 1 | 3 | 3 | 3 | 3 | 3 | 3 |
| ST5_1_t19 | 12 | 1 | 1 | 1 | 1 | 15 | 15 | 15 | 15 | 15 | 15 |
| ST5_1_n4 | 12 | 1 | 1 | 1 | 1 | 32 | 15 | 15 | 15 | 15 | 15 |
| ST5_1_t10 | 12 | 1 | 1 | 1 | 1 | 66 | 15 | 15 | 15 | 15 | 15 |
| ST5_1_n10 | 12 | 9 | 9 | 1 | 1 | 37 | 36 | 33 | 32 | 15 | 15 |
| ST5_1_t14 | 11 | 13 | 13 | 10 | 1 | 60 | 56 | 50 | 48 | 45 | 44 |
| ST5_2_t18 | 25 | 2 | 2 | 5 | 1 | 63 | 59 | 53 | 51 | 48 | 47 |
| ST5_2_n21 | 25 | 2 | 2 | 5 | 1 | 76 | 59 | 53 | 51 | 48 | 47 |
| ST5_3_n2 | 31 | 14 | 14 | 11 | 1 | 82 | 73 | 67 | 64 | 59 | 57 |
| ST5_2_t8 | 24 | 12 | 12 | 4 | 1 | 58 | 54 | 48 | 46 | 20 | 19 |
| ST5_2_t11 | 24 | 8 | 8 | 4 | 1 | 33 | 32 | 29 | 28 | 20 | 19 |
| ST5_2_n17 | 24 | 7 | 7 | 4 | 1 | 20 | 20 | 20 | 20 | 20 | 19 |
| ST5_2_n10 | 24 | 19 | 19 | 4 | 1 | 116 | 100 | 93 | 88 | 79 | 75 |
| ST5_2_t19 | 23 | 17 | 17 | 13 | 1 | 105 | 89 | 82 | 78 | 69 | 67 |
| ST5_2_t14 | 22 | 15 | 15 | 12 | 1 | 91 | 79 | 73 | 70 | 64 | 62 |
| ST5_2_n5 | 211 | 6 | 6 | 7 | 1 | 6 | 6 | 6 | 6 | 6 | 6 |
| ST5_3_t9 | 33 | 3 | 3 | 6 | 2 | 27 | 27 | 25 | 25 | 25 | 24 |
| ST5_3_n7 | 33 | 3 | 3 | 6 | 2 | 41 | 27 | 25 | 25 | 25 | 24 |
| ST5_3_n14 | 34 | 11 | 11 | 9 | 2 | 57 | 53 | 47 | 45 | 43 | 42 |
