## Supplementary Data 2 for "THRESHER: A tool for evidence-based, phylogeny-corrected SNP thresholds for whole genome molecular epidemiology": ST5_20_iter_6.pdf

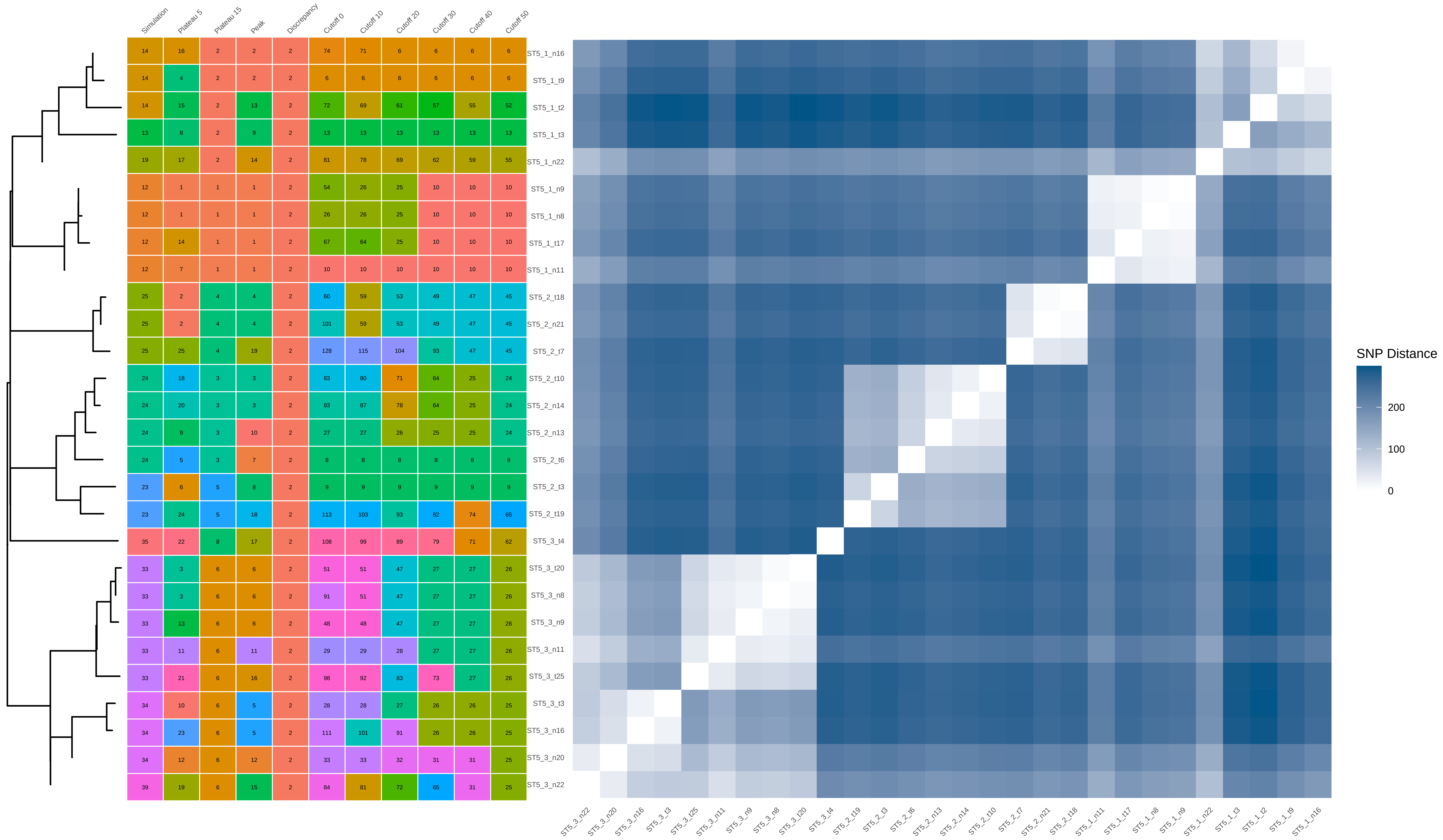


| | Simulation | Plateau 5 | Plateau 15 | Peak | Discrepancy | Cutoff 0 | Cutoff 10 | Cutoff 20 | Cutoff 30 | Cutoff 40 | Cutoff 50 |
|---|---|---|---|---|---|---|---|---|---|---|---|
| ST5_1_n16 | 14 | 16 | 2 | 2 | 2 | 74 | 71 | 6 | 6 | 6 | 6 |
| ST5_1_t9 | 14 | 4 | 2 | 2 | 2 | 6 | 6 | 6 | 6 | 6 | 6 |
| ST5_1_t2 | 14 | 15 | 2 | 13 | 2 | 72 | 69 | 61 | 57 | 55 | 52 |
| ST5_1_t3 | 13 | 8 | 2 | 9 | 2 | 13 | 13 | 13 | 13 | 13 | 13 |
| ST5_1_n22 | 19 | 17 | 2 | 14 | 2 | 81 | 78 | 69 | 62 | 59 | 55 |
| ST5_1_n9 | 12 | 1 | 1 | 1 | 2 | 54 | 26 | 25 | 10 | 10 | 10 |
| ST5_1_n8 | 12 | 1 | 1 | 1 | 2 | 26 | 26 | 25 | 10 | 10 | 10 |
| ST5_1_t17 | 12 | 14 | 1 | 1 | 2 | 67 | 64 | 25 | 10 | 10 | 10 |
| ST5_1_n11 | 12 | 7 | 1 | 1 | 2 | 10 | 10 | 10 | 10 | 10 | 10 |
| ST5_2_t18 | 25 | 2 | 4 | 4 | 2 | 60 | 59 | 53 | 49 | 47 | 45 |
| ST5_2_n21 | 25 | 2 | 4 | 4 | 2 | 101 | 59 | 53 | 49 | 47 | 45 |
| ST5_2_t7 | 25 | 25 | 4 | 19 | 2 | 128 | 115 | 104 | 93 | 47 | 45 |
| ST5_2_t10 | 24 | 18 | 3 | 3 | 2 | 83 | 80 | 71 | 64 | 25 | 24 |
| ST5_2_n14 | 24 | 20 | 3 | 3 | 2 | 93 | 87 | 78 | 64 | 25 | 24 |
| ST5_2_n13 | 24 | 9 | 3 | 10 | 2 | 27 | 27 | 26 | 25 | 25 | 24 |
| ST5_2_t6 | 24 | 5 | 3 | 7 | 2 | 8 | 8 | 8 | 8 | 8 | 8 |
| ST5_2_t3 | 23 | 6 | 5 | 8 | 2 | 9 | 9 | 9 | 9 | 9 | 9 |
| ST5_2_t19 | 23 | 24 | 5 | 18 | 2 | 113 | 103 | 93 | 82 | 74 | 65 |
| ST5_3_t4 | 35 | 22 | 8 | 17 | 2 | 108 | 99 | 89 | 79 | 71 | 62 |
| ST5_3_t20 | 33 | 3 | 6 | 6 | 2 | 51 | 51 | 47 | 27 | 27 | 26 |
| ST5_3_n8 | 33 | 3 | 6 | 6 | 2 | 91 | 51 | 47 | 27 | 27 | 26 |
| ST5_3_n9 | 33 | 13 | 6 | 6 | 2 | 48 | 48 | 47 | 27 | 27 | 26 |
| ST5_3_n11 | 33 | 11 | 6 | 11 | 2 | 29 | 29 | 28 | 27 | 27 | 26 |
| ST5_3_t25 | 33 | 21 | 6 | 16 | 2 | 98 | 92 | 83 | 73 | 27 | 26 |
| ST5_3_t3 | 34 | 10 | 6 | 5 | 2 | 28 | 28 | 27 | 26 | 26 | 25 |
| ST5_3_n16 | 34 | 23 | 6 | 5 | 2 | 111 | 101 | 91 | 26 | 26 | 25 |
| ST5_3_n20 | 34 | 12 | 6 | 12 | 2 | 33 | 33 | 32 | 31 | 31 | 25 |
| ST5_3_n22 | 39 | 19 | 6 | 15 | 2 | 84 | 81 | 72 | 65 | 31 | 25 |
