## Supplementary Data 2 for "THRESHER: A tool for evidence-based, phylogeny-corrected SNP thresholds for whole genome molecular epidemiology": ST5_20_iter_7.pdf

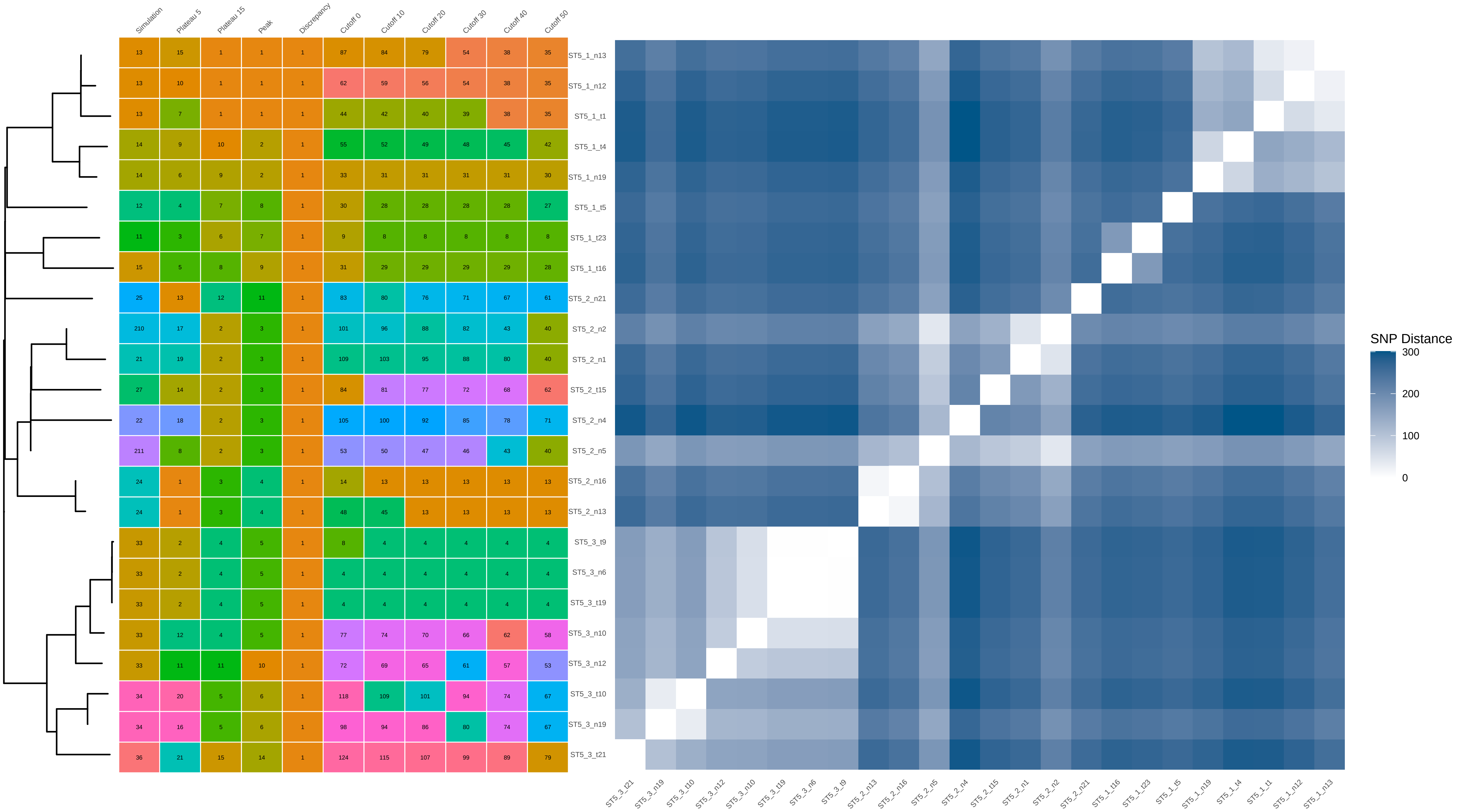


| | Simulation | Plateau 5 | Plateau 15 | Peak | Discrepancy | Cutoff 0 | Cutoff 10 | Cutoff 20 | Cutoff 30 | Cutoff 40 | Cutoff 50 |
|---|---|---|---|---|---|---|---|---|---|---|---|
| ST5_1_n13 | 13 | 15 | 1 | 1 | 1 | 87 | 84 | 79 | 54 | 38 | 35 |
| ST5_1_n12 | 13 | 10 | 1 | 1 | 1 | 62 | 59 | 56 | 54 | 38 | 35 |
| ST5_1_t1 | 13 | 7 | 1 | 1 | 1 | 44 | 42 | 40 | 39 | 38 | 35 |
| ST5_1_t4 | 14 | 9 | 10 | 2 | 1 | 55 | 52 | 49 | 48 | 45 | 42 |
| ST5_1_n19 | 14 | 6 | 9 | 2 | 1 | 33 | 31 | 31 | 31 | 31 | 30 |
| ST5_1_t5 | 12 | 4 | 7 | 8 | 1 | 30 | 28 | 28 | 28 | 28 | 27 |
| ST5_1_t23 | 11 | 3 | 6 | 7 | 1 | 9 | 8 | 8 | 8 | 8 | 8 |
| ST5_1_t16 | 15 | 5 | 8 | 9 | 1 | 31 | 29 | 29 | 29 | 29 | 28 |
| ST5_2_n21 | 25 | 13 | 12 | 11 | 1 | 83 | 80 | 76 | 71 | 67 | 61 |
| ST5_2_n2 | 210 | 17 | 2 | 3 | 1 | 101 | 96 | 88 | 82 | 43 | 40 |
| ST5_2_n1 | 21 | 19 | 2 | 3 | 1 | 109 | 103 | 95 | 88 | 80 | 40 |
| ST5_2_t15 | 27 | 14 | 2 | 3 | 1 | 84 | 81 | 77 | 72 | 68 | 62 |
| ST5_2_n4 | 22 | 18 | 2 | 3 | 1 | 105 | 100 | 92 | 85 | 78 | 71 |
| ST5_2_n5 | 211 | 8 | 2 | 3 | 1 | 53 | 50 | 47 | 46 | 43 | 40 |
| ST5_2_n16 | 24 | 1 | 3 | 4 | 1 | 14 | 13 | 13 | 13 | 13 | 13 |
| ST5_2_n13 | 24 | 1 | 3 | 4 | 1 | 48 | 45 | 13 | 13 | 13 | 13 |
| ST5_3_t9 | 33 | 2 | 4 | 5 | 1 | 8 | 4 | 4 | 4 | 4 | 4 |
| ST5_3_n6 | 33 | 2 | 4 | 5 | 1 | 4 | 4 | 4 | 4 | 4 | 4 |
| ST5_3_t19 | 33 | 2 | 4 | 5 | 1 | 4 | 4 | 4 | 4 | 4 | 4 |
| ST5_3_n10 | 33 | 12 | 4 | 5 | 1 | 77 | 74 | 70 | 66 | 62 | 58 |
| ST5_3_n12 | 33 | 11 | 11 | 10 | 1 | 72 | 69 | 65 | 61 | 57 | 53 |
| ST5_3_t10 | 34 | 20 | 5 | 6 | 1 | 118 | 109 | 101 | 94 | 74 | 67 |
| ST5_3_n19 | 34 | 16 | 5 | 6 | 1 | 98 | 94 | 86 | 80 | 74 | 67 |
| ST5_3_t21 | 36 | 21 | 15 | 14 | 1 | 124 | 115 | 107 | 99 | 89 | 79 |
