## Supplementary Data 2 for "THRESHER: A tool for evidence-based, phylogeny-corrected SNP thresholds for whole genome molecular epidemiology": ST5_40_iter_1.pdf

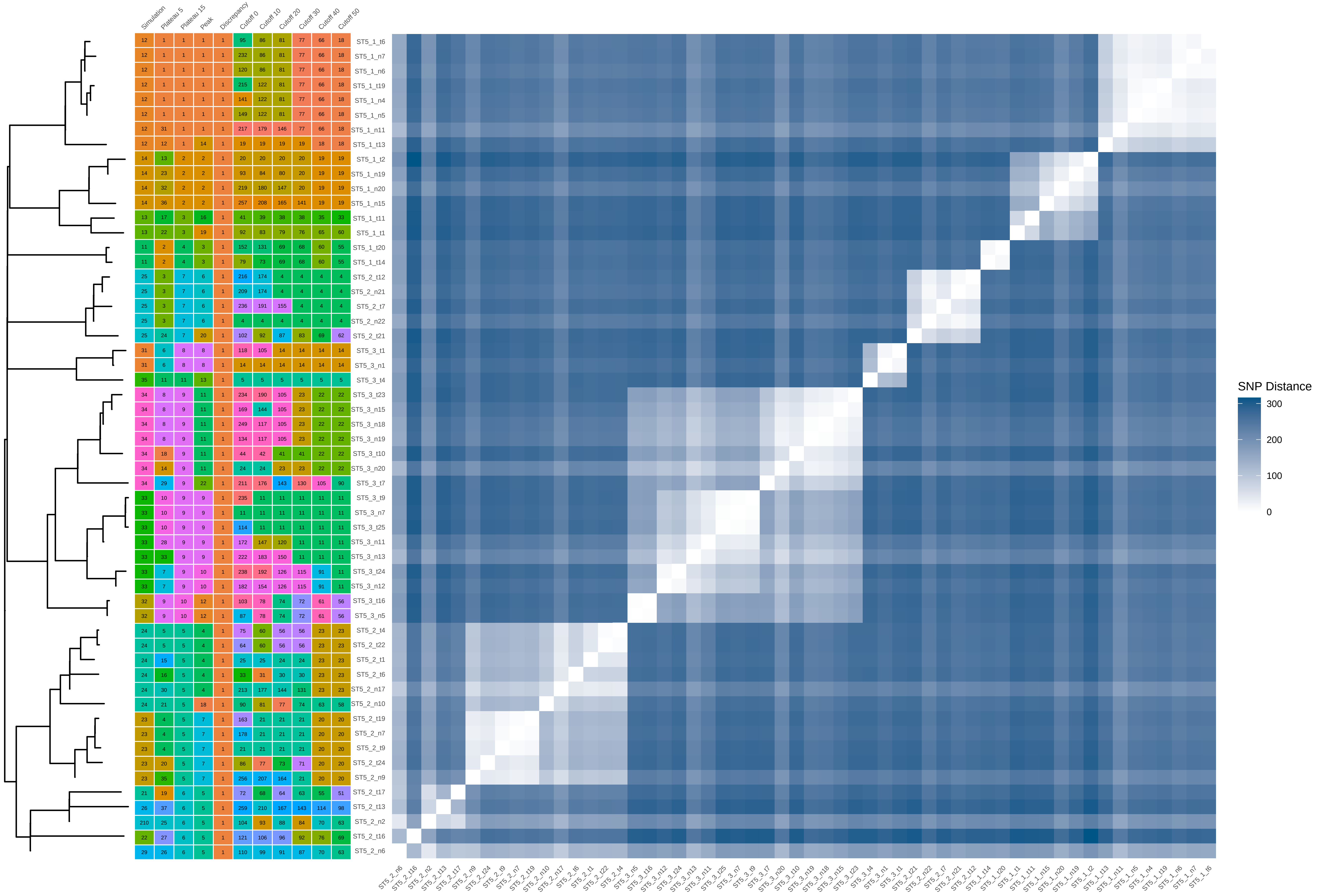


| | Simulation | Plateau 5 | Plateau 15 | Peak | Discrepancy | Cutoff 0 | Cutoff 10 | Cutoff 20 | Cutoff 30 | Cutoff 40 | Cutoff 50 |
|---|---|---|---|---|---|---|---|---|---|---|---|
| ST5_1_t6 | 12 | 1 | 1 | 1 | 1 | 95 | 86 | 81 | 77 | 66 | 18 |
| ST5_1_n7 | 12 | 1 | 1 | 1 | 1 | 232 | 86 | 81 | 77 | 66 | 18 |
| ST5_1_n6 | 12 | 1 | 1 | 1 | 1 | 120 | 86 | 81 | 77 | 66 | 18 |
| ST5_1_t19 | 12 | 1 | 1 | 1 | 1 | 215 | 122 | 81 | 77 | 66 | 18 |
| ST5_1_n4 | 12 | 1 | 1 | 1 | 1 | 141 | 122 | 81 | 77 | 66 | 18 |
| ST5_1_n5 | 12 | 1 | 1 | 1 | 1 | 149 | 122 | 81 | 77 | 66 | 18 |
| ST5_1_n11 | 12 | 31 | 1 | 1 | 1 | 217 | 179 | 146 | 77 | 66 | 18 |
| ST5_1_t13 | 12 | 12 | 1 | 14 | 1 | 19 | 19 | 19 | 19 | 18 | 18 |
| ST5_1_t2 | 14 | 13 | 2 | 2 | 1 | 20 | 20 | 20 | 20 | 19 | 19 |
| ST5_1_n19 | 14 | 23 | 2 | 2 | 1 | 93 | 84 | 80 | 20 | 19 | 19 |
| ST5_1_n20 | 14 | 32 | 2 | 2 | 1 | 219 | 180 | 147 | 20 | 19 | 19 |
| ST5_1_n15 | 14 | 36 | 2 | 2 | 1 | 257 | 208 | 165 | 141 | 19 | 19 |
| ST5_1_t11 | 13 | 17 | 3 | 16 | 1 | 41 | 39 | 38 | 38 | 35 | 33 |
| ST5_1_t1 | 13 | 22 | 3 | 19 | 1 | 92 | 83 | 79 | 76 | 65 | 60 |
| ST5_1_t20 | 11 | 2 | 4 | 3 | 1 | 152 | 131 | 69 | 68 | 60 | 55 |
| ST5_1_t14 | 11 | 2 | 4 | 3 | 1 | 79 | 73 | 69 | 68 | 60 | 55 |
| ST5_2_t12 | 25 | 3 | 7 | 6 | 1 | 216 | 174 | 4 | 4 | 4 | 4 |
| ST5_2_n21 | 25 | 3 | 7 | 6 | 1 | 209 | 174 | 4 | 4 | 4 | 4 |
| ST5_2_t7 | 25 | 3 | 7 | 6 | 1 | 236 | 191 | 155 | 4 | 4 | 4 |
| ST5_2_n22 | 25 | 3 | 7 | 6 | 1 | 4 | 4 | 4 | 4 | 4 | 4 |
| ST5_2_t21 | 25 | 24 | 7 | 20 | 1 | 102 | 92 | 87 | 83 | 69 | 62 |
| ST5_3_t1 | 31 | 6 | 8 | 8 | 1 | 118 | 105 | 14 | 14 | 14 | 14 |
| ST5_3_n1 | 31 | 6 | 8 | 8 | 1 | 14 | 14 | 14 | 14 | 14 | 14 |
| ST5_3_t4 | 35 | 11 | 11 | 13 | 1 | 5 | 5 | 5 | 5 | 5 | 5 |
| ST5_3_t23 | 34 | 8 | 9 | 11 | 1 | 234 | 190 | 105 | 23 | 22 | 22 |
| ST5_3_n15 | 34 | 8 | 9 | 11 | 1 | 169 | 144 | 105 | 23 | 22 | 22 |
| ST5_3_n18 | 34 | 8 | 9 | 11 | 1 | 249 | 117 | 105 | 23 | 22 | 22 |
| ST5_3_n19 | 34 | 8 | 9 | 11 | 1 | 134 | 117 | 105 | 23 | 22 | 22 |
| ST5_3_t10 | 34 | 18 | 9 | 11 | 1 | 44 | 42 | 41 | 41 | 22 | 22 |
| ST5_3_n20 | 34 | 14 | 9 | 11 | 1 | 24 | 24 | 23 | 23 | 22 | 22 |
| ST5_3_t7 | 34 | 29 | 9 | 22 | 1 | 211 | 176 | 143 | 130 | 105 | 90 |
| ST5_3_t9 | 33 | 10 | 9 | 9 | 1 | 235 | 11 | 11 | 11 | 11 | 11 |
| ST5_3_n7 | 33 | 10 | 9 | 9 | 1 | 11 | 11 | 11 | 11 | 11 | 11 |
| ST5_3_t25 | 33 | 10 | 9 | 9 | 1 | 114 | 11 | 11 | 11 | 11 | 11 |
| ST5_3_n11 | 33 | 28 | 9 | 9 | 1 | 172 | 147 | 120 | 11 | 11 | 11 |
| ST5_3_n13 | 33 | 33 | 9 | 9 | 1 | 222 | 183 | 150 | 11 | 11 | 11 |
| ST5_3_t24 | 33 | 7 | 9 | 10 | 1 | 238 | 192 | 126 | 115 | 91 | 11 |
| ST5_3_n12 | 33 | 7 | 9 | 10 | 1 | 182 | 154 | 126 | 115 | 91 | 11 |
| ST5_3_t16 | 32 | 9 | 10 | 12 | 1 | 103 | 78 | 74 | 72 | 61 | 56 |
| ST5_3_n5 | 32 | 9 | 10 | 12 | 1 | 87 | 78 | 74 | 72 | 61 | 56 |
| ST5_2_t4 | 24 | 5 | 5 | 4 | 1 | 75 | 60 | 56 | 56 | 23 | 23 |
| ST5_2_t22 | 24 | 5 | 5 | 4 | 1 | 64 | 60 | 56 | 56 | 23 | 23 |
| ST5_2_t1 | 24 | 15 | 5 | 4 | 1 | 25 | 25 | 24 | 24 | 23 | 23 |
| ST5_2_t6 | 24 | 16 | 5 | 4 | 1 | 33 | 31 | 30 | 30 | 23 | 23 |
| ST5_2_n17 | 24 | 30 | 5 | 4 | 1 | 213 | 177 | 144 | 131 | 23 | 23 |
| ST5_2_n10 | 24 | 21 | 5 | 18 | 1 | 90 | 81 | 77 | 74 | 63 | 58 |
| ST5_2_t19 | 23 | 4 | 5 | 7 | 1 | 163 | 21 | 21 | 21 | 20 | 20 |
| ST5_2_n7 | 23 | 4 | 5 | 7 | 1 | 178 | 21 | 21 | 21 | 20 | 20 |
| ST5_2_t9 | 23 | 4 | 5 | 7 | 1 | 21 | 21 | 21 | 21 | 20 | 20 |
| ST5_2_t24 | 23 | 20 | 5 | 7 | 1 | 86 | 77 | 73 | 71 | 20 | 20 |
| ST5_2_n9 | 23 | 35 | 5 | 7 | 1 | 256 | 207 | 164 | 21 | 20 | 20 |
| ST5_2_t17 | 21 | 19 | 6 | 5 | 1 | 72 | 68 | 64 | 63 | 55 | 51 |
| ST5_2_t13 | 26 | 37 | 6 | 5 | 1 | 259 | 210 | 167 | 143 | 114 | 98 |
| ST5_2_n2 | 210 | 25 | 6 | 5 | 1 | 104 | 93 | 88 | 84 | 70 | 63 |
| ST5_2_t16 | 22 | 27 | 6 | 5 | 1 | 121 | 106 | 96 | 92 | 76 | 69 |
| ST5_2_n6 | 29 | 26 | 6 | 5 | 1 | 110 | 99 | 91 | 87 | 70 | 63 |
