## Supplementary Data 2 for "THRESHER: A tool for evidence-based, phylogeny-corrected SNP thresholds for whole genome molecular epidemiology": ST5_40_iter_2.pdf

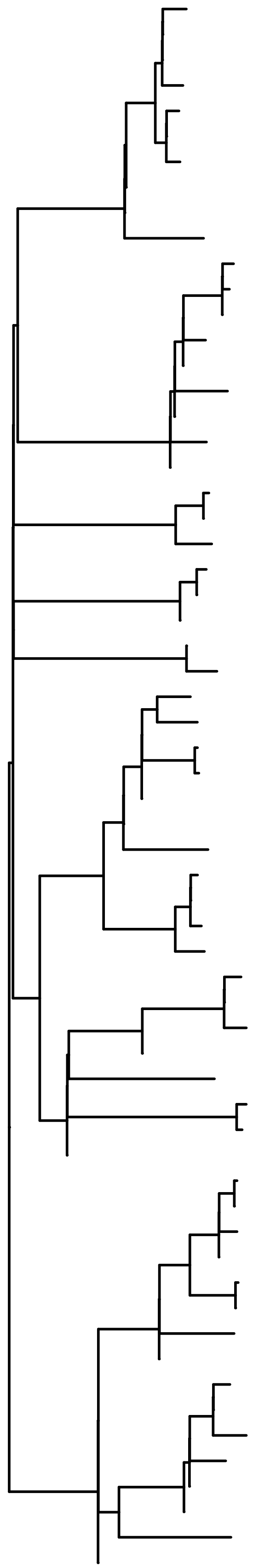

| Sample | Simulation | Plateau 5 | Plateau 15 | Peak | Discrepancy | Cutoff 0 | Cutoff 10 | Cutoff 20 | Cutoff 30 | Cutoff 40 | Cutoff 50 |
|---|---|---|---|---|---|---|---|---|---|---|---|
| ST5_1_t5 | 12 | 2 | 2 | 6 | 4 | 256 | 204 | 102 | 57 | 54 | 10 |
| ST5_1_n8 | 12 | 2 | 2 | 6 | 4 | 131 | 115 | 102 | 57 | 54 | 10 |
| ST5_1_n6 | 12 | 2 | 2 | 6 | 4 | 131 | 115 | 102 | 57 | 54 | 10 |
| ST5_1_n7 | 12 | 2 | 2 | 6 | 4 | 233 | 115 | 102 | 57 | 54 | 10 |
| ST5_1_t17 | 12 | 2 | 2 | 2 | 4 | 213 | 170 | 102 | 57 | 54 | 10 |
| ST5_1_n5 | 12 | 2 | 2 | 2 | 4 | 203 | 170 | 102 | 57 | 54 | 10 |
| ST5_1_t19 | 12 | 2 | 2 | 2 | 4 | 220 | 170 | 102 | 57 | 54 | 10 |
| ST5_1_n10 | 12 | 2 | 2 | 2 | 4 | 176 | 61 | 57 | 57 | 54 | 10 |
| ST5_1_n11 | 12 | 2 | 2 | 2 | 4 | 67 | 61 | 57 | 57 | 54 | 10 |
| ST5_1_t13 | 12 | 16 | 2 | 16 | 4 | 10 | 10 | 10 | 10 | 10 | 10 |
| ST5_1_t8 | 14 | 4 | 3 | 4 | 4 | 221 | 152 | 129 | 19 | 19 | 19 |
| ST5_1_t4 | 14 | 4 | 3 | 4 | 4 | 210 | 152 | 129 | 19 | 19 | 19 |
| ST5_1_n15 | 14 | 4 | 3 | 4 | 4 | 180 | 152 | 129 | 19 | 19 | 19 |
| ST5_1_t9 | 14 | 4 | 3 | 4 | 4 | 215 | 177 | 108 | 19 | 19 | 19 |
| ST5_1_n16 | 14 | 4 | 3 | 4 | 4 | 246 | 122 | 108 | 19 | 19 | 19 |
| ST5_1_t25 | 14 | 4 | 3 | 4 | 4 | 181 | 153 | 130 | 119 | 19 | 19 |
| ST5_1_n17 | 14 | 4 | 3 | 4 | 4 | 142 | 122 | 108 | 19 | 19 | 19 |
| ST5_1_n19 | 14 | 4 | 3 | 4 | 4 | 20 | 19 | 19 | 19 | 19 | 19 |
| ST5_1_n20 | 14 | 4 | 3 | 4 | 4 | 139 | 122 | 108 | 19 | 19 | 19 |
| ST5_1_t20 | 11 | 1 | 1 | 1 | 4 | 23 | 22 | 22 | 22 | 22 | 22 |
| ST5_1_n1 | 11 | 1 | 1 | 1 | 4 | 197 | 22 | 22 | 22 | 22 | 22 |
| ST5_1_t23 | 11 | 34 | 1 | 43 | 4 | 260 | 207 | 168 | 145 | 120 | 22 |
| ST5_2_t18 | 25 | 8 | 6 | 8 | 4 | 251 | 197 | 139 | 125 | 105 | 84 |
| ST5_2_n21 | 25 | 8 | 6 | 8 | 4 | 244 | 197 | 139 | 125 | 105 | 84 |
| ST5_2_n22 | 25 | 8 | 6 | 36 | 4 | 198 | 165 | 139 | 125 | 105 | 84 |
| ST5_3_n3 | 31 | 28 | 9 | 34 | 4 | 190 | 159 | 134 | 121 | 102 | 82 |
| ST5_3_n1 | 31 | 32 | 9 | 40 | 4 | 248 | 200 | 164 | 121 | 102 | 82 |
| ST5_2_t8 | 24 | 7 | 5 | 32 | 4 | 151 | 129 | 111 | 104 | 33 | 33 |
| ST5_2_t6 | 24 | 7 | 5 | 20 | 4 | 71 | 65 | 60 | 60 | 33 | 33 |
| ST5_2_t4 | 24 | 7 | 5 | 11 | 4 | 224 | 34 | 33 | 33 | 33 | 33 |
| ST5_2_t22 | 24 | 7 | 5 | 11 | 4 | 35 | 34 | 33 | 33 | 33 | 33 |
| ST5_2_n17 | 24 | 7 | 5 | 39 | 4 | 247 | 199 | 78 | 77 | 33 | 33 |
| ST5_2_n18 | 24 | 7 | 5 | 24 | 4 | 95 | 86 | 78 | 77 | 33 | 33 |
| ST5_2_n10 | 24 | 23 | 5 | 27 | 4 | 119 | 107 | 95 | 92 | 78 | 33 |
| ST5_2_t19 | 23 | 9 | 7 | 9 | 4 | 102 | 91 | 82 | 81 | 69 | 61 |
| ST5_2_n7 | 23 | 9 | 7 | 9 | 4 | 205 | 91 | 82 | 81 | 69 | 61 |
| ST5_2_t9 | 23 | 9 | 7 | 9 | 4 | 155 | 91 | 82 | 81 | 69 | 61 |
| ST5_2_t24 | 23 | 29 | 7 | 35 | 4 | 192 | 160 | 135 | 122 | 69 | 61 |
| ST5_2_t17 | 21 | 6 | 4 | 7 | 4 | 96 | 87 | 18 | 18 | 18 | 18 |
| ST5_2_n1 | 21 | 6 | 4 | 7 | 4 | 19 | 18 | 18 | 18 | 18 | 18 |
| ST5_2_t5 | 21 | 6 | 4 | 7 | 4 | 150 | 18 | 18 | 18 | 18 | 18 |
| ST5_2_n2 | 210 | 31 | 4 | 38 | 4 | 230 | 186 | 155 | 138 | 51 | 18 |
| ST5_2_t15 | 27 | 25 | 4 | 30 | 4 | 136 | 120 | 106 | 100 | 86 | 71 |
| ST5_2_t16 | 22 | 10 | 4 | 10 | 4 | 44 | 41 | 39 | 39 | 39 | 38 |
| ST5_2_t14 | 22 | 10 | 4 | 10 | 4 | 83 | 41 | 39 | 39 | 39 | 38 |
| ST5_2_n5 | 211 | 18 | 4 | 19 | 4 | 62 | 58 | 54 | 54 | 51 | 18 |
| ST5_3_t17 | 33 | 12 | 10 | 14 | 4 | 70 | 64 | 6 | 6 | 6 | 6 |
| ST5_3_t15 | 33 | 12 | 10 | 14 | 4 | 257 | 64 | 6 | 6 | 6 | 6 |
| ST5_3_t18 | 33 | 12 | 10 | 31 | 4 | 141 | 124 | 6 | 6 | 6 | 6 |
| ST5_3_n10 | 33 | 12 | 10 | 15 | 4 | 6 | 6 | 6 | 6 | 6 | 6 |
| ST5_3_t9 | 33 | 15 | 10 | 13 | 4 | 209 | 93 | 84 | 83 | 71 | 63 |
| ST5_3_n6 | 33 | 15 | 10 | 13 | 4 | 105 | 93 | 84 | 83 | 71 | 63 |
| ST5_3_t24 | 33 | 24 | 10 | 28 | 4 | 122 | 110 | 97 | 94 | 80 | 68 |
| ST5_3_n13 | 33 | 33 | 10 | 41 | 4 | 255 | 203 | 166 | 75 | 65 | 6 |
| ST5_3_t3 | 34 | 14 | 10 | 12 | 4 | 132 | 116 | 25 | 25 | 25 | 25 |
| ST5_3_n17 | 34 | 14 | 10 | 12 | 4 | 26 | 25 | 25 | 25 | 25 | 25 |
| ST5_3_t5 | 34 | 14 | 10 | 12 | 4 | 76 | 70 | 25 | 25 | 25 | 25 |
| ST5_3_t23 | 34 | 14 | 10 | 12 | 4 | 109 | 97 | 87 | 25 | 25 | 25 |
| ST5_3_n18 | 34 | 14 | 10 | 12 | 4 | 124 | 36 | 25 | 25 | 25 | 25 |
| ST5_3_n19 | 34 | 14 | 10 | 12 | 4 | 37 | 36 | 25 | 25 | 25 | 25 |
| ST5_3_t21 | 36 | 20 | 10 | 22 | 4 | 84 | 76 | 69 | 69 | 62 | 56 |
| ST5_3_n22 | 39 | 21 | 10 | 23 | 4 | 93 | 84 | 76 | 75 | 65 | 6 |

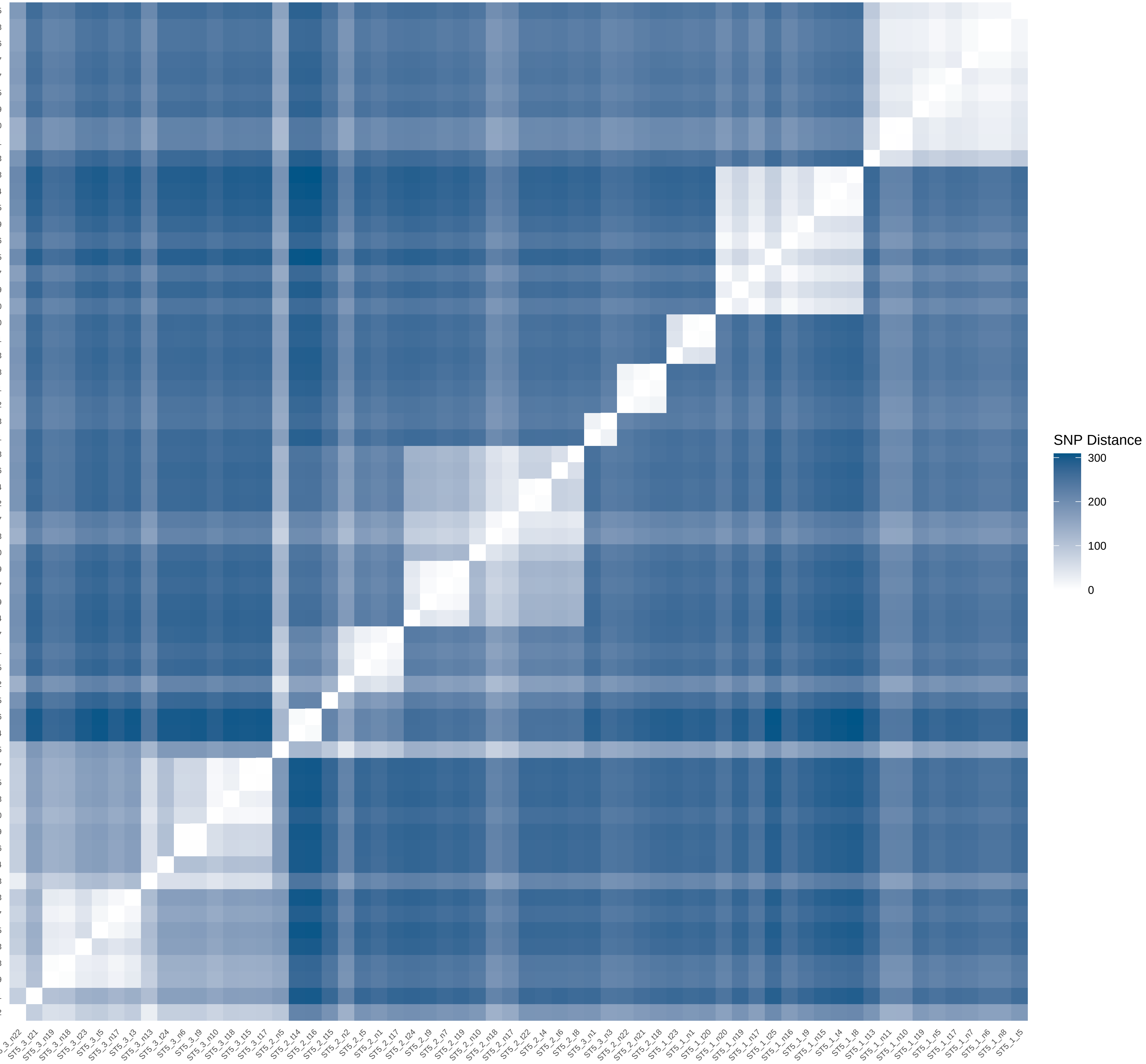
