## Supplementary Data 2 for "THRESHER: A tool for evidence-based, phylogeny-corrected SNP thresholds for whole genome molecular epidemiology": ST8_20_iter_2.pdf

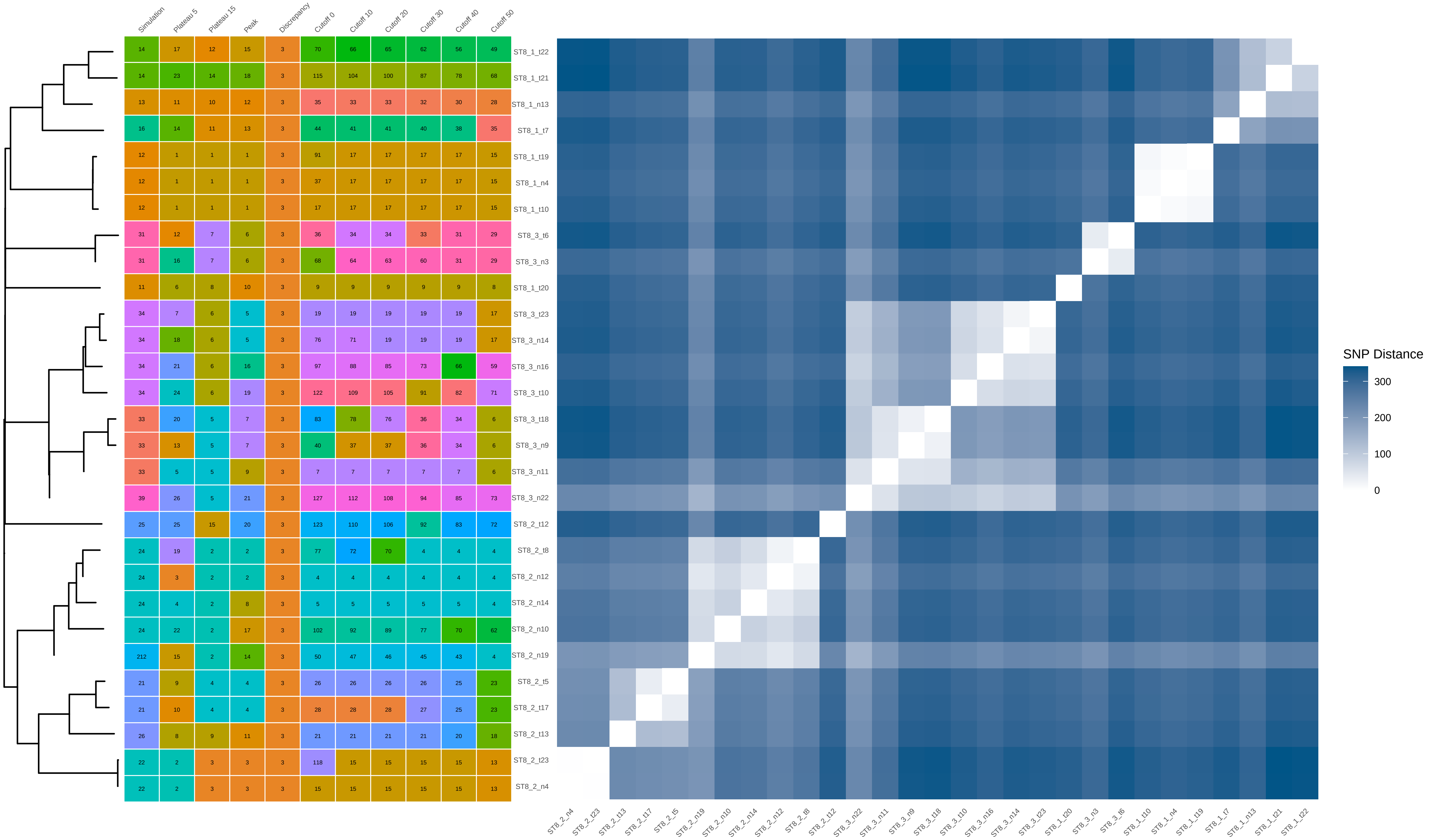


| | Simulation | Plateau 5 | Plateau 15 | Peak | Discrepancy | Cutoff 0 | Cutoff 10 | Cutoff 20 | Cutoff 30 | Cutoff 40 | Cutoff 50 |
|---|---|---|---|---|---|---|---|---|---|---|---|
| ST8_1_t22 | 14 | 17 | 12 | 15 | 3 | 70 | 66 | 65 | 62 | 56 | 49 |
| ST8_1_t21 | 14 | 23 | 14 | 18 | 3 | 115 | 104 | 100 | 87 | 78 | 68 |
| ST8_1_n13 | 13 | 11 | 10 | 12 | 3 | 35 | 33 | 33 | 32 | 30 | 28 |
| ST8_1_t7 | 16 | 14 | 11 | 13 | 3 | 44 | 41 | 41 | 40 | 38 | 35 |
| ST8_1_t19 | 12 | 1 | 1 | 1 | 3 | 91 | 17 | 17 | 17 | 17 | 15 |
| ST8_1_n4 | 12 | 1 | 1 | 1 | 3 | 37 | 17 | 17 | 17 | 17 | 15 |
| ST8_1_t10 | 12 | 1 | 1 | 1 | 3 | 17 | 17 | 17 | 17 | 17 | 15 |
| ST8_3_t6 | 31 | 12 | 7 | 6 | 3 | 36 | 34 | 34 | 33 | 31 | 29 |
| ST8_3_n3 | 31 | 16 | 7 | 6 | 3 | 68 | 64 | 63 | 60 | 31 | 29 |
| ST8_1_t20 | 11 | 6 | 8 | 10 | 3 | 9 | 9 | 9 | 9 | 9 | 8 |
| ST8_3_t23 | 34 | 7 | 6 | 5 | 3 | 19 | 19 | 19 | 19 | 19 | 17 |
| ST8_3_n14 | 34 | 18 | 6 | 5 | 3 | 76 | 71 | 19 | 19 | 19 | 17 |
| ST8_3_n16 | 34 | 21 | 6 | 16 | 3 | 97 | 88 | 85 | 73 | 66 | 59 |
| ST8_3_t10 | 34 | 24 | 6 | 19 | 3 | 122 | 109 | 105 | 91 | 82 | 71 |
| ST8_3_t18 | 33 | 20 | 5 | 7 | 3 | 83 | 78 | 76 | 36 | 34 | 6 |
| ST8_3_n9 | 33 | 13 | 5 | 7 | 3 | 40 | 37 | 37 | 36 | 34 | 6 |
| ST8_3_n11 | 33 | 5 | 5 | 9 | 3 | 7 | 7 | 7 | 7 | 7 | 6 |
| ST8_3_n22 | 39 | 26 | 5 | 21 | 3 | 127 | 112 | 108 | 94 | 85 | 73 |
| ST8_2_t12 | 25 | 25 | 15 | 20 | 3 | 123 | 110 | 106 | 92 | 83 | 72 |
| ST8_2_t8 | 24 | 19 | 2 | 2 | 3 | 77 | 72 | 70 | 4 | 4 | 4 |
| ST8_2_n12 | 24 | 3 | 2 | 2 | 3 | 4 | 4 | 4 | 4 | 4 | 4 |
| ST8_2_n14 | 24 | 4 | 2 | 8 | 3 | 5 | 5 | 5 | 5 | 5 | 4 |
| ST8_2_n10 | 24 | 22 | 2 | 17 | 3 | 102 | 92 | 89 | 77 | 70 | 62 |
| ST8_2_n19 | 212 | 15 | 2 | 14 | 3 | 50 | 47 | 46 | 45 | 43 | 4 |
| ST8_2_t5 | 21 | 9 | 4 | 4 | 3 | 26 | 26 | 26 | 26 | 25 | 23 |
| ST8_2_t17 | 21 | 10 | 4 | 4 | 3 | 28 | 28 | 28 | 27 | 25 | 23 |
| ST8_2_t13 | 26 | 8 | 9 | 11 | 3 | 21 | 21 | 21 | 21 | 20 | 18 |
| ST8_2_t23 | 22 | 2 | 3 | 3 | 3 | 118 | 15 | 15 | 15 | 15 | 13 |
| ST8_2_n4 | 22 | 2 | 3 | 3 | 3 | 15 | 15 | 15 | 15 | 15 | 13 |
