## Supplementary Data 2 for "THRESHER: A tool for evidence-based, phylogeny-corrected SNP thresholds for whole genome molecular epidemiology": ST8_20_iter_4.pdf

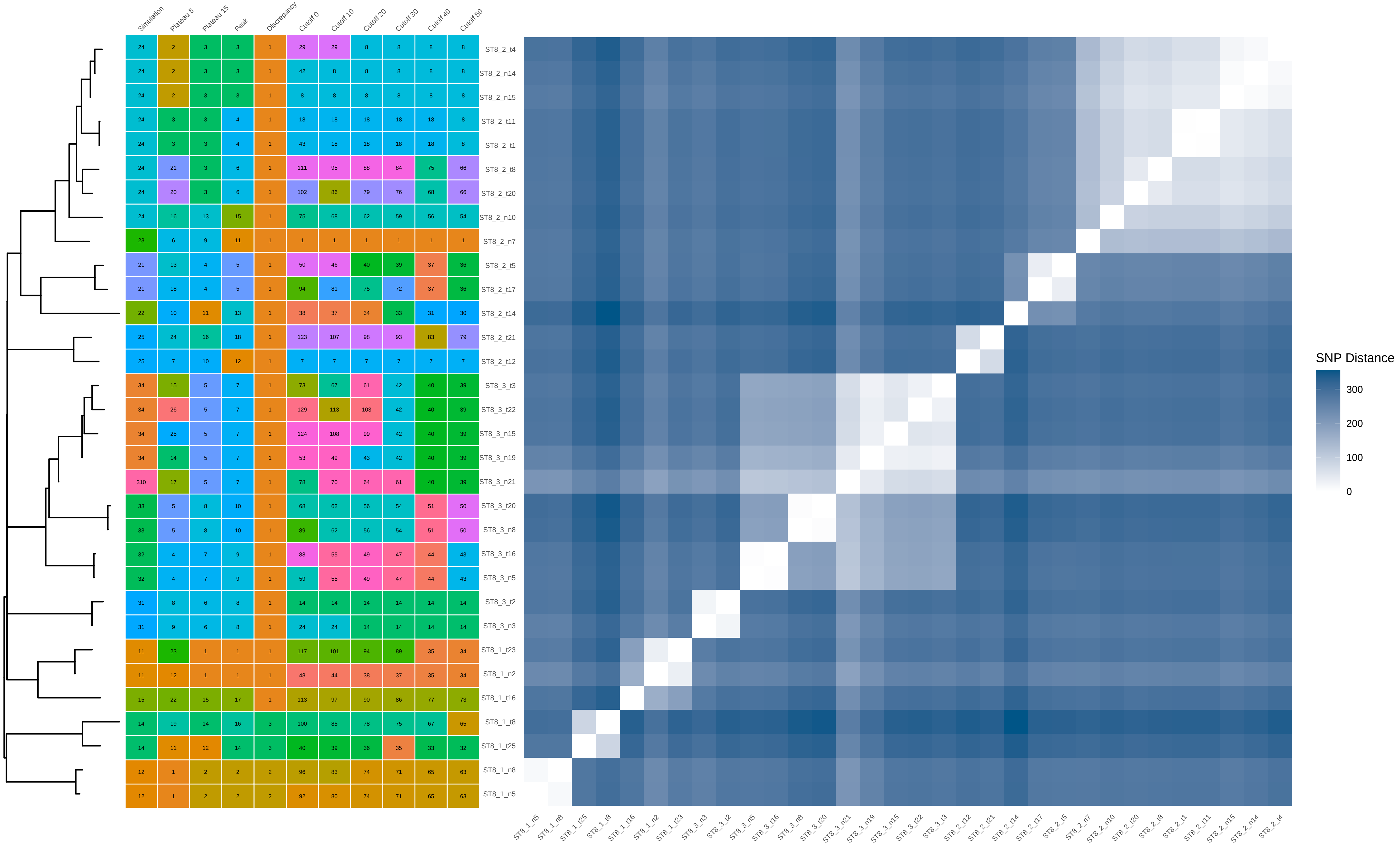


| | Simulation | Plateau 5 | Plateau 15 | Peak | Discrepancy | Cutoff 0 | Cutoff 10 | Cutoff 20 | Cutoff 30 | Cutoff 40 | Cutoff 50 |
|---|---|---|---|---|---|---|---|---|---|---|---|
| ST8_2_t4 | 24 | 2 | 3 | 3 | 1 | 29 | 29 | 8 | 8 | 8 | 8 |
| ST8_2_n14 | 24 | 2 | 3 | 3 | 1 | 42 | 8 | 8 | 8 | 8 | 8 |
| ST8_2_n15 | 24 | 2 | 3 | 3 | 1 | 8 | 8 | 8 | 8 | 8 | 8 |
| ST8_2_t11 | 24 | 3 | 3 | 4 | 1 | 18 | 18 | 18 | 18 | 18 | 8 |
| ST8_2_t1 | 24 | 3 | 3 | 4 | 1 | 43 | 18 | 18 | 18 | 18 | 8 |
| ST8_2_t8 | 24 | 21 | 3 | 6 | 1 | 111 | 95 | 88 | 84 | 75 | 66 |
| ST8_2_t20 | 24 | 20 | 3 | 6 | 1 | 102 | 86 | 79 | 76 | 68 | 66 |
| ST8_2_n10 | 24 | 16 | 13 | 15 | 1 | 75 | 68 | 62 | 59 | 56 | 54 |
| ST8_2_n7 | 23 | 6 | 9 | 11 | 1 | 1 | 1 | 1 | 1 | 1 | 1 |
| ST8_2_t5 | 21 | 13 | 4 | 5 | 1 | 50 | 46 | 40 | 39 | 37 | 36 |
| ST8_2_t17 | 21 | 18 | 4 | 5 | 1 | 94 | 81 | 75 | 72 | 37 | 36 |
| ST8_2_t14 | 22 | 10 | 11 | 13 | 1 | 38 | 37 | 34 | 33 | 31 | 30 |
| ST8_2_t21 | 25 | 24 | 16 | 18 | 1 | 123 | 107 | 98 | 93 | 83 | 79 |
| ST8_2_t12 | 25 | 7 | 10 | 12 | 1 | 7 | 7 | 7 | 7 | 7 | 7 |
| ST8_3_t3 | 34 | 15 | 5 | 7 | 1 | 73 | 67 | 61 | 42 | 40 | 39 |
| ST8_3_t22 | 34 | 26 | 5 | 7 | 1 | 129 | 113 | 103 | 42 | 40 | 39 |
| ST8_3_n15 | 34 | 25 | 5 | 7 | 1 | 124 | 108 | 99 | 42 | 40 | 39 |
| ST8_3_n19 | 34 | 14 | 5 | 7 | 1 | 53 | 49 | 43 | 42 | 40 | 39 |
| ST8_3_n21 | 310 | 17 | 5 | 7 | 1 | 78 | 70 | 64 | 61 | 40 | 39 |
| ST8_3_t20 | 33 | 5 | 8 | 10 | 1 | 68 | 62 | 56 | 54 | 51 | 50 |
| ST8_3_n8 | 33 | 5 | 8 | 10 | 1 | 89 | 62 | 56 | 54 | 51 | 50 |
| ST8_3_t16 | 32 | 4 | 7 | 9 | 1 | 88 | 55 | 49 | 47 | 44 | 43 |
| ST8_3_n5 | 32 | 4 | 7 | 9 | 1 | 59 | 55 | 49 | 47 | 44 | 43 |
| ST8_3_t2 | 31 | 8 | 6 | 8 | 1 | 14 | 14 | 14 | 14 | 14 | 14 |
| ST8_3_n3 | 31 | 9 | 6 | 8 | 1 | 24 | 24 | 14 | 14 | 14 | 14 |
| ST8_1_t23 | 11 | 23 | 1 | 1 | 1 | 117 | 101 | 94 | 89 | 35 | 34 |
| ST8_1_n2 | 11 | 12 | 1 | 1 | 1 | 48 | 44 | 38 | 37 | 35 | 34 |
| ST8_1_t16 | 15 | 22 | 15 | 17 | 1 | 113 | 97 | 90 | 86 | 77 | 73 |
| ST8_1_t8 | 14 | 19 | 14 | 16 | 3 | 100 | 85 | 78 | 75 | 67 | 65 |
| ST8_1_t25 | 14 | 11 | 12 | 14 | 3 | 40 | 39 | 36 | 35 | 33 | 32 |
| ST8_1_n8 | 12 | 1 | 2 | 2 | 2 | 96 | 83 | 74 | 71 | 65 | 63 |
| ST8_1_n5 | 12 | 1 | 2 | 2 | 2 | 92 | 80 | 74 | 71 | 65 | 63 |
