## Supplementary Data 2 for "THRESHER: A tool for evidence-based, phylogeny-corrected SNP thresholds for whole genome molecular epidemiology": ST8_20_iter_5.pdf

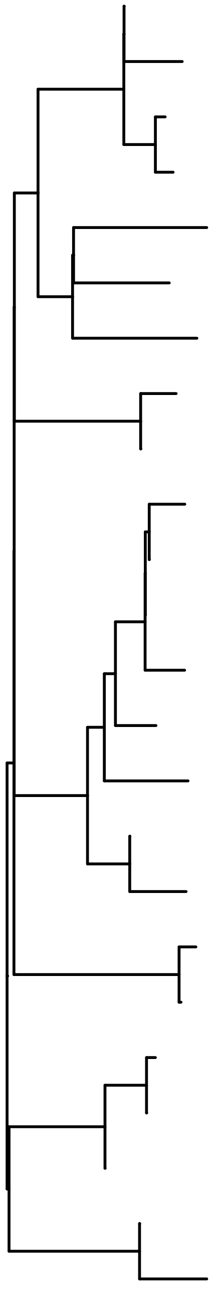

| | Simulation | Plateau 5 | Plateau 15 | Peak | Discrepancy | Cutoff 0 | Cutoff 10 | Cutoff 20 | Cutoff 30 | Cutoff 40 | Cutoff 50 |
|---|---|---|---|---|---|---|---|---|---|---|---|
| ST8_2_n18 | 24 | 21 | 3 | 3 | 1 | 119 | 112 | 102 | 93 | 81 | 1 |
| ST8_2_n10 | 24 | 11 | 3 | 3 | 1 | 77 | 75 | 70 | 65 | 59 | 1 |
| ST8_2_t20 | 24 | 7 | 3 | 3 | 1 | 31 | 31 | 30 | 1 | 1 | 1 |
| ST8_2_t6 | 24 | 3 | 3 | 3 | 1 | 1 | 1 | 1 | 1 | 1 | 1 |
| ST8_2_t23 | 22 | 17 | 11 | 11 | 1 | 103 | 97 | 89 | 83 | 71 | 62 |
| ST8_2_n1 | 21 | 13 | 10 | 10 | 1 | 86 | 84 | 77 | 71 | 63 | 55 |
| ST8_2_t15 | 27 | 8 | 9 | 9 | 1 | 37 | 37 | 35 | 31 | 31 | 27 |
| ST8_2_t12 | 25 | 20 | 4 | 4 | 1 | 116 | 109 | 99 | 80 | 70 | 61 |
| ST8_2_n22 | 25 | 16 | 4 | 4 | 1 | 99 | 94 | 86 | 80 | 70 | 61 |
| ST8_3_t22 | 34 | 2 | 7 | 7 | 1 | 109 | 103 | 94 | 50 | 48 | 43 |
| ST8_3_n18 | 34 | 2 | 7 | 7 | 1 | 58 | 58 | 55 | 50 | 48 | 43 |
| ST8_3_n19 | 34 | 2 | 7 | 7 | 1 | 68 | 58 | 55 | 50 | 48 | 43 |
| ST8_3_t10 | 34 | 10 | 7 | 7 | 1 | 76 | 74 | 69 | 64 | 48 | 43 |
| ST8_3_t7 | 34 | 4 | 8 | 8 | 1 | 17 | 17 | 17 | 16 | 16 | 16 |
| ST8_3_t21 | 36 | 18 | 12 | 12 | 1 | 104 | 98 | 90 | 84 | 72 | 63 |
| ST8_3_n13 | 33 | 14 | 6 | 6 | 1 | 96 | 92 | 84 | 78 | 68 | 59 |
| ST8_3_n10 | 33 | 6 | 6 | 6 | 1 | 19 | 19 | 19 | 18 | 18 | 18 |
| ST8_3_t1 | 31 | 5 | 5 | 5 | 1 | 18 | 18 | 18 | 17 | 17 | 17 |
| ST8_3_n1 | 31 | 12 | 5 | 5 | 1 | 82 | 80 | 18 | 17 | 17 | 17 |
| ST8_1_t10 | 12 | 1 | 1 | 1 | 1 | 92 | 13 | 13 | 13 | 13 | 13 |
| ST8_1_n4 | 12 | 1 | 1 | 1 | 1 | 13 | 13 | 13 | 13 | 13 | 13 |
| ST8_1_n11 | 12 | 22 | 1 | 1 | 1 | 122 | 115 | 104 | 94 | 82 | 13 |
| ST8_1_n20 | 14 | 15 | 2 | 2 | 1 | 98 | 93 | 85 | 79 | 69 | 60 |
| ST8_1_n15 | 14 | 9 | 2 | 2 | 1 | 64 | 63 | 60 | 55 | 51 | 46 |

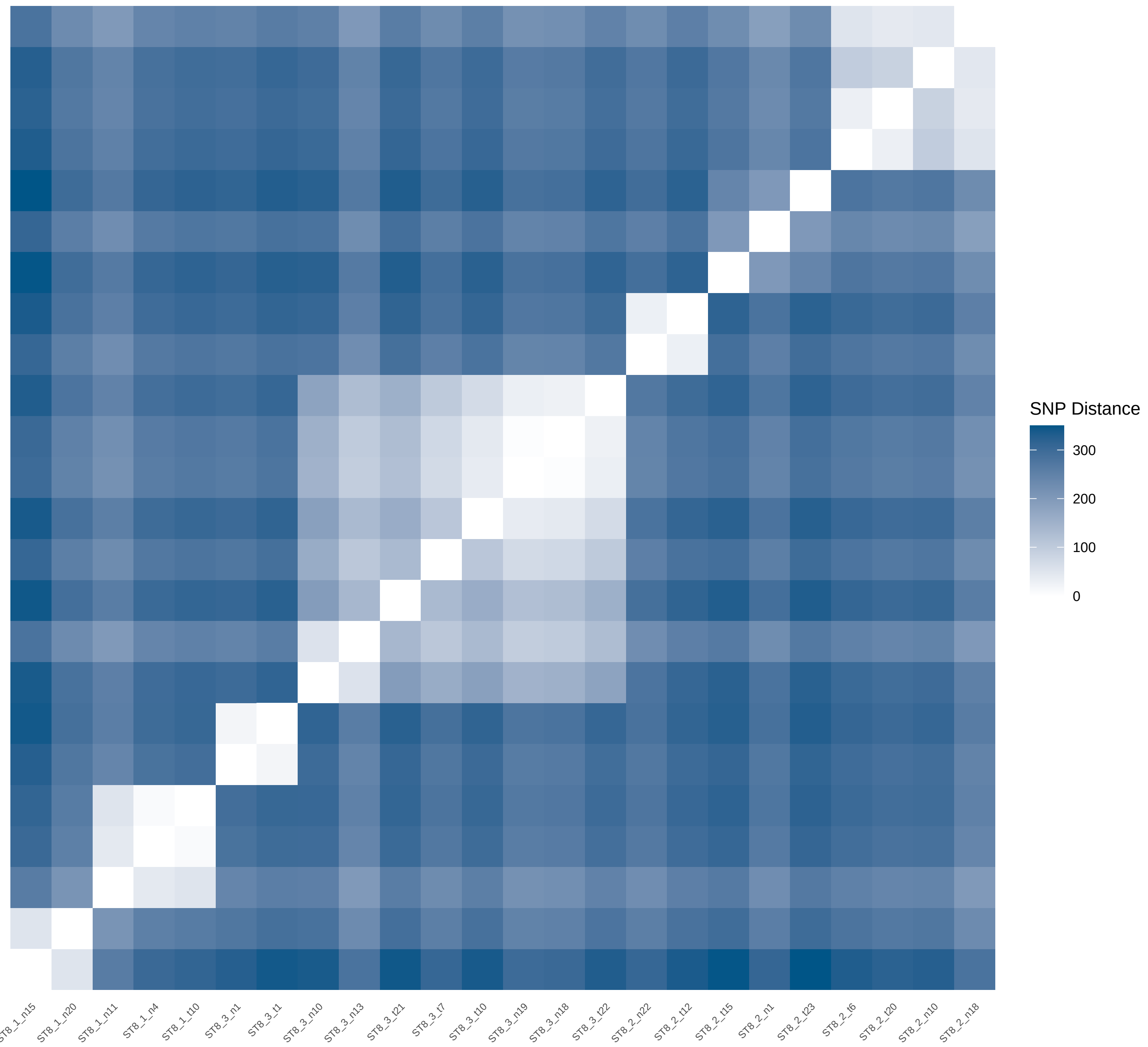
