## Supplementary Data 2 for "THRESHER: A tool for evidence-based, phylogeny-corrected SNP thresholds for whole genome molecular epidemiology": ST8_20_iter_6.pdf

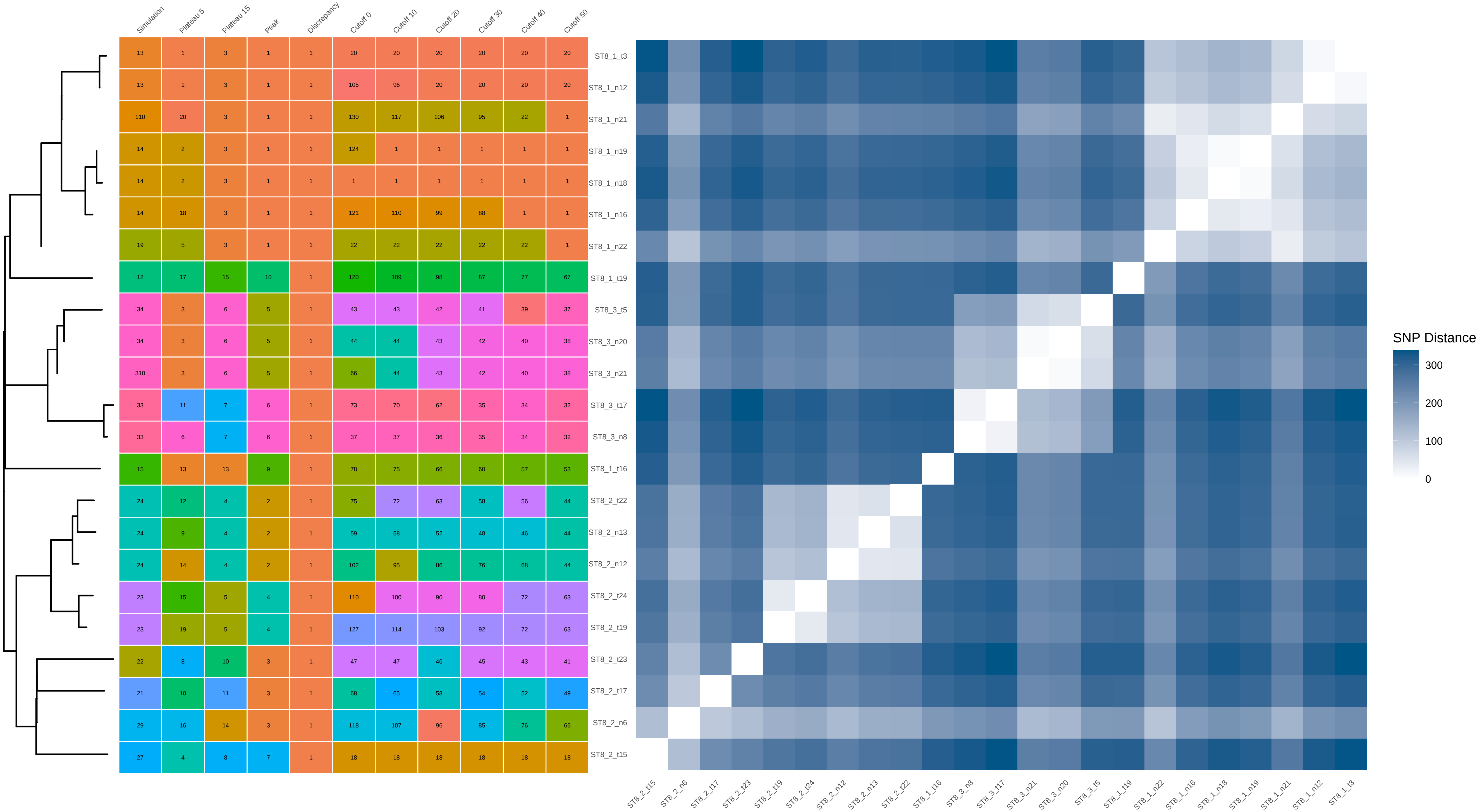


| | Simulation | Plateau 5 | Plateau 15 | Peak | Discrepancy | Cutoff 0 | Cutoff 10 | Cutoff 20 | Cutoff 30 | Cutoff 40 | Cutoff 50 |
|---|---|---|---|---|---|---|---|---|---|---|---|
| ST8_1_t3 | 13 | 1 | 3 | 1 | 1 | 20 | 20 | 20 | 20 | 20 | 20 |
| ST8_1_n12 | 13 | 1 | 3 | 1 | 1 | 105 | 96 | 20 | 20 | 20 | 20 |
| ST8_1_n21 | 110 | 20 | 3 | 1 | 1 | 130 | 117 | 106 | 95 | 22 | 1 |
| ST8_1_n19 | 14 | 2 | 3 | 1 | 1 | 124 | 1 | 1 | 1 | 1 | 1 |
| ST8_1_n18 | 14 | 2 | 3 | 1 | 1 | 1 | 1 | 1 | 1 | 1 | 1 |
| ST8_1_n16 | 14 | 18 | 3 | 1 | 1 | 121 | 110 | 99 | 88 | 1 | 1 |
| ST8_1_n22 | 19 | 5 | 3 | 1 | 1 | 22 | 22 | 22 | 22 | 22 | 1 |
| ST8_1_t19 | 12 | 17 | 15 | 10 | 1 | 120 | 109 | 98 | 87 | 77 | 67 |
| ST8_3_t5 | 34 | 3 | 6 | 5 | 1 | 43 | 43 | 42 | 41 | 39 | 37 |
| ST8_3_n20 | 34 | 3 | 6 | 5 | 1 | 44 | 44 | 43 | 42 | 40 | 38 |
| ST8_3_n21 | 310 | 3 | 6 | 5 | 1 | 66 | 44 | 43 | 42 | 40 | 38 |
| ST8_3_t17 | 33 | 11 | 7 | 6 | 1 | 73 | 70 | 62 | 35 | 34 | 32 |
| ST8_3_n8 | 33 | 6 | 7 | 6 | 1 | 37 | 37 | 36 | 35 | 34 | 32 |
| ST8_1_t16 | 15 | 13 | 13 | 9 | 1 | 78 | 75 | 66 | 60 | 57 | 53 |
| ST8_2_t22 | 24 | 12 | 4 | 2 | 1 | 75 | 72 | 63 | 58 | 56 | 44 |
| ST8_2_n13 | 24 | 9 | 4 | 2 | 1 | 59 | 58 | 52 | 48 | 46 | 44 |
| ST8_2_n12 | 24 | 14 | 4 | 2 | 1 | 102 | 95 | 86 | 76 | 68 | 44 |
| ST8_2_t24 | 23 | 15 | 5 | 4 | 1 | 110 | 100 | 90 | 80 | 72 | 63 |
| ST8_2_t19 | 23 | 19 | 5 | 4 | 1 | 127 | 114 | 103 | 92 | 72 | 63 |
| ST8_2_t23 | 22 | 8 | 10 | 3 | 1 | 47 | 47 | 46 | 45 | 43 | 41 |
| ST8_2_t17 | 21 | 10 | 11 | 3 | 1 | 68 | 65 | 58 | 54 | 52 | 49 |
| ST8_2_n6 | 29 | 16 | 14 | 3 | 1 | 118 | 107 | 96 | 85 | 76 | 66 |
| ST8_2_t15 | 27 | 4 | 8 | 7 | 1 | 18 | 18 | 18 | 18 | 18 | 18 |
