## Supplementary Data 2 for "THRESHER: A tool for evidence-based, phylogeny-corrected SNP thresholds for whole genome molecular epidemiology": ST8_20_iter_8.pdf

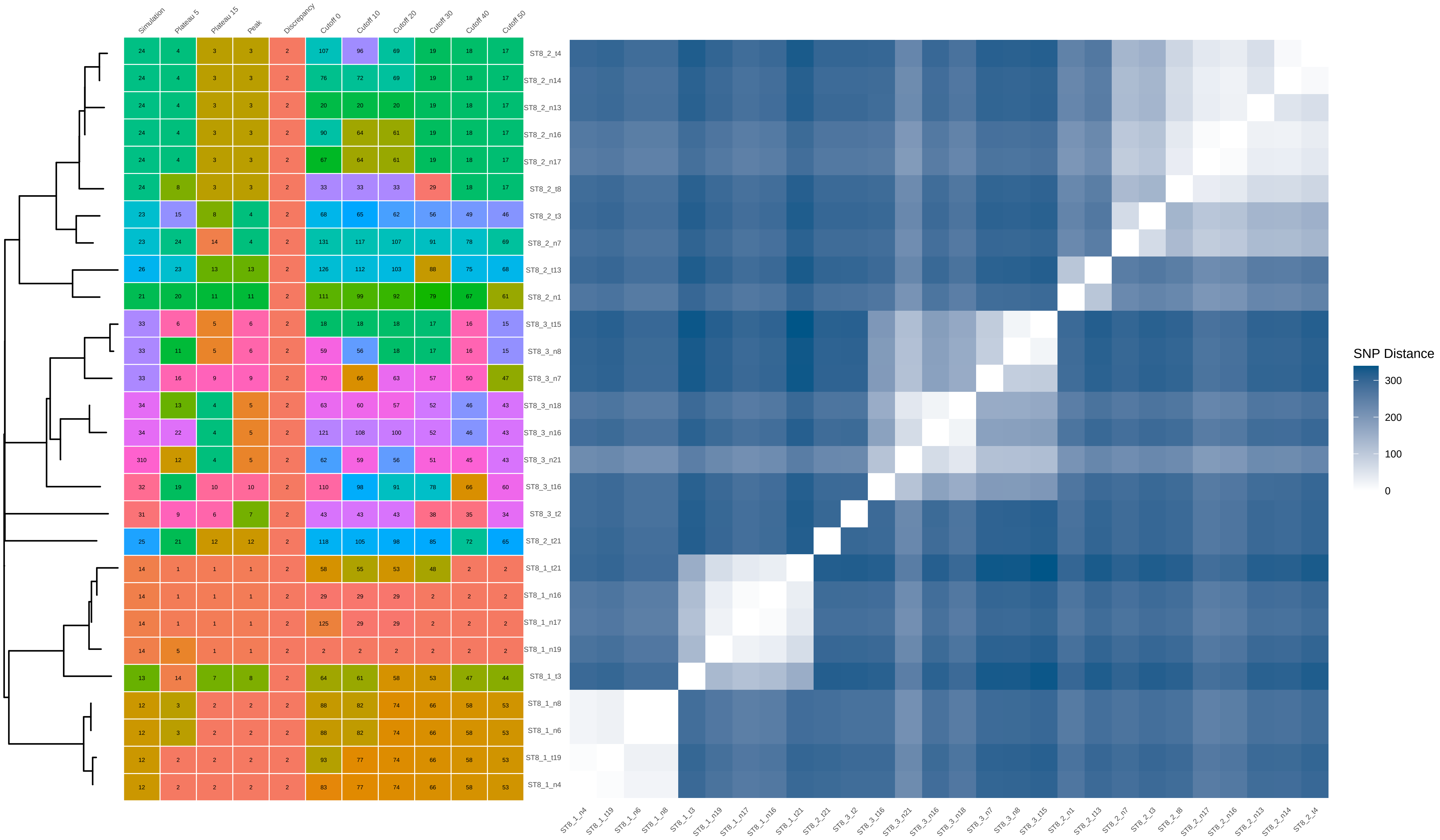


| | Simulation | Plateau 5 | Plateau 15 | Peak | Discrepancy | Cutoff 0 | Cutoff 10 | Cutoff 20 | Cutoff 30 | Cutoff 40 | Cutoff 50 |
|---|---|---|---|---|---|---|---|---|---|---|---|
| ST8_2_t4 | 24 | 4 | 3 | 3 | 2 | 107 | 96 | 69 | 19 | 18 | 17 |
| ST8_2_n14 | 24 | 4 | 3 | 3 | 2 | 76 | 72 | 69 | 19 | 18 | 17 |
| ST8_2_n13 | 24 | 4 | 3 | 3 | 2 | 20 | 20 | 20 | 19 | 18 | 17 |
| ST8_2_n16 | 24 | 4 | 3 | 3 | 2 | 90 | 64 | 61 | 19 | 18 | 17 |
| ST8_2_n17 | 24 | 4 | 3 | 3 | 2 | 67 | 64 | 61 | 19 | 18 | 17 |
| ST8_2_t8 | 24 | 8 | 3 | 3 | 2 | 33 | 33 | 33 | 29 | 18 | 17 |
| ST8_2_t3 | 23 | 15 | 8 | 4 | 2 | 68 | 65 | 62 | 56 | 49 | 46 |
| ST8_2_n7 | 23 | 24 | 14 | 4 | 2 | 131 | 117 | 107 | 91 | 78 | 69 |
| ST8_2_t13 | 26 | 23 | 13 | 13 | 2 | 126 | 112 | 103 | 88 | 75 | 68 |
| ST8_2_n1 | 21 | 20 | 11 | 11 | 2 | 111 | 99 | 92 | 79 | 67 | 61 |
| ST8_3_t15 | 33 | 6 | 5 | 6 | 2 | 18 | 18 | 18 | 17 | 16 | 15 |
| ST8_3_n8 | 33 | 11 | 5 | 6 | 2 | 59 | 56 | 18 | 17 | 16 | 15 |
| ST8_3_n7 | 33 | 16 | 9 | 9 | 2 | 70 | 66 | 63 | 57 | 50 | 47 |
| ST8_3_n18 | 34 | 13 | 4 | 5 | 2 | 63 | 60 | 57 | 52 | 46 | 43 |
| ST8_3_n16 | 34 | 22 | 4 | 5 | 2 | 121 | 108 | 100 | 52 | 46 | 43 |
| ST8_3_n21 | 310 | 12 | 4 | 5 | 2 | 62 | 59 | 56 | 51 | 45 | 43 |
| ST8_3_t16 | 32 | 19 | 10 | 10 | 2 | 110 | 98 | 91 | 78 | 66 | 60 |
| ST8_3_t2 | 31 | 9 | 6 | 7 | 2 | 43 | 43 | 43 | 38 | 35 | 34 |
| ST8_2_t21 | 25 | 21 | 12 | 12 | 2 | 118 | 105 | 98 | 85 | 72 | 65 |
| ST8_1_t21 | 14 | 1 | 1 | 1 | 2 | 58 | 55 | 53 | 48 | 2 | 2 |
| ST8_1_n16 | 14 | 1 | 1 | 1 | 2 | 29 | 29 | 29 | 2 | 2 | 2 |
| ST8_1_n17 | 14 | 1 | 1 | 1 | 2 | 125 | 29 | 29 | 2 | 2 | 2 |
| ST8_1_n19 | 14 | 5 | 1 | 1 | 2 | 2 | 2 | 2 | 2 | 2 | 2 |
| ST8_1_t3 | 13 | 14 | 7 | 8 | 2 | 64 | 61 | 58 | 53 | 47 | 44 |
| ST8_1_n8 | 12 | 3 | 2 | 2 | 2 | 88 | 82 | 74 | 66 | 58 | 53 |
| ST8_1_n6 | 12 | 3 | 2 | 2 | 2 | 88 | 82 | 74 | 66 | 58 | 53 |
| ST8_1_t19 | 12 | 2 | 2 | 2 | 2 | 93 | 77 | 74 | 66 | 58 | 53 |
| ST8_1_n4 | 12 | 2 | 2 | 2 | 2 | 83 | 77 | 74 | 66 | 58 | 53 |
