## Supplementary Data 2 for "THRESHER: A tool for evidence-based, phylogeny-corrected SNP thresholds for whole genome molecular epidemiology": ST8_20_iter_9.pdf

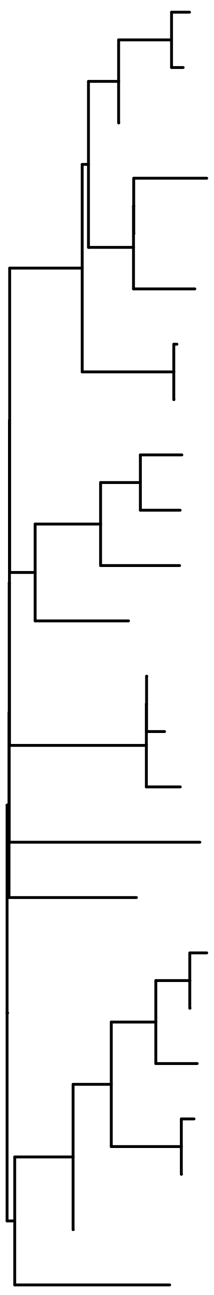

| | Simulation | Plateau 5 | Plateau 15 | Peak | Discrepancy | Cutoff 0 | Cutoff 10 | Cutoff 20 | Cutoff 30 | Cutoff 40 | Cutoff 50 |
|---|---|---|---|---|---|---|---|---|---|---|---|
| ST8_3_t5 | 34 | 9 | 5 | 6 | 1 | 26 | 25 | 25 | 7 | 7 | 7 |
| ST8_3_t3 | 34 | 4 | 5 | 6 | 1 | 7 | 7 | 7 | 7 | 7 | 7 |
| ST8_3_n20 | 34 | 8 | 10 | 10 | 1 | 22 | 21 | 21 | 21 | 21 | 21 |
| ST8_3_t14 | 33 | 6 | 8 | 8 | 1 | 16 | 15 | 15 | 15 | 15 | 15 |
| ST8_3_n13 | 33 | 12 | 11 | 4 | 1 | 43 | 40 | 39 | 37 | 35 | 34 |
| ST8_3_n10 | 33 | 3 | 6 | 4 | 1 | 1 | 1 | 1 | 1 | 1 | 1 |
| ST8_3_t16 | 32 | 2 | 4 | 5 | 1 | 27 | 26 | 26 | 25 | 24 | 24 |
| ST8_3_n5 | 32 | 2 | 4 | 5 | 1 | 131 | 26 | 26 | 25 | 24 | 24 |
| ST8_2_t8 | 24 | 18 | 14 | 13 | 1 | 97 | 92 | 84 | 77 | 72 | 69 |
| ST8_2_t22 | 24 | 5 | 7 | 7 | 1 | 8 | 8 | 8 | 8 | 8 | 8 |
| ST8_2_t24 | 23 | 20 | 16 | 15 | 1 | 104 | 99 | 90 | 82 | 77 | 74 |
| ST8_2_n2 | 210 | 22 | 18 | 17 | 1 | 125 | 118 | 106 | 97 | 88 | 83 |
| ST8_2_n22 | 25 | 11 | 3 | 3 | 1 | 42 | 39 | 38 | 36 | 34 | 33 |
| ST8_2_n21 | 25 | 17 | 3 | 3 | 1 | 89 | 84 | 38 | 36 | 34 | 33 |
| ST8_2_t7 | 25 | 15 | 3 | 3 | 1 | 80 | 77 | 74 | 36 | 34 | 33 |
| ST8_3_t4 | 35 | 21 | 17 | 16 | 1 | 124 | 117 | 105 | 96 | 87 | 82 |
| ST8_1_n2 | 11 | 7 | 9 | 9 | 1 | 17 | 16 | 16 | 16 | 16 | 16 |
| ST8_1_t21 | 14 | 10 | 2 | 2 | 1 | 28 | 27 | 27 | 26 | 25 | 25 |
| ST8_1_n14 | 14 | 16 | 2 | 2 | 1 | 81 | 78 | 27 | 26 | 25 | 25 |
| ST8_1_t2 | 14 | 19 | 15 | 14 | 1 | 99 | 94 | 86 | 78 | 73 | 70 |
| ST8_1_t3 | 13 | 1 | 1 | 1 | 1 | 84 | 80 | 76 | 69 | 64 | 61 |
| ST8_1_n12 | 13 | 1 | 1 | 1 | 1 | 102 | 97 | 76 | 69 | 64 | 61 |
| ST8_1_n22 | 19 | 13 | 12 | 11 | 1 | 57 | 54 | 52 | 48 | 45 | 44 |
| ST8_1_t5 | 12 | 14 | 13 | 12 | 1 | 77 | 74 | 71 | 65 | 60 | 58 |

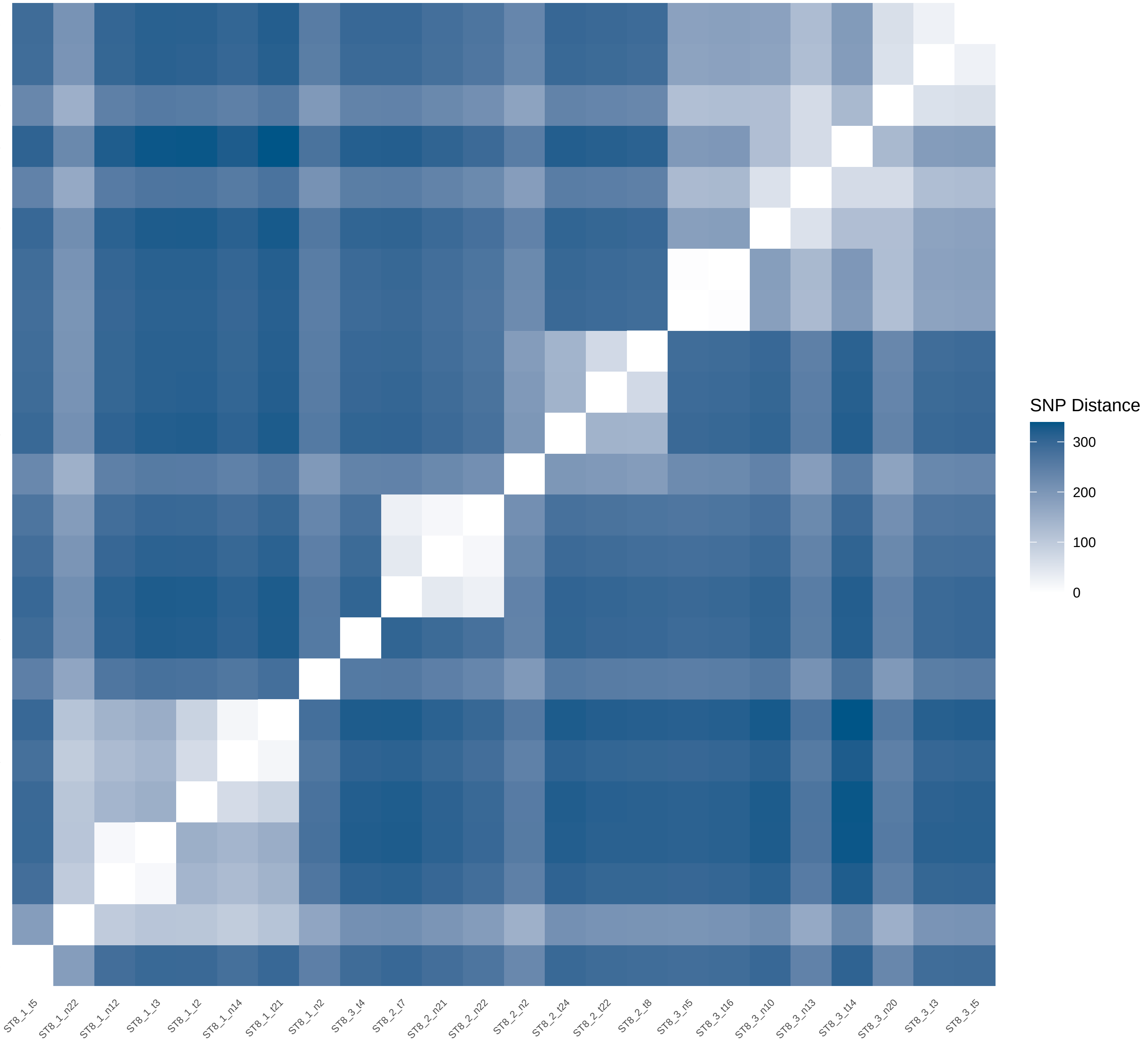
