## Supplementary Data 2 for "THRESHER: A tool for evidence-based, phylogeny-corrected SNP thresholds for whole genome molecular epidemiology": ST22_20_iter_4.pdf

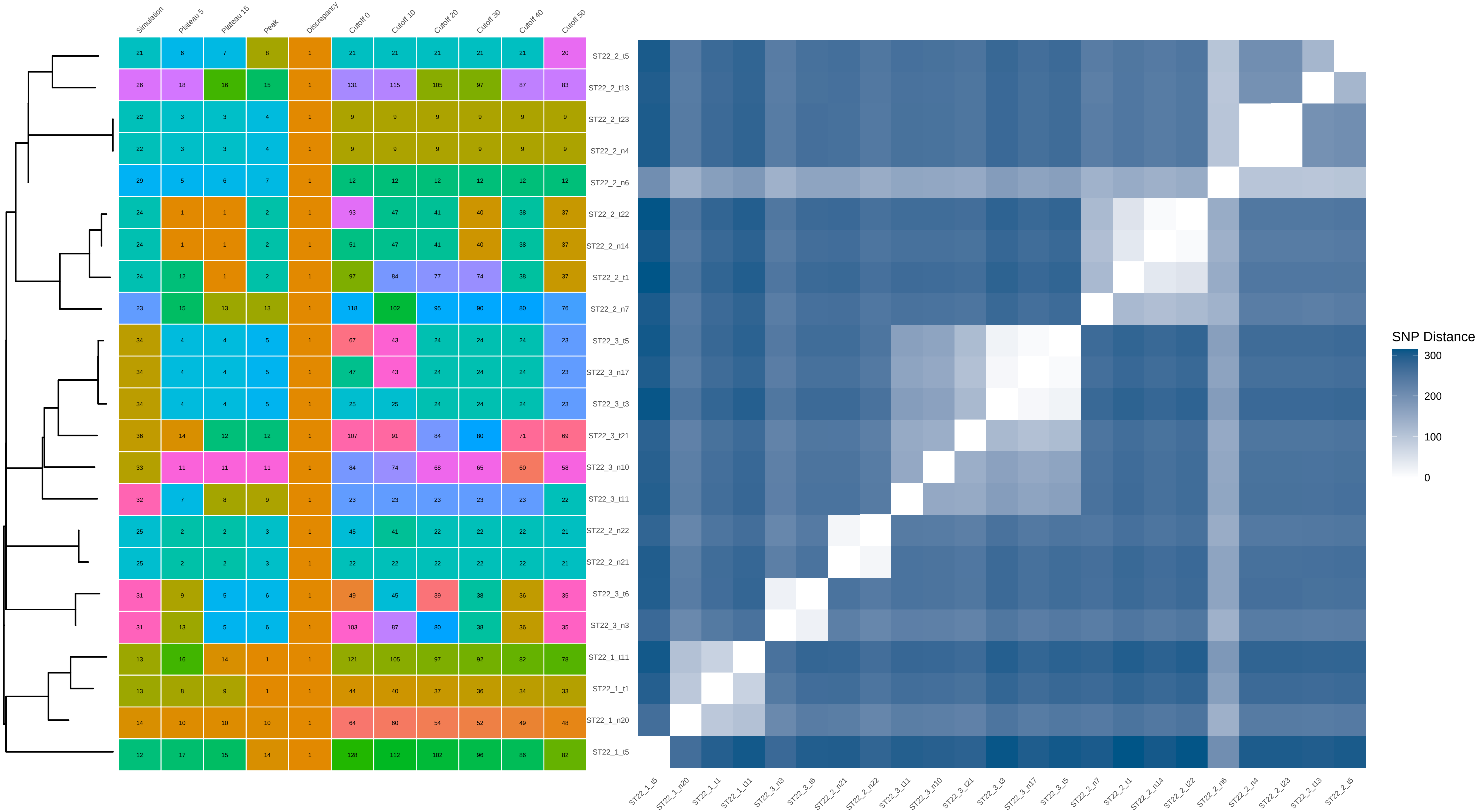


| | Simulation | Plateau 5 | Plateau 15 | Peak | Discrepancy | Cutoff 0 | Cutoff 10 | Cutoff 20 | Cutoff 30 | Cutoff 40 | Cutoff 50 |
|---|---|---|---|---|---|---|---|---|---|---|---|
| ST22_2_t5 | 21 | 6 | 7 | 8 | 1 | 21 | 21 | 21 | 21 | 21 | 20 |
| ST22_2_t13 | 26 | 18 | 16 | 15 | 1 | 131 | 115 | 105 | 97 | 87 | 83 |
| ST22_2_t23 | 22 | 3 | 3 | 4 | 1 | 9 | 9 | 9 | 9 | 9 | 9 |
| ST22_2_n4 | 22 | 3 | 3 | 4 | 1 | 9 | 9 | 9 | 9 | 9 | 9 |
| ST22_2_n6 | 29 | 5 | 6 | 7 | 1 | 12 | 12 | 12 | 12 | 12 | 12 |
| ST22_2_t22 | 24 | 1 | 1 | 2 | 1 | 93 | 47 | 41 | 40 | 38 | 37 |
| ST22_2_n14 | 24 | 1 | 1 | 2 | 1 | 51 | 47 | 41 | 40 | 38 | 37 |
| ST22_2_t1 | 24 | 12 | 1 | 2 | 1 | 97 | 84 | 77 | 74 | 38 | 37 |
| ST22_2_n7 | 23 | 15 | 13 | 13 | 1 | 118 | 102 | 95 | 90 | 80 | 76 |
| ST22_3_t5 | 34 | 4 | 4 | 5 | 1 | 67 | 43 | 24 | 24 | 24 | 23 |
| ST22_3_n17 | 34 | 4 | 4 | 5 | 1 | 47 | 43 | 24 | 24 | 24 | 23 |
| ST22_3_t3 | 34 | 4 | 4 | 5 | 1 | 25 | 25 | 24 | 24 | 24 | 23 |
| ST22_3_t21 | 36 | 14 | 12 | 12 | 1 | 107 | 91 | 84 | 80 | 71 | 69 |
| ST22_3_n10 | 33 | 11 | 11 | 11 | 1 | 84 | 74 | 68 | 65 | 60 | 58 |
| ST22_3_t11 | 32 | 7 | 8 | 9 | 1 | 23 | 23 | 23 | 23 | 23 | 22 |
| ST22_2_n22 | 25 | 2 | 2 | 3 | 1 | 45 | 41 | 22 | 22 | 22 | 21 |
| ST22_2_n21 | 25 | 2 | 2 | 3 | 1 | 22 | 22 | 22 | 22 | 22 | 21 |
| ST22_3_t6 | 31 | 9 | 5 | 6 | 1 | 49 | 45 | 39 | 38 | 36 | 35 |
| ST22_3_n3 | 31 | 13 | 5 | 6 | 1 | 103 | 87 | 80 | 38 | 36 | 35 |
| ST22_1_t11 | 13 | 16 | 14 | 1 | 1 | 121 | 105 | 97 | 92 | 82 | 78 |
| ST22_1_t1 | 13 | 8 | 9 | 1 | 1 | 44 | 40 | 37 | 36 | 34 | 33 |
| ST22_1_n20 | 14 | 10 | 10 | 10 | 1 | 64 | 60 | 54 | 52 | 49 | 48 |
| ST22_1_t5 | 12 | 17 | 15 | 14 | 1 | 128 | 112 | 102 | 96 | 86 | 82 |
