## Supplementary Data 2 for "THRESHER: A tool for evidence-based, phylogeny-corrected SNP thresholds for whole genome molecular epidemiology": ST22_20_iter_5.pdf

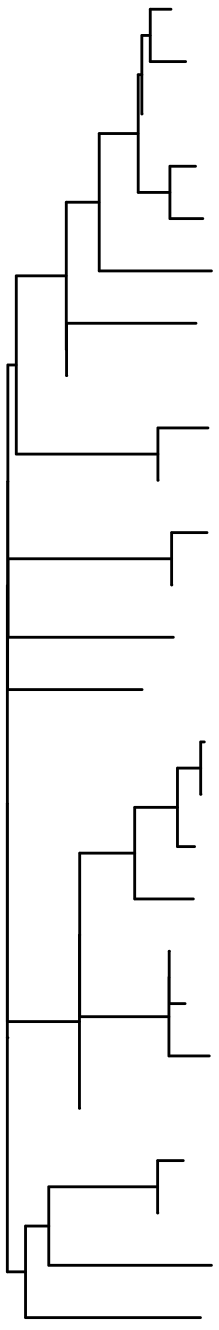

| | Simulation | Plateau 5 | Plateau 15 | Peak | Discrepancy | Cutoff 0 | Cutoff 10 | Cutoff 20 | Cutoff 30 | Cutoff 40 | Cutoff 50 |
|---|---|---|---|---|---|---|---|---|---|---|---|
| ST22_1_n14 | 14 | 23 | 2 | 2 | 1 | 121 | 114 | 103 | 81 | 2 | 2 |
| ST22_1_t8 | 14 | 3 | 2 | 2 | 1 | 2 | 2 | 2 | 2 | 2 | 2 |
| ST22_1_n17 | 14 | 17 | 2 | 2 | 1 | 101 | 95 | 87 | 81 | 2 | 2 |
| ST22_1_t24 | 14 | 9 | 2 | 4 | 1 | 25 | 25 | 24 | 23 | 23 | 2 |
| ST22_1_t22 | 14 | 22 | 2 | 4 | 1 | 114 | 107 | 97 | 89 | 77 | 2 |
| ST22_1_t11 | 13 | 21 | 15 | 16 | 1 | 113 | 106 | 96 | 88 | 76 | 67 |
| ST22_1_t7 | 16 | 15 | 13 | 14 | 1 | 88 | 86 | 78 | 72 | 64 | 56 |
| ST22_1_n22 | 19 | 8 | 9 | 10 | 1 | 20 | 20 | 20 | 19 | 19 | 19 |
| ST22_1_t12 | 12 | 4 | 1 | 1 | 1 | 9 | 9 | 9 | 9 | 9 | 9 |
| ST22_1_n11 | 12 | 16 | 1 | 1 | 1 | 93 | 90 | 82 | 76 | 9 | 9 |
| ST22_1_t14 | 11 | 12 | 3 | 3 | 1 | 50 | 50 | 47 | 43 | 42 | 37 |
| ST22_1_n2 | 11 | 19 | 3 | 3 | 1 | 105 | 99 | 91 | 43 | 42 | 37 |
| ST22_2_t12 | 25 | 14 | 12 | 13 | 1 | 72 | 70 | 66 | 61 | 56 | 50 |
| ST22_3_n3 | 31 | 11 | 11 | 12 | 1 | 47 | 47 | 44 | 40 | 39 | 34 |
| ST22_3_t15 | 33 | 2 | 6 | 7 | 1 | 22 | 22 | 22 | 21 | 21 | 20 |
| ST22_3_n9 | 33 | 2 | 6 | 7 | 1 | 124 | 22 | 22 | 21 | 21 | 20 |
| ST22_3_t18 | 33 | 18 | 6 | 7 | 1 | 102 | 96 | 88 | 82 | 21 | 20 |
| ST22_3_t25 | 33 | 6 | 8 | 9 | 1 | 14 | 14 | 14 | 14 | 14 | 14 |
| ST22_3_n18 | 34 | 1 | 5 | 6 | 1 | 127 | 119 | 8 | 8 | 8 | 8 |
| ST22_3_n17 | 34 | 1 | 5 | 6 | 1 | 8 | 8 | 8 | 8 | 8 | 8 |
| ST22_3_t23 | 34 | 7 | 5 | 6 | 1 | 15 | 15 | 15 | 8 | 8 | 8 |
| ST22_3_n22 | 39 | 5 | 7 | 8 | 1 | 10 | 10 | 10 | 10 | 10 | 10 |
| ST22_2_t5 | 21 | 24 | 4 | 5 | 3 | 128 | 120 | 48 | 44 | 43 | 38 |
| ST22_2_n1 | 21 | 13 | 4 | 5 | 3 | 51 | 51 | 48 | 44 | 43 | 38 |
| ST22_2_n3 | 22 | 10 | 10 | 11 | 4 | 40 | 40 | 38 | 34 | 34 | 30 |
| ST22_2_t3 | 23 | 20 | 14 | 15 | 6 | 111 | 105 | 95 | 87 | 75 | 66 |

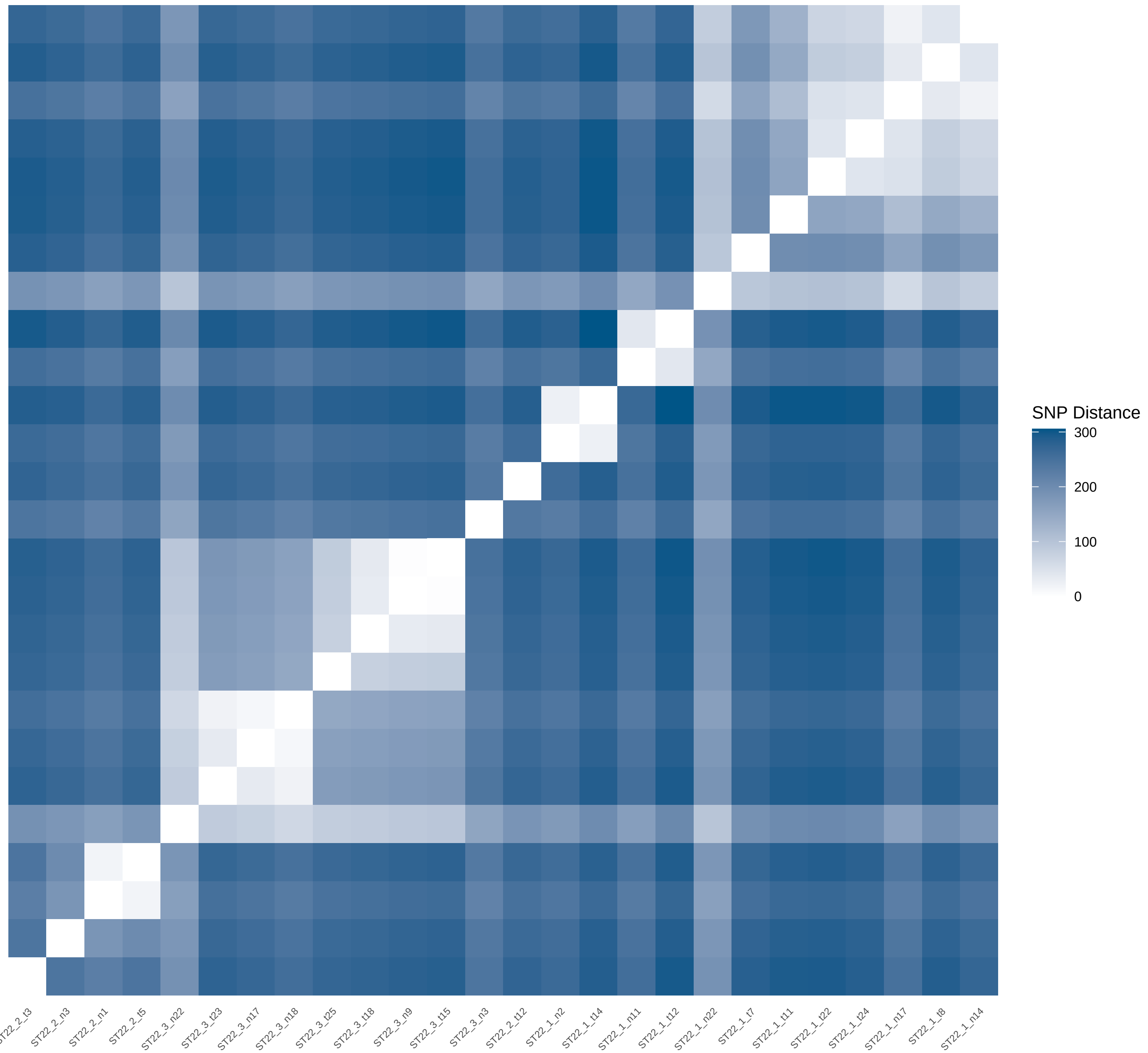
