## Supplementary Data 2 for "THRESHER: A tool for evidence-based, phylogeny-corrected SNP thresholds for whole genome molecular epidemiology": ST22_20_iter_6.pdf

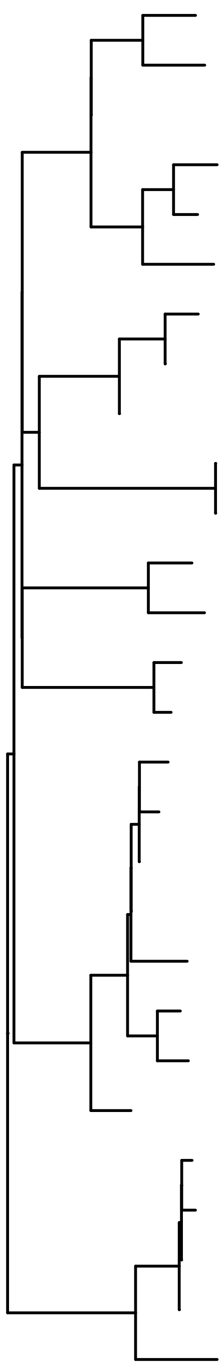

| | Simulation | Plateau 5 | Plateau 15 | Peak | Discrepancy | Cutoff 0 | Cutoff 10 | Cutoff 20 | Cutoff 30 | Cutoff 40 | Cutoff 50 |
|---|---|---|---|---|---|---|---|---|---|---|---|
| ST22_3_t19 | 33 | 17 | 17 | 7 | 1 | 79 | 76 | 67 | 61 | 58 | 54 |
| ST22_3_n9 | 33 | 20 | 20 | 7 | 1 | 90 | 85 | 76 | 68 | 62 | 57 |
| ST22_3_n22 | 39 | 12 | 12 | 7 | 1 | 52 | 52 | 50 | 47 | 45 | 43 |
| ST22_3_n14 | 34 | 14 | 14 | 6 | 1 | 65 | 63 | 57 | 53 | 51 | 27 |
| ST22_3_t22 | 34 | 7 | 7 | 6 | 1 | 30 | 30 | 29 | 28 | 28 | 27 |
| ST22_3_t7 | 34 | 25 | 25 | 10 | 1 | 123 | 112 | 101 | 90 | 79 | 69 |
| ST22_2_t4 | 24 | 16 | 16 | 3 | 1 | 77 | 74 | 65 | 2 | 2 | 2 |
| ST22_2_n17 | 24 | 4 | 4 | 3 | 1 | 2 | 2 | 2 | 2 | 2 | 2 |
| ST22_2_n19 | 212 | 21 | 21 | 3 | 1 | 94 | 88 | 79 | 70 | 2 | 2 |
| ST22_2_n4 | 22 | 3 | 3 | 5 | 1 | 3 | 3 | 3 | 3 | 3 | 3 |
| ST22_2_n3 | 22 | 3 | 3 | 5 | 1 | 3 | 3 | 3 | 3 | 3 | 3 |
| ST22_3_t6 | 31 | 8 | 8 | 8 | 1 | 31 | 31 | 30 | 29 | 29 | 28 |
| ST22_3_t1 | 31 | 10 | 10 | 8 | 1 | 41 | 41 | 40 | 39 | 37 | 35 |
| ST22_2_t7 | 25 | 19 | 19 | 4 | 1 | 88 | 84 | 75 | 67 | 12 | 12 |
| ST22_2_n21 | 25 | 5 | 5 | 4 | 1 | 12 | 12 | 12 | 12 | 12 | 12 |
| ST22_1_n15 | 14 | 1 | 1 | 1 | 1 | 86 | 82 | 73 | 24 | 24 | 23 |
| ST22_1_n14 | 14 | 1 | 1 | 1 | 1 | 25 | 25 | 24 | 24 | 24 | 23 |
| ST22_1_n16 | 14 | 1 | 1 | 1 | 1 | 103 | 56 | 24 | 24 | 24 | 23 |
| ST22_1_n17 | 14 | 1 | 1 | 1 | 1 | 57 | 56 | 24 | 24 | 24 | 23 |
| ST22_1_t25 | 14 | 15 | 15 | 1 | 1 | 76 | 73 | 64 | 59 | 24 | 23 |
| ST22_1_t24 | 14 | 23 | 23 | 1 | 1 | 116 | 105 | 95 | 84 | 75 | 23 |
| ST22_1_t22 | 14 | 22 | 22 | 1 | 1 | 106 | 97 | 87 | 77 | 69 | 23 |
| ST22_1_n13 | 13 | 13 | 13 | 1 | 1 | 61 | 60 | 54 | 50 | 48 | 46 |
| ST22_1_t6 | 12 | 2 | 2 | 2 | 2 | 50 | 50 | 49 | 46 | 44 | 42 |
| ST22_1_t5 | 12 | 2 | 2 | 2 | 2 | 117 | 106 | 49 | 46 | 44 | 42 |
| ST22_1_n6 | 12 | 2 | 2 | 2 | 2 | 56 | 55 | 49 | 46 | 44 | 42 |
| ST22_1_n9 | 12 | 2 | 2 | 2 | 2 | 62 | 55 | 49 | 46 | 44 | 42 |
| ST22_1_t13 | 12 | 9 | 9 | 9 | 2 | 40 | 40 | 39 | 38 | 36 | 34 |

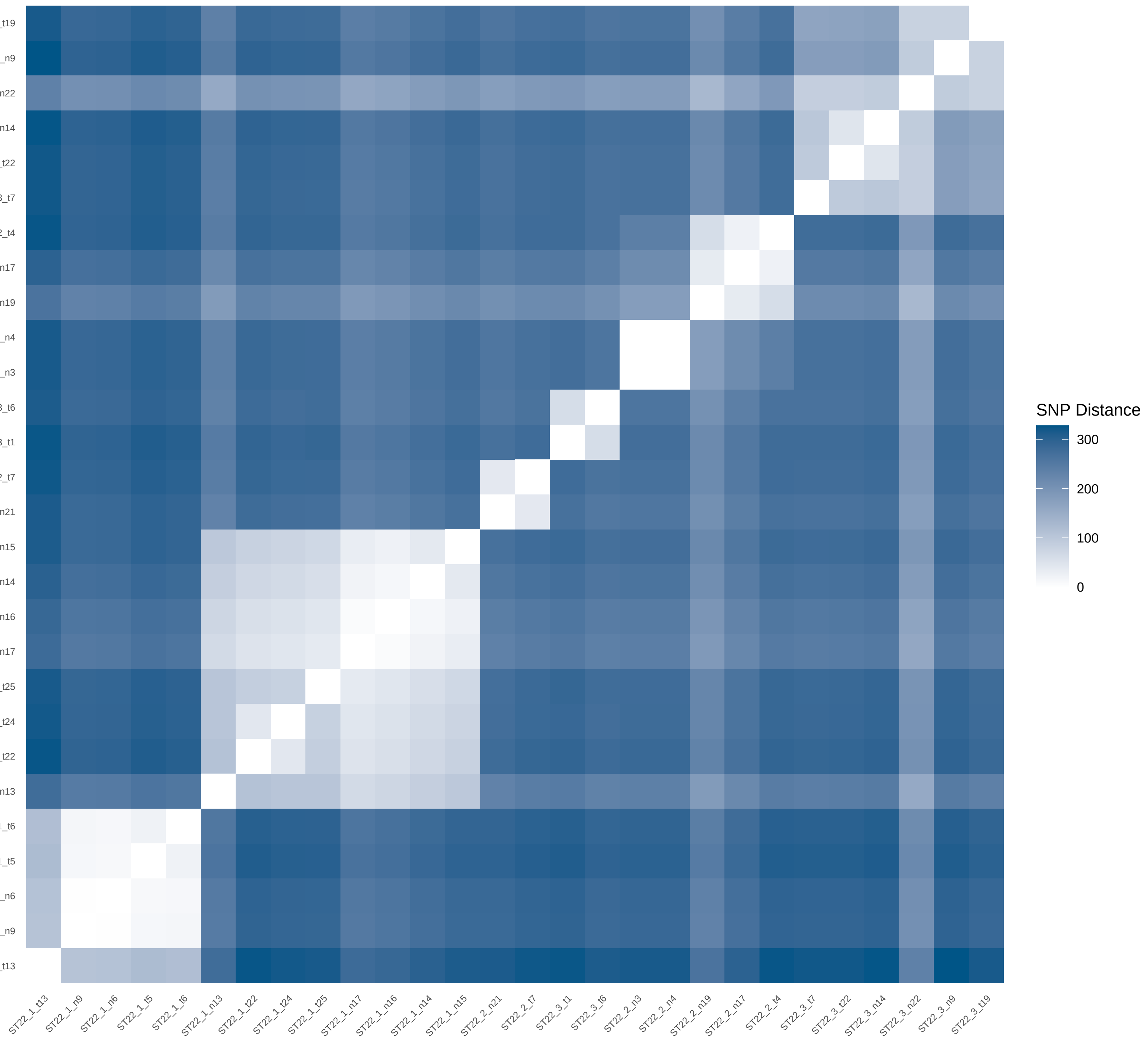
