## Supplementary Data 2 for "THRESHER: A tool for evidence-based, phylogeny-corrected SNP thresholds for whole genome molecular epidemiology": ST22_40_iter_2.pdf

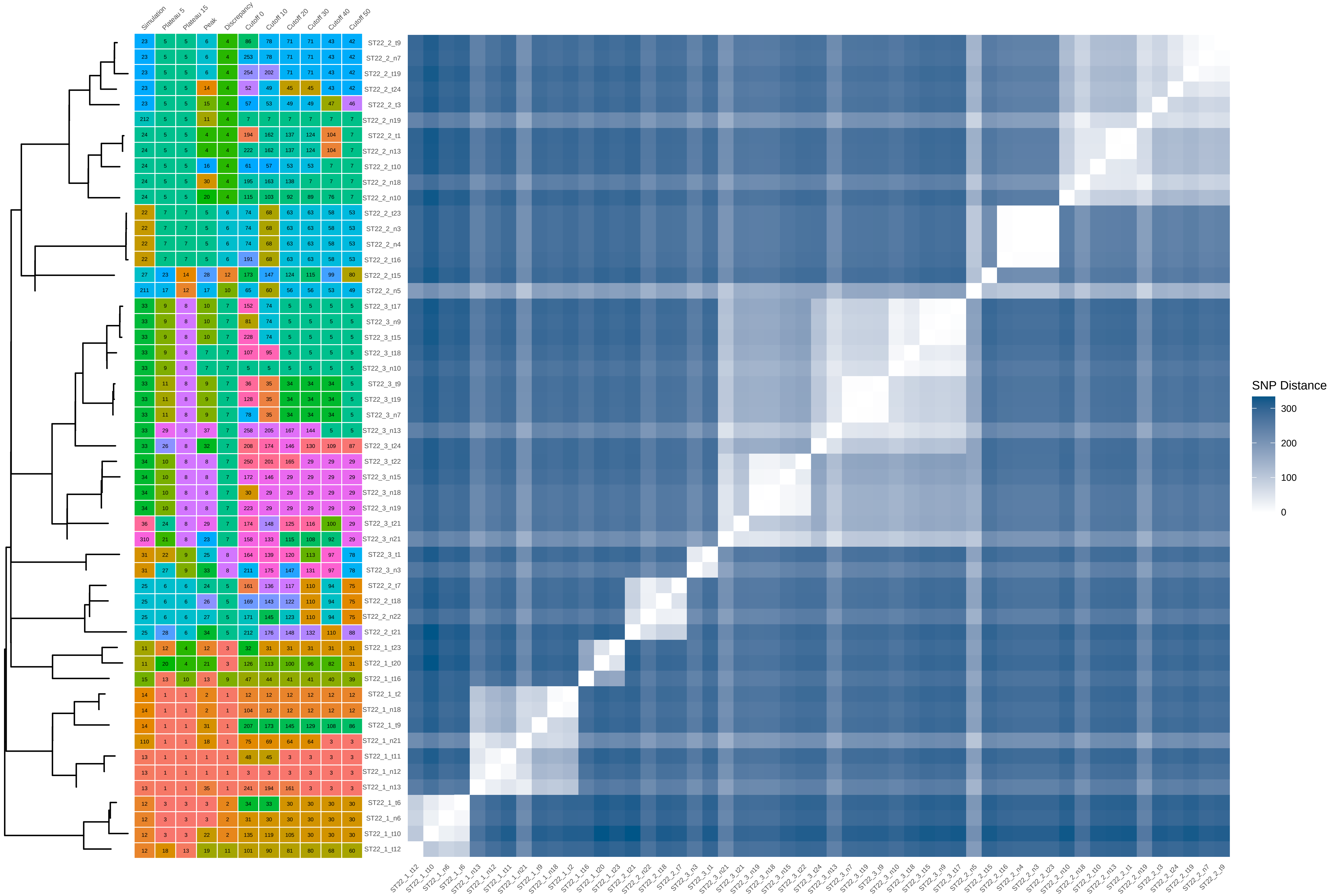


| Sample | Simulation | Plateau 5 | Plateau 15 | Peak | Discrepancy | Cutoff 0 | Cutoff 10 | Cutoff 20 | Cutoff 30 | Cutoff 40 | Cutoff 50 |
|---|---|---|---|---|---|---|---|---|---|---|---|
| ST22_2_t9 | 23 | 5 | 5 | 6 | 4 | 86 | 78 | 71 | 71 | 43 | 42 |
| ST22_2_n7 | 23 | 5 | 5 | 6 | 4 | 253 | 78 | 71 | 71 | 43 | 42 |
| ST22_2_t19 | 23 | 5 | 5 | 6 | 4 | 254 | 202 | 71 | 71 | 43 | 42 |
| ST22_2_t24 | 23 | 5 | 5 | 14 | 4 | 52 | 49 | 45 | 45 | 43 | 42 |
| ST22_2_t3 | 23 | 5 | 5 | 15 | 4 | 57 | 53 | 49 | 49 | 47 | 46 |
| ST22_2_n19 | 212 | 5 | 5 | 11 | 4 | 7 | 7 | 7 | 7 | 7 | 7 |
| ST22_2_t1 | 24 | 5 | 5 | 4 | 4 | 194 | 162 | 137 | 124 | 104 | 7 |
| ST22_2_n13 | 24 | 5 | 5 | 4 | 4 | 222 | 162 | 137 | 124 | 104 | 7 |
| ST22_2_t10 | 24 | 5 | 5 | 16 | 4 | 61 | 57 | 53 | 53 | 7 | 7 |
| ST22_2_n18 | 24 | 5 | 5 | 30 | 4 | 195 | 163 | 138 | 7 | 7 | 7 |
| ST22_2_n10 | 24 | 5 | 5 | 20 | 4 | 115 | 103 | 92 | 89 | 76 | 7 |
| ST22_2_t23 | 22 | 7 | 7 | 5 | 6 | 74 | 68 | 63 | 63 | 58 | 53 |
| ST22_2_n3 | 22 | 7 | 7 | 5 | 6 | 74 | 68 | 63 | 63 | 58 | 53 |
| ST22_2_n4 | 22 | 7 | 7 | 5 | 6 | 74 | 68 | 63 | 63 | 58 | 53 |
| ST22_2_t16 | 22 | 7 | 7 | 5 | 6 | 191 | 68 | 63 | 63 | 58 | 53 |
| ST22_2_t15 | 27 | 23 | 14 | 28 | 12 | 173 | 147 | 124 | 115 | 99 | 80 |
| ST22_2_n5 | 211 | 17 | 12 | 17 | 10 | 65 | 60 | 56 | 56 | 53 | 49 |
| ST22_3_t17 | 33 | 9 | 8 | 10 | 7 | 152 | 74 | 5 | 5 | 5 | 5 |
| ST22_3_n9 | 33 | 9 | 8 | 10 | 7 | 81 | 74 | 5 | 5 | 5 | 5 |
| ST22_3_t15 | 33 | 9 | 8 | 10 | 7 | 228 | 74 | 5 | 5 | 5 | 5 |
| ST22_3_t18 | 33 | 9 | 8 | 7 | 7 | 107 | 95 | 5 | 5 | 5 | 5 |
| ST22_3_n10 | 33 | 9 | 8 | 7 | 7 | 5 | 5 | 5 | 5 | 5 | 5 |
| ST22_3_t9 | 33 | 11 | 8 | 9 | 7 | 36 | 35 | 34 | 34 | 34 | 5 |
| ST22_3_t19 | 33 | 11 | 8 | 9 | 7 | 128 | 35 | 34 | 34 | 34 | 5 |
| ST22_3_n7 | 33 | 11 | 8 | 9 | 7 | 78 | 35 | 34 | 34 | 34 | 5 |
| ST22_3_n13 | 33 | 29 | 8 | 37 | 7 | 258 | 205 | 167 | 144 | 5 | 5 |
| ST22_3_t24 | 33 | 26 | 8 | 32 | 7 | 208 | 174 | 146 | 130 | 109 | 87 |
| ST22_3_t22 | 34 | 10 | 8 | 8 | 7 | 250 | 201 | 165 | 29 | 29 | 29 |
| ST22_3_n15 | 34 | 10 | 8 | 8 | 7 | 172 | 146 | 29 | 29 | 29 | 29 |
| ST22_3_n18 | 34 | 10 | 8 | 8 | 7 | 30 | 29 | 29 | 29 | 29 | 29 |
| ST22_3_n19 | 34 | 10 | 8 | 8 | 7 | 223 | 29 | 29 | 29 | 29 | 29 |
| ST22_3_t21 | 36 | 24 | 8 | 29 | 7 | 174 | 148 | 125 | 116 | 100 | 29 |
| ST22_3_n21 | 310 | 21 | 8 | 23 | 7 | 158 | 133 | 115 | 108 | 92 | 29 |
| ST22_3_t1 | 31 | 22 | 9 | 25 | 8 | 164 | 139 | 120 | 113 | 97 | 78 |
| ST22_3_n3 | 31 | 27 | 9 | 33 | 8 | 211 | 175 | 147 | 131 | 97 | 78 |
| ST22_2_t7 | 25 | 6 | 6 | 24 | 5 | 161 | 136 | 117 | 110 | 94 | 75 |
| ST22_2_t18 | 25 | 6 | 6 | 26 | 5 | 169 | 143 | 122 | 110 | 94 | 75 |
| ST22_2_n22 | 25 | 6 | 6 | 27 | 5 | 171 | 145 | 123 | 110 | 94 | 75 |
| ST22_2_t21 | 25 | 28 | 6 | 34 | 5 | 212 | 176 | 148 | 132 | 110 | 88 |
| ST22_1_t23 | 11 | 12 | 4 | 12 | 3 | 32 | 31 | 31 | 31 | 31 | 31 |
| ST22_1_t20 | 11 | 20 | 4 | 21 | 3 | 126 | 113 | 100 | 96 | 82 | 31 |
| ST22_1_t16 | 15 | 13 | 10 | 13 | 9 | 47 | 44 | 41 | 41 | 40 | 39 |
| ST22_1_t2 | 14 | 1 | 1 | 2 | 1 | 12 | 12 | 12 | 12 | 12 | 12 |
| ST22_1_n18 | 14 | 1 | 1 | 2 | 1 | 104 | 12 | 12 | 12 | 12 | 12 |
| ST22_1_t9 | 14 | 1 | 1 | 31 | 1 | 207 | 173 | 145 | 129 | 108 | 86 |
| ST22_1_n21 | 110 | 1 | 1 | 18 | 1 | 75 | 69 | 64 | 64 | 3 | 3 |
| ST22_1_t11 | 13 | 1 | 1 | 1 | 1 | 48 | 45 | 3 | 3 | 3 | 3 |
| ST22_1_n12 | 13 | 1 | 1 | 1 | 1 | 3 | 3 | 3 | 3 | 3 | 3 |
| ST22_1_n13 | 13 | 1 | 1 | 35 | 1 | 241 | 194 | 161 | 3 | 3 | 3 |
| ST22_1_t6 | 12 | 3 | 3 | 3 | 2 | 34 | 33 | 30 | 30 | 30 | 30 |
| ST22_1_n6 | 12 | 3 | 3 | 3 | 2 | 31 | 30 | 30 | 30 | 30 | 30 |
| ST22_1_t10 | 12 | 3 | 3 | 22 | 2 | 135 | 119 | 105 | 30 | 30 | 30 |
| ST22_1_t12 | 12 | 18 | 13 | 19 | 11 | 101 | 90 | 81 | 80 | 68 | 60 |
