## Supplementary Data 2 for "THRESHER: A tool for evidence-based, phylogeny-corrected SNP thresholds for whole genome molecular epidemiology": ST22_40_iter_3.pdf

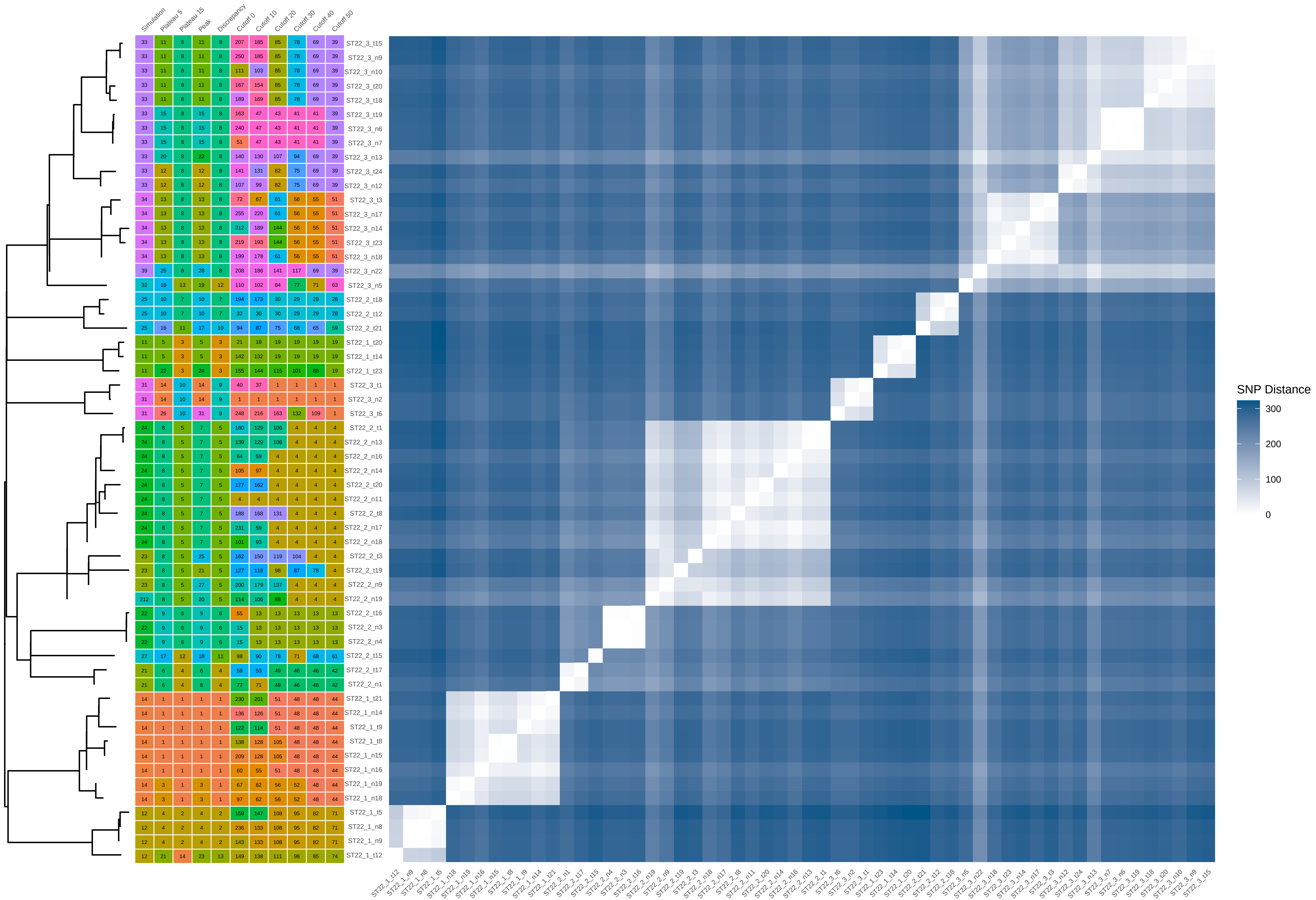


| Sample | Simulation | Plateau 5 | Plateau 15 | Peak | Discrepancy | Cutoff 0 | Cutoff 10 | Cutoff 20 | Cutoff 30 | Cutoff 40 | Cutoff 50 |
|---|---|---|---|---|---|---|---|---|---|---|---|
| ST22_3_t15 | 33 | 11 | 8 | 11 | 8 | 207 | 185 | 85 | 78 | 69 | 39 |
| ST22_3_n9 | 33 | 11 | 8 | 11 | 8 | 250 | 185 | 85 | 78 | 69 | 39 |
| ST22_3_n10 | 33 | 11 | 8 | 11 | 8 | 111 | 103 | 85 | 78 | 69 | 39 |
| ST22_3_t20 | 33 | 11 | 8 | 11 | 8 | 167 | 154 | 85 | 78 | 69 | 39 |
| ST22_3_t18 | 33 | 11 | 8 | 11 | 8 | 189 | 169 | 85 | 78 | 69 | 39 |
| ST22_3_t19 | 33 | 15 | 8 | 15 | 8 | 163 | 47 | 43 | 41 | 41 | 39 |
| ST22_3_n6 | 33 | 15 | 8 | 15 | 8 | 240 | 47 | 43 | 41 | 41 | 39 |
| ST22_3_n7 | 33 | 15 | 8 | 15 | 8 | 51 | 47 | 43 | 41 | 41 | 39 |
| ST22_3_n13 | 33 | 20 | 8 | 22 | 8 | 140 | 130 | 107 | 94 | 69 | 39 |
| ST22_3_t24 | 33 | 12 | 8 | 12 | 8 | 141 | 131 | 82 | 75 | 69 | 39 |
| ST22_3_n12 | 33 | 12 | 8 | 12 | 8 | 107 | 99 | 82 | 75 | 69 | 39 |
| ST22_3_t3 | 34 | 13 | 8 | 13 | 8 | 72 | 67 | 61 | 56 | 55 | 51 |
| ST22_3_n17 | 34 | 13 | 8 | 13 | 8 | 255 | 220 | 61 | 56 | 55 | 51 |
| ST22_3_n14 | 34 | 13 | 8 | 13 | 8 | 212 | 189 | 144 | 56 | 55 | 51 |
| ST22_3_t23 | 34 | 13 | 8 | 13 | 8 | 219 | 193 | 144 | 56 | 55 | 51 |
| ST22_3_n18 | 34 | 13 | 8 | 13 | 8 | 199 | 178 | 61 | 56 | 55 | 51 |
| ST22_3_n22 | 39 | 25 | 8 | 28 | 8 | 208 | 186 | 141 | 117 | 69 | 39 |
| ST22_3_n5 | 32 | 18 | 13 | 19 | 12 | 110 | 102 | 84 | 77 | 71 | 63 |
| ST22_2_t18 | 25 | 10 | 7 | 10 | 7 | 194 | 173 | 30 | 29 | 29 | 28 |
| ST22_2_t12 | 25 | 10 | 7 | 10 | 7 | 32 | 30 | 30 | 29 | 29 | 28 |
| ST22_2_t21 | 25 | 16 | 11 | 17 | 10 | 94 | 87 | 75 | 68 | 65 | 59 |
| ST22_1_t20 | 11 | 5 | 3 | 5 | 3 | 21 | 19 | 19 | 19 | 19 | 19 |
| ST22_1_t14 | 11 | 5 | 3 | 5 | 3 | 142 | 132 | 19 | 19 | 19 | 19 |
| ST22_1_t23 | 11 | 22 | 3 | 24 | 3 | 155 | 144 | 115 | 101 | 88 | 19 |
| ST22_3_t1 | 31 | 14 | 10 | 14 | 9 | 40 | 37 | 1 | 1 | 1 | 1 |
| ST22_3_n2 | 31 | 14 | 10 | 14 | 9 | 1 | 1 | 1 | 1 | 1 | 1 |
| ST22_3_t6 | 31 | 26 | 10 | 31 | 9 | 248 | 216 | 163 | 132 | 109 | 1 |
| ST22_2_t1 | 24 | 8 | 5 | 7 | 5 | 180 | 129 | 106 | 4 | 4 | 4 |
| ST22_2_n13 | 24 | 8 | 5 | 7 | 5 | 139 | 129 | 106 | 4 | 4 | 4 |
| ST22_2_n16 | 24 | 8 | 5 | 7 | 5 | 64 | 59 | 4 | 4 | 4 | 4 |
| ST22_2_n14 | 24 | 8 | 5 | 7 | 5 | 105 | 97 | 4 | 4 | 4 | 4 |
| ST22_2_t20 | 24 | 8 | 5 | 7 | 5 | 177 | 162 | 4 | 4 | 4 | 4 |
| ST22_2_n11 | 24 | 8 | 5 | 7 | 5 | 4 | 4 | 4 | 4 | 4 | 4 |
| ST22_2_t8 | 24 | 8 | 5 | 7 | 5 | 188 | 168 | 131 | 4 | 4 | 4 |
| ST22_2_n17 | 24 | 8 | 5 | 7 | 5 | 231 | 59 | 4 | 4 | 4 | 4 |
| ST22_2_n18 | 24 | 8 | 5 | 7 | 5 | 101 | 93 | 4 | 4 | 4 | 4 |
| ST22_2_t3 | 23 | 8 | 5 | 25 | 5 | 162 | 150 | 119 | 104 | 4 | 4 |
| ST22_2_t19 | 23 | 8 | 5 | 21 | 5 | 127 | 118 | 98 | 87 | 78 | 4 |
| ST22_2_n9 | 23 | 8 | 5 | 27 | 5 | 200 | 179 | 137 | 4 | 4 | 4 |
| ST22_2_n19 | 212 | 8 | 5 | 20 | 5 | 114 | 106 | 88 | 4 | 4 | 4 |
| ST22_2_t16 | 22 | 9 | 6 | 9 | 6 | 55 | 13 | 13 | 13 | 13 | 13 |
| ST22_2_n3 | 22 | 9 | 6 | 9 | 6 | 15 | 13 | 13 | 13 | 13 | 13 |
| ST22_2_n4 | 22 | 9 | 6 | 9 | 6 | 15 | 13 | 13 | 13 | 13 | 13 |
| ST22_2_t15 | 27 | 17 | 12 | 18 | 11 | 98 | 90 | 78 | 71 | 68 | 61 |
| ST22_2_t17 | 21 | 6 | 4 | 6 | 4 | 58 | 53 | 49 | 46 | 46 | 42 |
| ST22_2_n1 | 21 | 6 | 4 | 6 | 4 | 77 | 71 | 49 | 46 | 46 | 42 |
| ST22_1_t21 | 14 | 1 | 1 | 1 | 1 | 230 | 201 | 51 | 48 | 48 | 44 |
| ST22_1_n14 | 14 | 1 | 1 | 1 | 1 | 136 | 126 | 51 | 48 | 48 | 44 |
| ST22_1_t9 | 14 | 1 | 1 | 1 | 1 | 122 | 114 | 51 | 48 | 48 | 44 |
| ST22_1_t8 | 14 | 1 | 1 | 1 | 1 | 138 | 128 | 105 | 48 | 48 | 44 |
| ST22_1_n15 | 14 | 1 | 1 | 1 | 1 | 209 | 128 | 105 | 48 | 48 | 44 |
| ST22_1_n16 | 14 | 1 | 1 | 1 | 1 | 60 | 55 | 51 | 48 | 48 | 44 |
| ST22_1_n19 | 14 | 3 | 1 | 3 | 1 | 67 | 62 | 56 | 52 | 48 | 44 |
| ST22_1_n18 | 14 | 3 | 1 | 3 | 1 | 97 | 62 | 56 | 52 | 48 | 44 |
| ST22_1_t5 | 12 | 4 | 2 | 4 | 2 | 159 | 147 | 108 | 95 | 82 | 71 |
| ST22_1_n8 | 12 | 4 | 2 | 4 | 2 | 236 | 133 | 108 | 95 | 82 | 71 |
| ST22_1_n9 | 12 | 4 | 2 | 4 | 2 | 143 | 133 | 108 | 95 | 82 | 71 |
| ST22_1_t12 | 12 | 21 | 14 | 23 | 13 | 149 | 138 | 111 | 98 | 85 | 74 |
