## Supplementary Data 2 for "THRESHER: A tool for evidence-based, phylogeny-corrected SNP thresholds for whole genome molecular epidemiology": ST398_20_iter_8.pdf

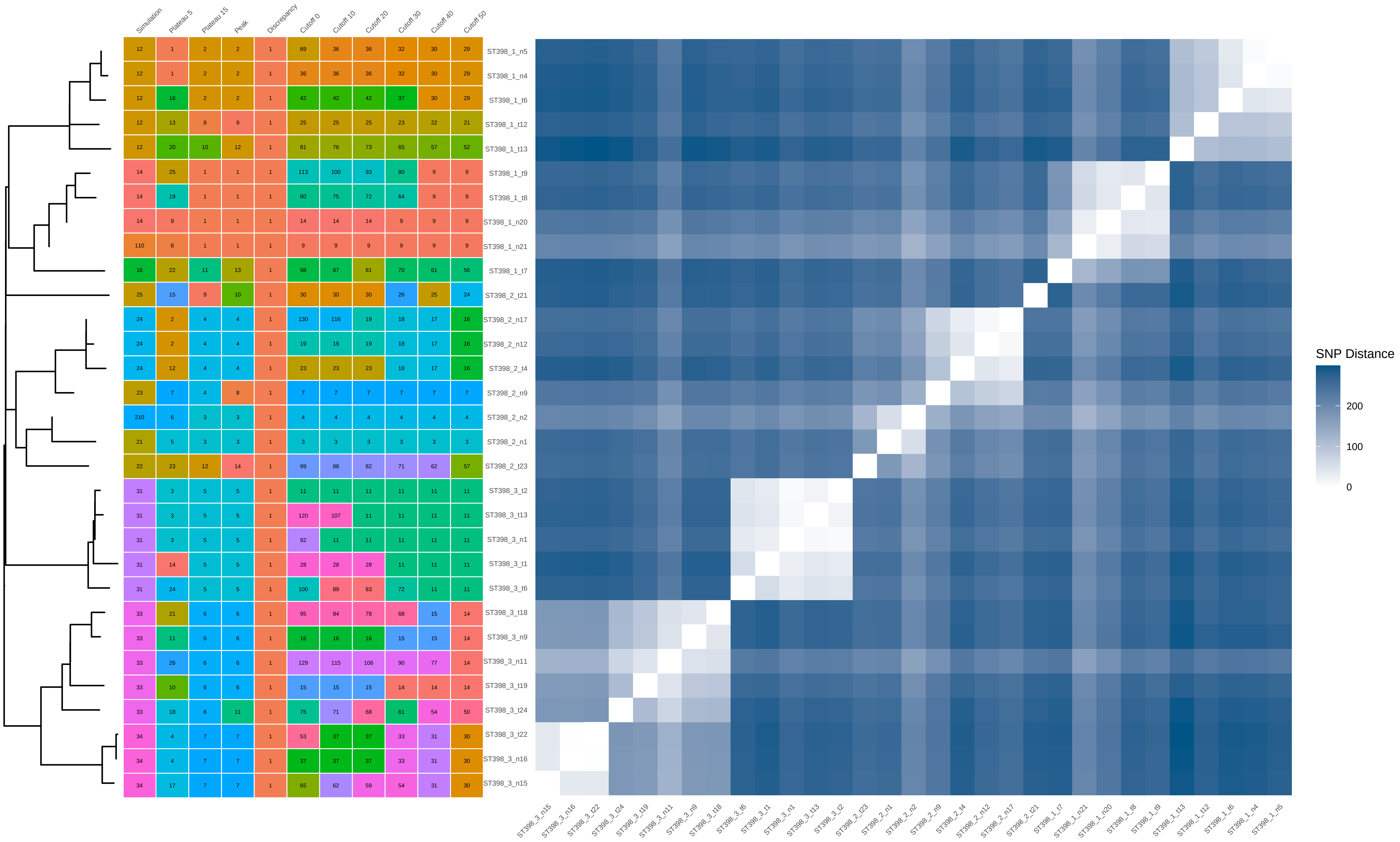


| | Simulation | Plateau 5 | Plateau 15 | Peak | Discrepancy | Cutoff 0 | Cutoff 10 | Cutoff 20 | Cutoff 30 | Cutoff 40 | Cutoff 50 |
|---|---|---|---|---|---|---|---|---|---|---|---|
| ST398_1_n5 | 12 | 1 | 2 | 2 | 1 | 69 | 36 | 36 | 32 | 30 | 29 |
| ST398_1_n4 | 12 | 1 | 2 | 2 | 1 | 36 | 36 | 36 | 32 | 30 | 29 |
| ST398_1_t6 | 12 | 16 | 2 | 2 | 1 | 42 | 42 | 42 | 37 | 30 | 29 |
| ST398_1_t12 | 12 | 13 | 8 | 9 | 1 | 25 | 25 | 25 | 23 | 22 | 21 |
| ST398_1_t13 | 12 | 20 | 10 | 12 | 1 | 81 | 76 | 73 | 65 | 57 | 52 |
| ST398_1_t9 | 14 | 25 | 1 | 1 | 1 | 113 | 100 | 93 | 80 | 9 | 9 |
| ST398_1_t8 | 14 | 19 | 1 | 1 | 1 | 80 | 75 | 72 | 64 | 9 | 9 |
| ST398_1_n20 | 14 | 9 | 1 | 1 | 1 | 14 | 14 | 14 | 9 | 9 | 9 |
| ST398_1_n21 | 110 | 8 | 1 | 1 | 1 | 9 | 9 | 9 | 9 | 9 | 9 |
| ST398_1_t7 | 16 | 22 | 11 | 13 | 1 | 98 | 87 | 81 | 70 | 61 | 56 |
| ST398_2_t21 | 25 | 15 | 9 | 10 | 1 | 30 | 30 | 30 | 26 | 25 | 24 |
| ST398_2_n17 | 24 | 2 | 4 | 4 | 1 | 130 | 116 | 19 | 18 | 17 | 16 |
| ST398_2_n12 | 24 | 2 | 4 | 4 | 1 | 19 | 19 | 19 | 18 | 17 | 16 |
| ST398_2_t4 | 24 | 12 | 4 | 4 | 1 | 23 | 23 | 23 | 18 | 17 | 16 |
| ST398_2_n9 | 23 | 7 | 4 | 8 | 1 | 7 | 7 | 7 | 7 | 7 | 7 |
| ST398_2_n2 | 210 | 6 | 3 | 3 | 1 | 4 | 4 | 4 | 4 | 4 | 4 |
| ST398_2_n1 | 21 | 5 | 3 | 3 | 1 | 3 | 3 | 3 | 3 | 3 | 3 |
| ST398_2_t23 | 22 | 23 | 12 | 14 | 1 | 99 | 88 | 82 | 71 | 62 | 57 |
| ST398_3_t2 | 31 | 3 | 5 | 5 | 1 | 11 | 11 | 11 | 11 | 11 | 11 |
| ST398_3_t13 | 31 | 3 | 5 | 5 | 1 | 120 | 107 | 11 | 11 | 11 | 11 |
| ST398_3_n1 | 31 | 3 | 5 | 5 | 1 | 92 | 11 | 11 | 11 | 11 | 11 |
| ST398_3_t1 | 31 | 14 | 5 | 5 | 1 | 28 | 28 | 28 | 11 | 11 | 11 |
| ST398_3_t6 | 31 | 24 | 5 | 5 | 1 | 100 | 89 | 83 | 72 | 11 | 11 |
| ST398_3_t18 | 33 | 21 | 6 | 6 | 1 | 95 | 84 | 78 | 68 | 15 | 14 |
| ST398_3_n9 | 33 | 11 | 6 | 6 | 1 | 16 | 16 | 16 | 15 | 15 | 14 |
| ST398_3_n11 | 33 | 26 | 6 | 6 | 1 | 129 | 115 | 106 | 90 | 77 | 14 |
| ST398_3_t19 | 33 | 10 | 6 | 6 | 1 | 15 | 15 | 15 | 14 | 14 | 14 |
| ST398_3_t24 | 33 | 18 | 6 | 11 | 1 | 75 | 71 | 68 | 61 | 54 | 50 |
| ST398_3_t22 | 34 | 4 | 7 | 7 | 1 | 53 | 37 | 37 | 33 | 31 | 30 |
| ST398_3_n16 | 34 | 4 | 7 | 7 | 1 | 37 | 37 | 37 | 33 | 31 | 30 |
| ST398_3_n15 | 34 | 17 | 7 | 7 | 1 | 65 | 62 | 59 | 54 | 31 | 30 |
