## Supplementary Data 2 for "THRESHER: A tool for evidence-based, phylogeny-corrected SNP thresholds for whole genome molecular epidemiology": ST398_20_iter_9.pdf

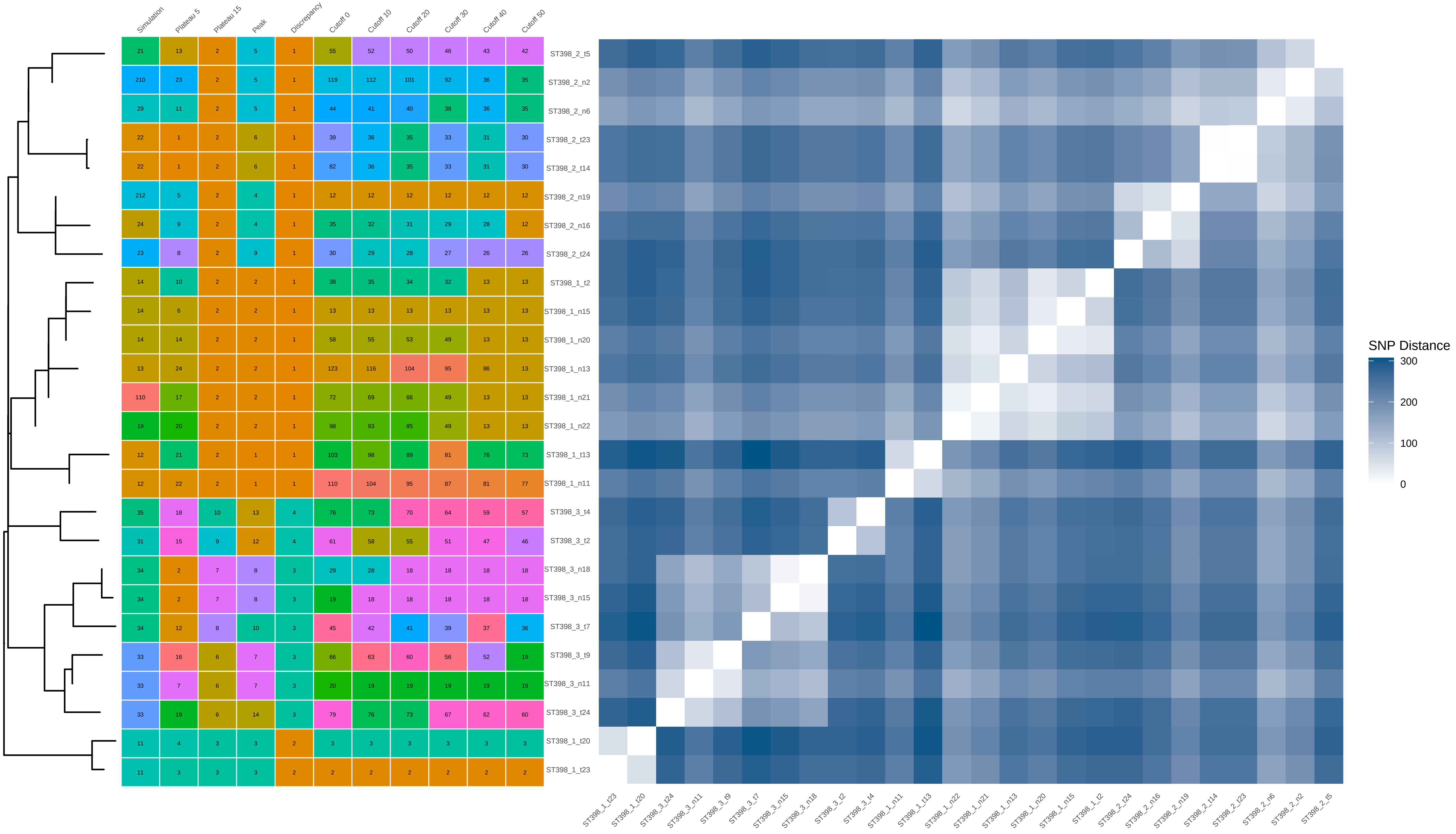

| | Simulation | Plateau 5 | Plateau 15 | Peak | Discrepancy | Cutoff 0 | Cutoff 10 | Cutoff 20 | Cutoff 30 | Cutoff 40 | Cutoff 50 |
|---|---|---|---|---|---|---|---|---|---|---|---|
| ST398_2_t5 | 21 | 13 | 2 | 5 | 1 | 55 | 52 | 50 | 46 | 43 | 42 |
| ST398_2_n2 | 210 | 23 | 2 | 5 | 1 | 119 | 112 | 101 | 92 | 36 | 35 |
| ST398_2_n6 | 29 | 11 | 2 | 5 | 1 | 44 | 41 | 40 | 38 | 36 | 35 |
| ST398_2_t23 | 22 | 1 | 2 | 6 | 1 | 39 | 36 | 35 | 33 | 31 | 30 |
| ST398_2_t14 | 22 | 1 | 2 | 6 | 1 | 82 | 36 | 35 | 33 | 31 | 30 |
| ST398_2_n19 | 212 | 5 | 2 | 4 | 1 | 12 | 12 | 12 | 12 | 12 | 12 |
| ST398_2_n16 | 24 | 9 | 2 | 4 | 1 | 35 | 32 | 31 | 29 | 28 | 12 |
| ST398_2_t24 | 23 | 8 | 2 | 9 | 1 | 30 | 29 | 28 | 27 | 26 | 26 |
| ST398_1_t2 | 14 | 10 | 2 | 2 | 1 | 38 | 35 | 34 | 32 | 13 | 13 |
| ST398_1_n15 | 14 | 6 | 2 | 2 | 1 | 13 | 13 | 13 | 13 | 13 | 13 |
| ST398_1_n20 | 14 | 14 | 2 | 2 | 1 | 58 | 55 | 53 | 49 | 13 | 13 |
| ST398_1_n13 | 13 | 24 | 2 | 2 | 1 | 123 | 116 | 104 | 95 | 86 | 13 |
| ST398_1_n21 | 110 | 17 | 2 | 2 | 1 | 72 | 69 | 66 | 49 | 13 | 13 |
| ST398_1_n22 | 19 | 20 | 2 | 2 | 1 | 98 | 93 | 85 | 49 | 13 | 13 |
| ST398_1_t13 | 12 | 21 | 2 | 1 | 1 | 103 | 98 | 89 | 81 | 76 | 73 |
| ST398_1_n11 | 12 | 22 | 2 | 1 | 1 | 110 | 104 | 95 | 87 | 81 | 77 |
| ST398_3_t4 | 35 | 18 | 10 | 13 | 4 | 76 | 73 | 70 | 64 | 59 | 57 |
| ST398_3_t2 | 31 | 15 | 9 | 12 | 4 | 61 | 58 | 55 | 51 | 47 | 46 |
| ST398_3_n18 | 34 | 2 | 7 | 8 | 3 | 29 | 28 | 18 | 18 | 18 | 18 |
| ST398_3_n15 | 34 | 2 | 7 | 8 | 3 | 19 | 18 | 18 | 18 | 18 | 18 |
| ST398_3_t7 | 34 | 12 | 8 | 10 | 3 | 45 | 42 | 41 | 39 | 37 | 36 |
| ST398_3_t9 | 33 | 16 | 6 | 7 | 3 | 66 | 63 | 60 | 56 | 52 | 19 |
| ST398_3_n11 | 33 | 7 | 6 | 7 | 3 | 20 | 19 | 19 | 19 | 19 | 19 |
| ST398_3_t24 | 33 | 19 | 6 | 14 | 3 | 79 | 76 | 73 | 67 | 62 | 60 |
| ST398_1_t20 | 11 | 4 | 3 | 3 | 2 | 3 | 3 | 3 | 3 | 3 | 3 |
| ST398_1_t23 | 11 | 3 | 3 | 3 | 2 | 2 | 2 | 2 | 2 | 2 | 2 |
