## Supplementary Data 2 for "THRESHER: A tool for evidence-based, phylogeny-corrected SNP thresholds for whole genome molecular epidemiology": ST398_40_iter_2.pdf

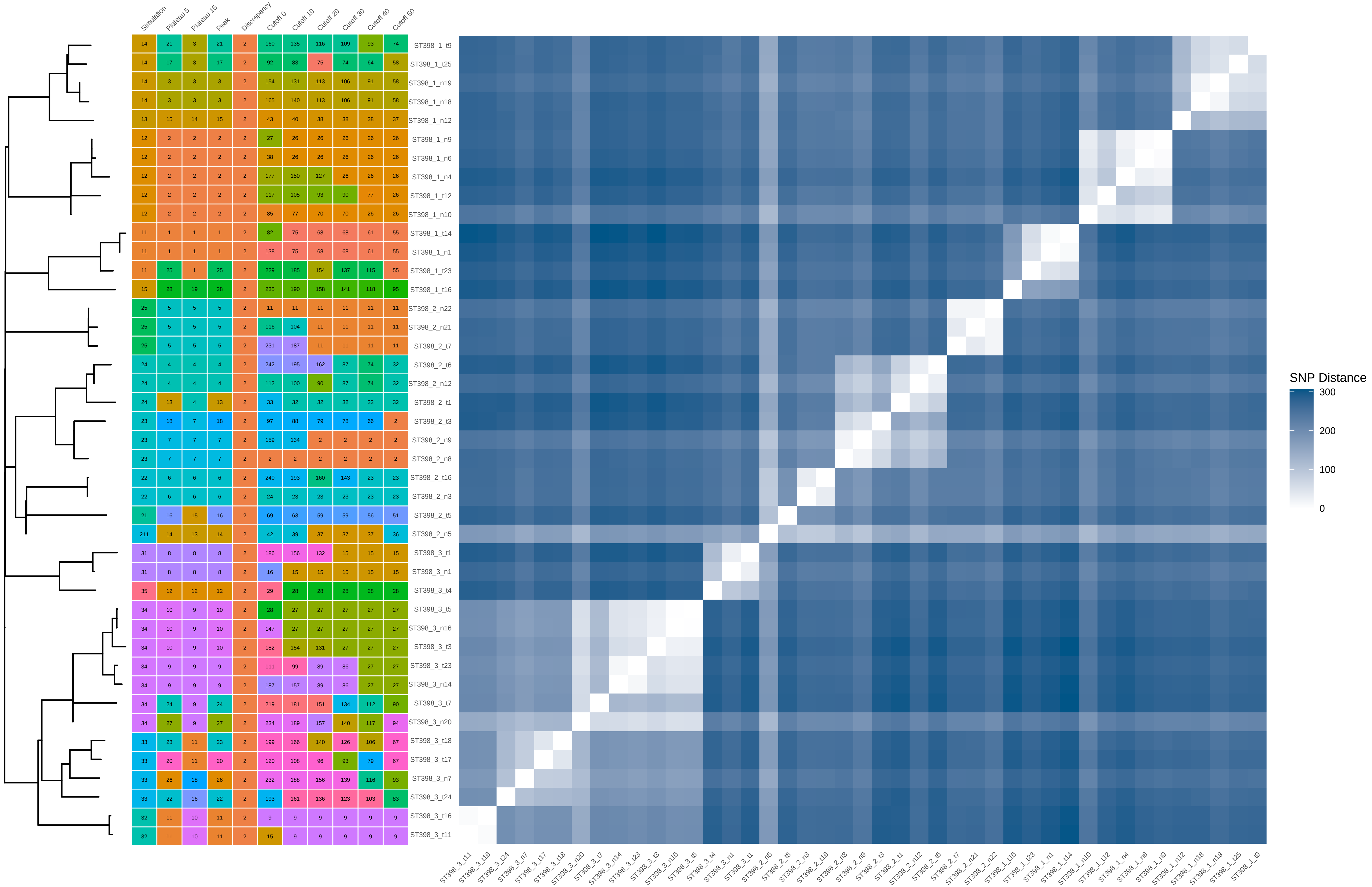


| | Simulation | Plateau 5 | Plateau 15 | Peak | Discrepancy | Cutoff 0 | Cutoff 10 | Cutoff 20 | Cutoff 30 | Cutoff 40 | Cutoff 50 |
|---|---|---|---|---|---|---|---|---|---|---|---|
| ST398_1_t9 | 14 | 21 | 3 | 21 | 2 | 160 | 135 | 116 | 109 | 93 | 74 |
| ST398_1_t25 | 14 | 17 | 3 | 17 | 2 | 92 | 83 | 75 | 74 | 64 | 58 |
| ST398_1_n19 | 14 | 3 | 3 | 3 | 2 | 154 | 131 | 113 | 106 | 91 | 58 |
| ST398_1_n18 | 14 | 3 | 3 | 3 | 2 | 165 | 140 | 113 | 106 | 91 | 58 |
| ST398_1_n12 | 13 | 15 | 14 | 15 | 2 | 43 | 40 | 38 | 38 | 38 | 37 |
| ST398_1_n9 | 12 | 2 | 2 | 2 | 2 | 27 | 26 | 26 | 26 | 26 | 26 |
| ST398_1_n6 | 12 | 2 | 2 | 2 | 2 | 38 | 26 | 26 | 26 | 26 | 26 |
| ST398_1_n4 | 12 | 2 | 2 | 2 | 2 | 177 | 150 | 127 | 26 | 26 | 26 |
| ST398_1_t12 | 12 | 2 | 2 | 2 | 2 | 117 | 105 | 93 | 90 | 77 | 26 |
| ST398_1_n10 | 12 | 2 | 2 | 2 | 2 | 85 | 77 | 70 | 70 | 26 | 26 |
| ST398_1_t14 | 11 | 1 | 1 | 1 | 2 | 82 | 75 | 68 | 68 | 61 | 55 |
| ST398_1_n1 | 11 | 1 | 1 | 1 | 2 | 138 | 75 | 68 | 68 | 61 | 55 |
| ST398_1_t23 | 11 | 25 | 1 | 25 | 2 | 229 | 185 | 154 | 137 | 115 | 55 |
| ST398_1_t16 | 15 | 28 | 19 | 28 | 2 | 235 | 190 | 158 | 141 | 118 | 95 |
| ST398_2_n22 | 25 | 5 | 5 | 5 | 2 | 11 | 11 | 11 | 11 | 11 | 11 |
| ST398_2_n21 | 25 | 5 | 5 | 5 | 2 | 116 | 104 | 11 | 11 | 11 | 11 |
| ST398_2_t7 | 25 | 5 | 5 | 5 | 2 | 231 | 187 | 11 | 11 | 11 | 11 |
| ST398_2_t6 | 24 | 4 | 4 | 4 | 2 | 242 | 195 | 162 | 87 | 74 | 32 |
| ST398_2_n12 | 24 | 4 | 4 | 4 | 2 | 112 | 100 | 90 | 87 | 74 | 32 |
| ST398_2_t1 | 24 | 13 | 4 | 13 | 2 | 33 | 32 | 32 | 32 | 32 | 32 |
| ST398_2_t3 | 23 | 18 | 7 | 18 | 2 | 97 | 88 | 79 | 78 | 66 | 2 |
| ST398_2_n9 | 23 | 7 | 7 | 7 | 2 | 159 | 134 | 2 | 2 | 2 | 2 |
| ST398_2_n8 | 23 | 7 | 7 | 7 | 2 | 2 | 2 | 2 | 2 | 2 | 2 |
| ST398_2_t16 | 22 | 6 | 6 | 6 | 2 | 240 | 193 | 160 | 143 | 23 | 23 |
| ST398_2_n3 | 22 | 6 | 6 | 6 | 2 | 24 | 23 | 23 | 23 | 23 | 23 |
| ST398_2_t5 | 21 | 16 | 15 | 16 | 2 | 69 | 63 | 59 | 59 | 56 | 51 |
| ST398_2_n5 | 211 | 14 | 13 | 14 | 2 | 42 | 39 | 37 | 37 | 37 | 36 |
| ST398_3_t1 | 31 | 8 | 8 | 8 | 2 | 186 | 156 | 132 | 15 | 15 | 15 |
| ST398_3_n1 | 31 | 8 | 8 | 8 | 2 | 16 | 15 | 15 | 15 | 15 | 15 |
| ST398_3_t4 | 35 | 12 | 12 | 12 | 2 | 29 | 28 | 28 | 28 | 28 | 28 |
| ST398_3_t5 | 34 | 10 | 9 | 10 | 2 | 28 | 27 | 27 | 27 | 27 | 27 |
| ST398_3_n16 | 34 | 10 | 9 | 10 | 2 | 147 | 27 | 27 | 27 | 27 | 27 |
| ST398_3_t3 | 34 | 10 | 9 | 10 | 2 | 182 | 154 | 131 | 27 | 27 | 27 |
| ST398_3_t23 | 34 | 9 | 9 | 9 | 2 | 111 | 99 | 89 | 86 | 27 | 27 |
| ST398_3_n14 | 34 | 9 | 9 | 9 | 2 | 187 | 157 | 89 | 86 | 27 | 27 |
| ST398_3_t7 | 34 | 24 | 9 | 24 | 2 | 219 | 181 | 151 | 134 | 112 | 90 |
| ST398_3_n20 | 34 | 27 | 9 | 27 | 2 | 234 | 189 | 157 | 140 | 117 | 94 |
| ST398_3_t18 | 33 | 23 | 11 | 23 | 2 | 199 | 166 | 140 | 126 | 106 | 67 |
| ST398_3_t17 | 33 | 20 | 11 | 20 | 2 | 120 | 108 | 96 | 93 | 79 | 67 |
| ST398_3_n7 | 33 | 26 | 18 | 26 | 2 | 232 | 188 | 156 | 139 | 116 | 93 |
| ST398_3_t24 | 33 | 22 | 16 | 22 | 2 | 193 | 161 | 136 | 123 | 103 | 83 |
| ST398_3_t16 | 32 | 11 | 10 | 11 | 2 | 9 | 9 | 9 | 9 | 9 | 9 |
| ST398_3_t11 | 32 | 11 | 10 | 11 | 2 | 15 | 9 | 9 | 9 | 9 | 9 |
