## Supplementary Data 2 for "THRESHER: A tool for evidence-based, phylogeny-corrected SNP thresholds for whole genome molecular epidemiology": ST398_40_iter_9.pdf

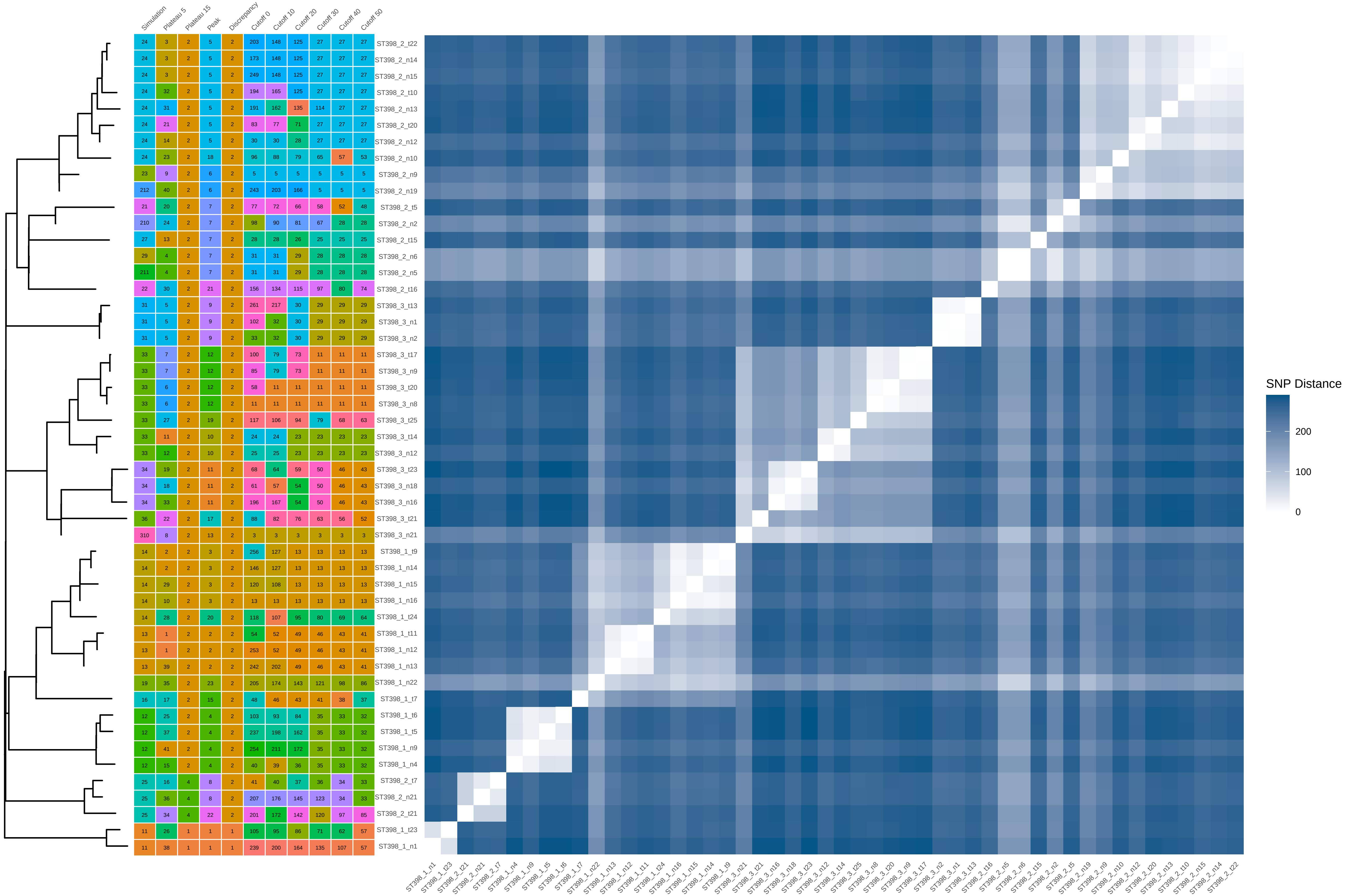


| Simulation | Plateau 5 | Plateau 15 | Peak | Discrepancy | Cutoff 0 | Cutoff 10 | Cutoff 20 | Cutoff 30 | Cutoff 40 | Cutoff 50 | |
|---|---|---|---|---|---|---|---|---|---|---|---|
| 24 | 3 | 2 | 5 | 2 | 203 | 148 | 125 | 27 | 27 | 27 | ST398_2_t22 |
| 24 | 3 | 2 | 5 | 2 | 173 | 148 | 125 | 27 | 27 | 27 | ST398_2_n14 |
| 24 | 3 | 2 | 5 | 2 | 249 | 148 | 125 | 27 | 27 | 27 | ST398_2_n15 |
| 24 | 32 | 2 | 5 | 2 | 194 | 165 | 125 | 27 | 27 | 27 | ST398_2_t10 |
| 24 | 31 | 2 | 5 | 2 | 191 | 162 | 135 | 114 | 27 | 27 | ST398_2_n13 |
| 24 | 21 | 2 | 5 | 2 | 83 | 77 | 71 | 27 | 27 | 27 | ST398_2_t20 |
| 24 | 14 | 2 | 5 | 2 | 30 | 30 | 28 | 27 | 27 | 27 | ST398_2_n12 |
| 24 | 23 | 2 | 18 | 2 | 96 | 88 | 79 | 65 | 57 | 53 | ST398_2_n10 |
| 23 | 9 | 2 | 6 | 2 | 5 | 5 | 5 | 5 | 5 | 5 | ST398_2_n9 |
| 212 | 40 | 2 | 6 | 2 | 243 | 203 | 166 | 5 | 5 | 5 | ST398_2_n19 |
| 21 | 20 | 2 | 7 | 2 | 77 | 72 | 66 | 58 | 52 | 48 | ST398_2_t5 |
| 210 | 24 | 2 | 7 | 2 | 98 | 90 | 81 | 67 | 28 | 28 | ST398_2_n2 |
| 27 | 13 | 2 | 7 | 2 | 28 | 28 | 26 | 25 | 25 | 25 | ST398_2_t15 |
| 29 | 4 | 2 | 7 | 2 | 31 | 31 | 29 | 28 | 28 | 28 | ST398_2_n6 |
| 211 | 4 | 2 | 7 | 2 | 31 | 31 | 29 | 28 | 28 | 28 | ST398_2_n5 |
| 22 | 30 | 2 | 21 | 2 | 156 | 134 | 115 | 97 | 80 | 74 | ST398_2_t16 |
| 31 | 5 | 2 | 9 | 2 | 261 | 217 | 30 | 29 | 29 | 29 | ST398_3_t13 |
| 31 | 5 | 2 | 9 | 2 | 102 | 32 | 30 | 29 | 29 | 29 | ST398_3_n1 |
| 31 | 5 | 2 | 9 | 2 | 33 | 32 | 30 | 29 | 29 | 29 | ST398_3_n2 |
| 33 | 7 | 2 | 12 | 2 | 100 | 79 | 73 | 11 | 11 | 11 | ST398_3_t17 |
| 33 | 7 | 2 | 12 | 2 | 85 | 79 | 73 | 11 | 11 | 11 | ST398_3_n9 |
| 33 | 6 | 2 | 12 | 2 | 58 | 11 | 11 | 11 | 11 | 11 | ST398_3_t20 |
| 33 | 6 | 2 | 12 | 2 | 11 | 11 | 11 | 11 | 11 | 11 | ST398_3_n8 |
| 33 | 27 | 2 | 19 | 2 | 117 | 106 | 94 | 79 | 68 | 63 | ST398_3_t25 |
| 33 | 11 | 2 | 10 | 2 | 24 | 24 | 23 | 23 | 23 | 23 | ST398_3_t14 |
| 33 | 12 | 2 | 10 | 2 | 25 | 25 | 23 | 23 | 23 | 23 | ST398_3_n12 |
| 34 | 19 | 2 | 11 | 2 | 68 | 64 | 59 | 50 | 46 | 43 | ST398_3_t23 |
| 34 | 18 | 2 | 11 | 2 | 61 | 57 | 54 | 50 | 46 | 43 | ST398_3_n18 |
| 34 | 33 | 2 | 11 | 2 | 196 | 167 | 54 | 50 | 46 | 43 | ST398_3_n16 |
| 36 | 22 | 2 | 17 | 2 | 88 | 82 | 76 | 63 | 56 | 52 | ST398_3_t21 |
| 310 | 8 | 2 | 13 | 2 | 3 | 3 | 3 | 3 | 3 | 3 | ST398_3_n21 |
| 14 | 2 | 2 | 3 | 2 | 256 | 127 | 13 | 13 | 13 | 13 | ST398_1_t9 |
| 14 | 2 | 2 | 3 | 2 | 146 | 127 | 13 | 13 | 13 | 13 | ST398_1_n14 |
| 14 | 29 | 2 | 3 | 2 | 120 | 108 | 13 | 13 | 13 | 13 | ST398_1_n15 |
| 14 | 10 | 2 | 3 | 2 | 13 | 13 | 13 | 13 | 13 | 13 | ST398_1_n16 |
| 14 | 28 | 2 | 20 | 2 | 118 | 107 | 95 | 80 | 69 | 64 | ST398_1_t24 |
| 13 | 1 | 2 | 2 | 2 | 54 | 52 | 49 | 46 | 43 | 41 | ST398_1_t11 |
| 13 | 1 | 2 | 2 | 2 | 253 | 52 | 49 | 46 | 43 | 41 | ST398_1_n12 |
| 13 | 39 | 2 | 2 | 2 | 242 | 202 | 49 | 46 | 43 | 41 | ST398_1_n13 |
| 19 | 35 | 2 | 23 | 2 | 205 | 174 | 143 | 121 | 98 | 86 | ST398_1_n22 |
| 16 | 17 | 2 | 15 | 2 | 48 | 46 | 43 | 41 | 38 | 37 | ST398_1_t7 |
| 12 | 25 | 2 | 4 | 2 | 103 | 93 | 84 | 35 | 33 | 32 | ST398_1_t6 |
| 12 | 37 | 2 | 4 | 2 | 237 | 198 | 162 | 35 | 33 | 32 | ST398_1_t5 |
| 12 | 41 | 2 | 4 | 2 | 254 | 211 | 172 | 35 | 33 | 32 | ST398_1_n9 |
| 12 | 15 | 2 | 4 | 2 | 40 | 39 | 36 | 35 | 33 | 32 | ST398_1_n4 |
| 25 | 16 | 4 | 8 | 2 | 41 | 40 | 37 | 36 | 34 | 33 | ST398_2_t7 |
| 25 | 36 | 4 | 8 | 2 | 207 | 176 | 145 | 123 | 34 | 33 | ST398_2_n21 |
| 25 | 34 | 4 | 22 | 2 | 201 | 172 | 142 | 120 | 97 | 85 | ST398_2_t21 |
| 11 | 26 | 1 | 1 | 1 | 105 | 95 | 86 | 71 | 62 | 57 | ST398_1_t23 |
| 11 | 38 | 1 | 1 | 1 | 239 | 200 | 164 | 135 | 107 | 57 | ST398_1_n1 |
