## Supplementary figures and images for "THRESHER: A tool for evidence-based, phylogeny-corrected SNP thresholds for whole genome molecular epidemiology"

### ST1_40_iter_8.pdf

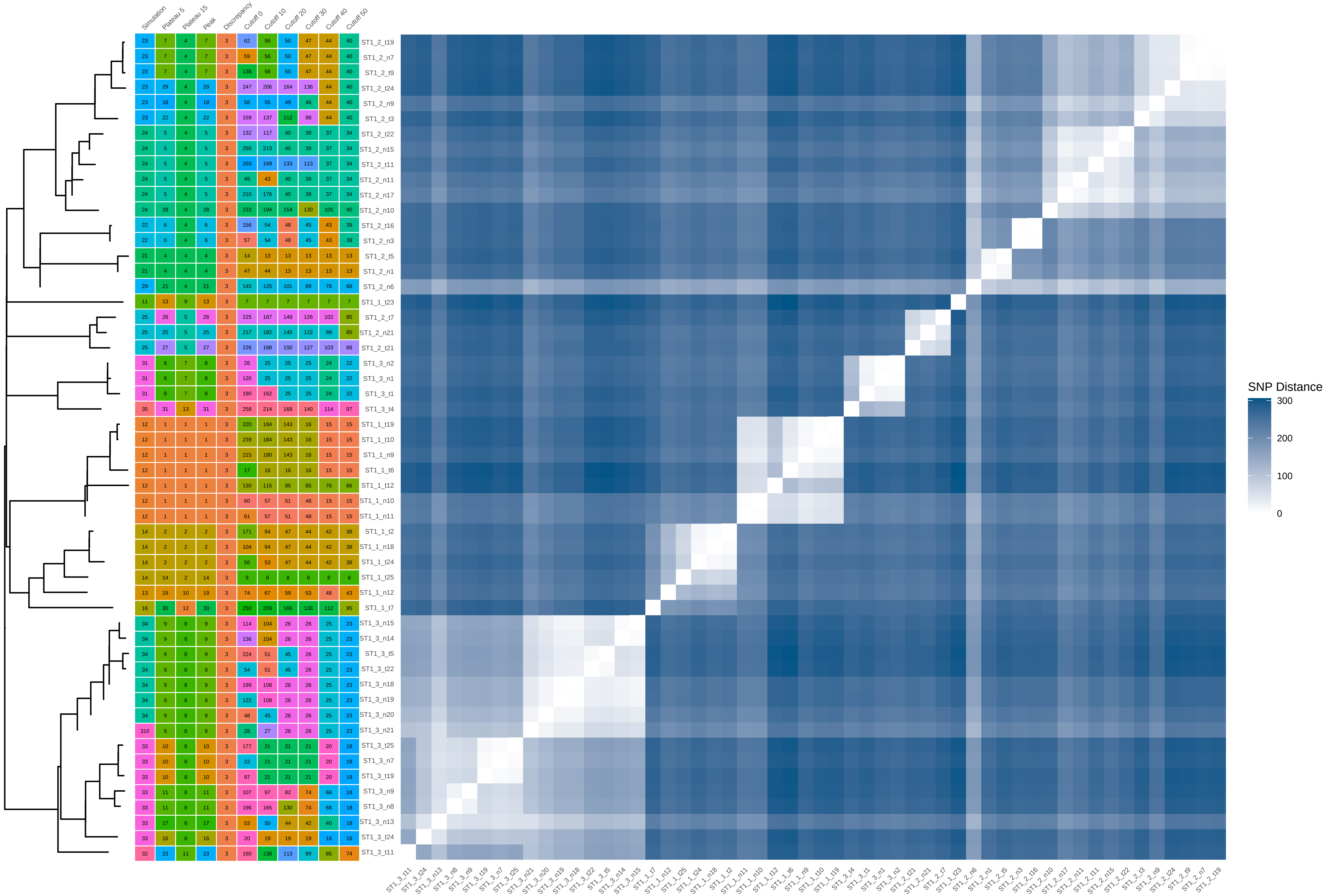

### ST1_40_iter_9.pdf

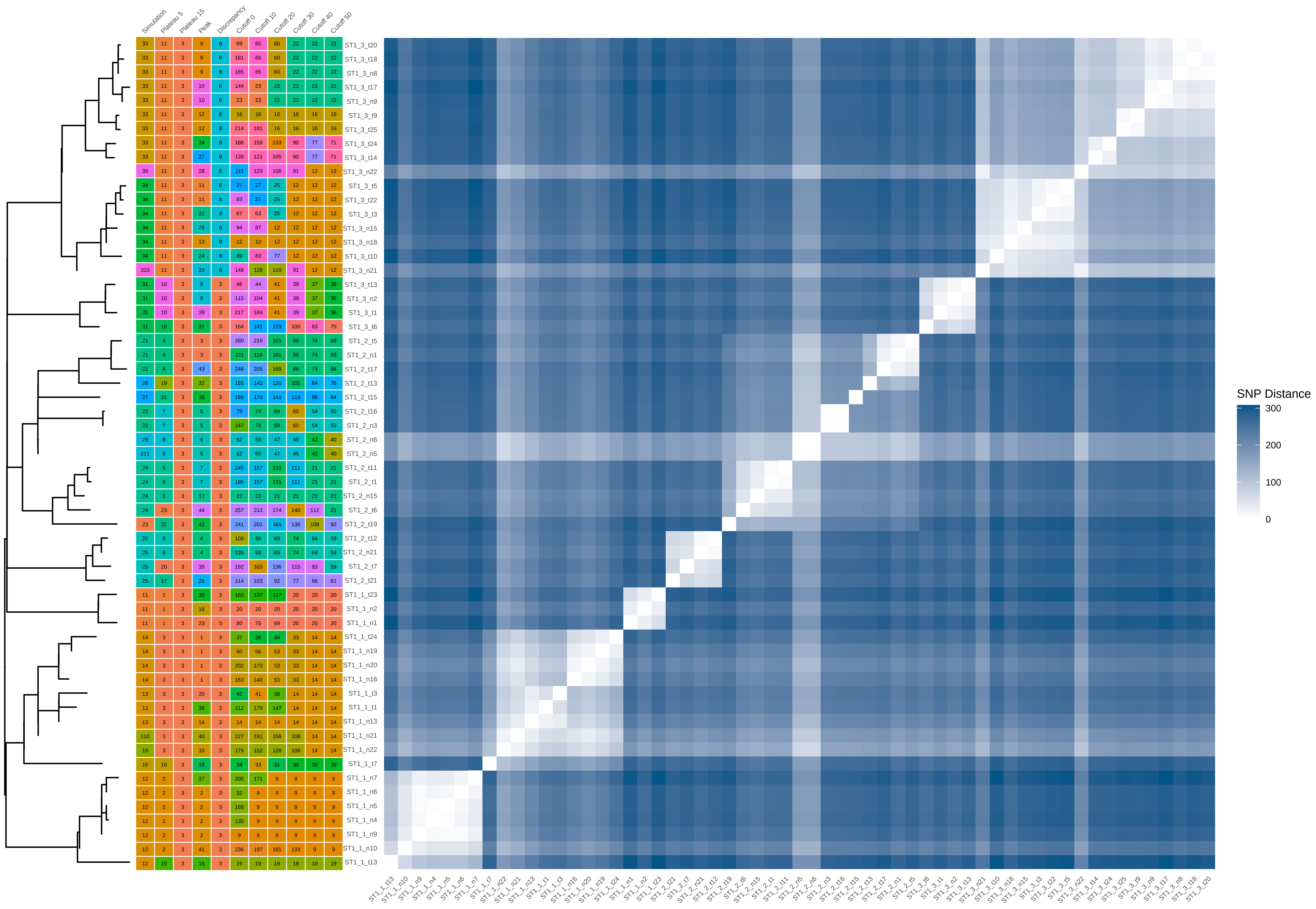

### ST1_60_iter_1.pdf

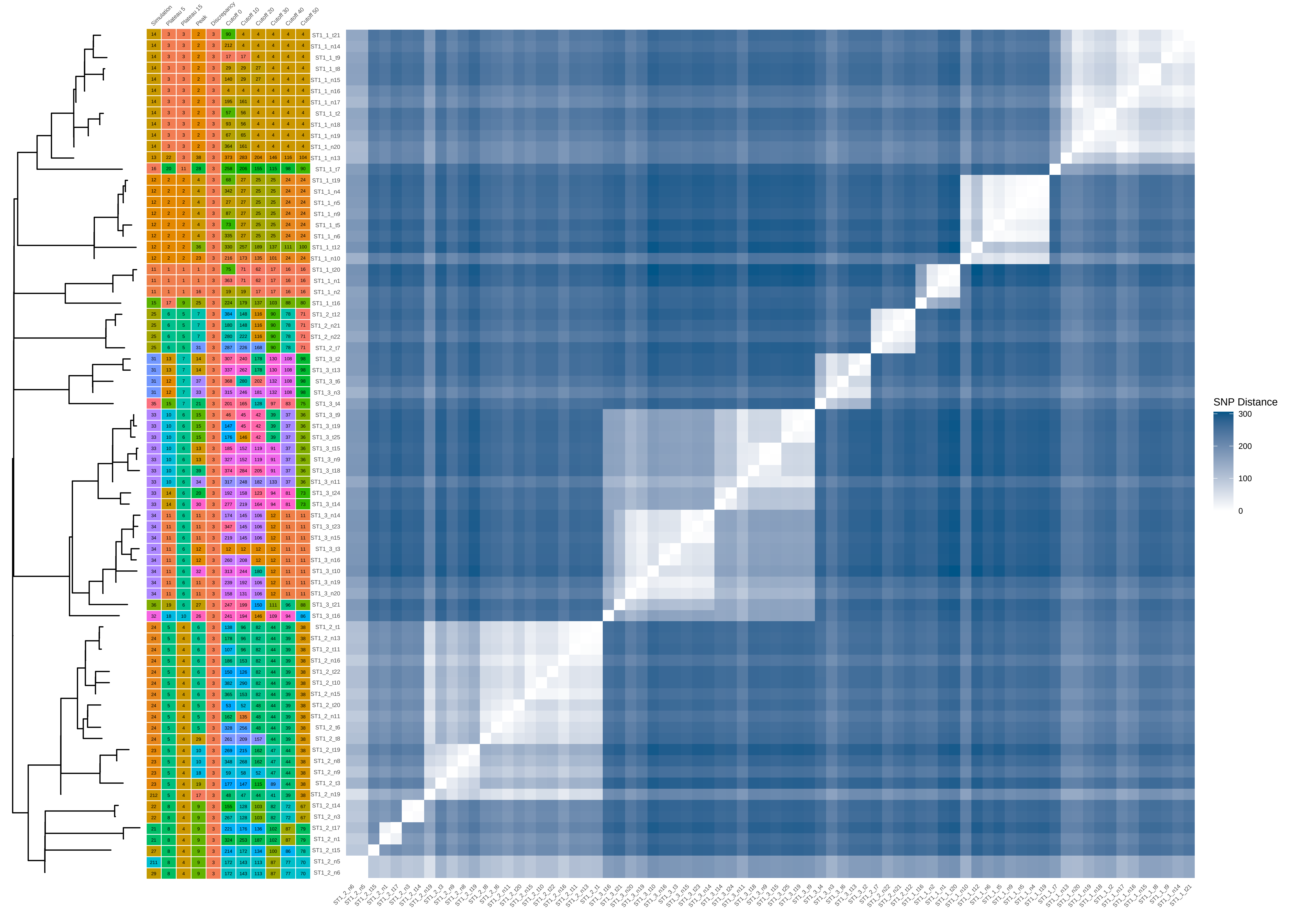

### ST1_60_iter_2.pdf

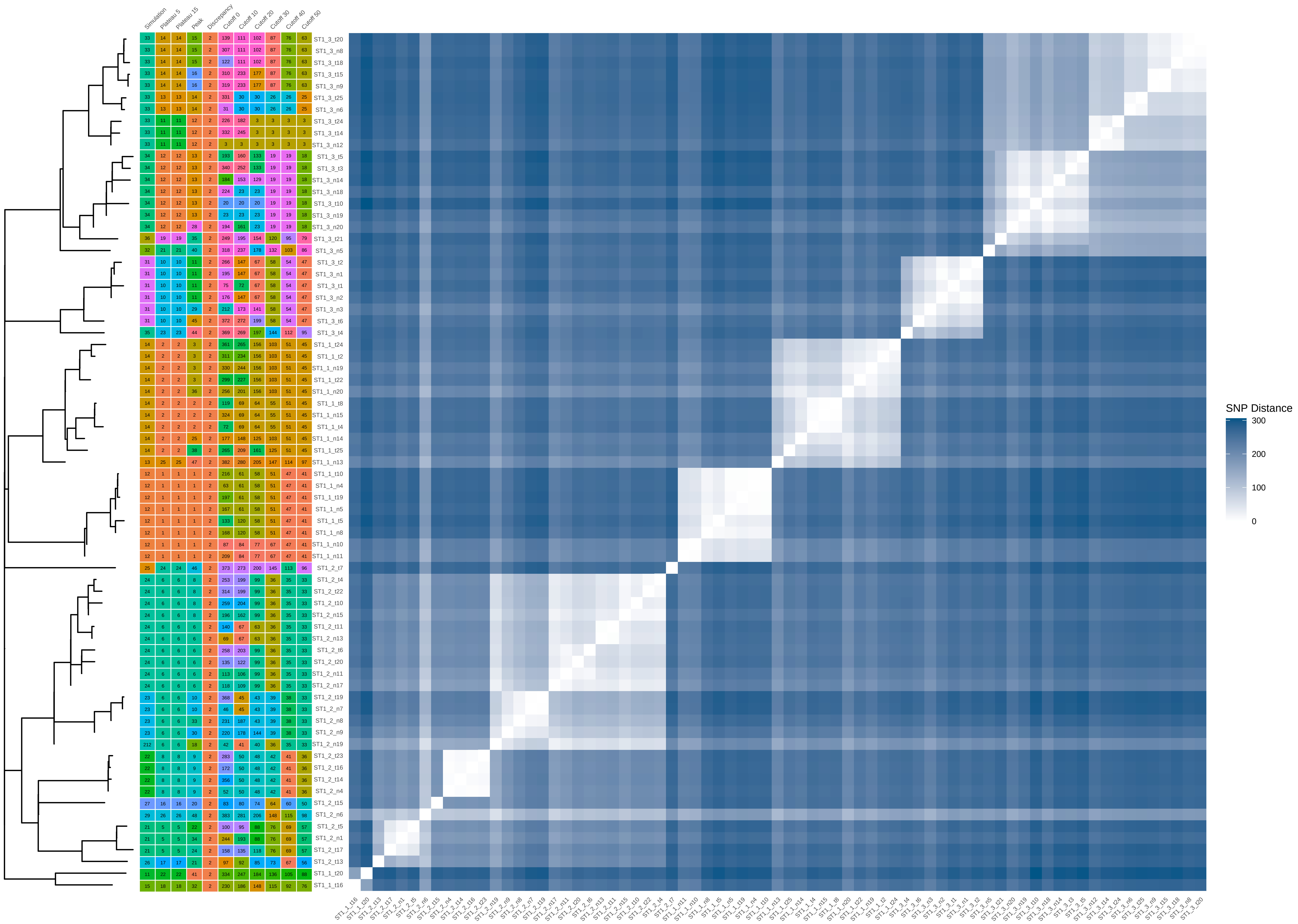

### ST1_60_iter_3.pdf

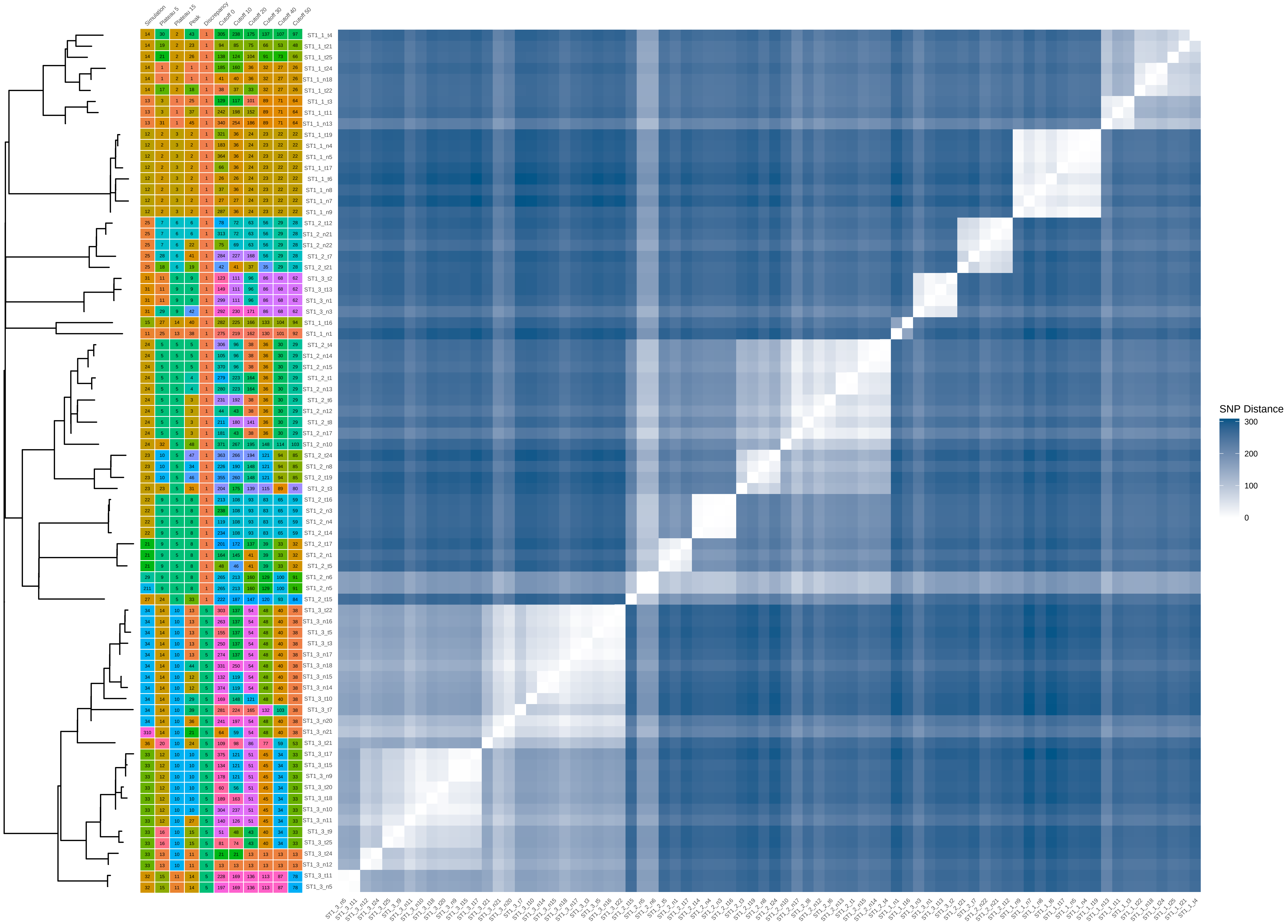

### ST1_60_iter_5.pdf

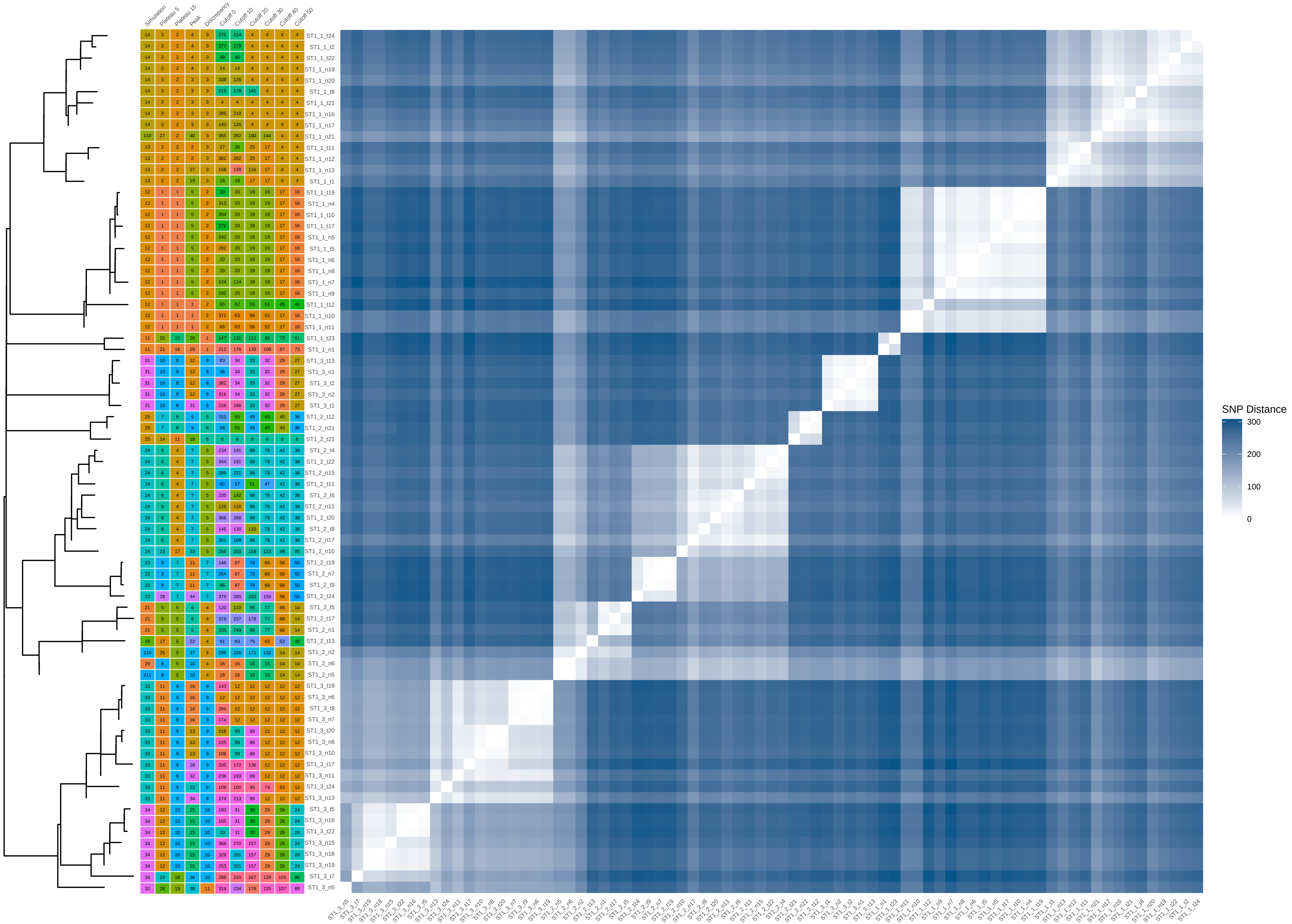

### ST1_60_iter_6.pdf

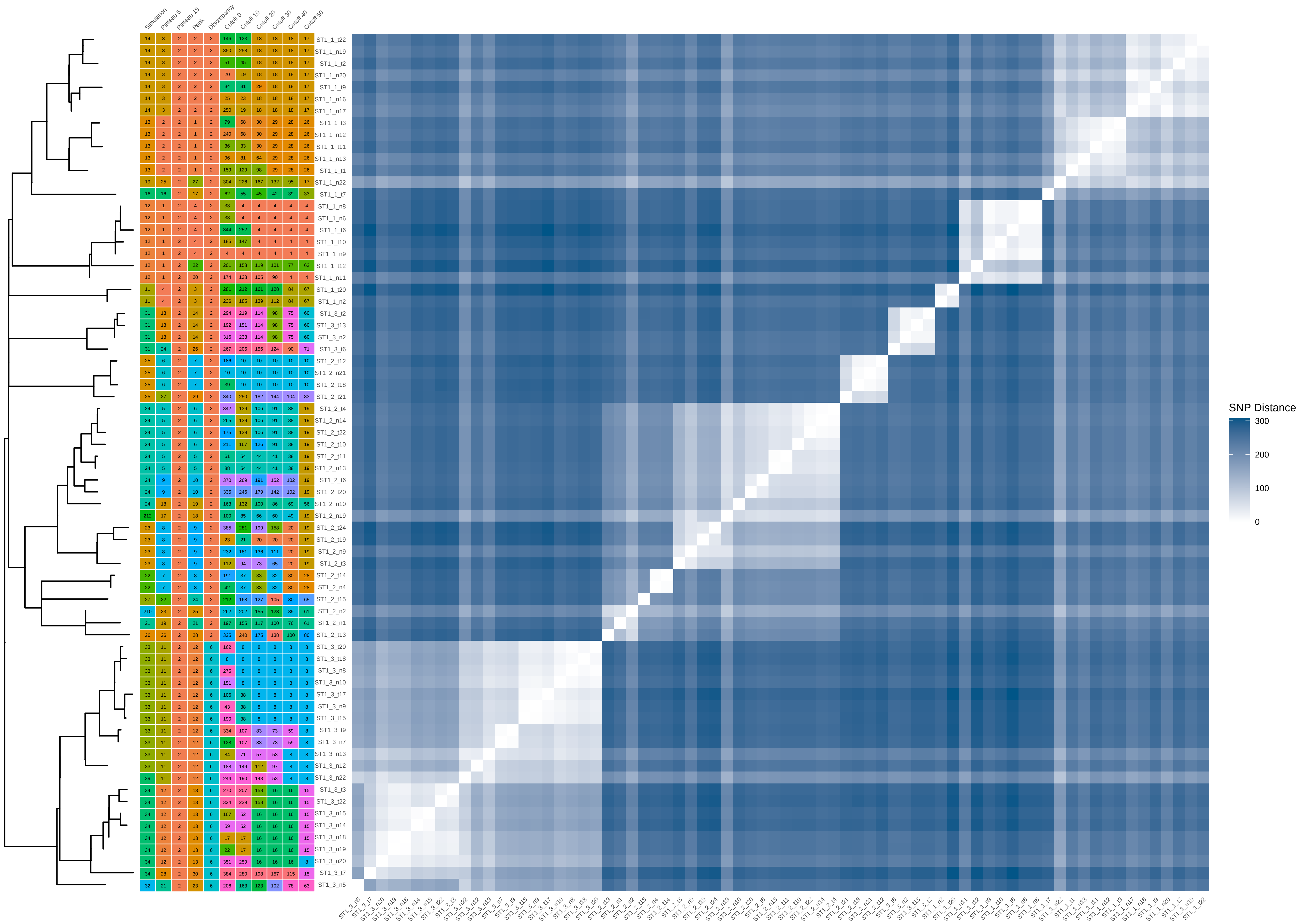

### ST1_60_iter_7.pdf

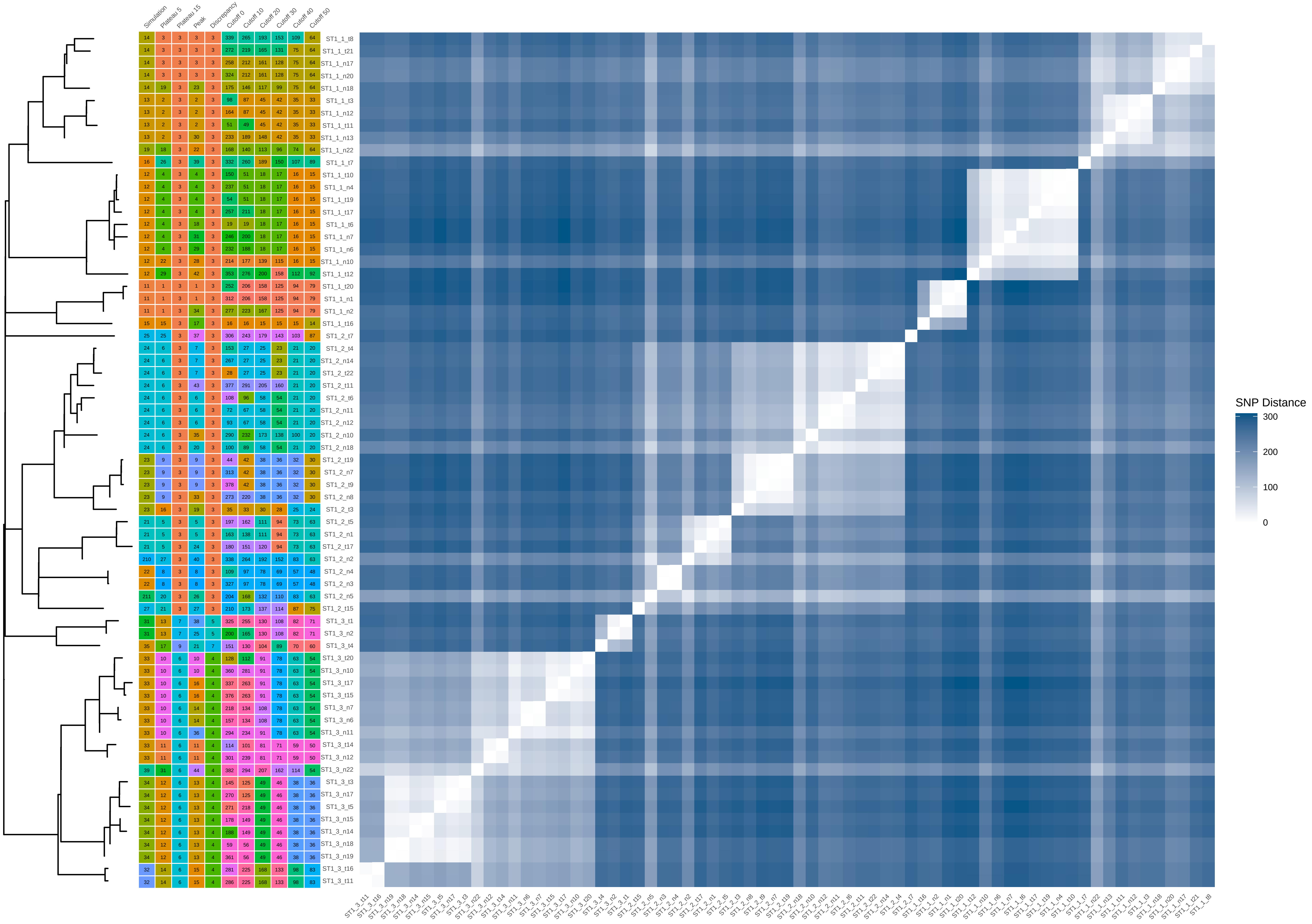

### ST1_80_iter_2.pdf

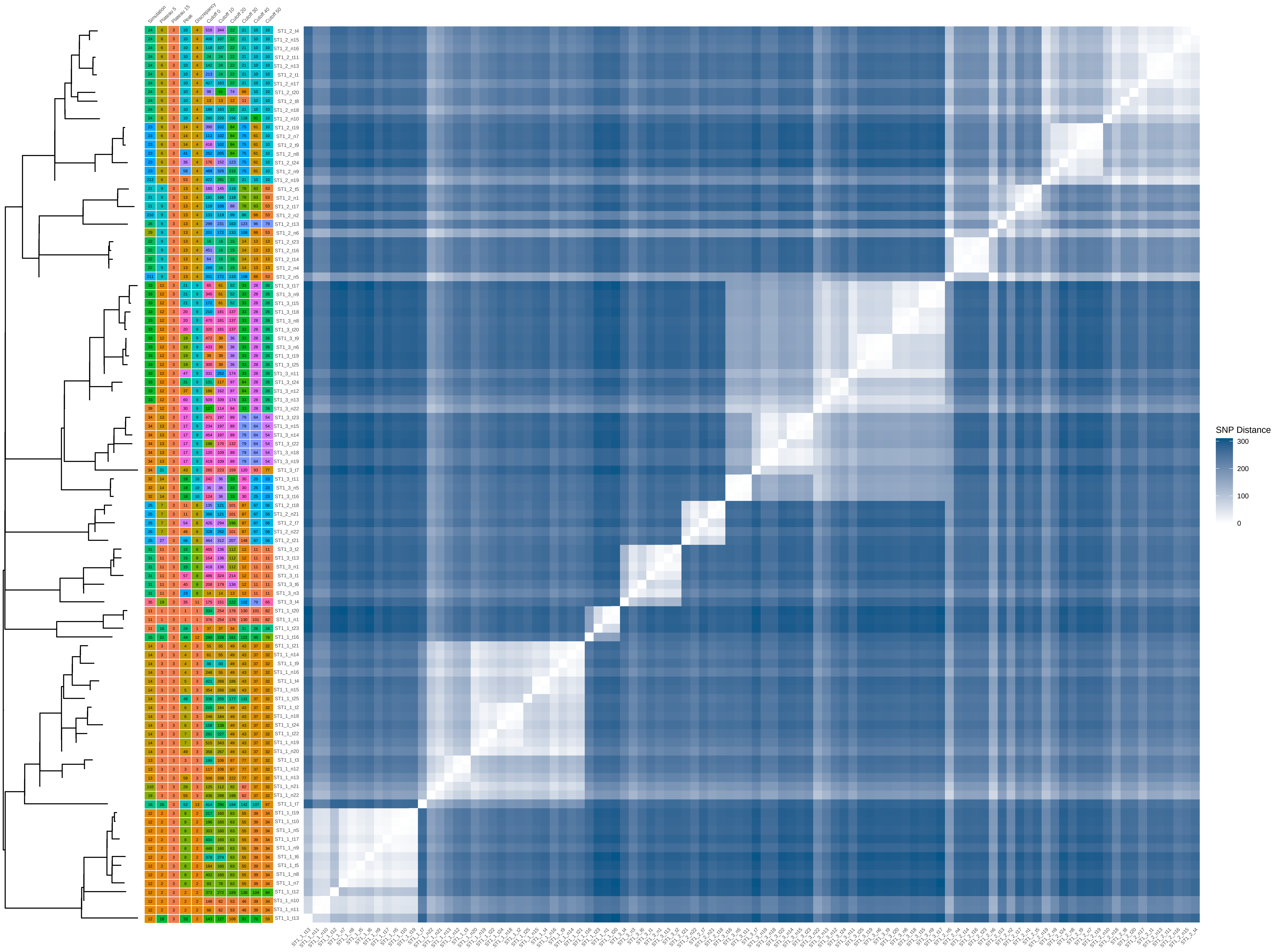

### ST1_80_iter_8.pdf

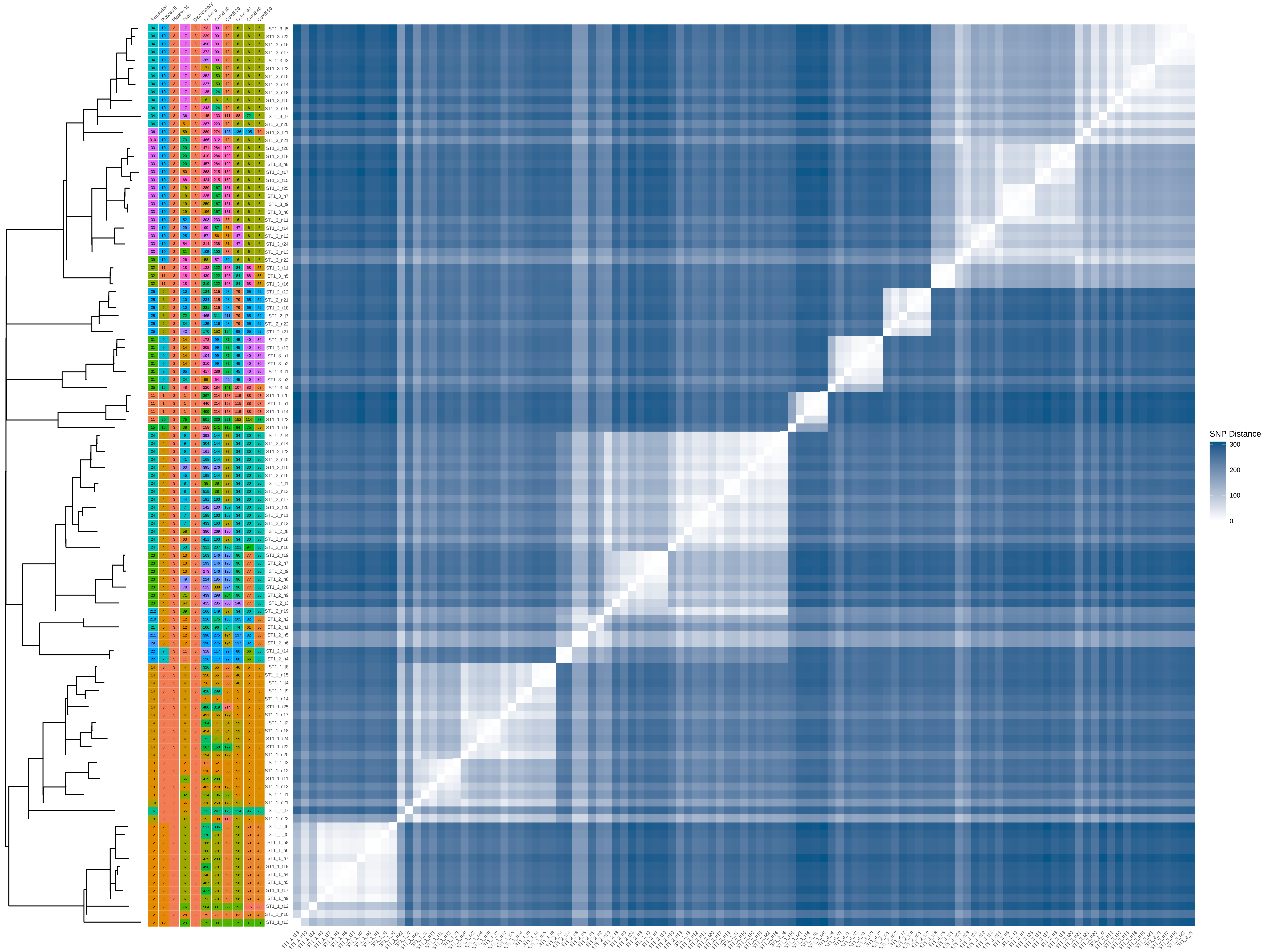

### ST1_80_iter_9.pdf

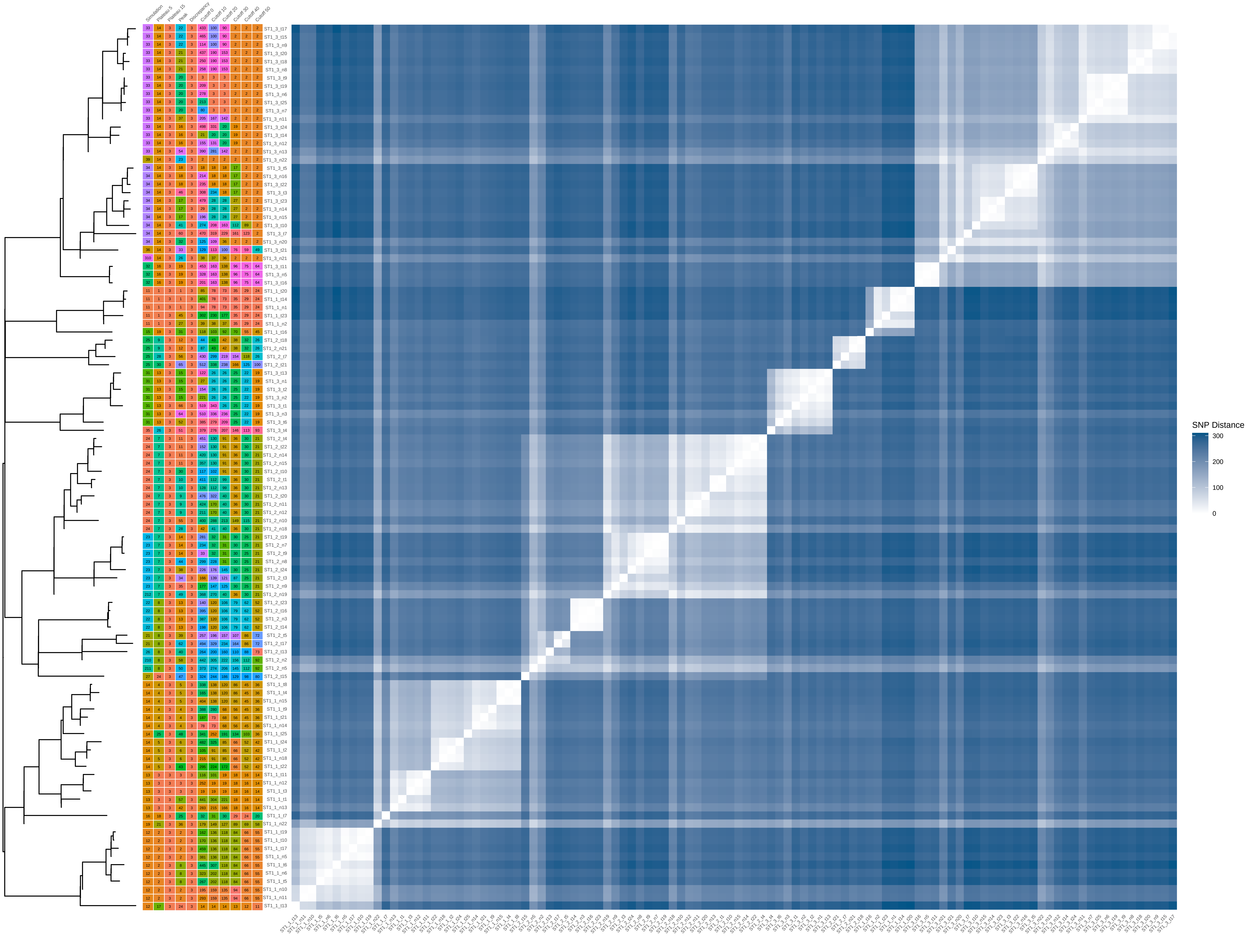

### ST1_80_iter_10.pdf

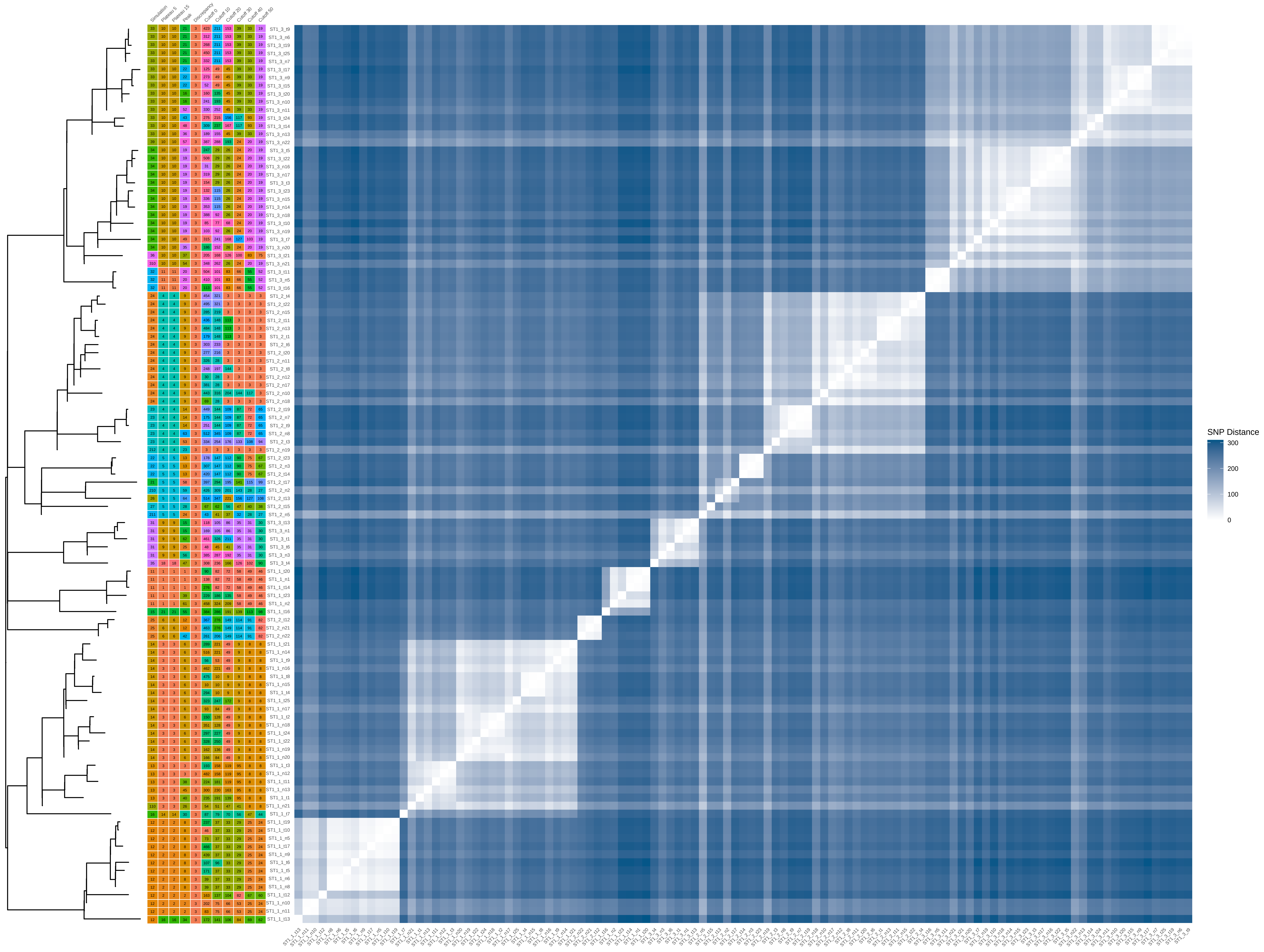

### ST1_100_iter_2.pdf

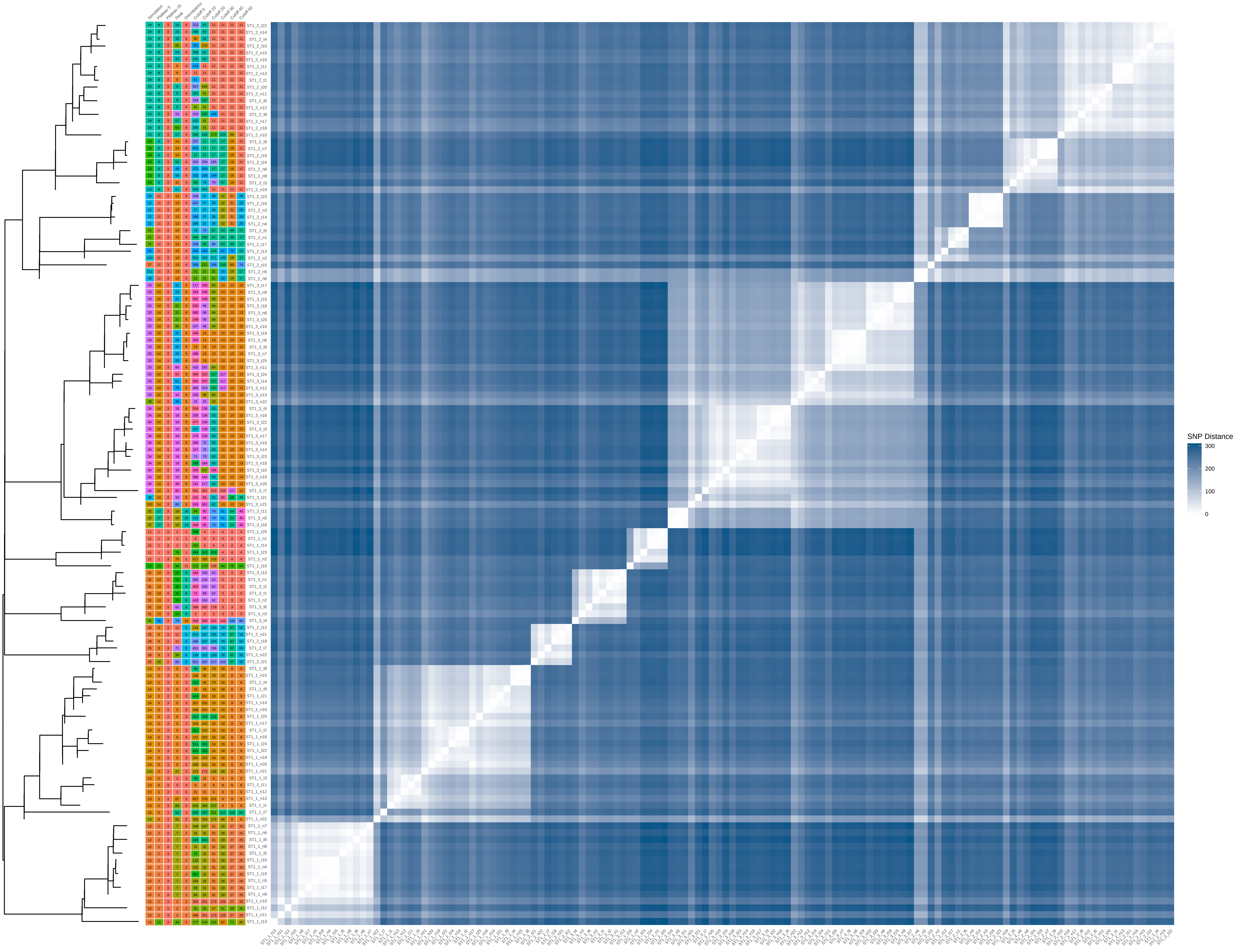

### ST1_100_iter_3.pdf

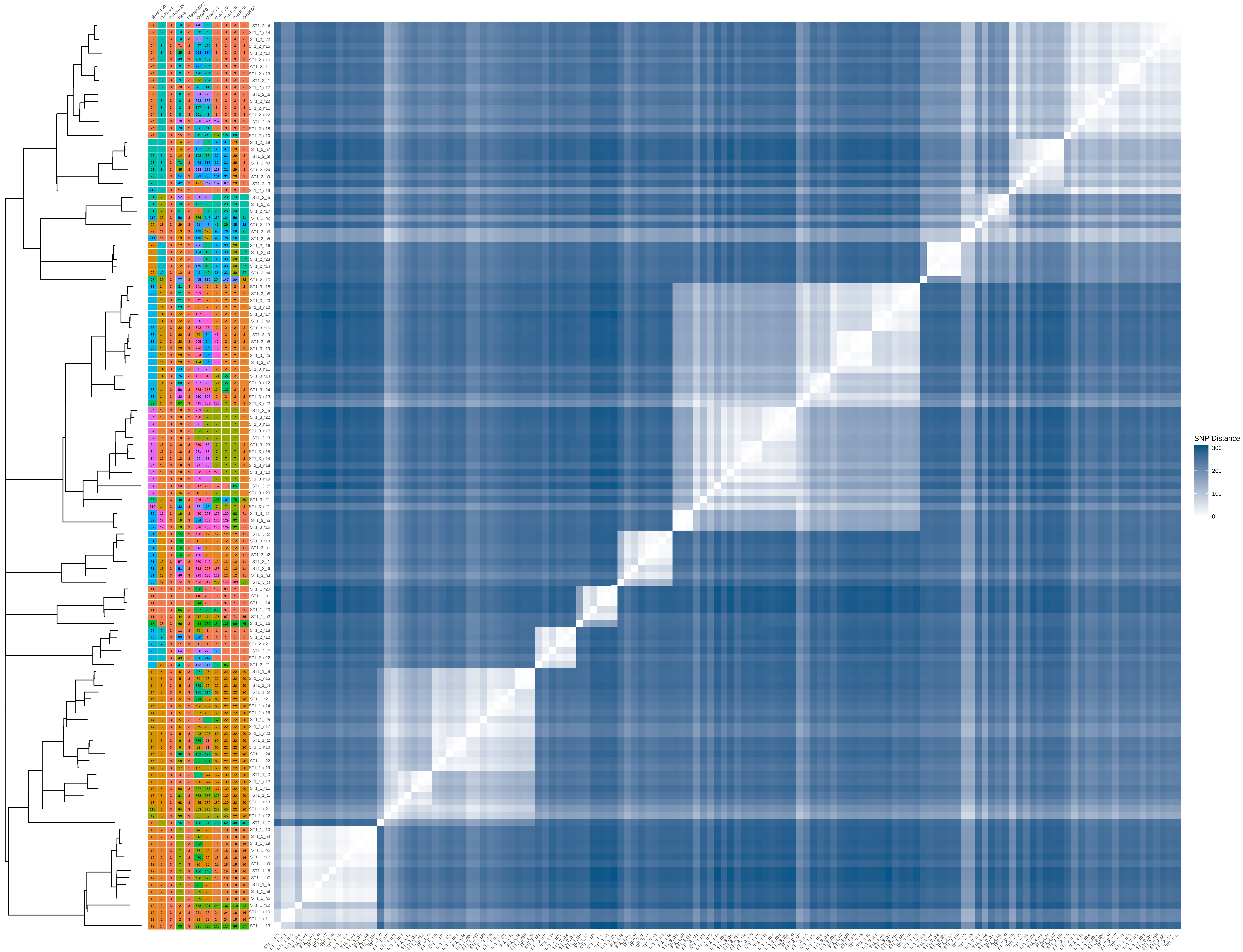

### ST1_100_iter_4.pdf

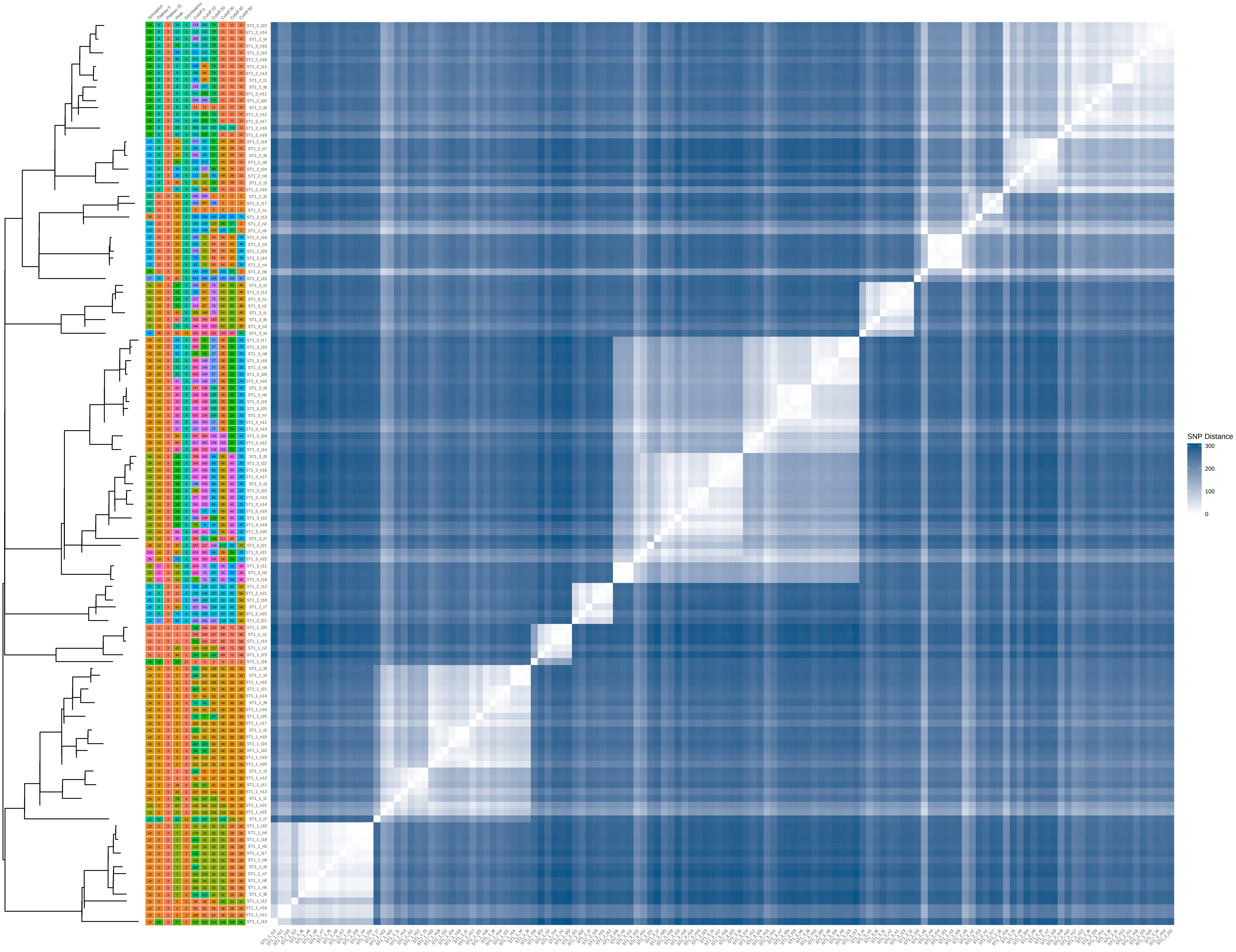

### ST1_100_iter_5.pdf

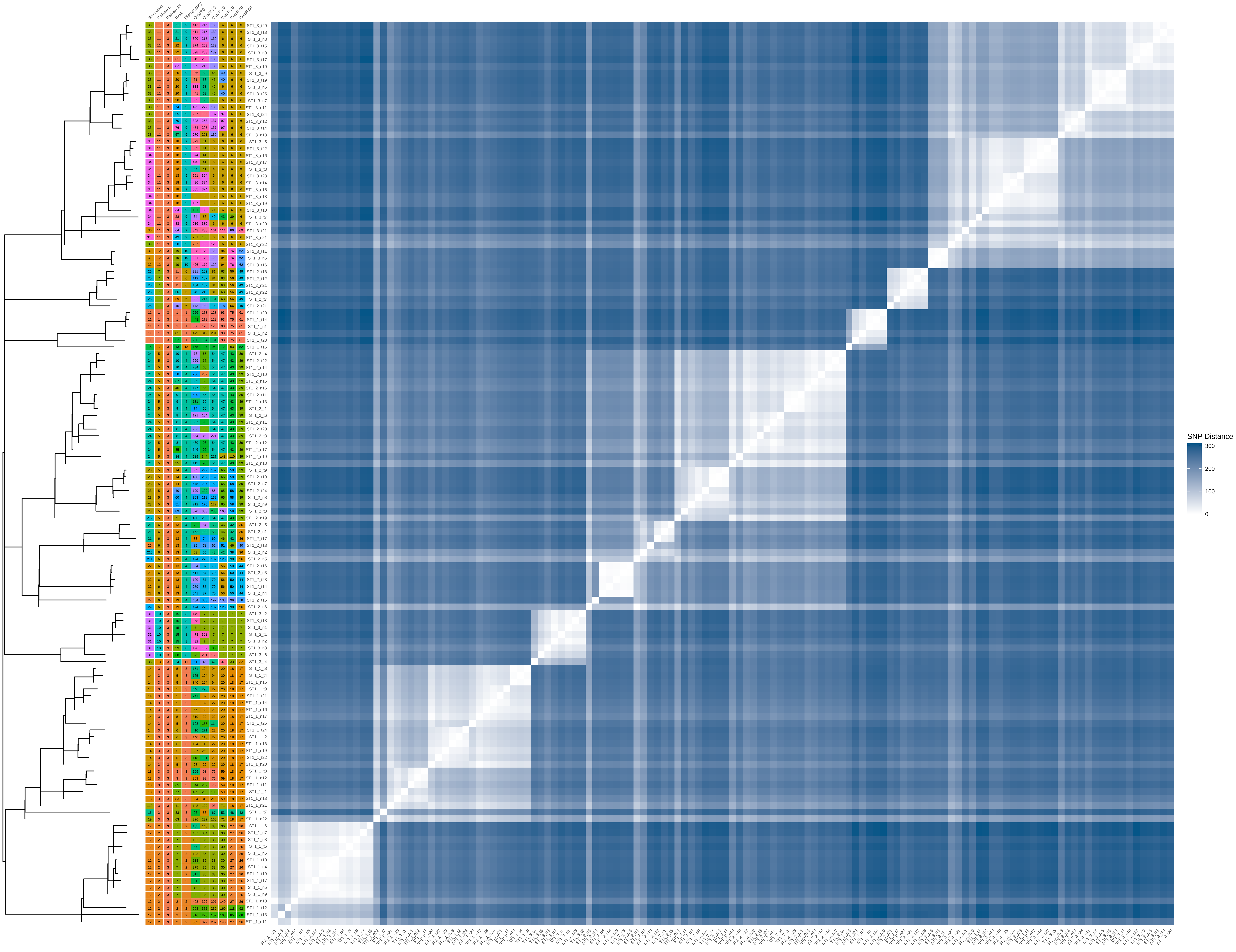

### ST1_100_iter_6.pdf

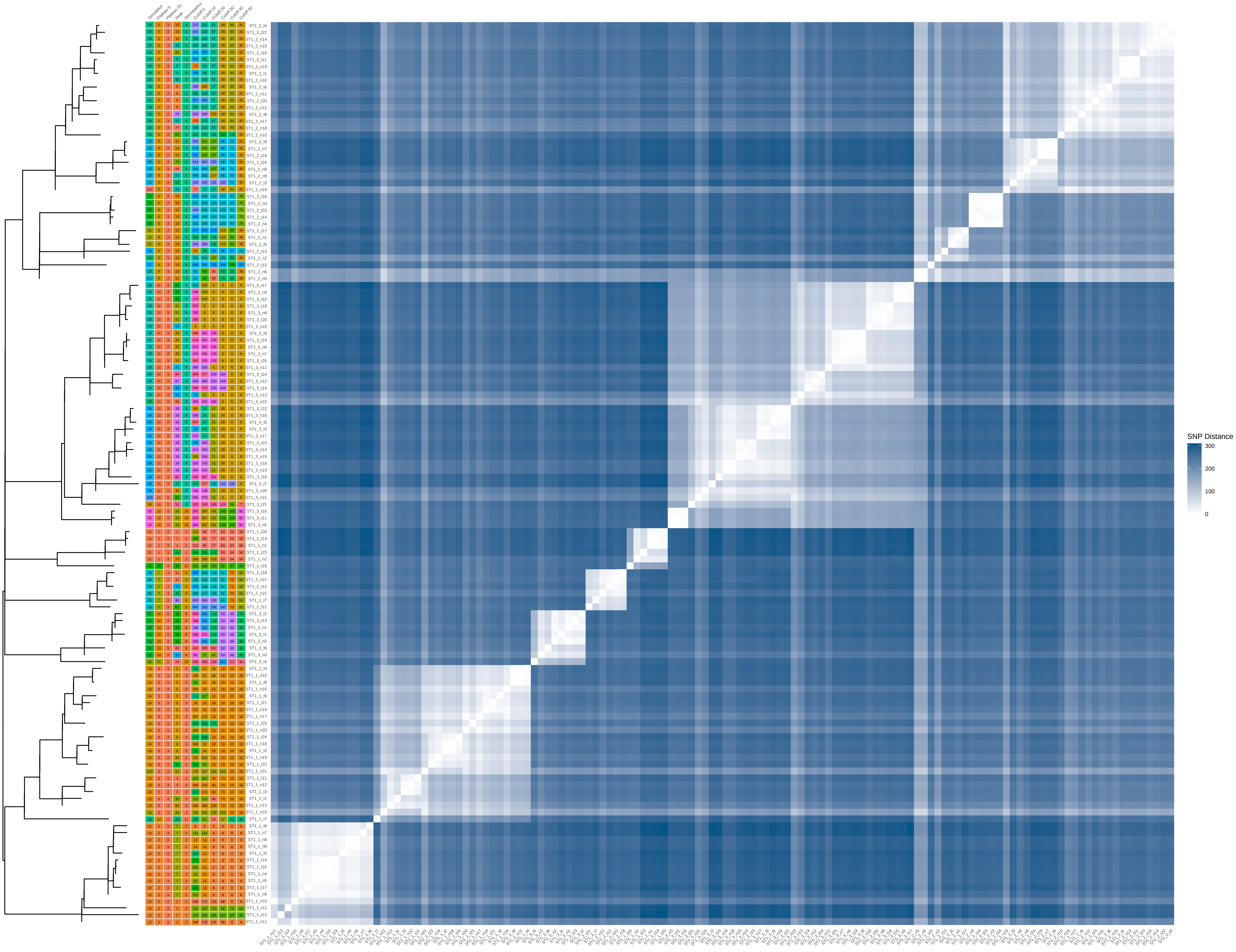

### ST1_100_iter_7.pdf

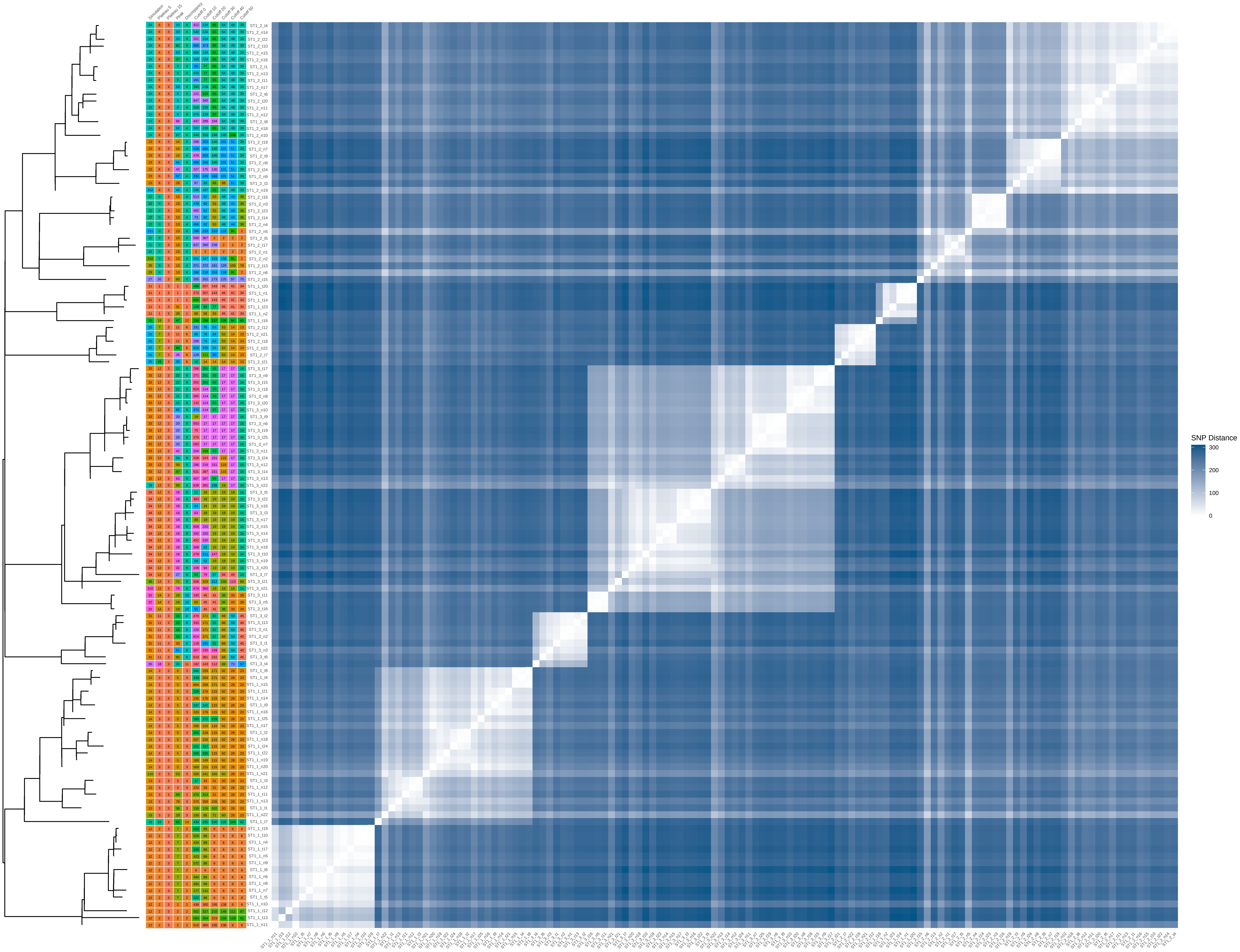

### ST1_100_iter_8.pdf

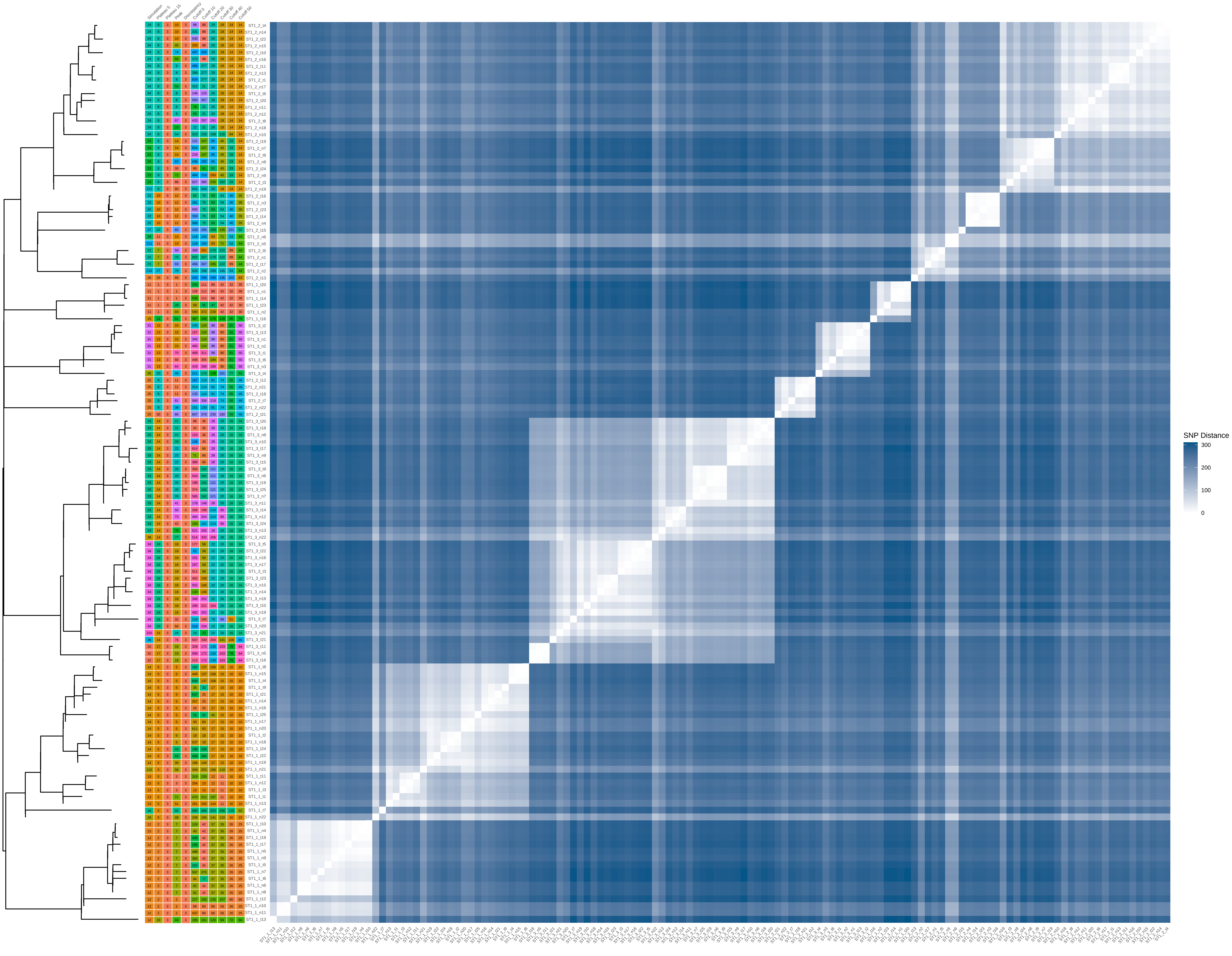

### ST1_100_iter_10.pdf

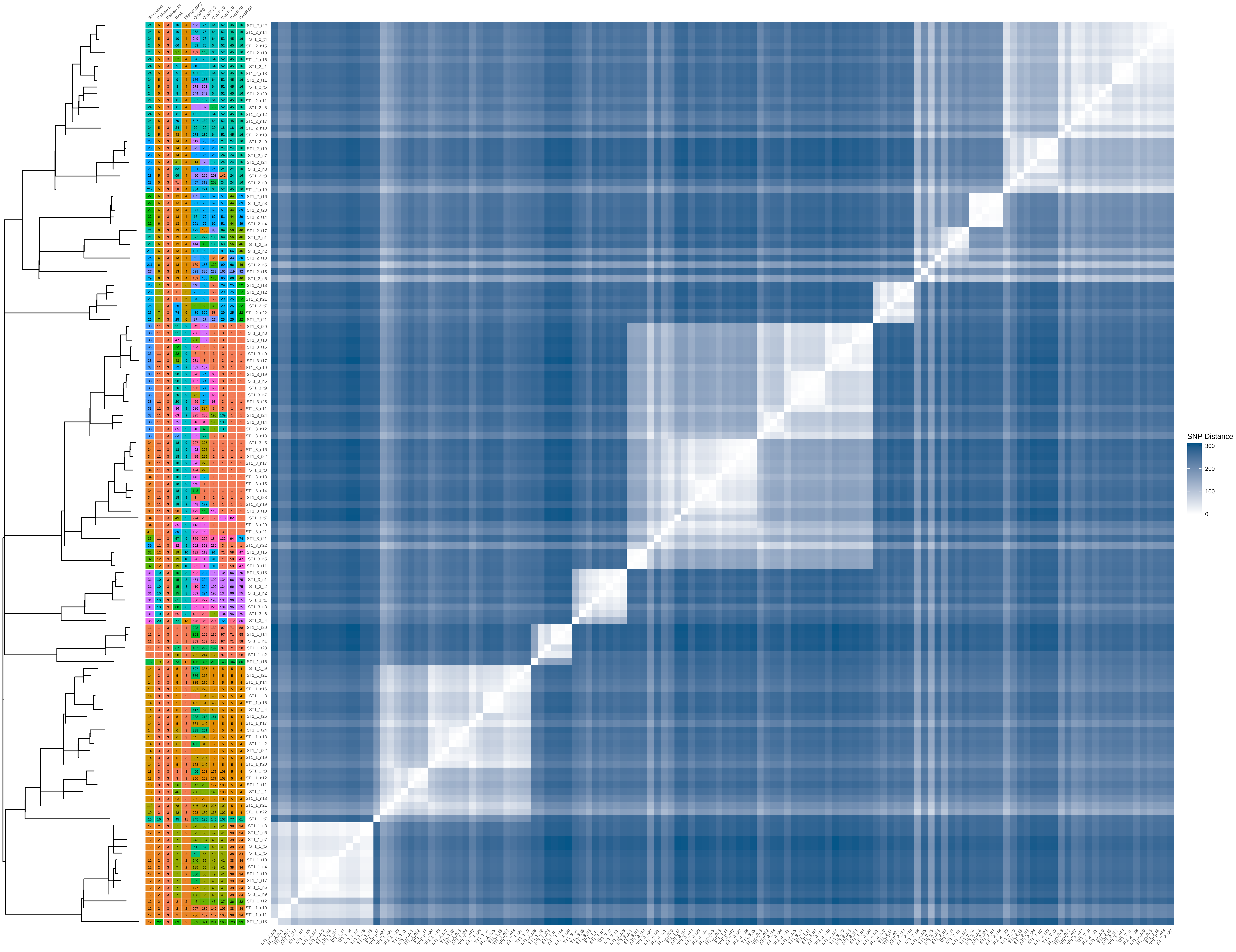

### ST5_20_iter_10.pdf

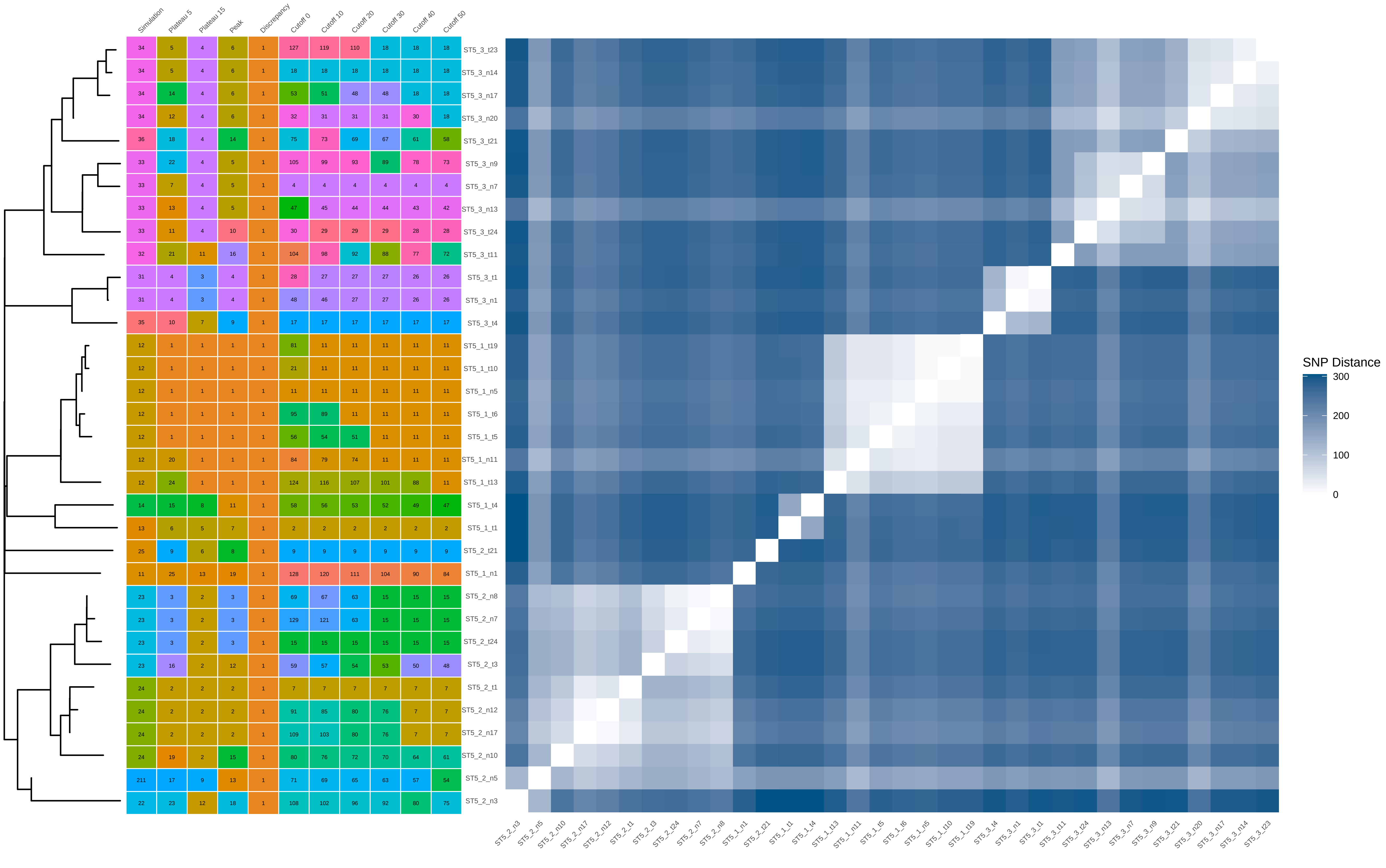

### ST5_40_iter_5.pdf

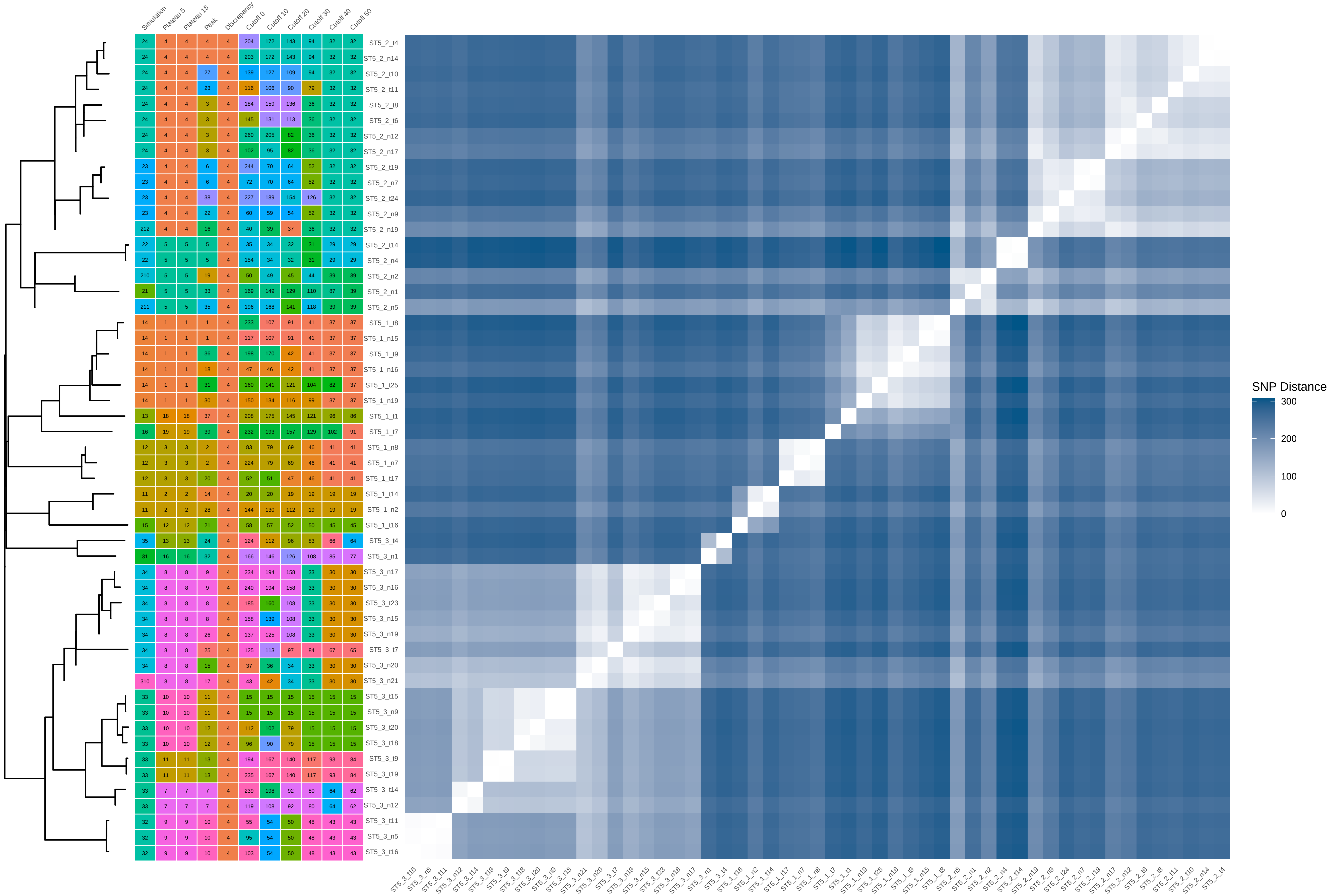

### ST5_40_iter_8.pdf

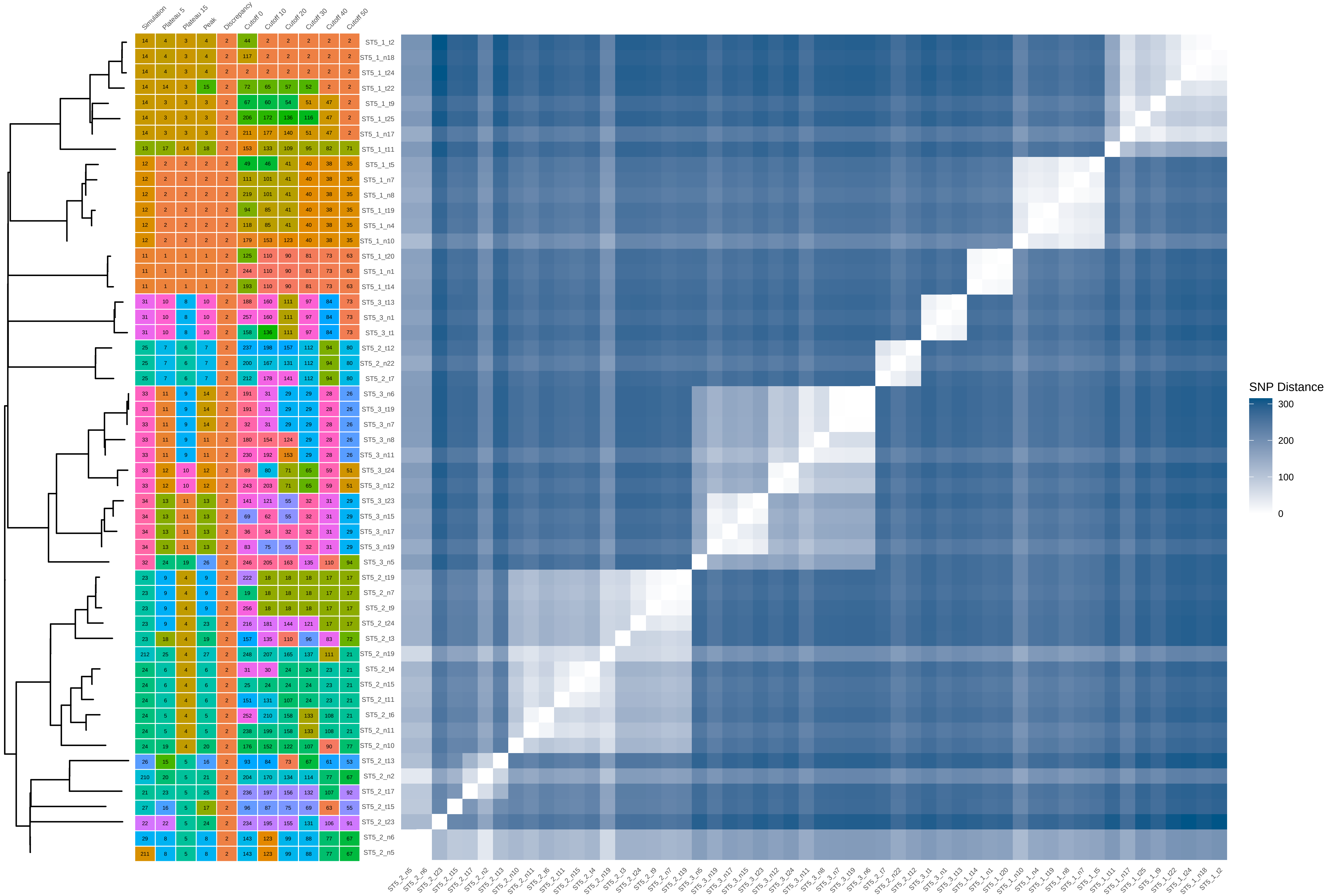

### ST5_40_iter_9.pdf

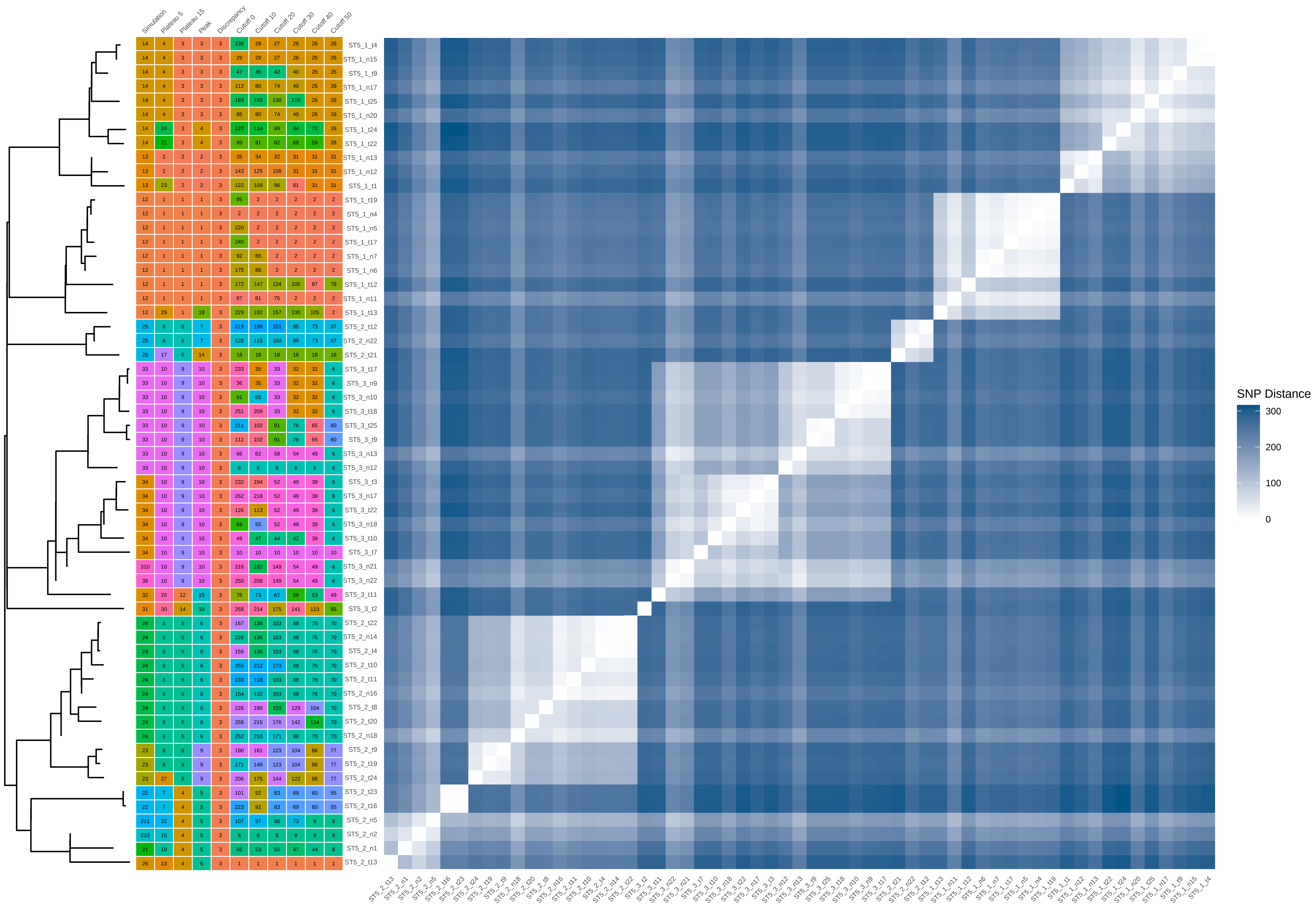

### ST5_40_iter_10.pdf

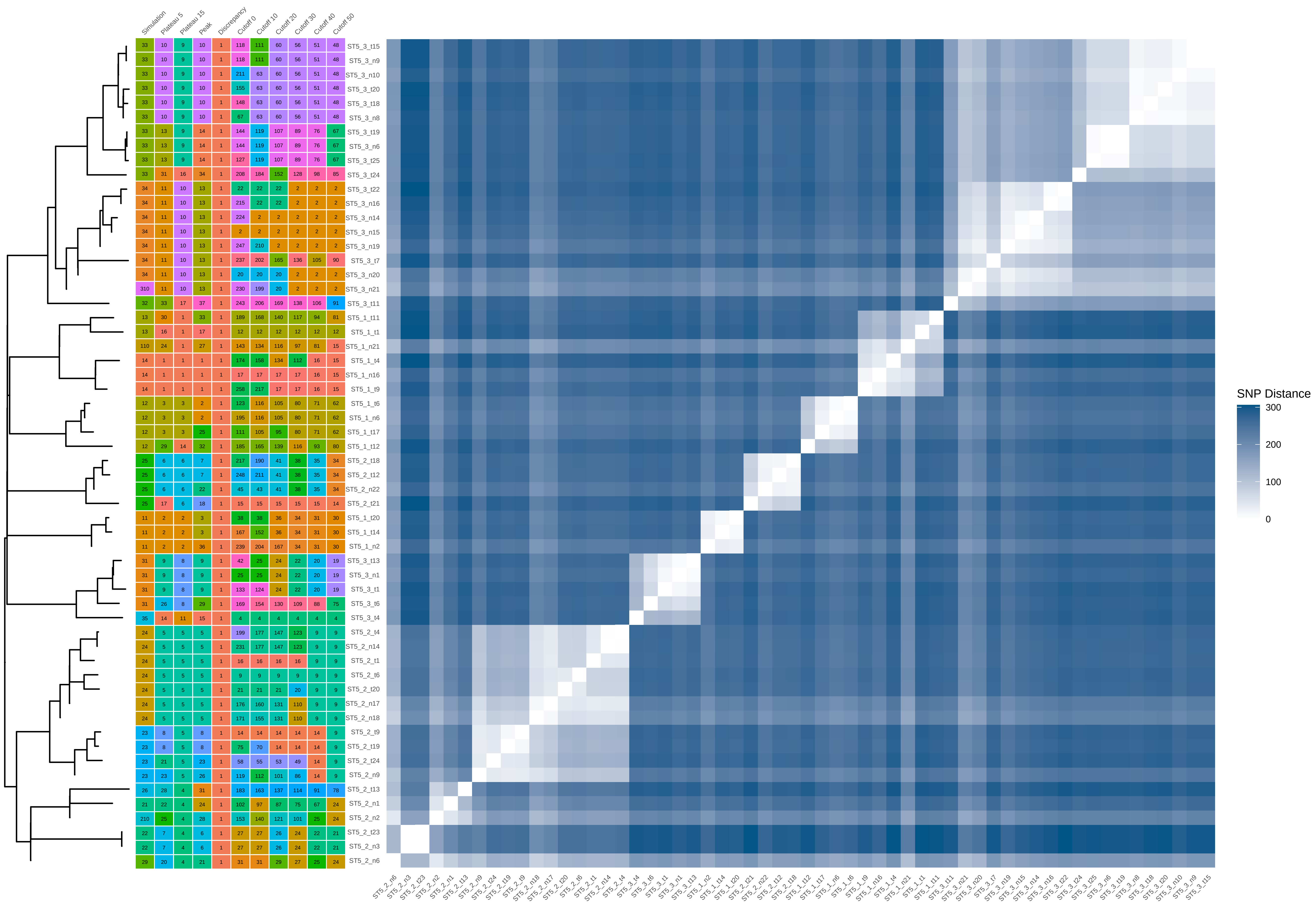

### ST5_60_iter_1.pdf

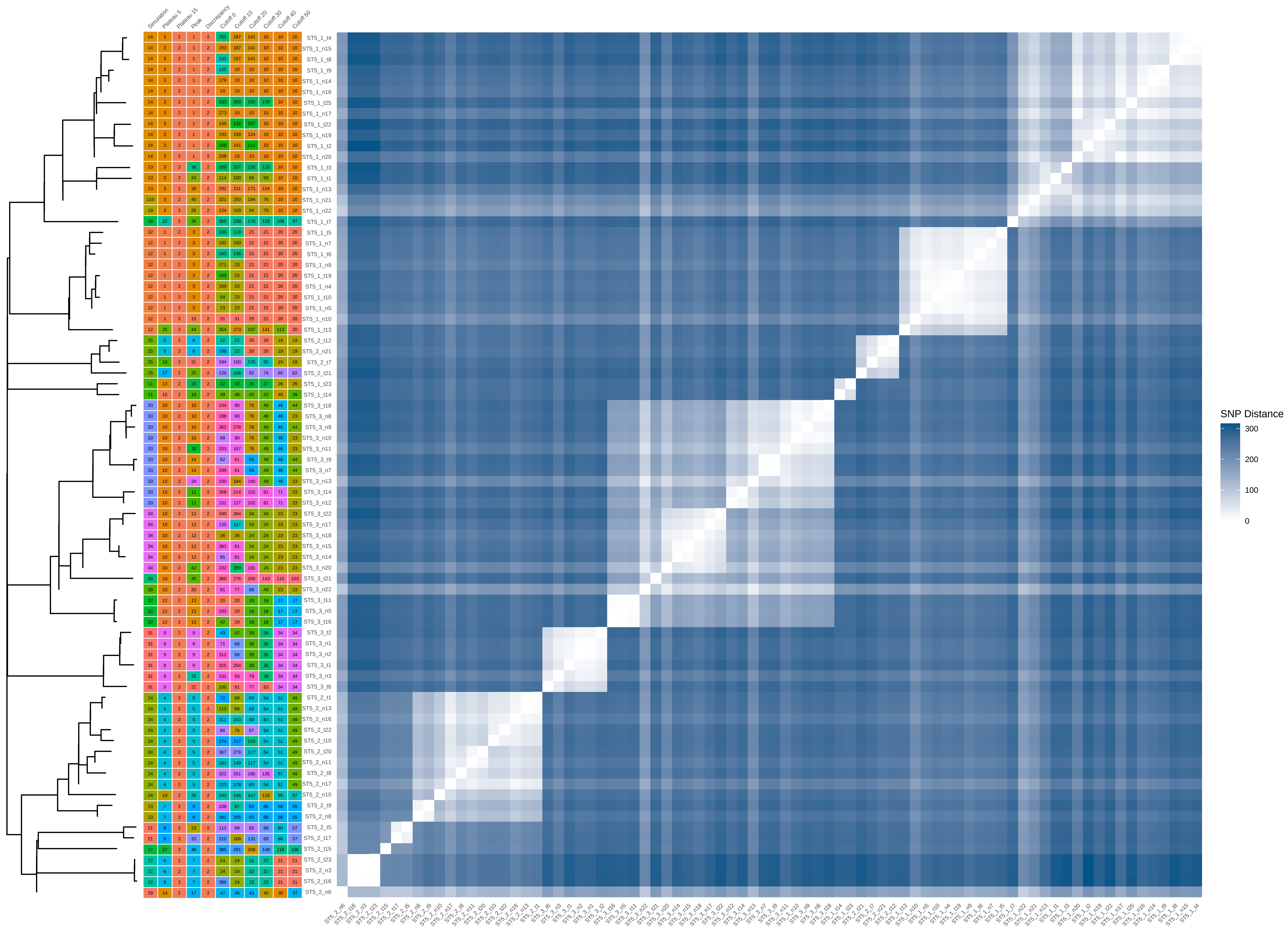

### ST5_60_iter_2.pdf

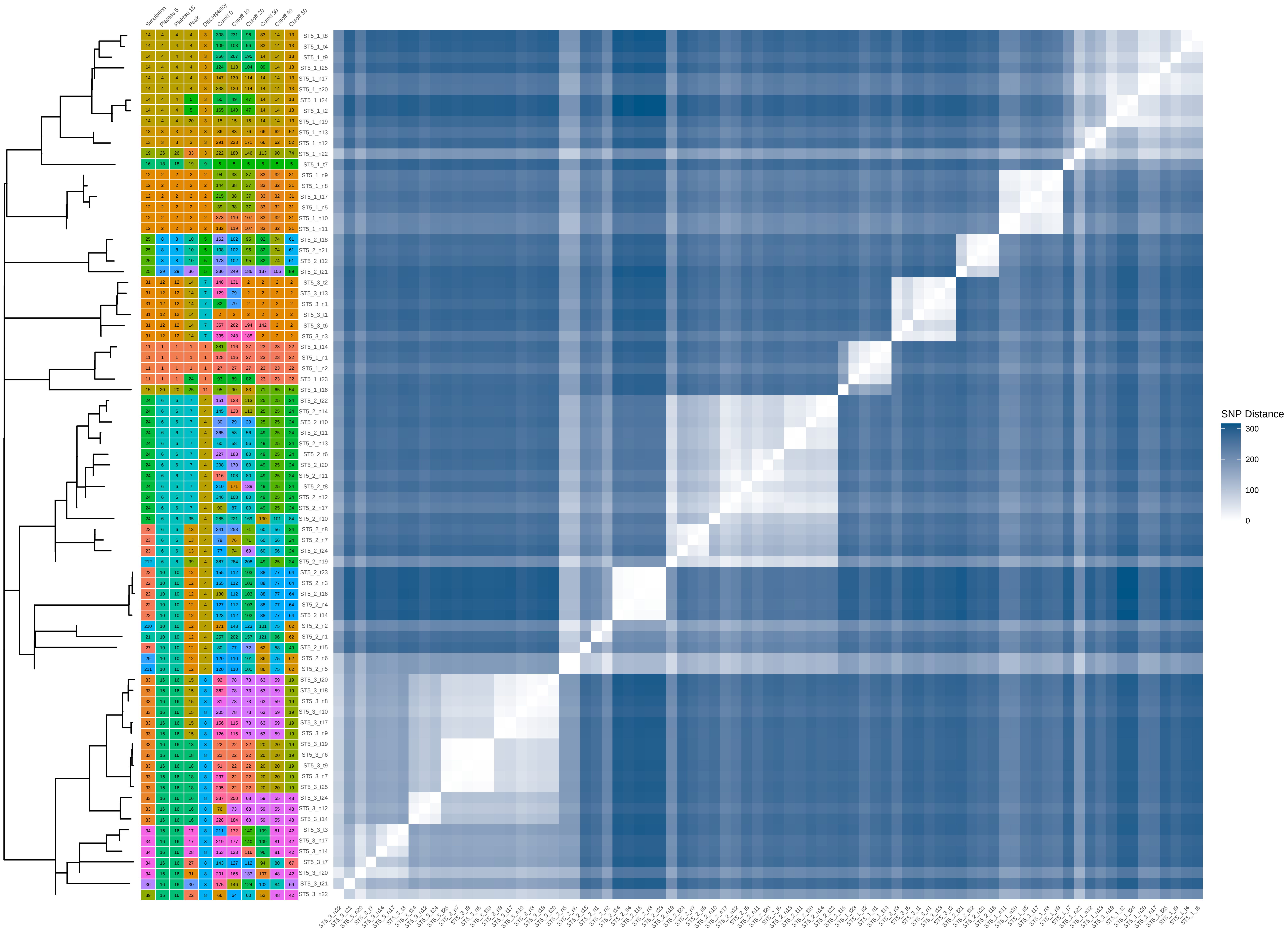

### ST5_60_iter_3.pdf

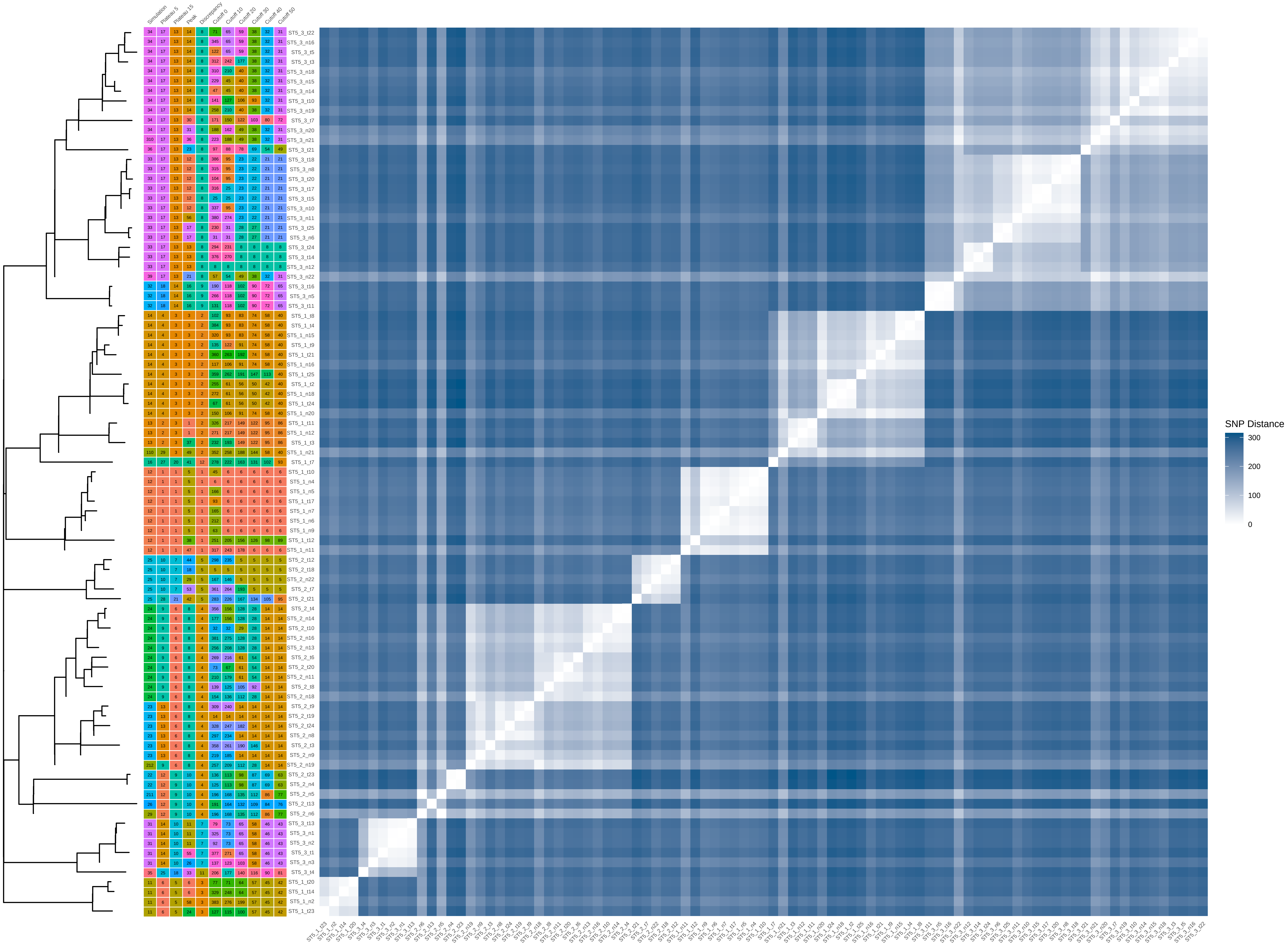

### ST5_60_iter_4.pdf

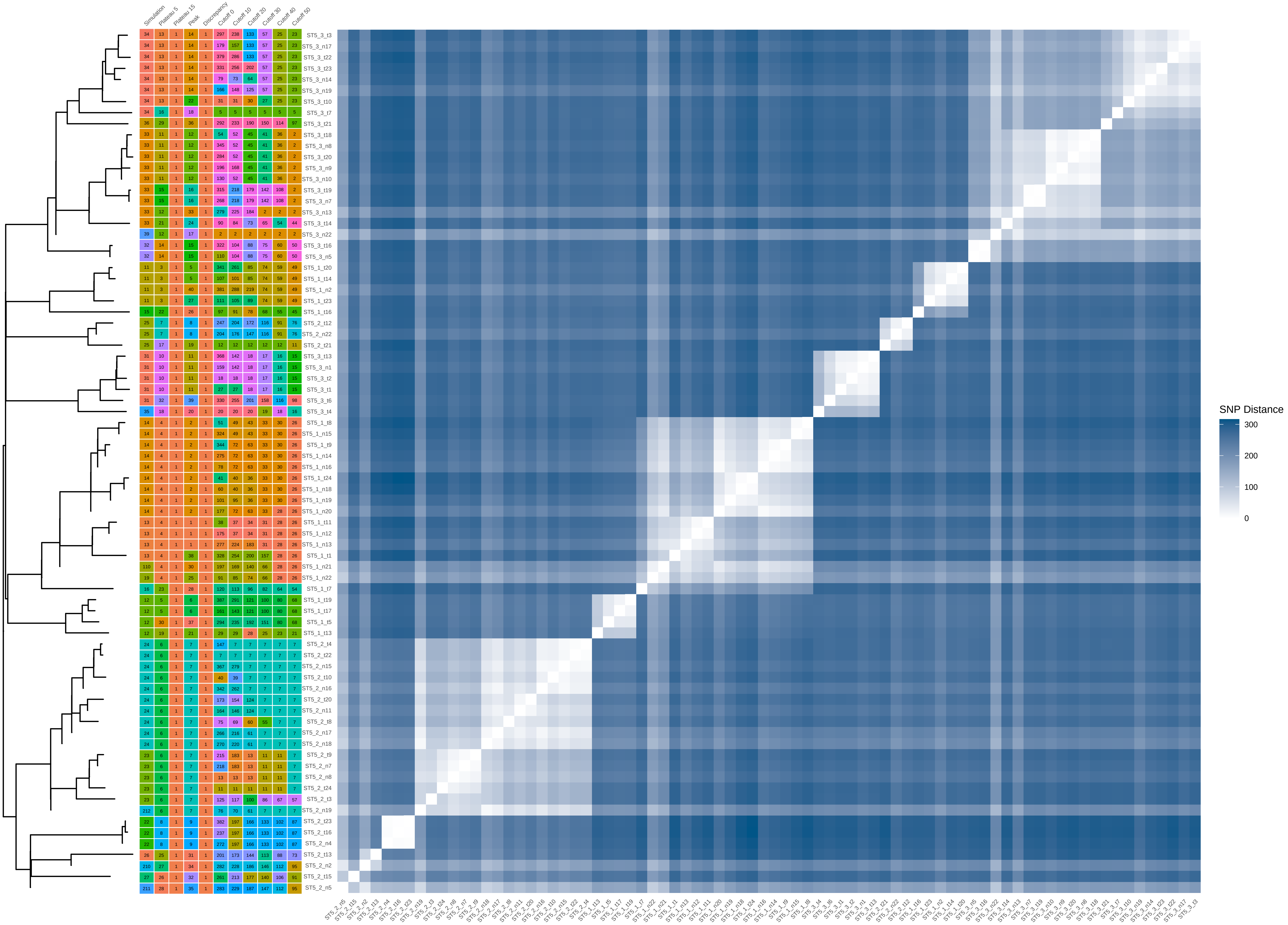

### ST5_60_iter_6.pdf

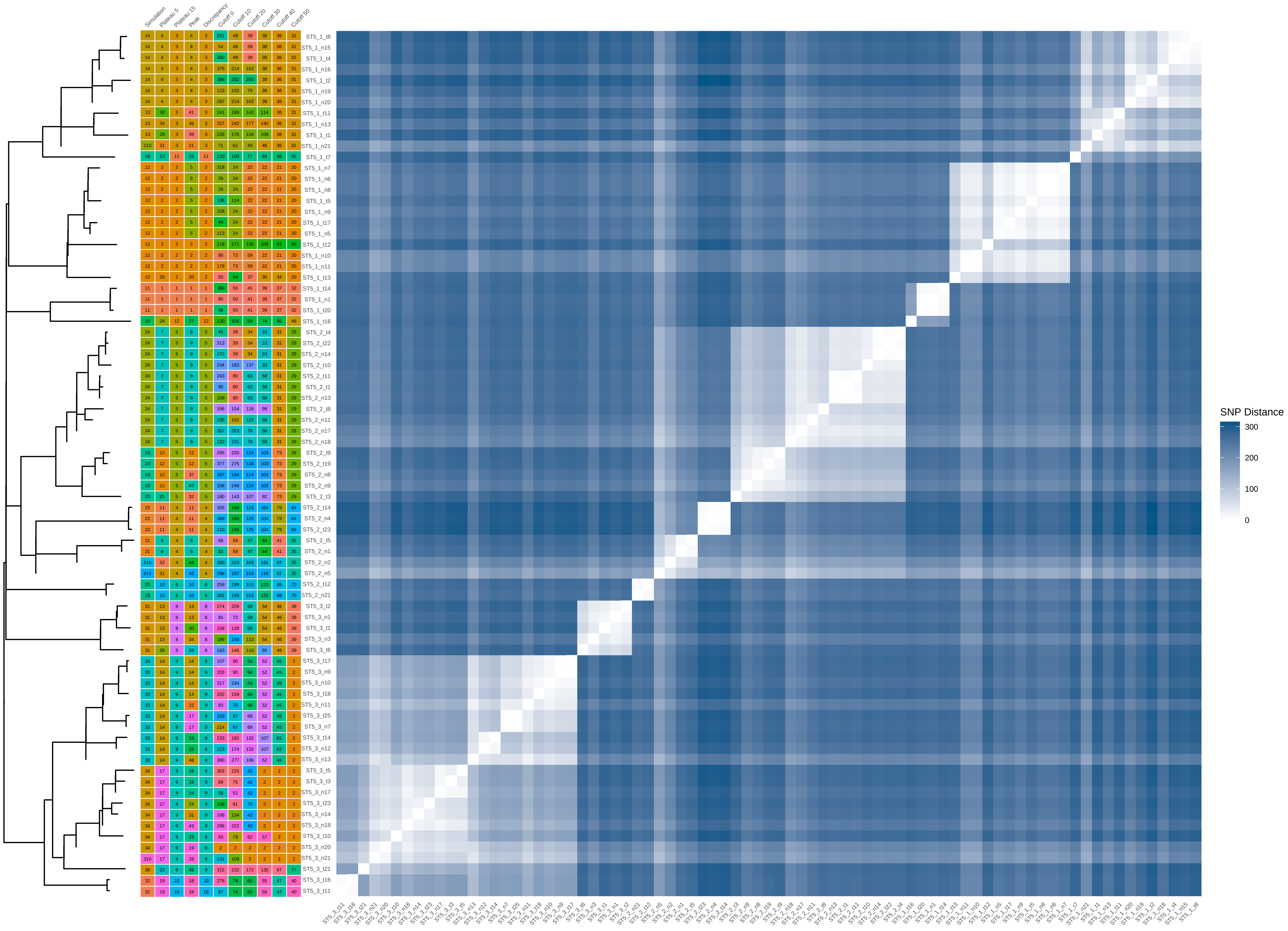

### ST5_60_iter_7.pdf

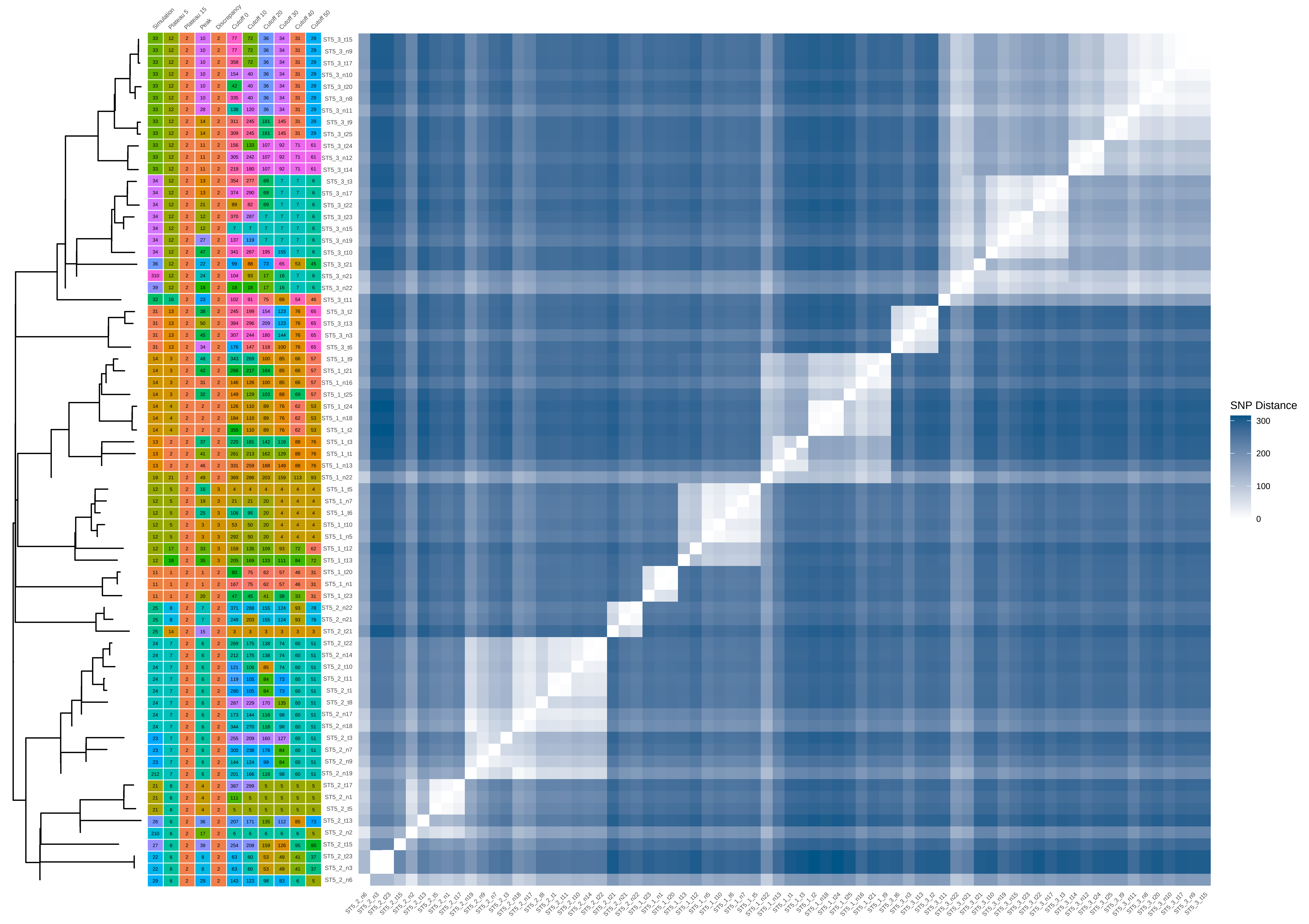

### ST5_80_iter_8.pdf

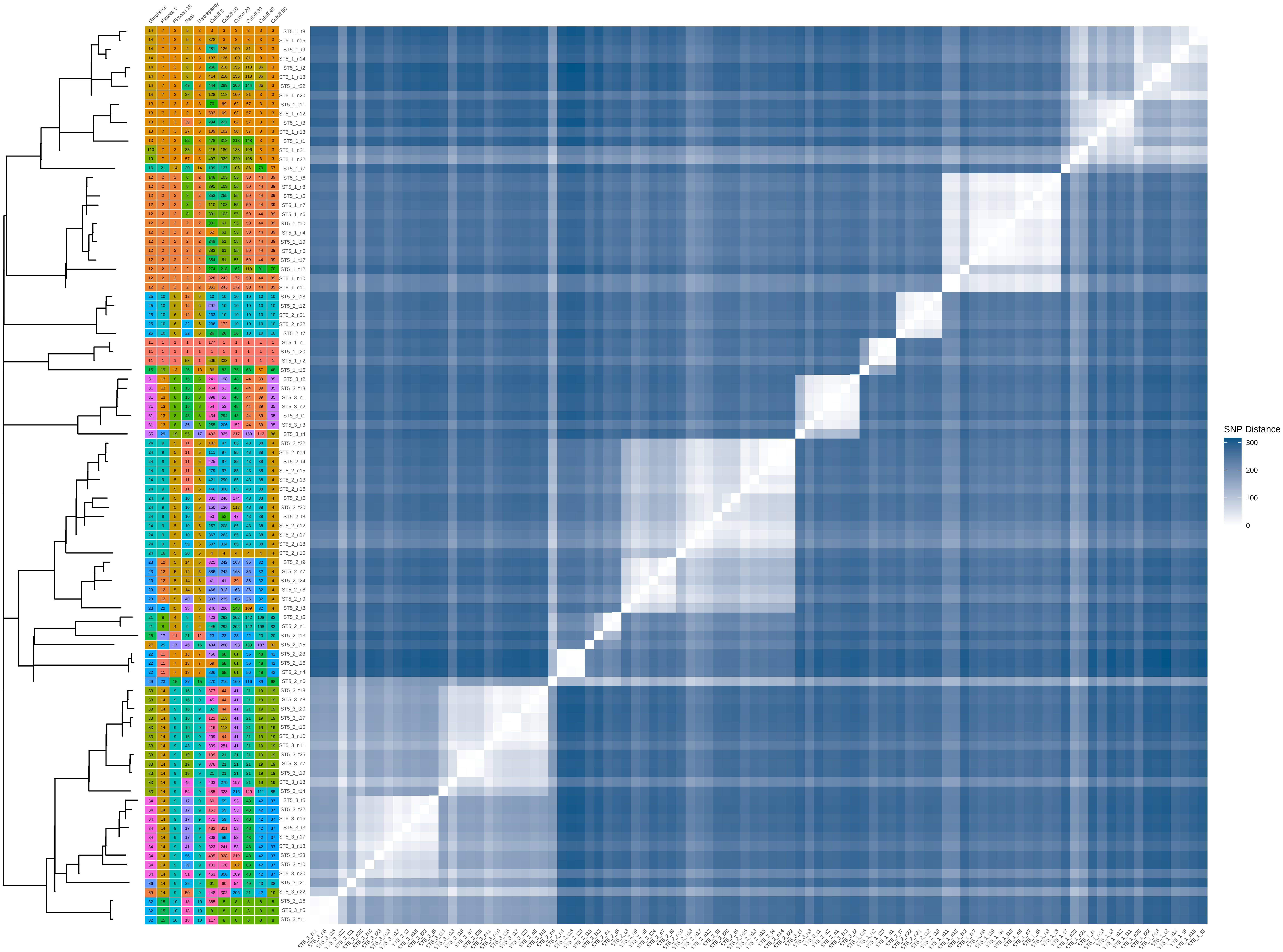

### ST5_80_iter_9.pdf

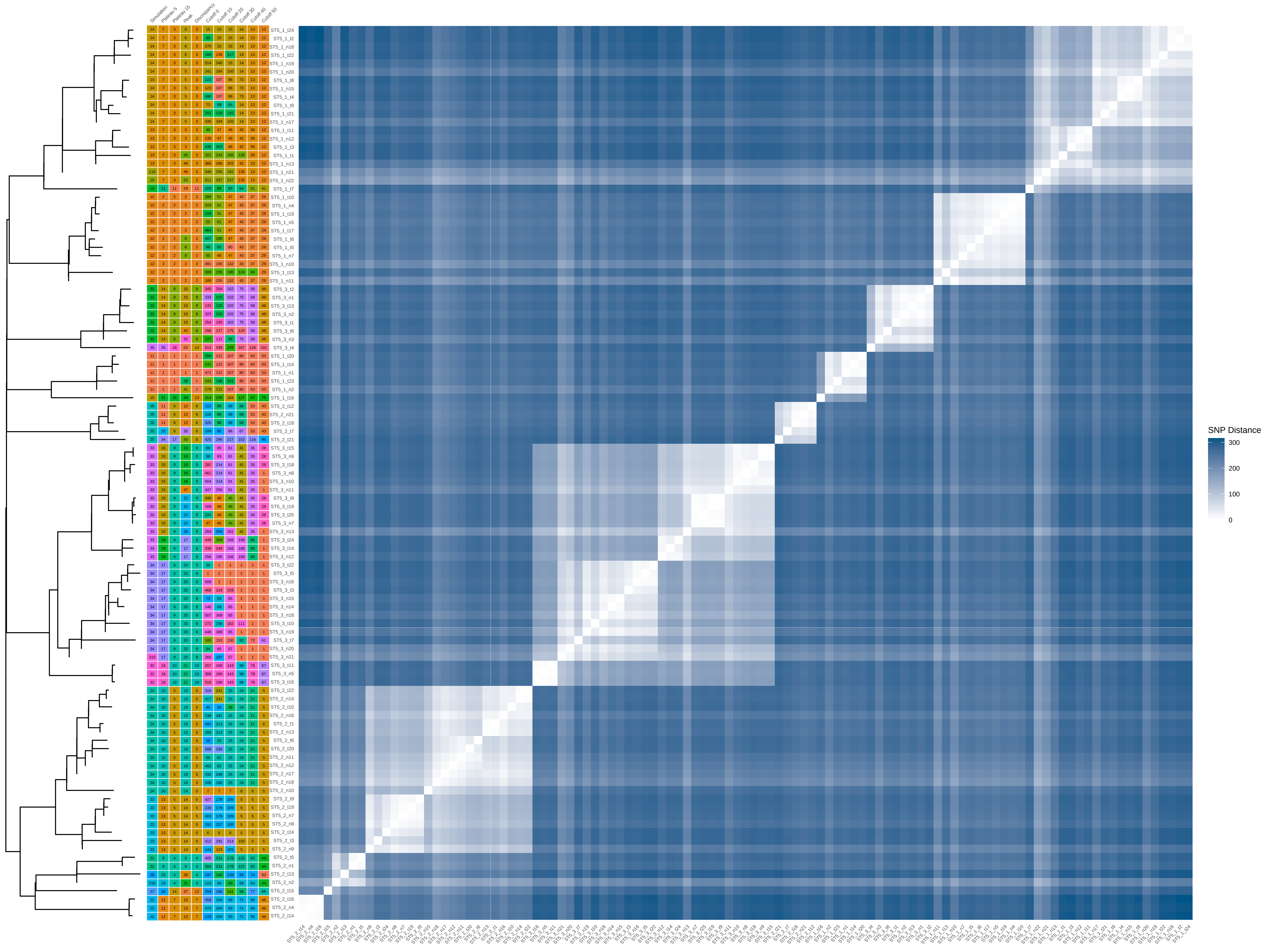

### ST8_80_iter_10.pdf

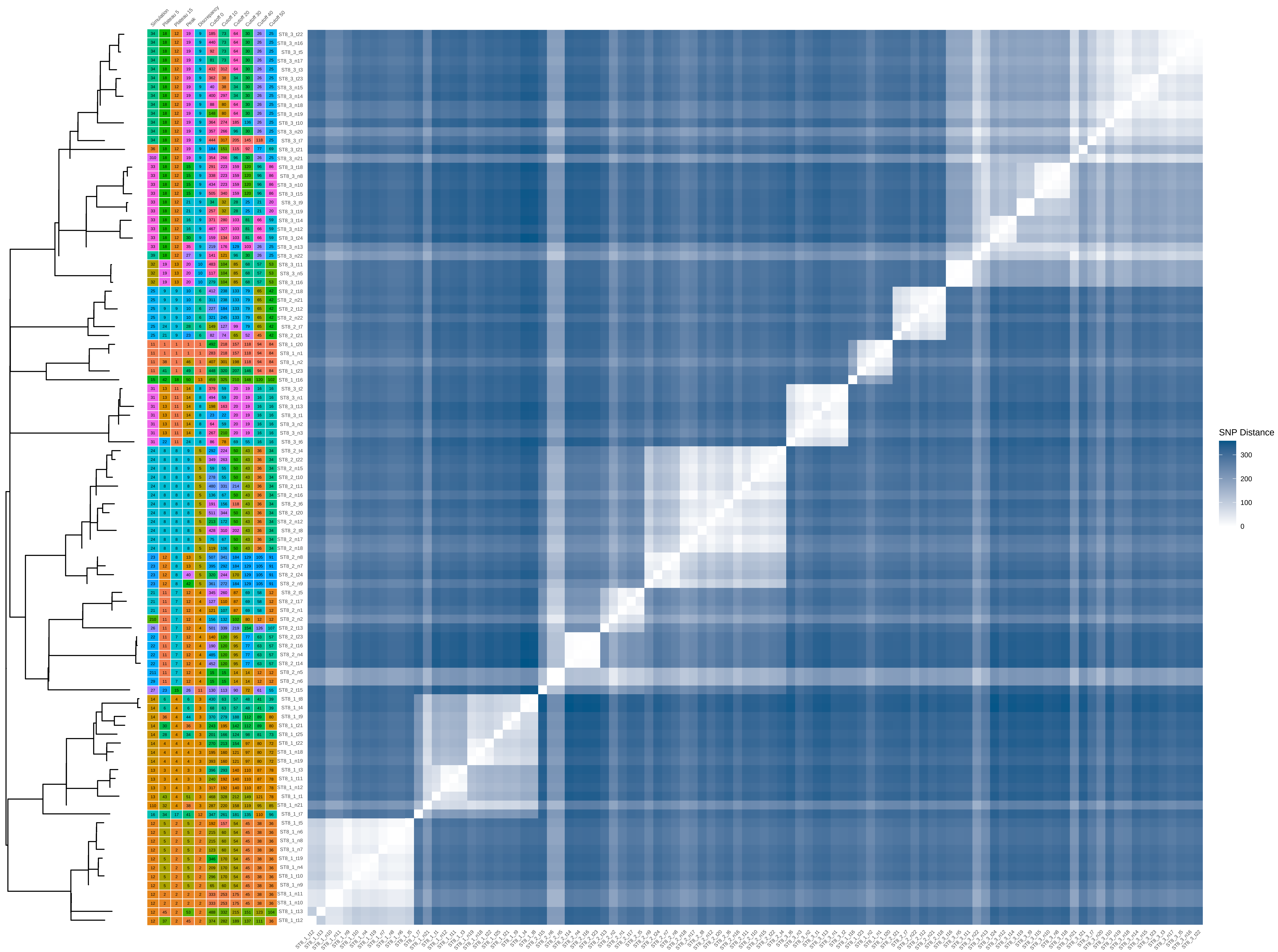

### ST8_100_iter_1.pdf

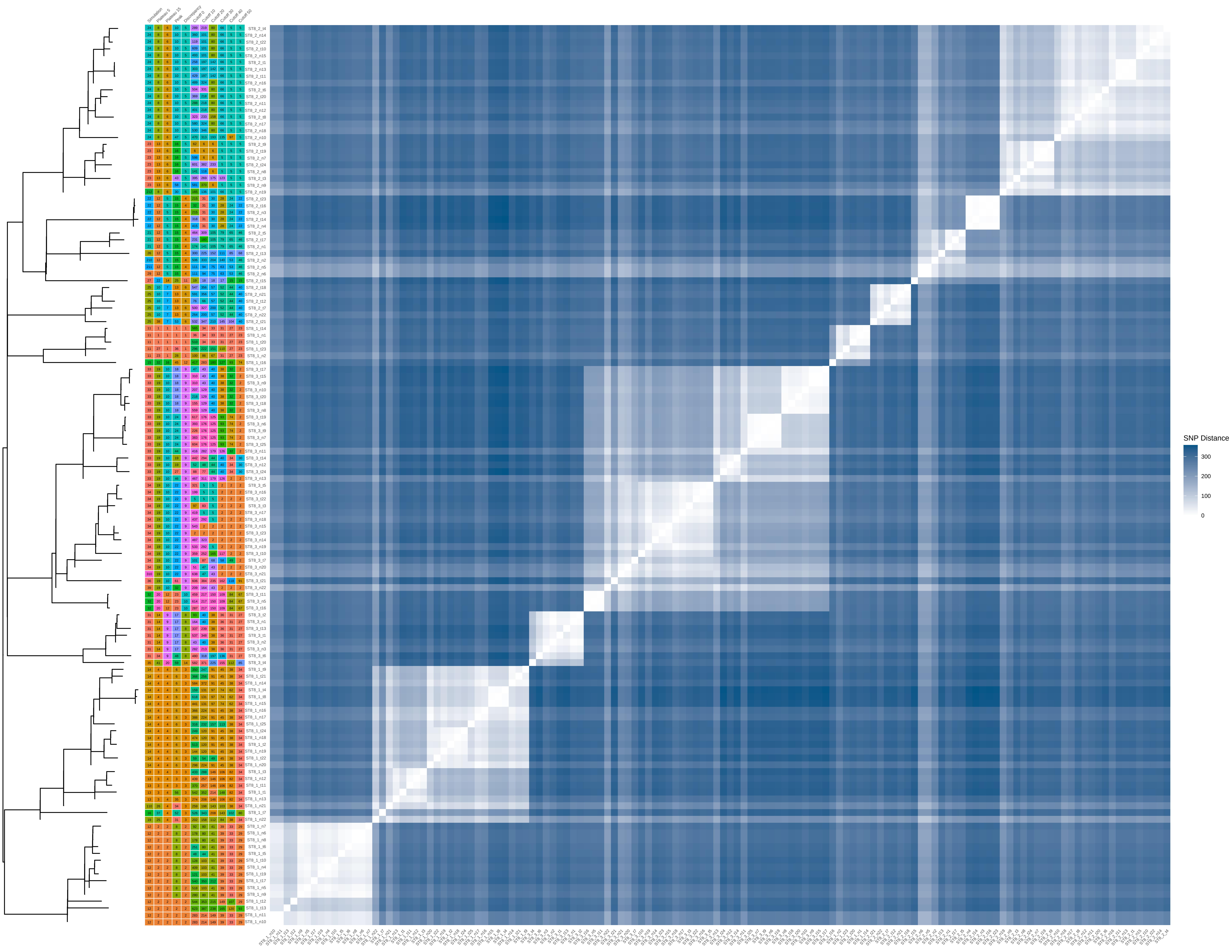

### ST8_100_iter_2.pdf

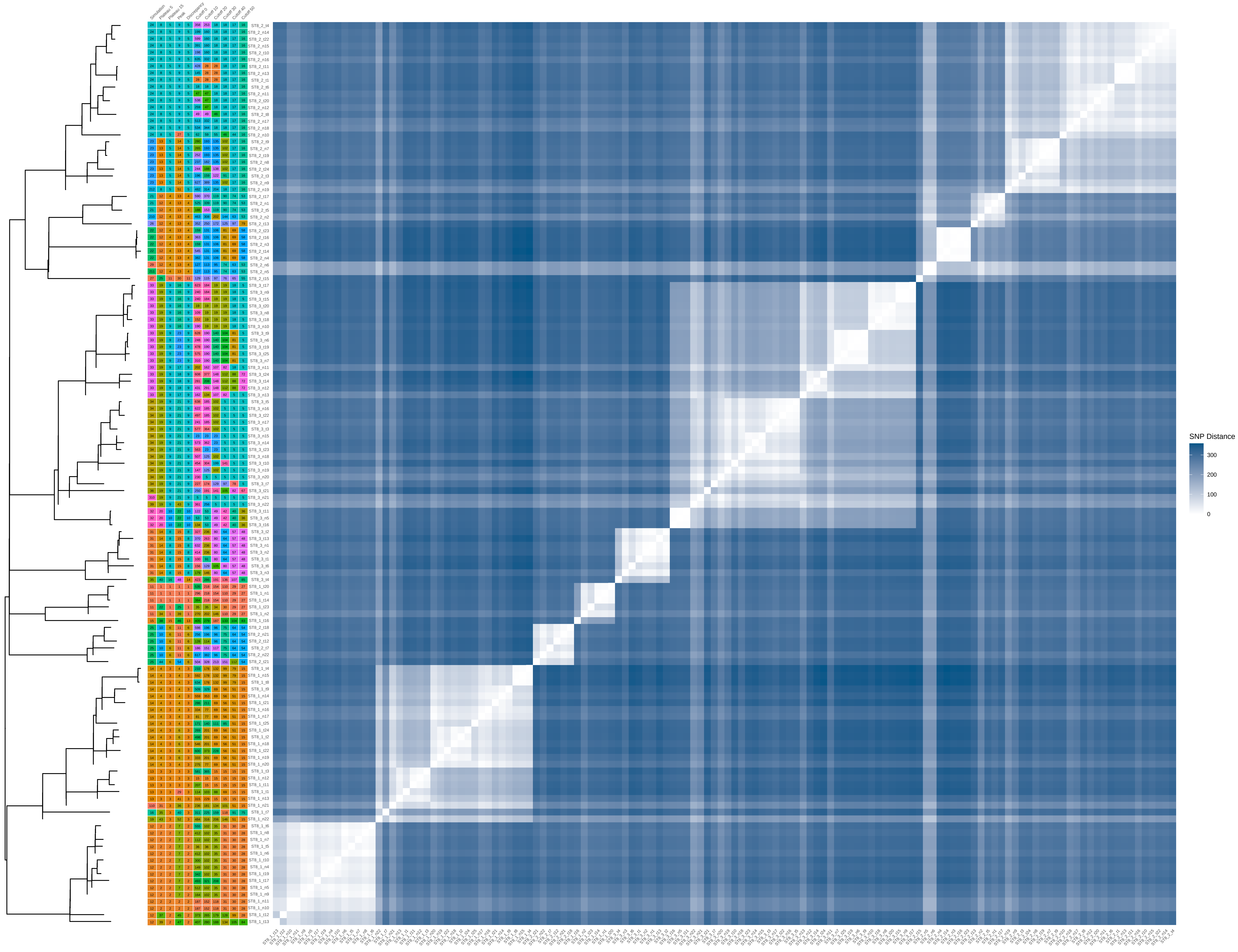

### ST8_100_iter_3.pdf

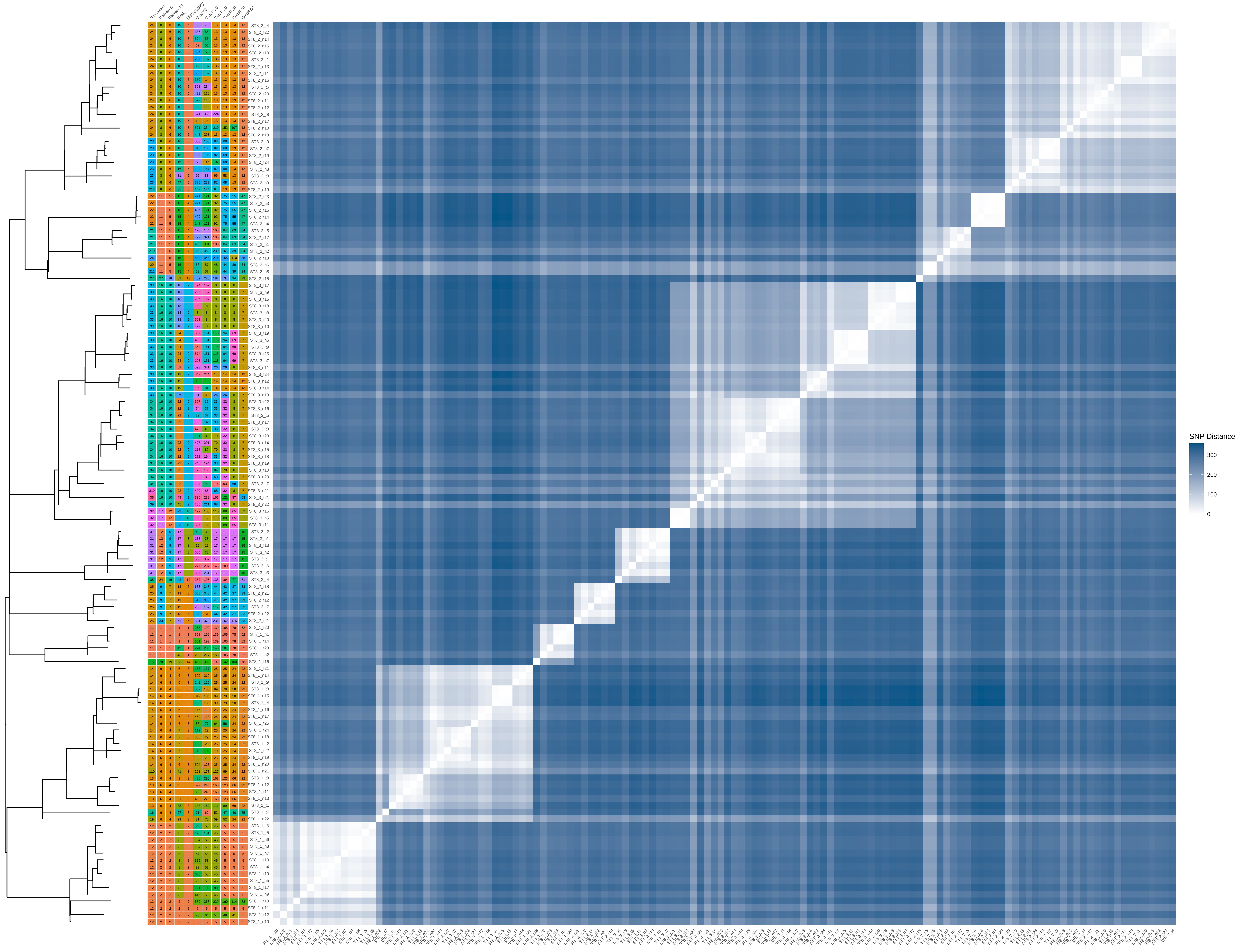

### ST8_100_iter_5.pdf

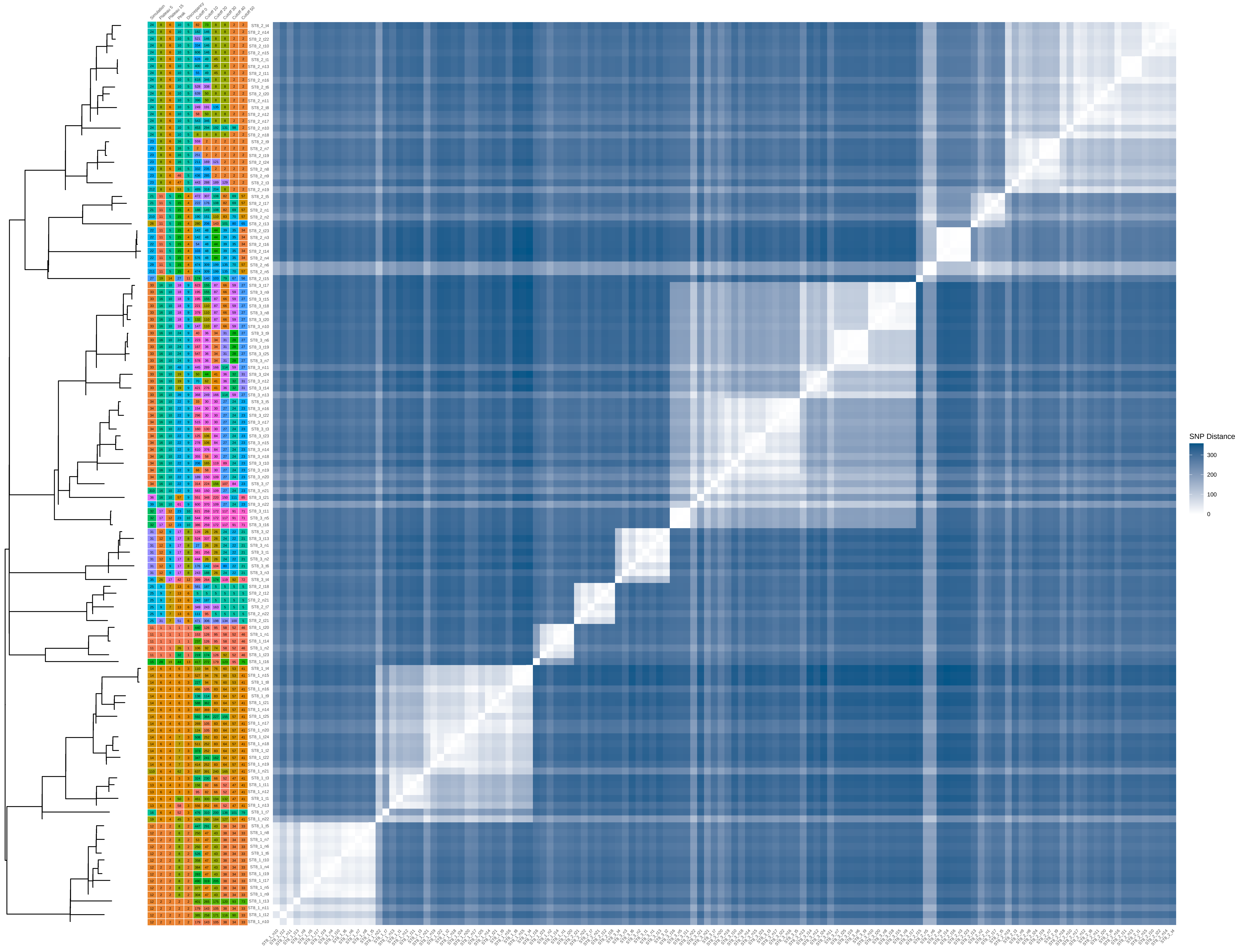

### ST8_100_iter_6.pdf

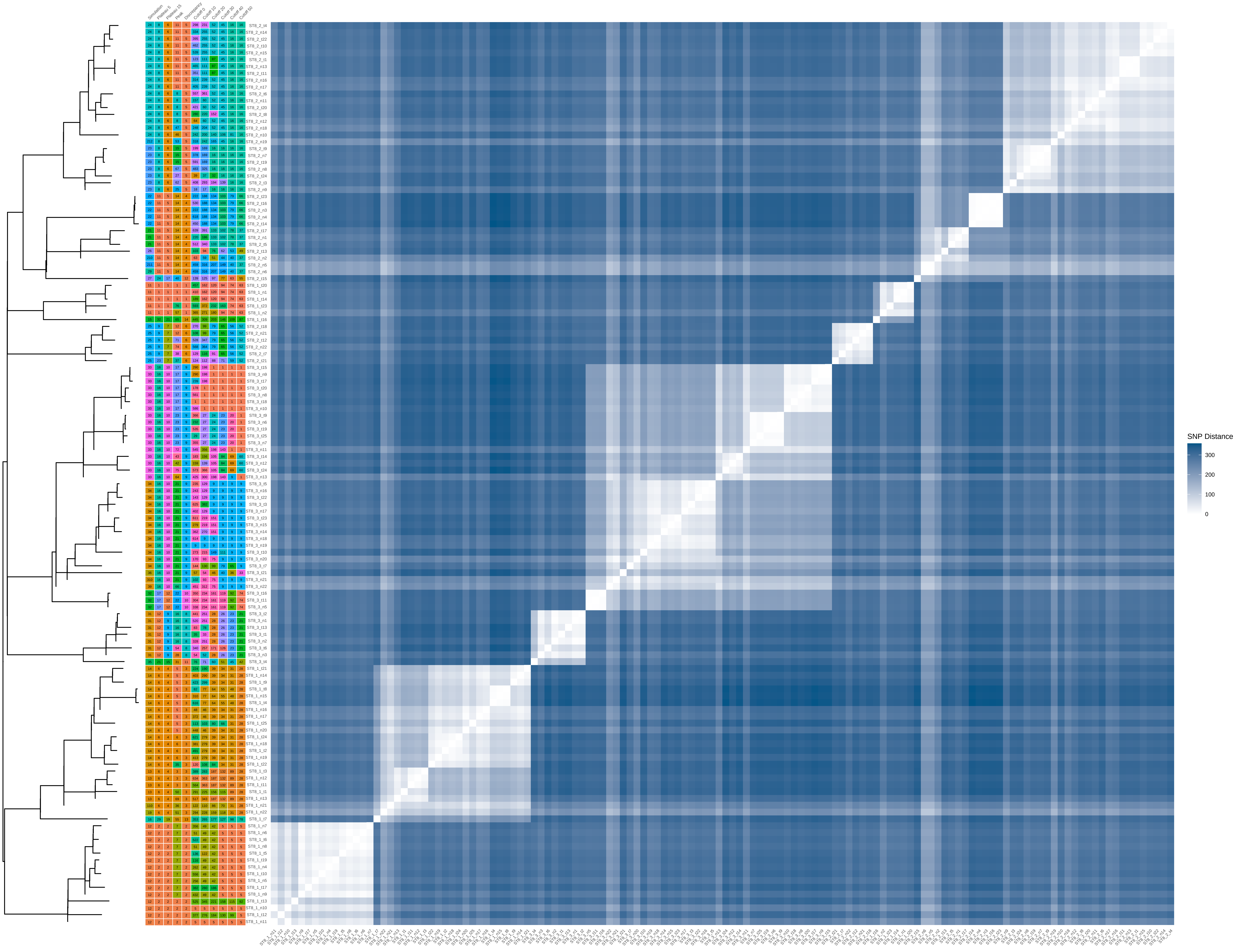

### ST8_100_iter_7.pdf

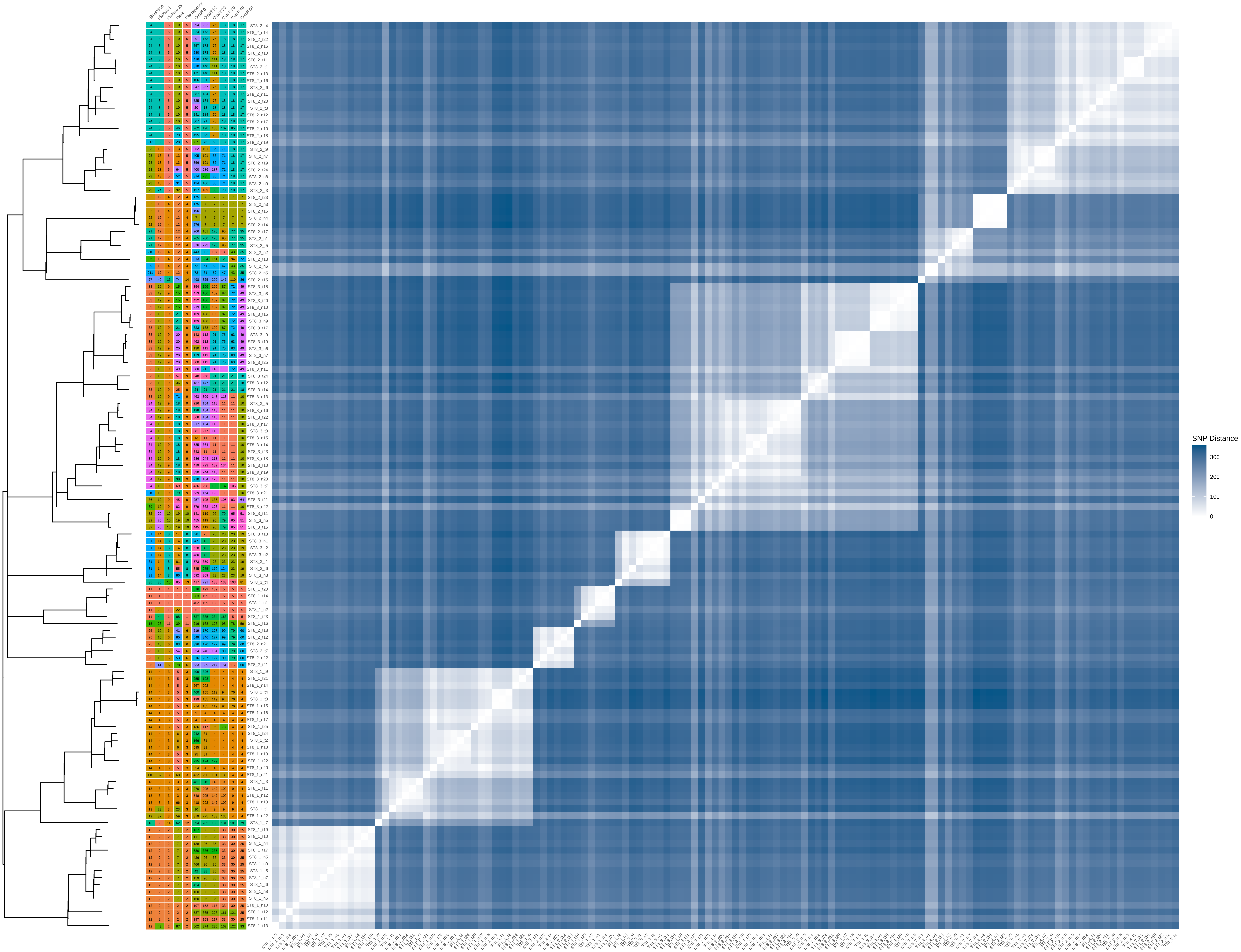

### ST22_40_iter_8.pdf

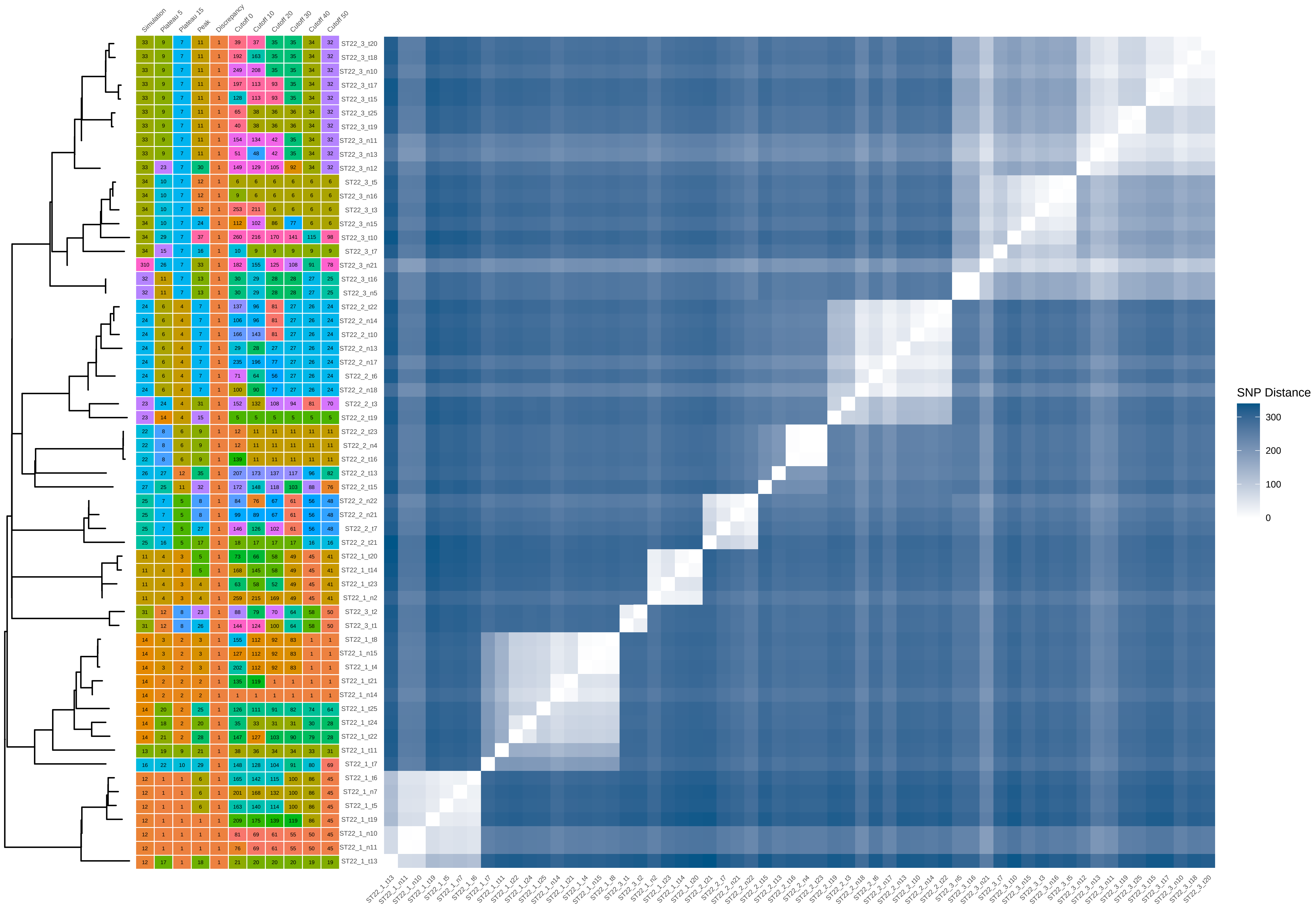

### ST22_40_iter_9.pdf

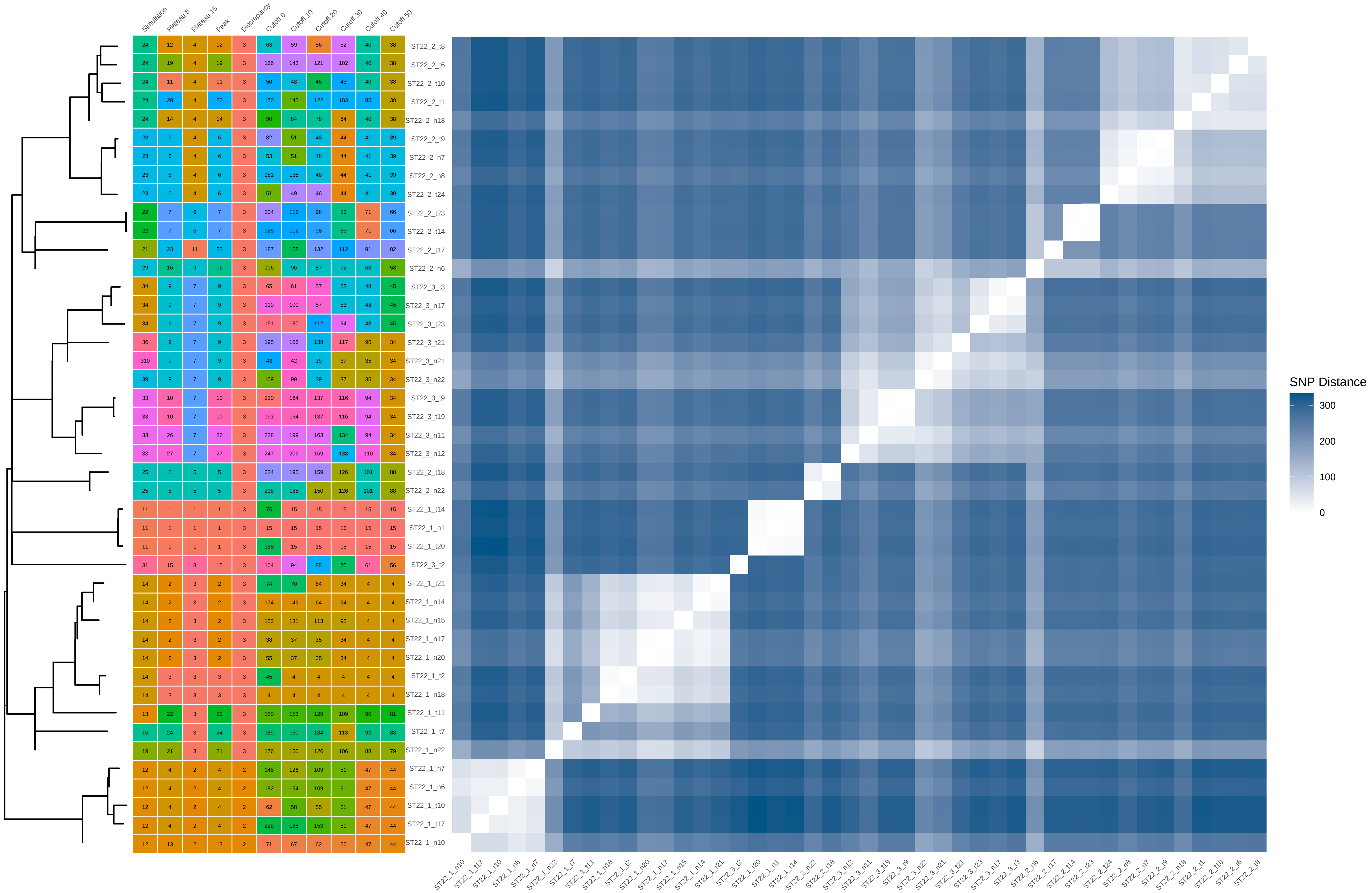

### ST22_60_iter_1.pdf

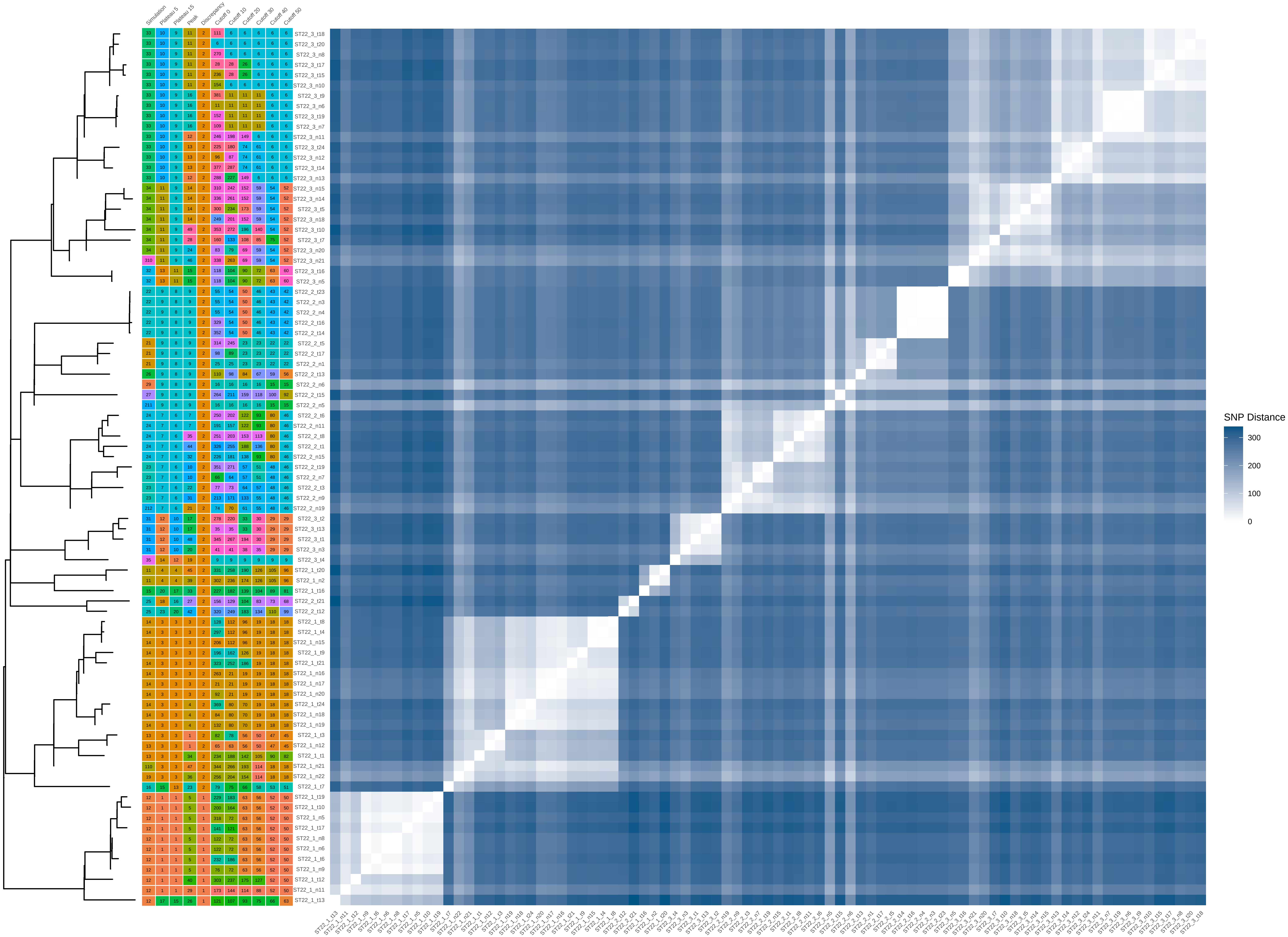

### ST22_60_iter_2.pdf

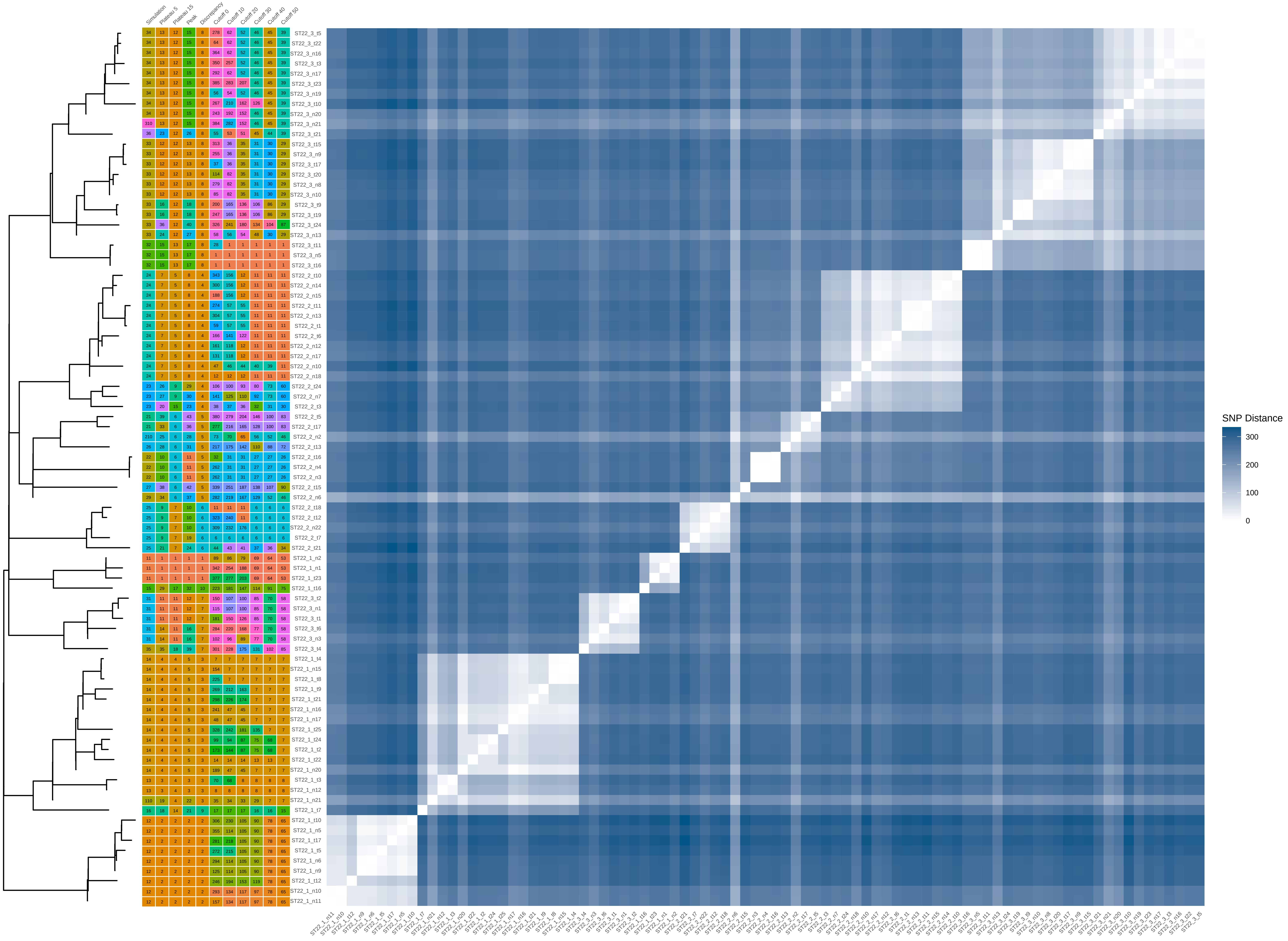

### ST22_60_iter_4.pdf

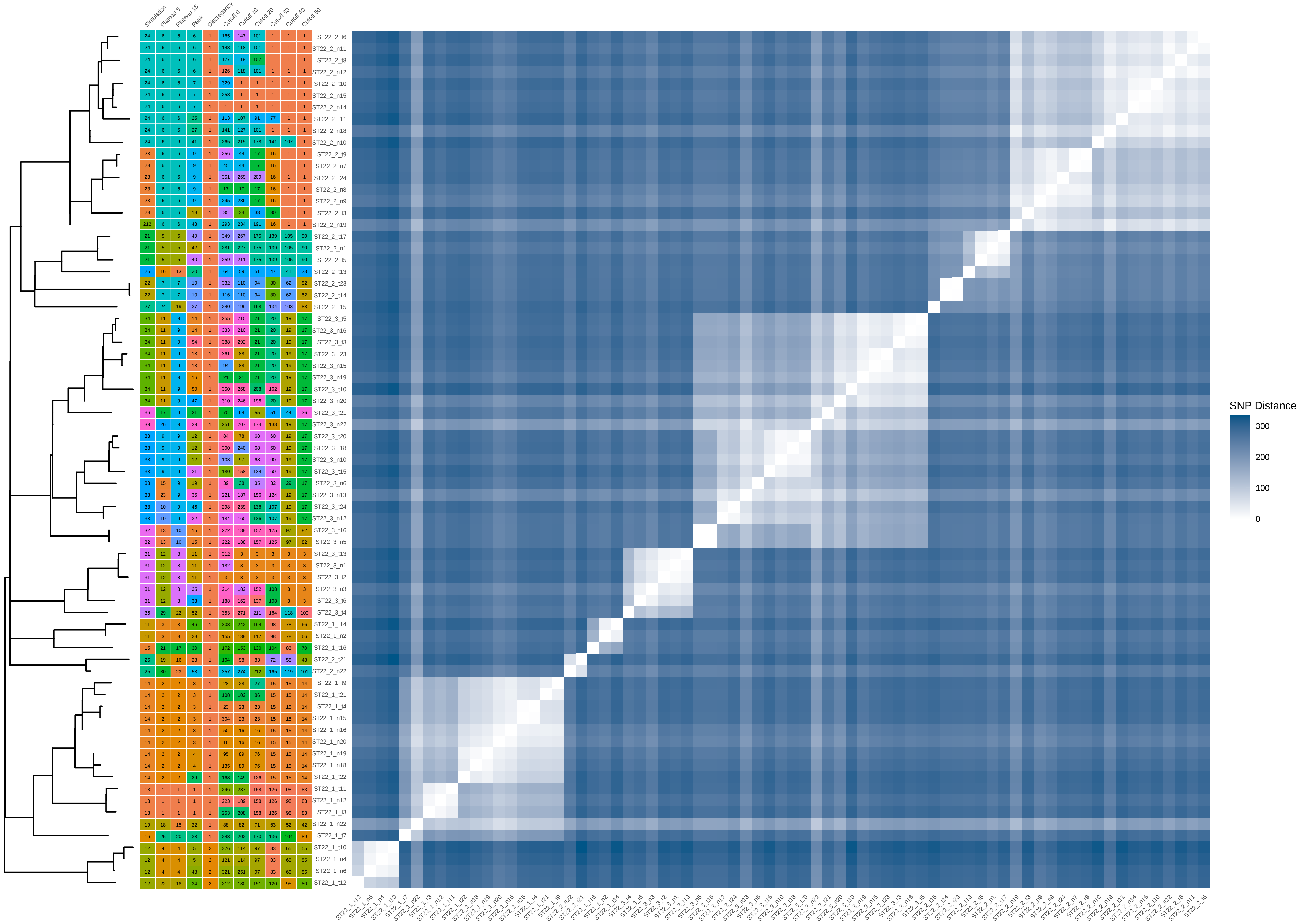

### ST22_60_iter_5.pdf

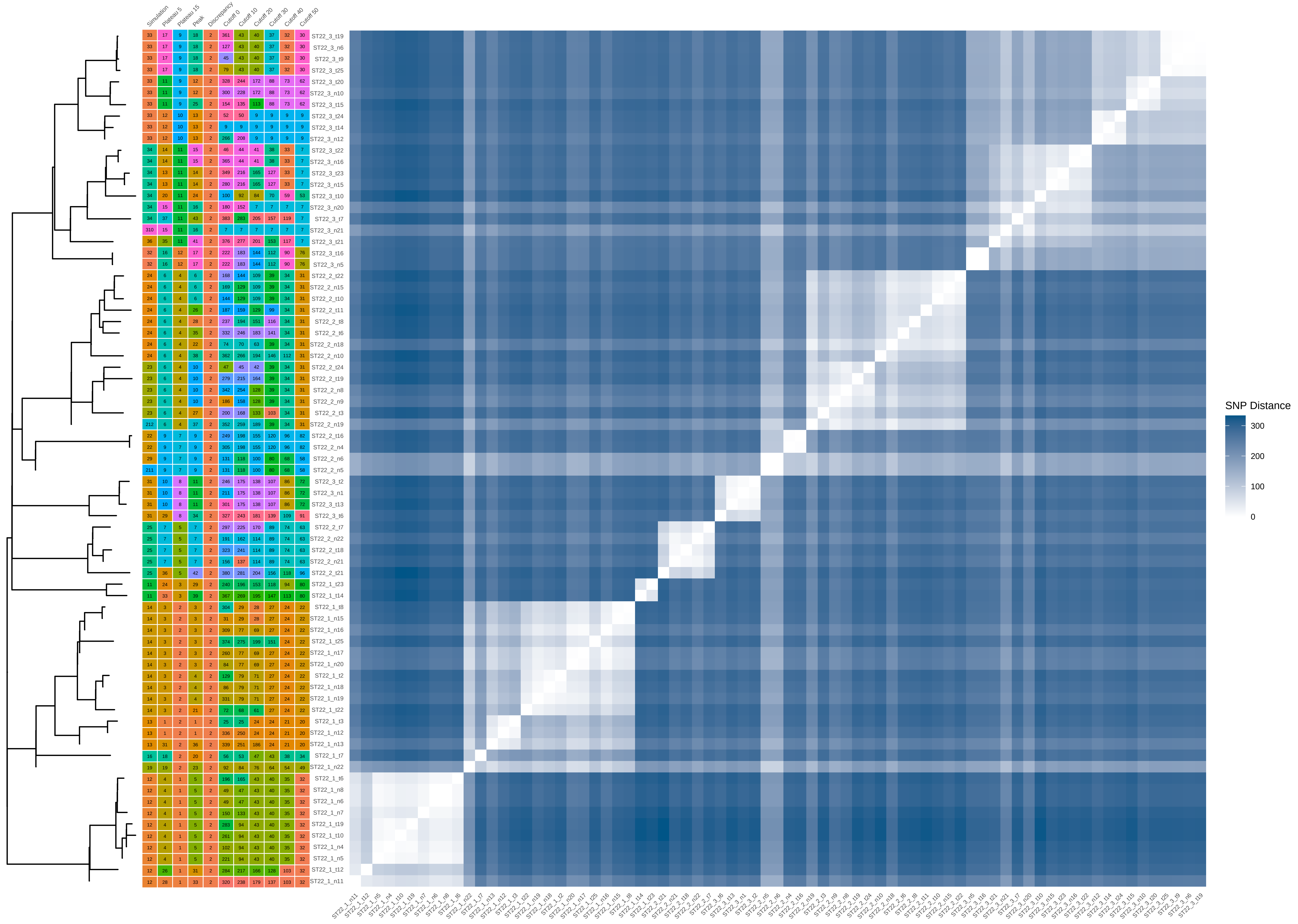

### ST22_60_iter_6.pdf

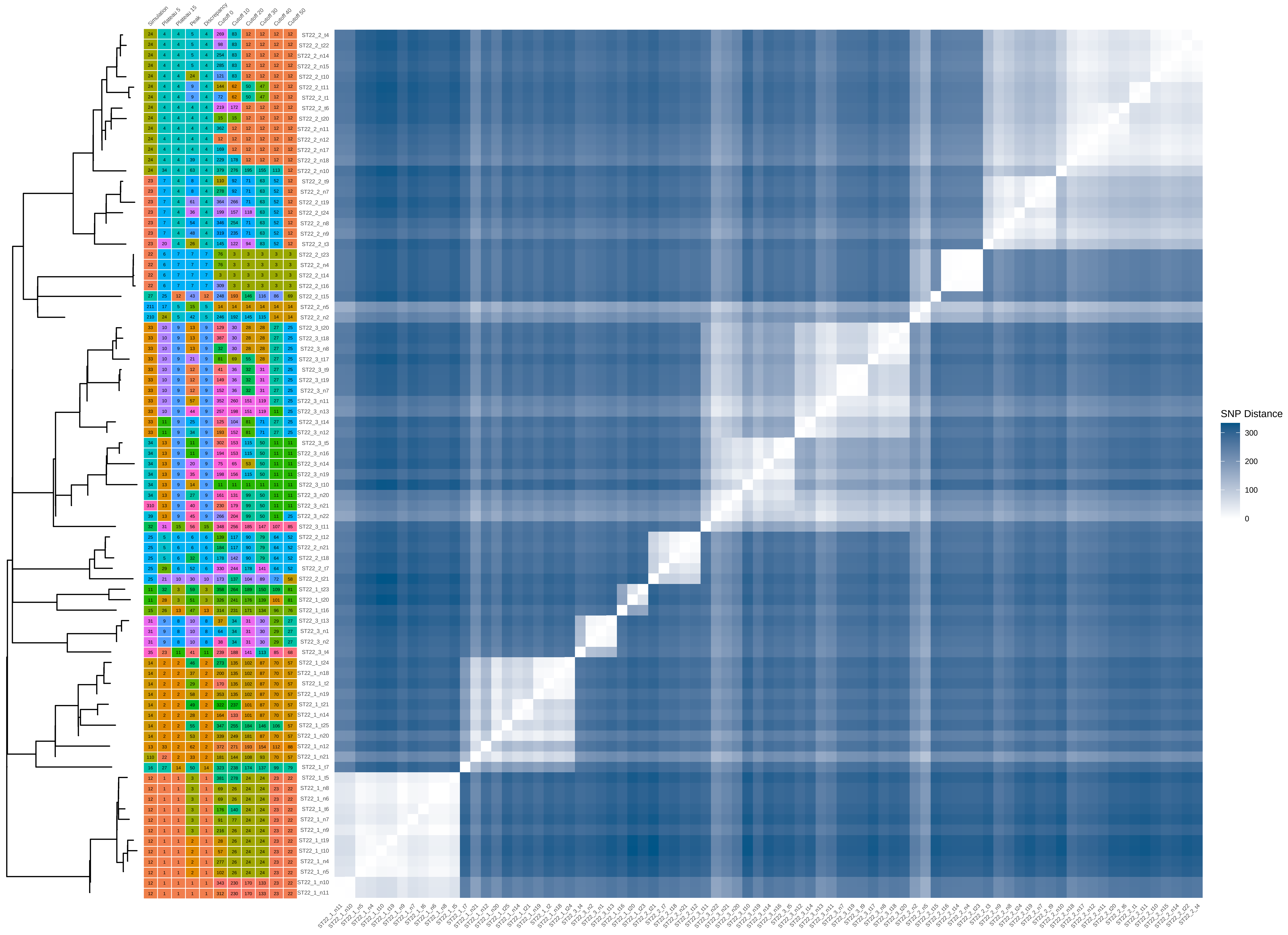

### ST22_60_iter_7.pdf

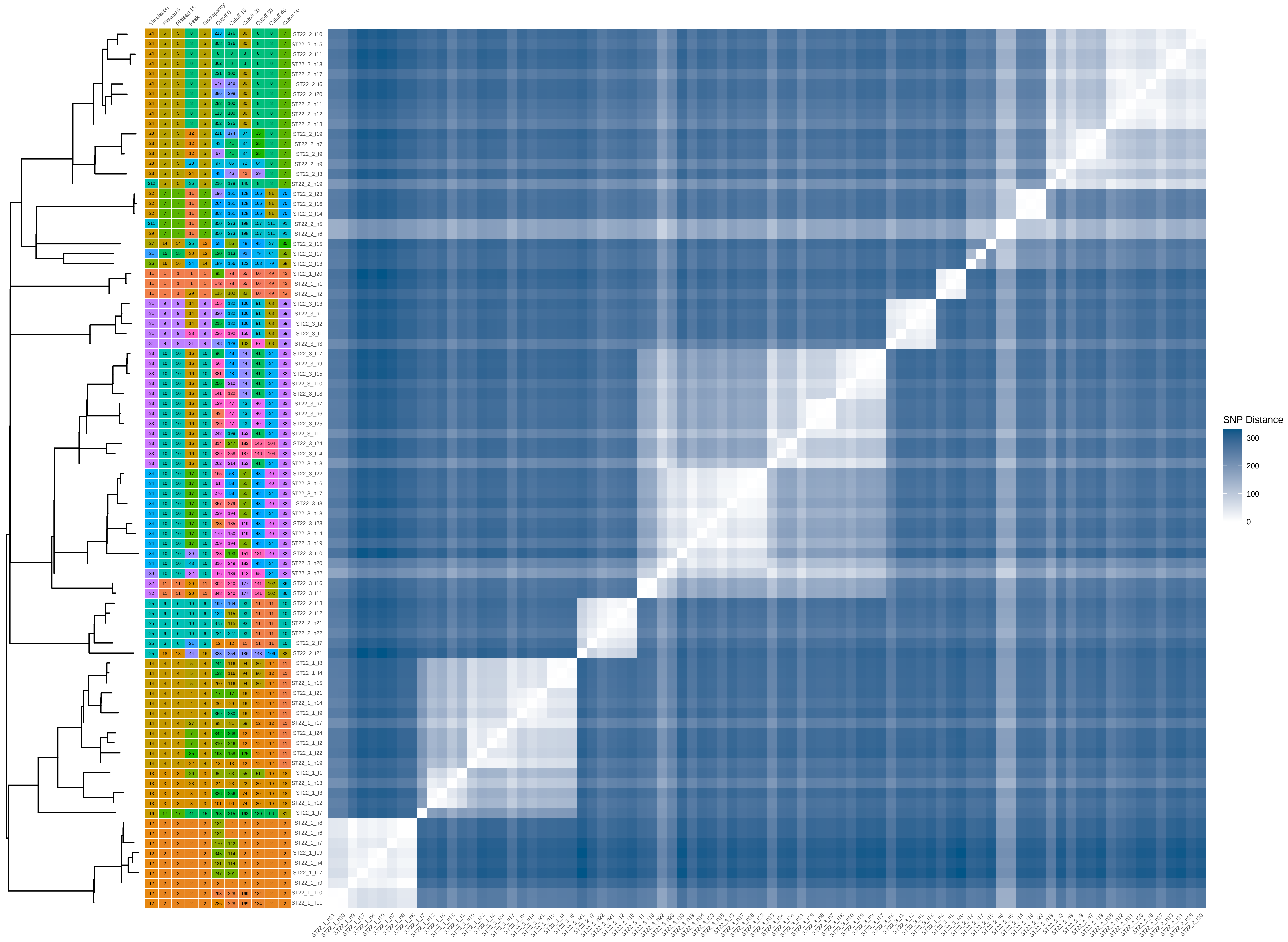

### ST22_80_iter_8.pdf

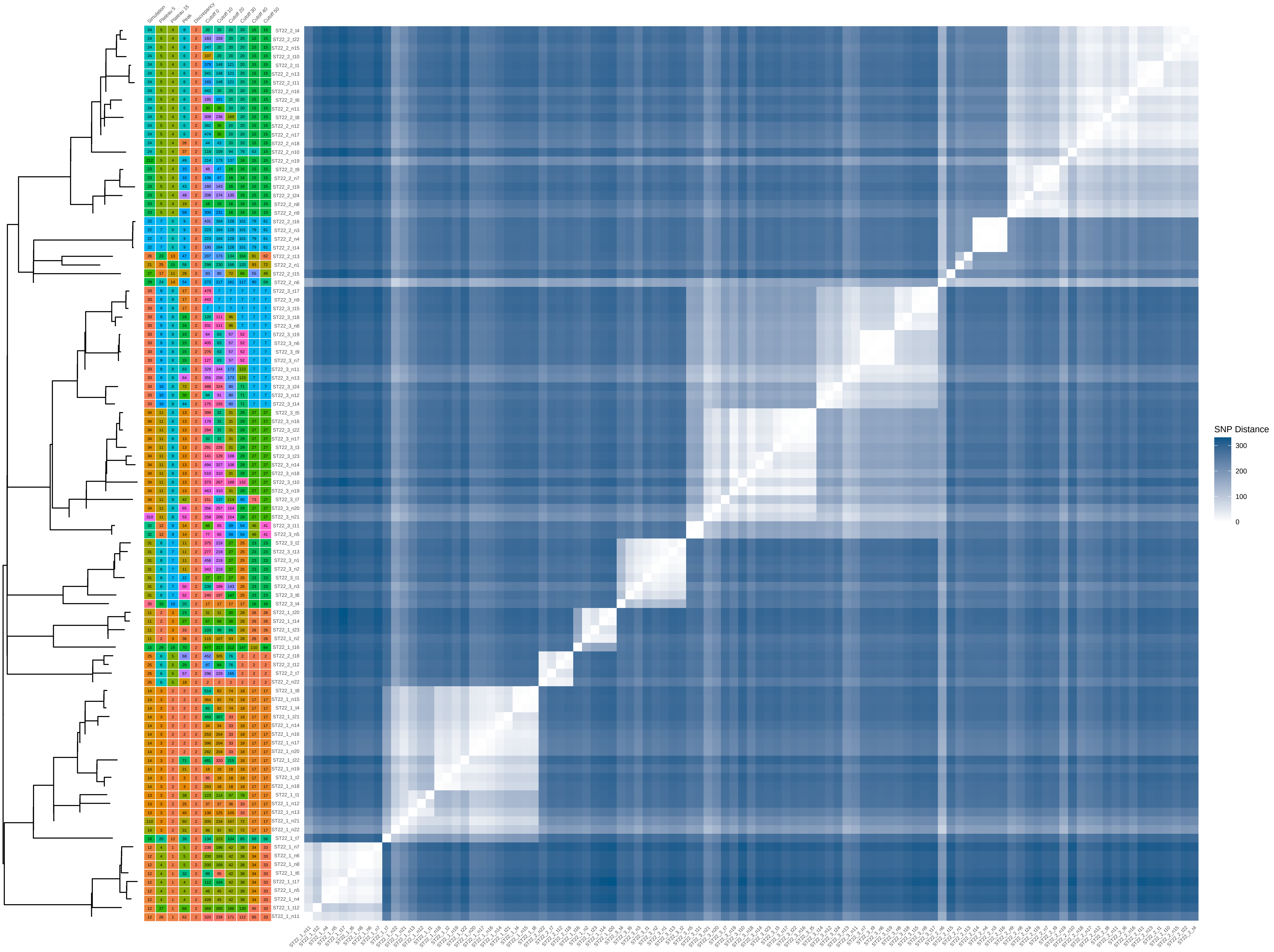

### ST22_80_iter_9.pdf

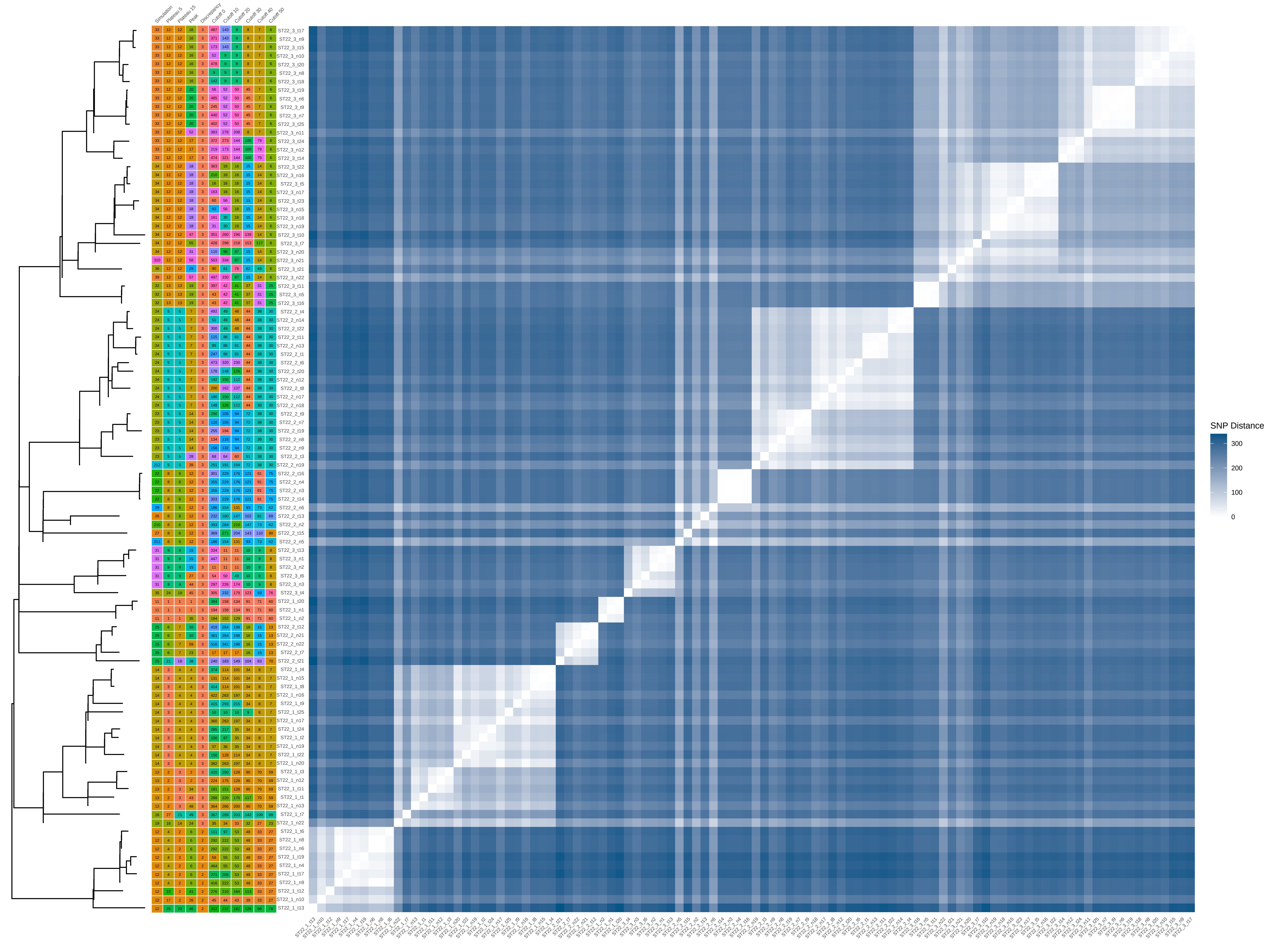

### ST22_100_iter_8.pdf

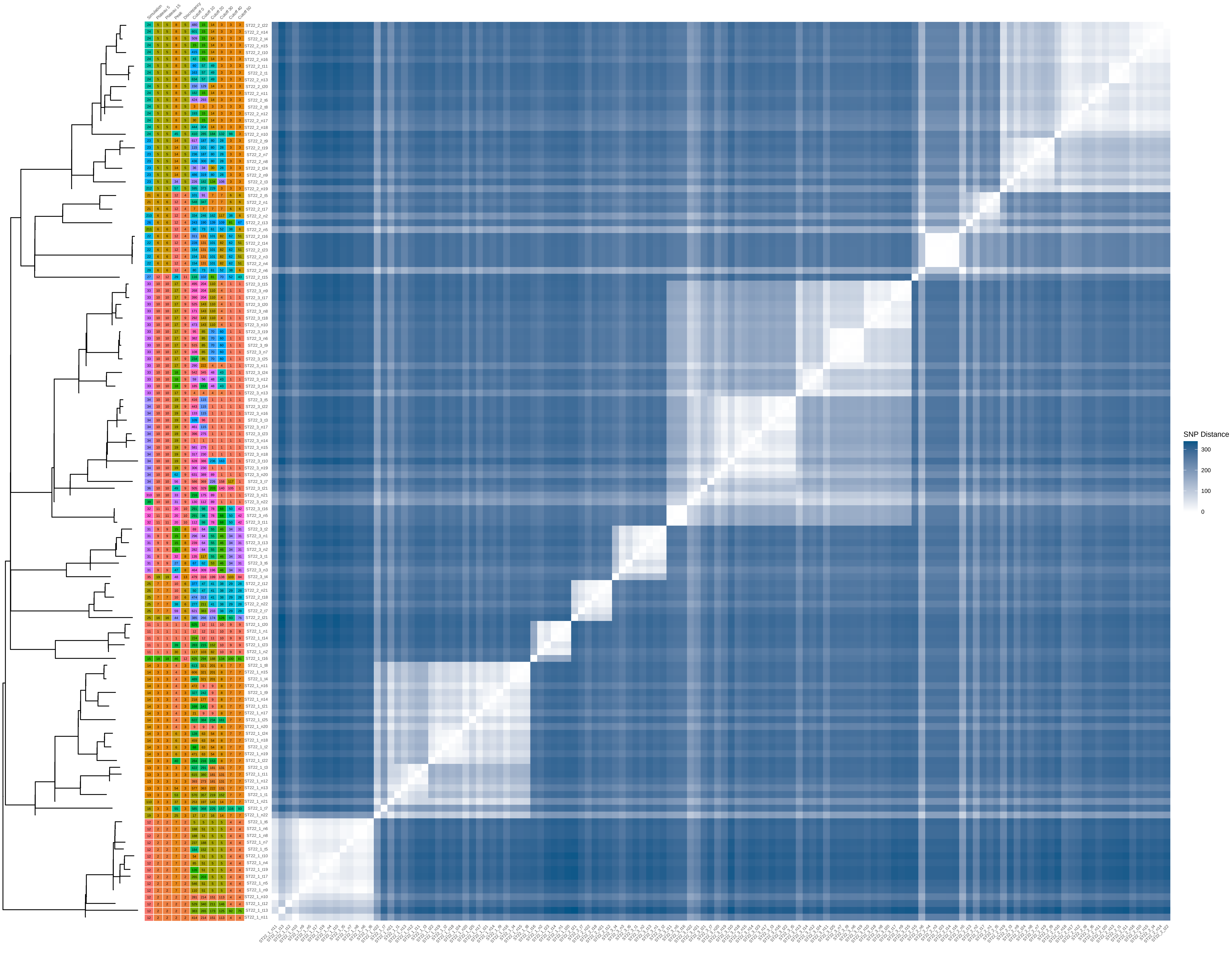

### ST398_40_iter_3.pdf

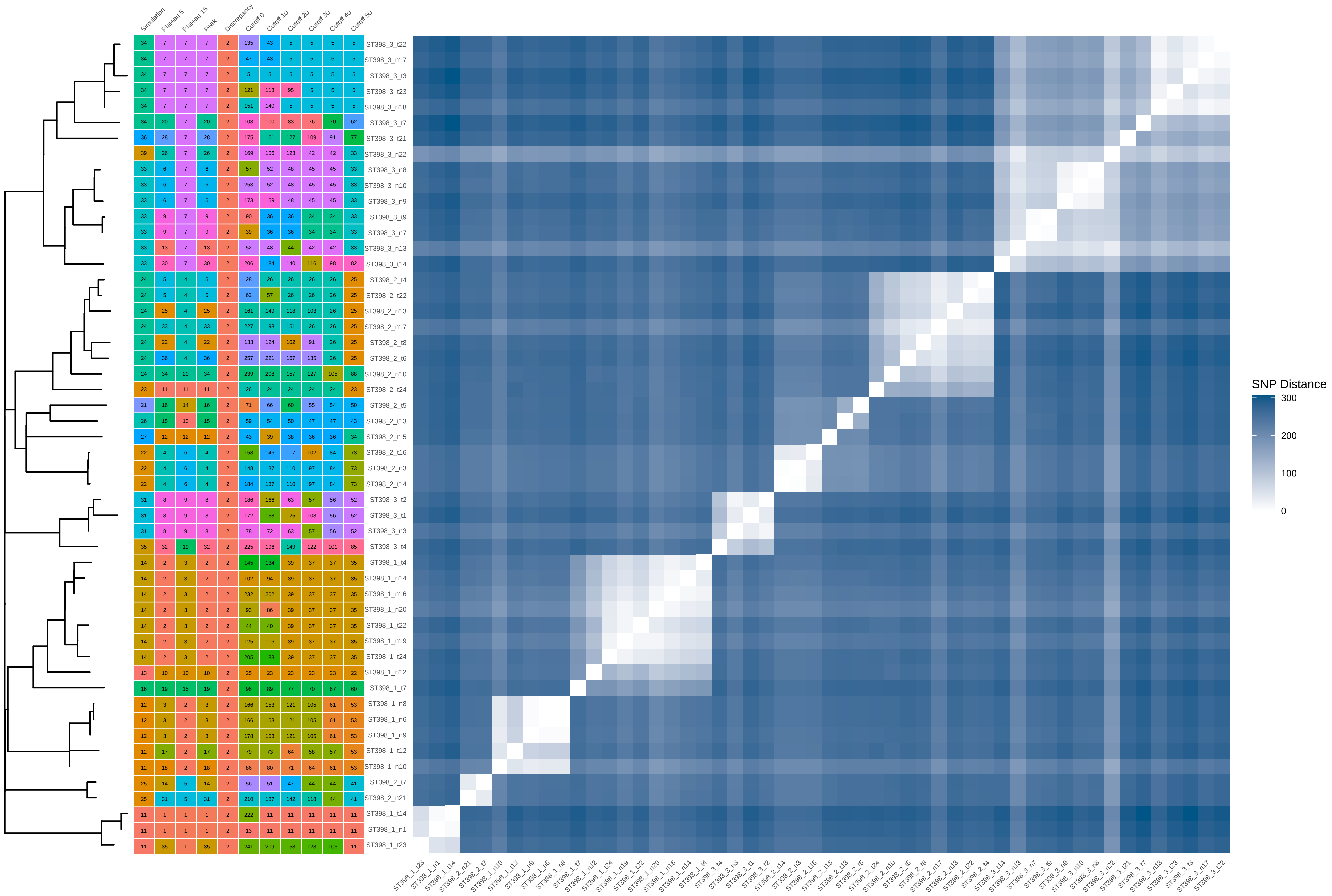

### ST398_40_iter_4.pdf

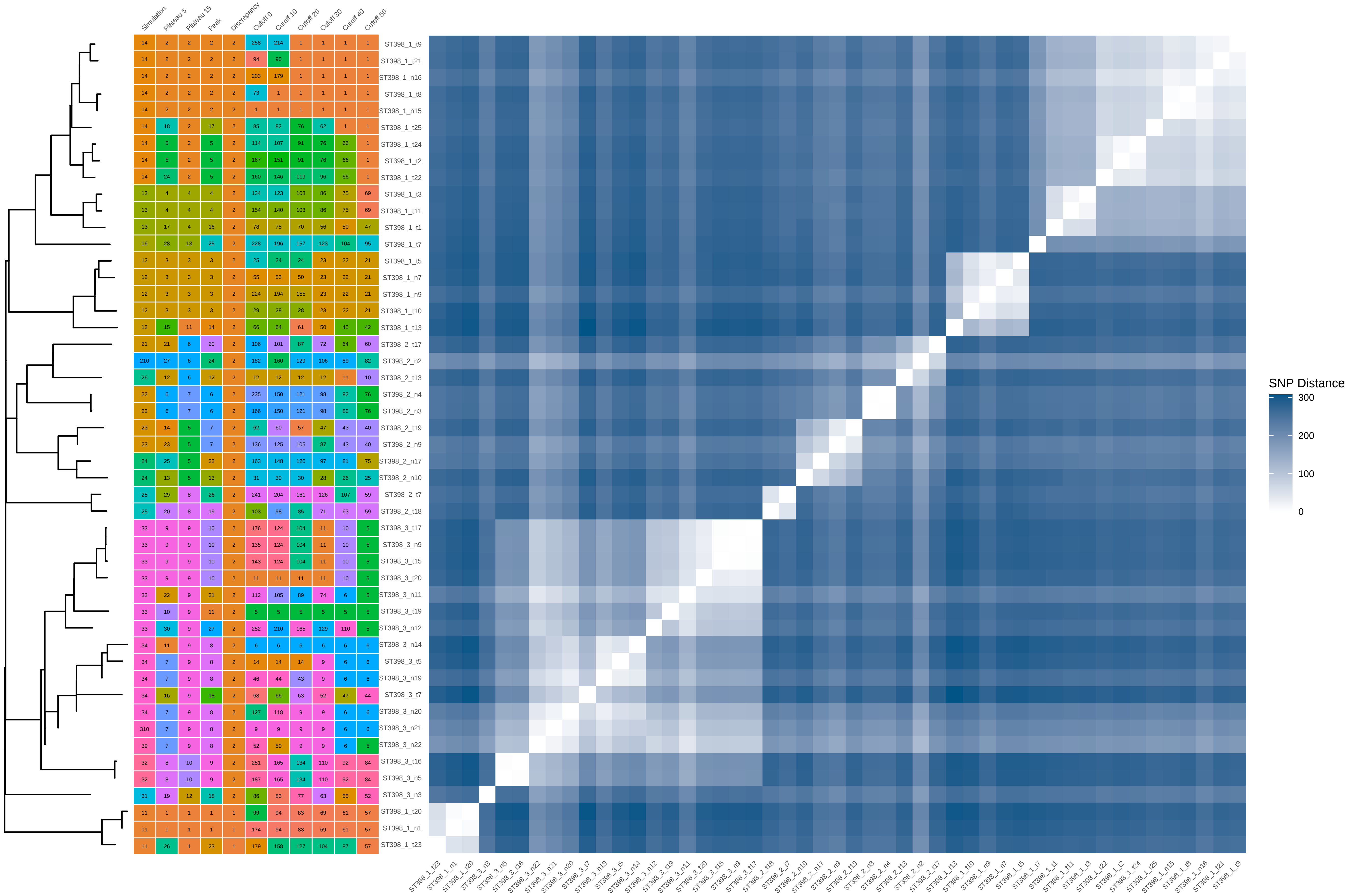

### ST398_40_iter_5.pdf

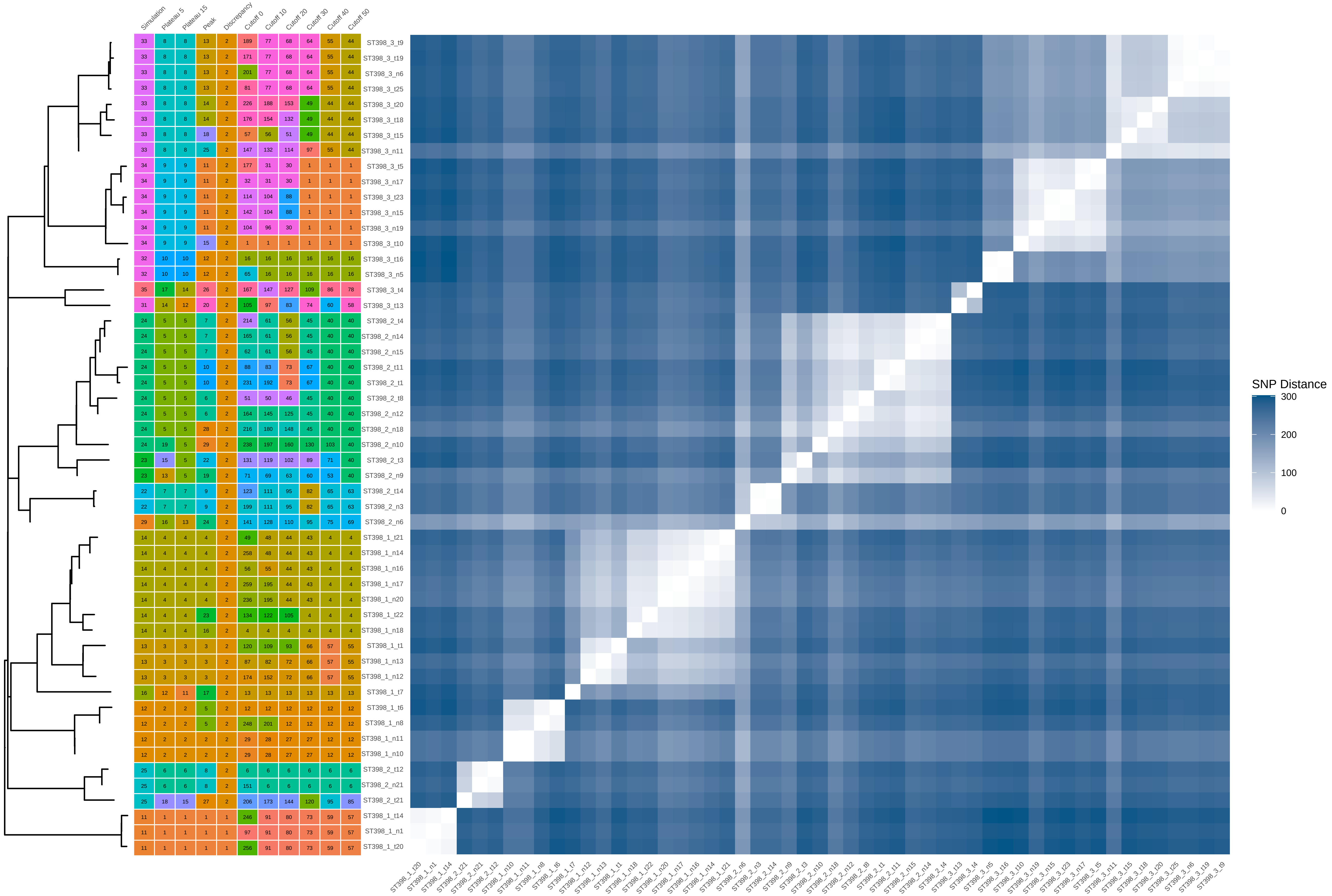

### ST398_40_iter_6.pdf

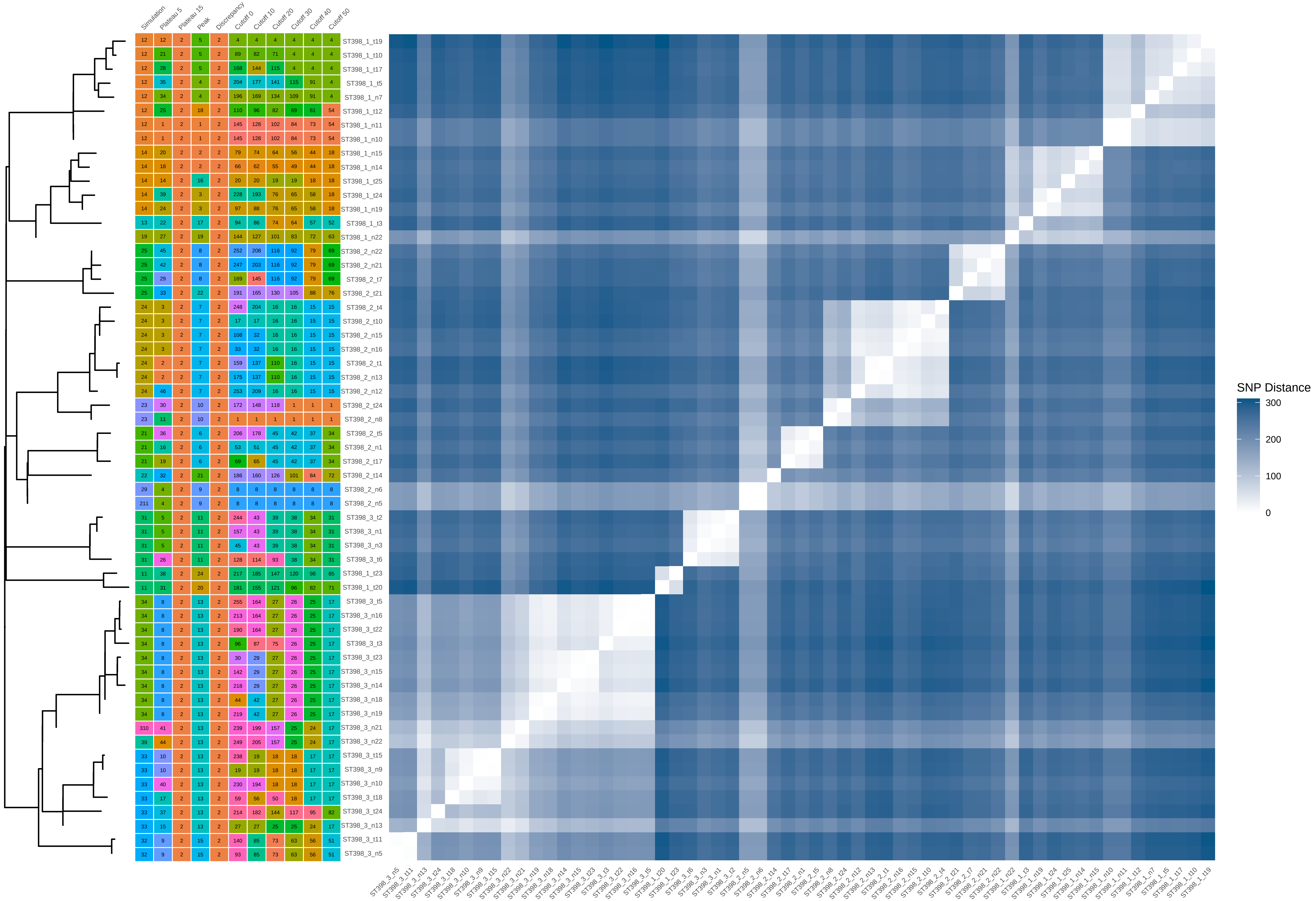

### ST398_40_iter_7.pdf

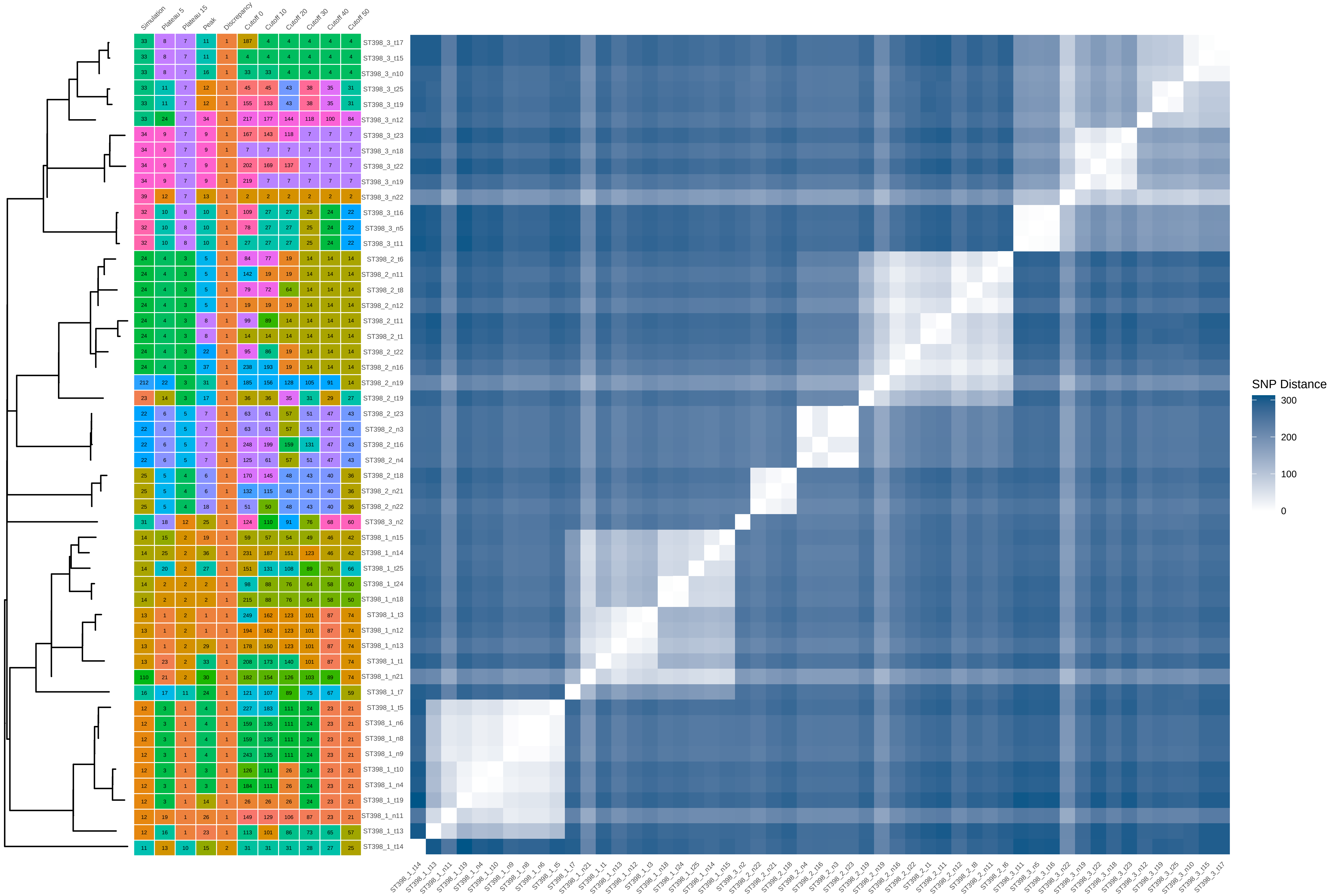

### ST398_40_iter_8.pdf

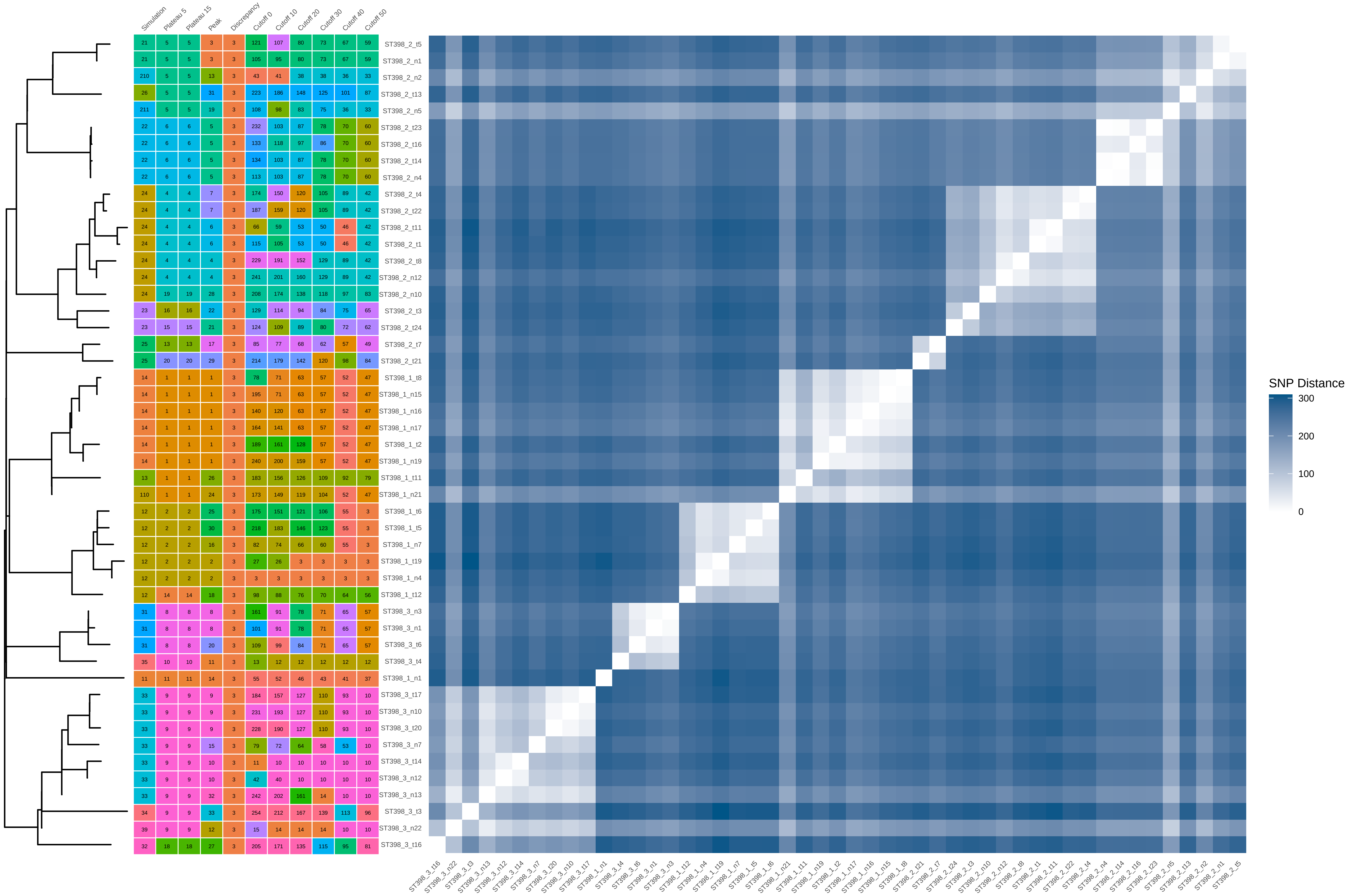

### ST398_60_iter_9.pdf

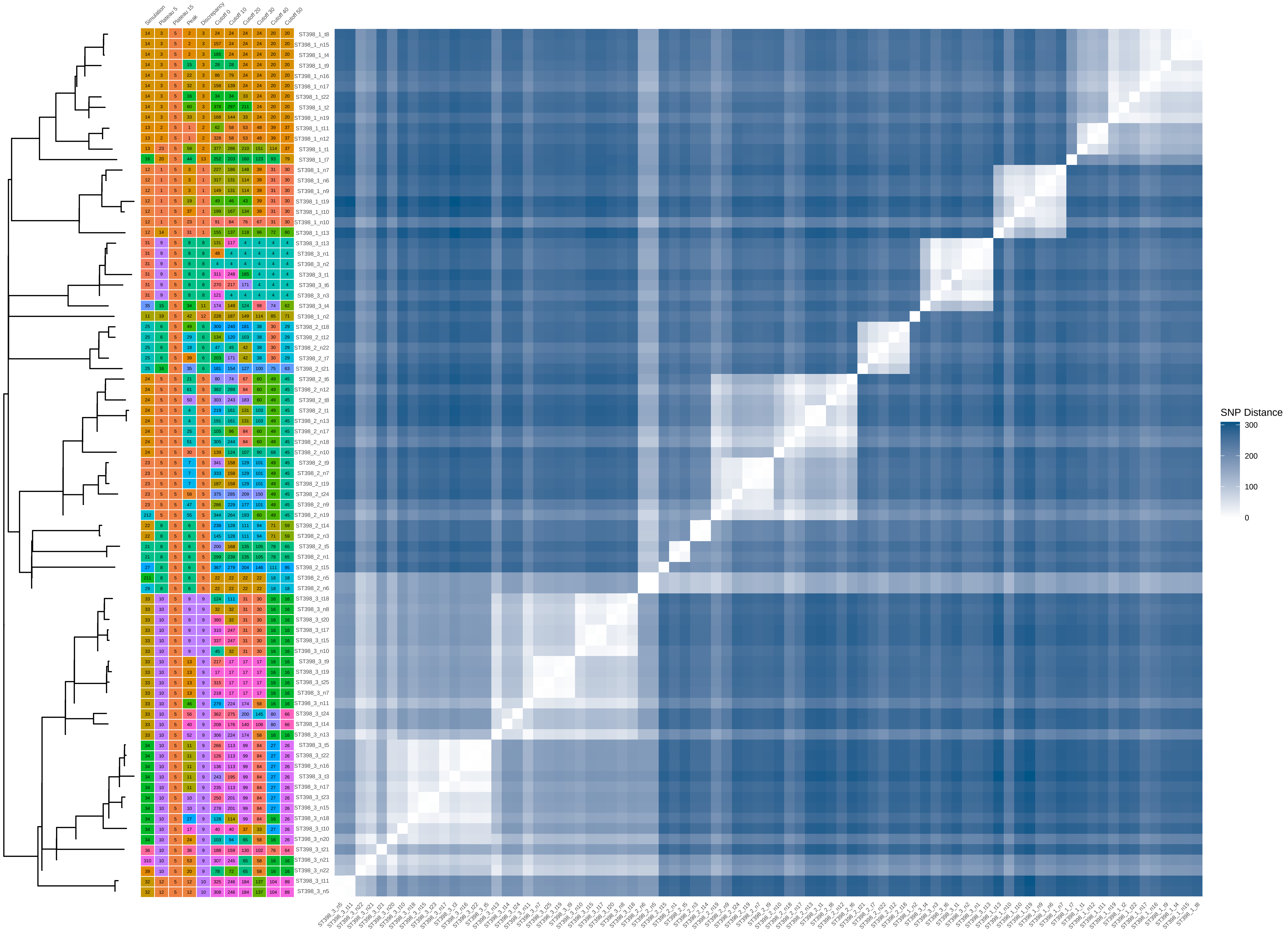

### ST398_80_iter_1.pdf

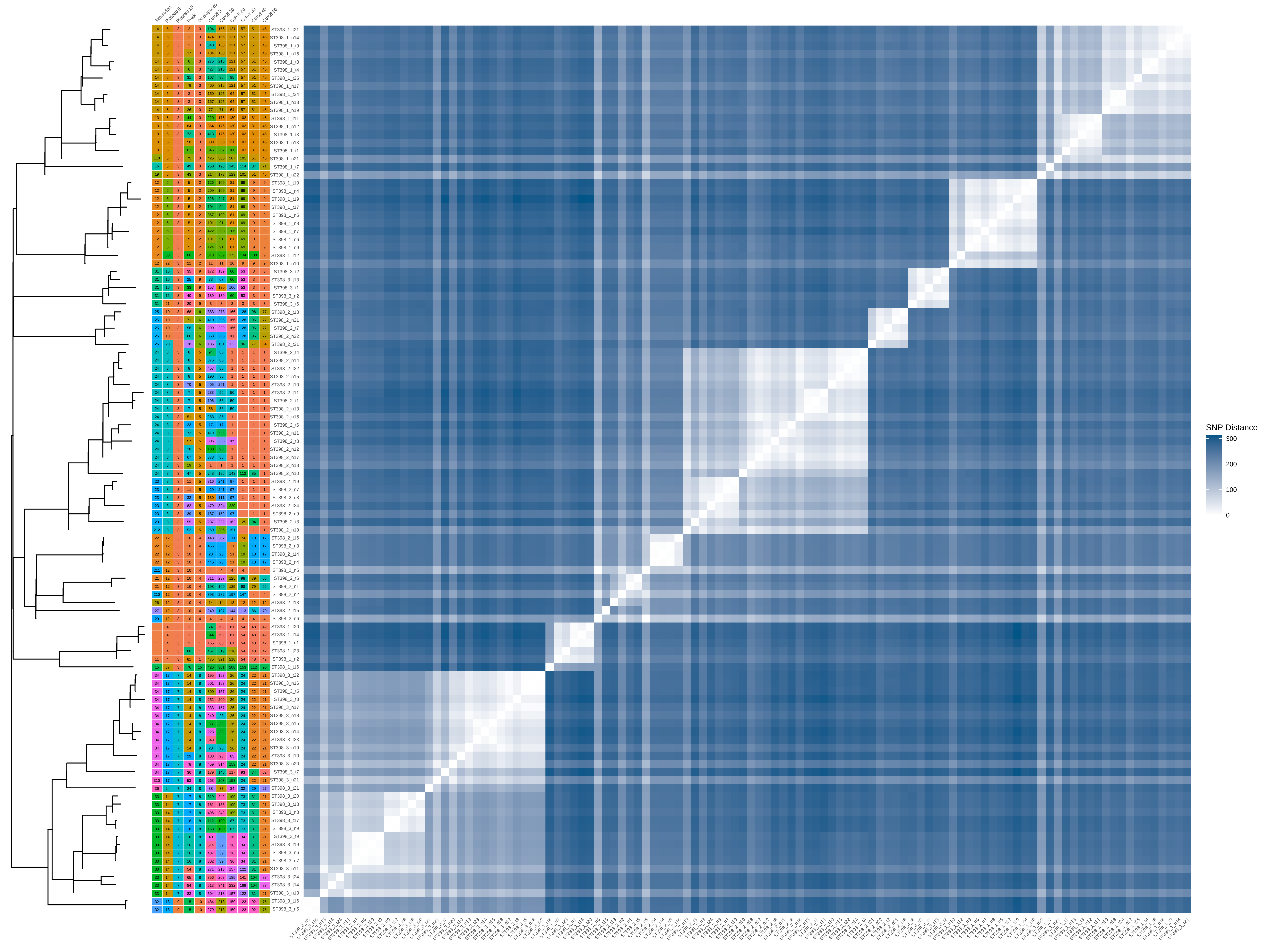

### ST398_80_iter_2.pdf

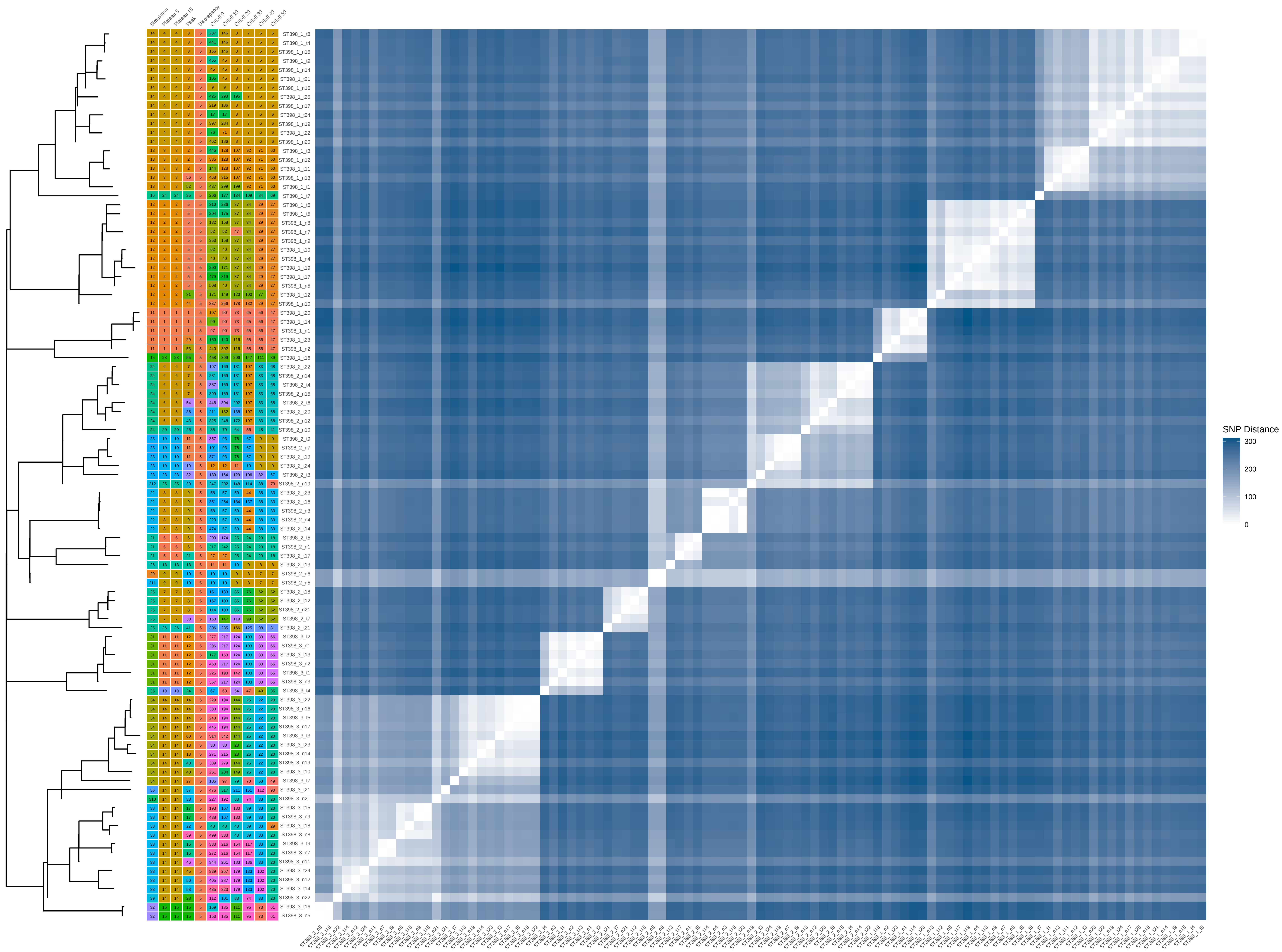

### ST398_80_iter_3.pdf

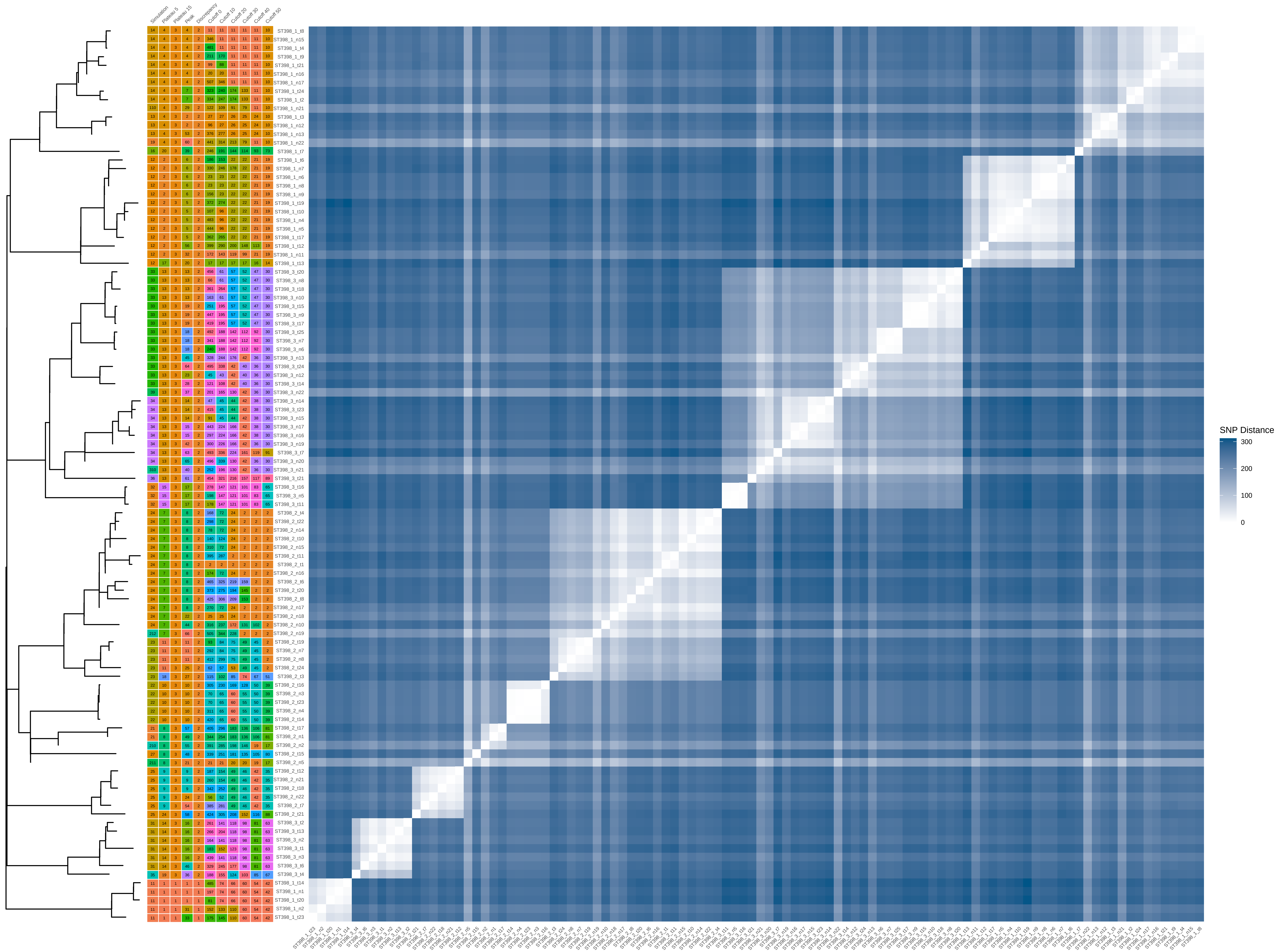

### ST398_80_iter_4.pdf

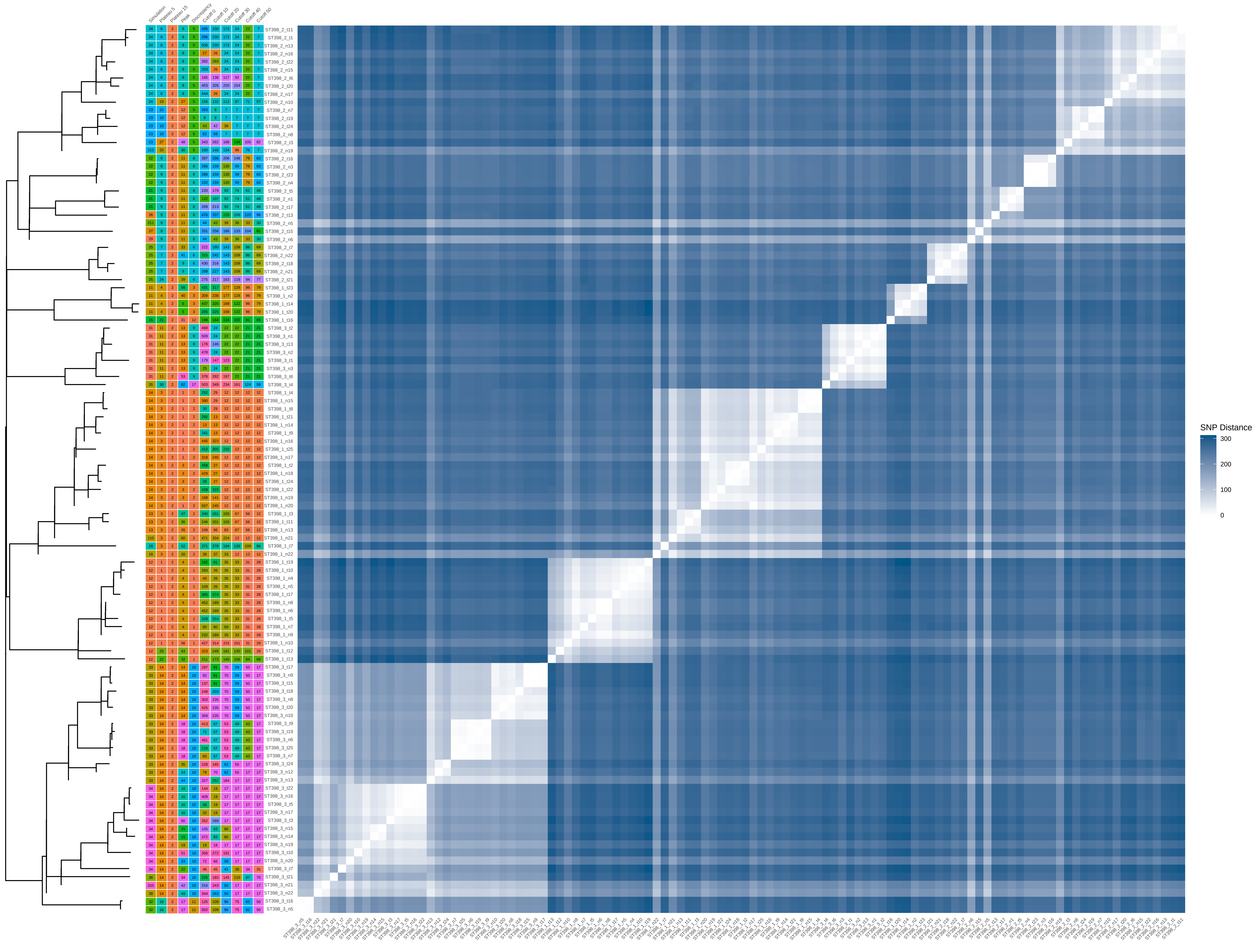

### ST398_80_iter_5.pdf

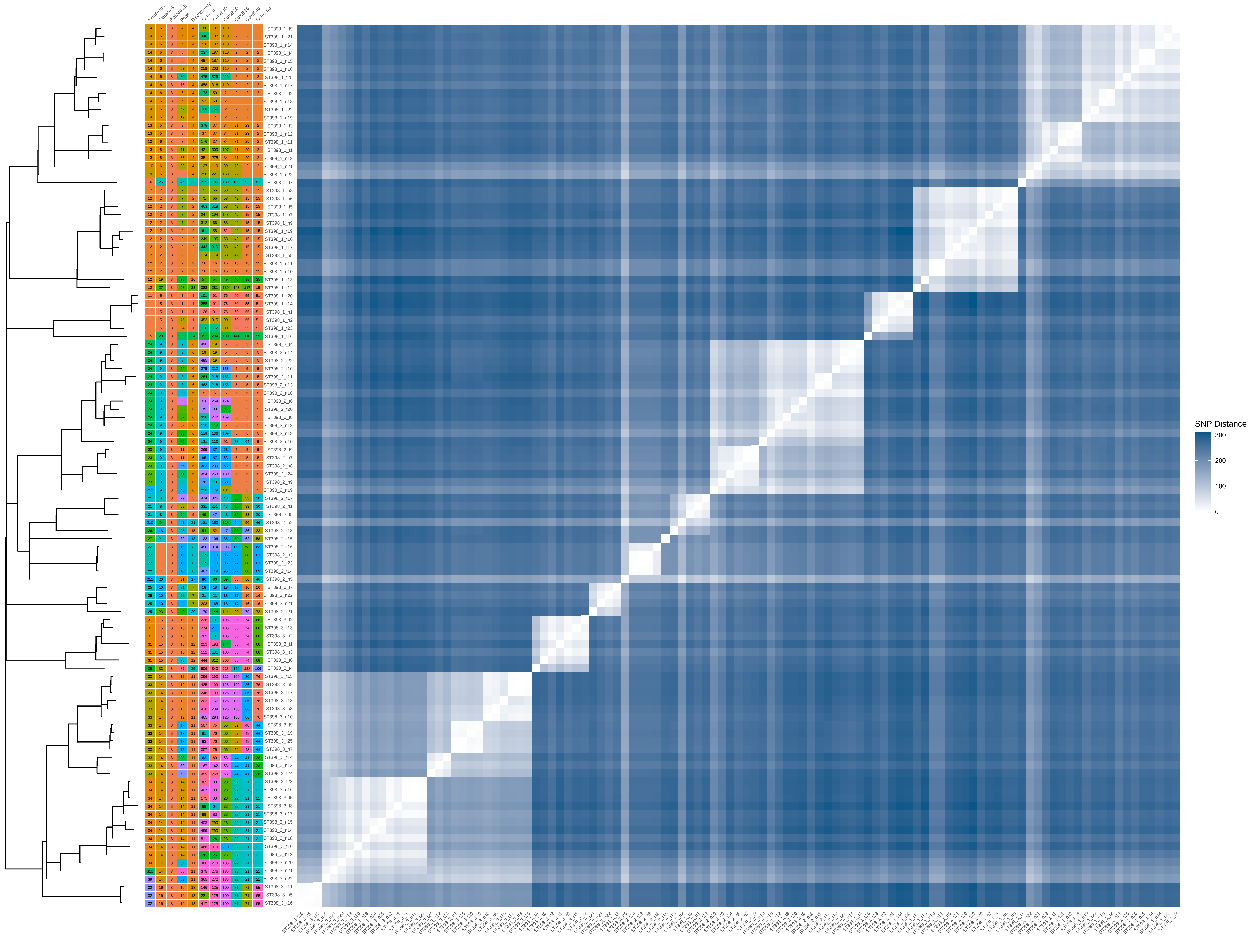

### ST398_80_iter_6.pdf

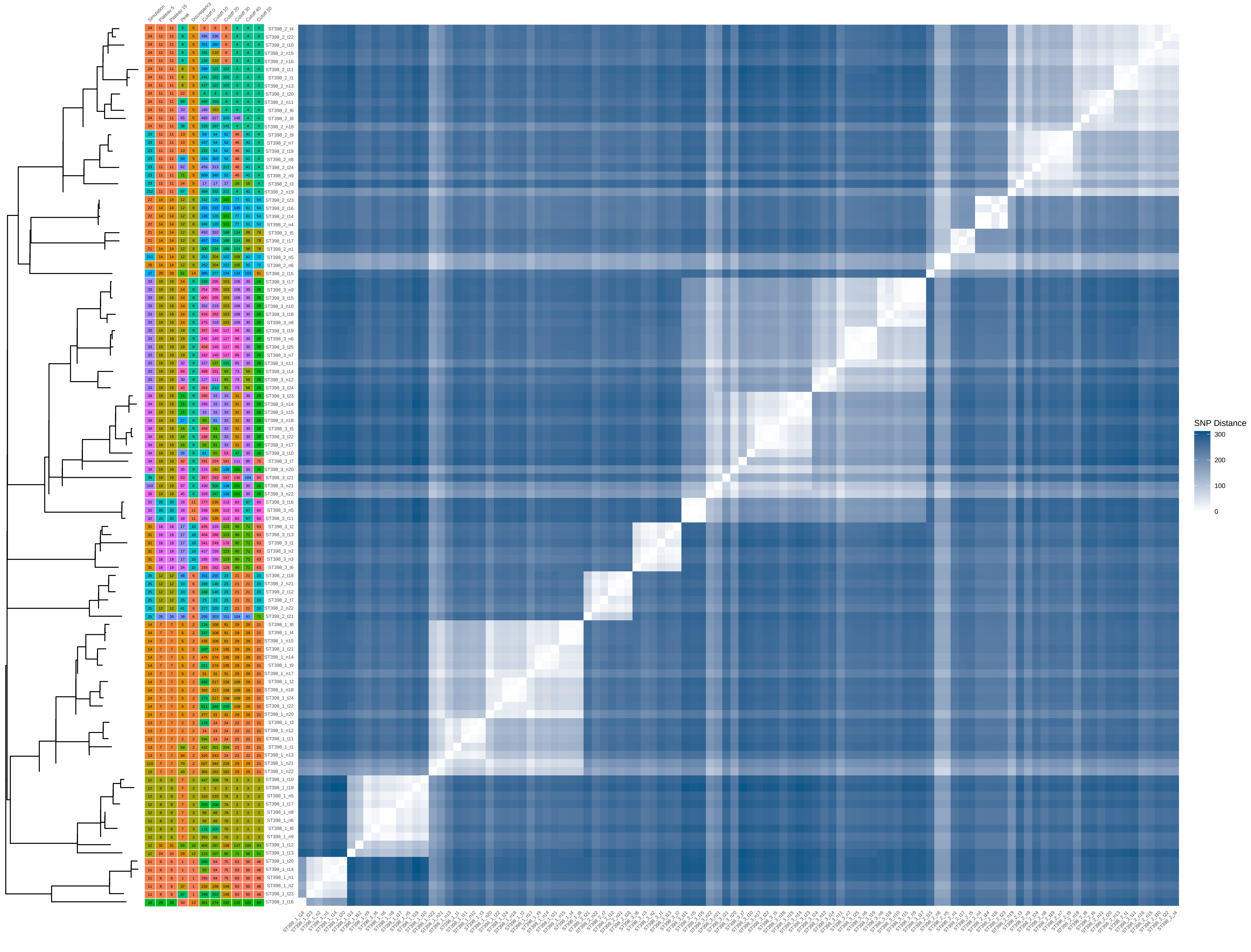

### ST398_80_iter_7.pdf

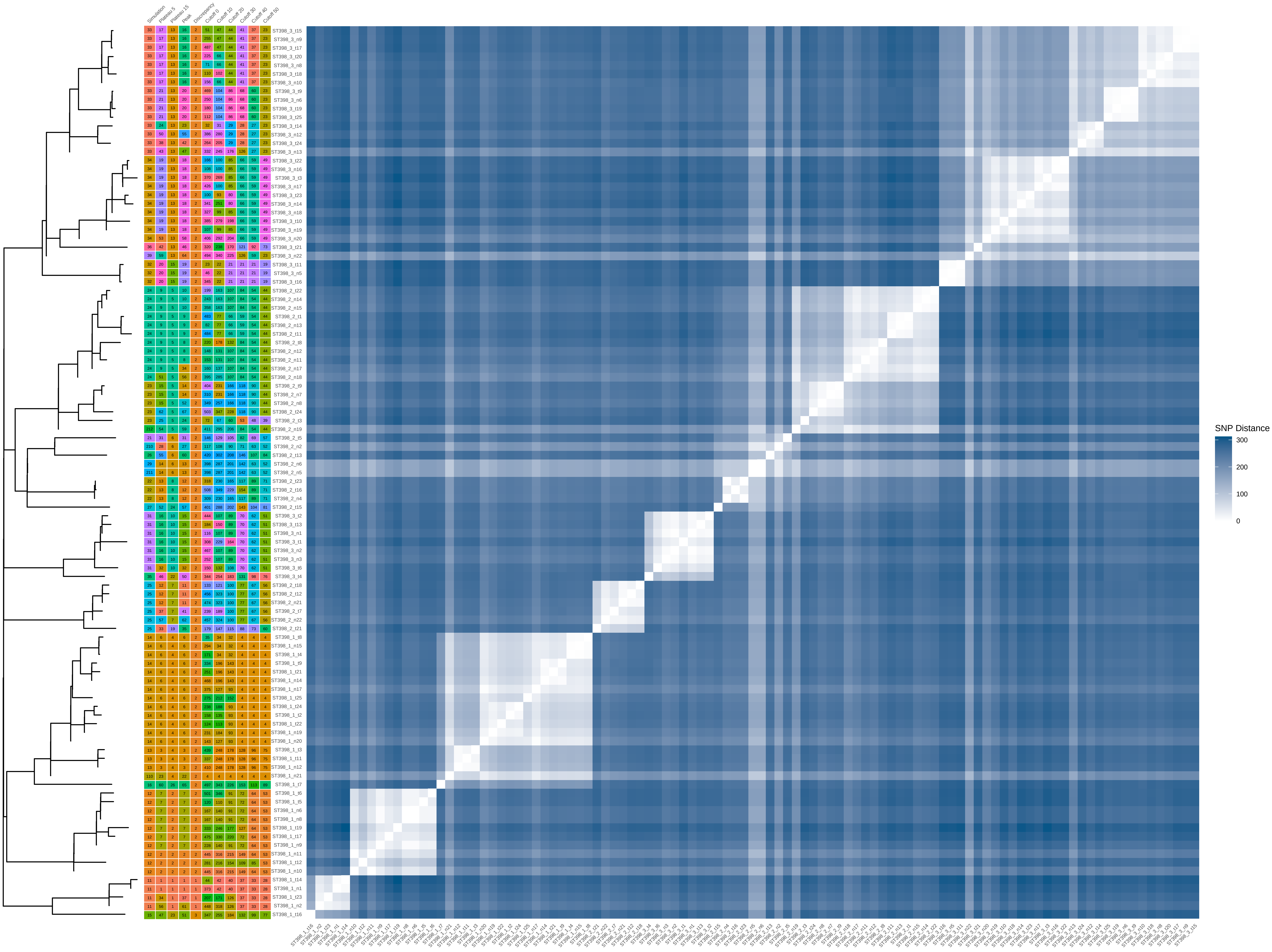

### ST398_80_iter_10.pdf

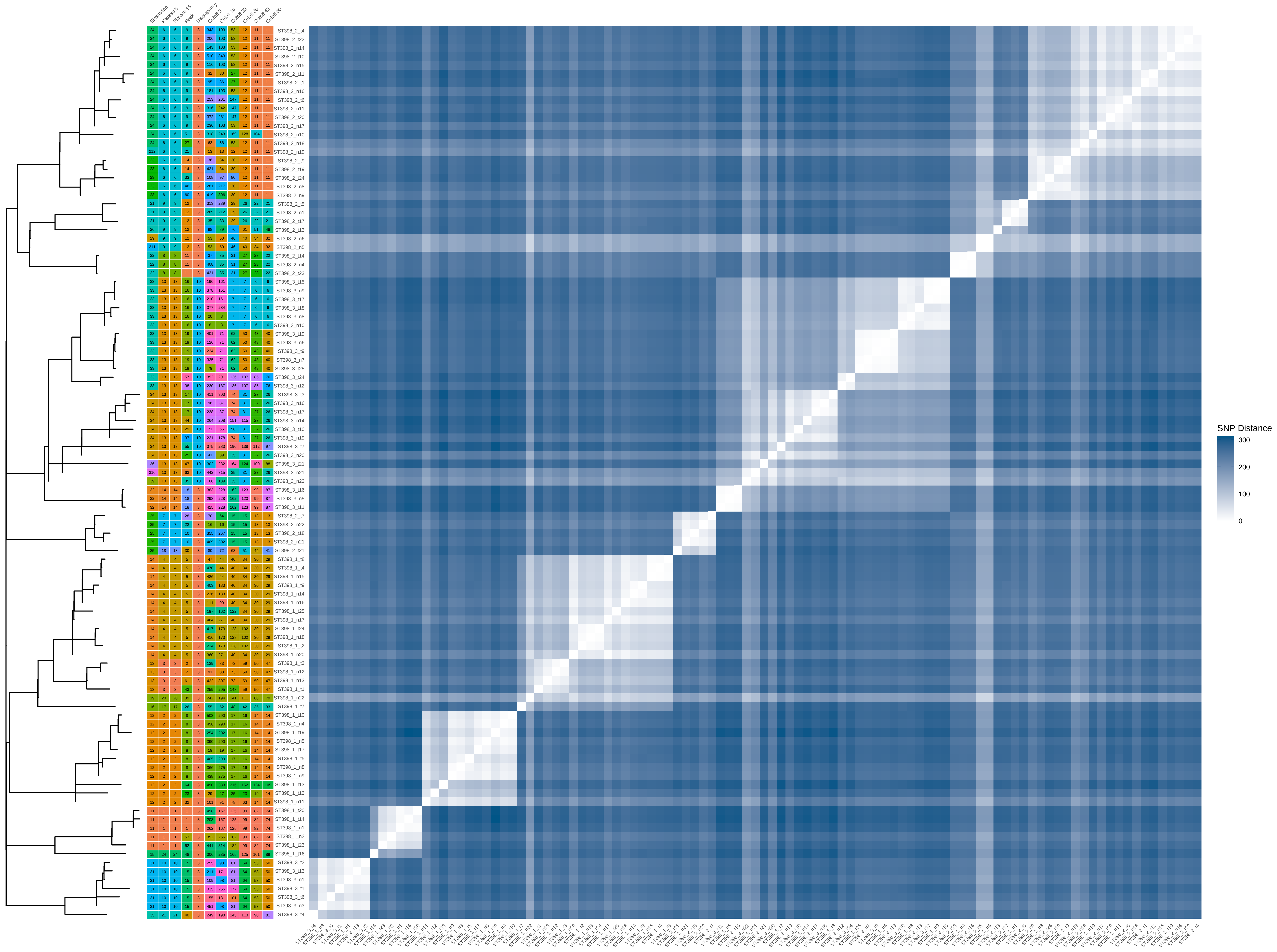

### ST398_100_iter_1.pdf

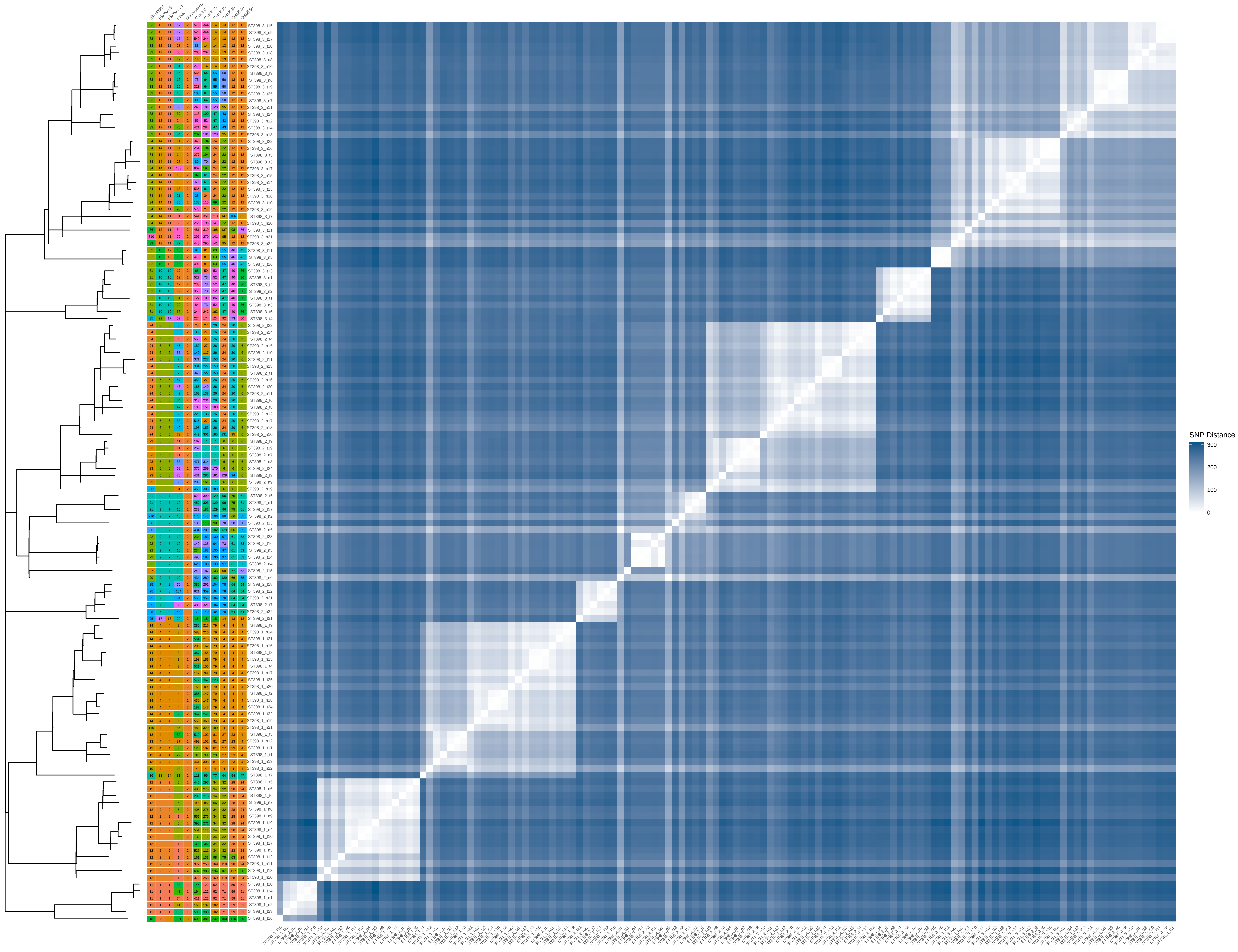

### ST398_100_iter_2.pdf

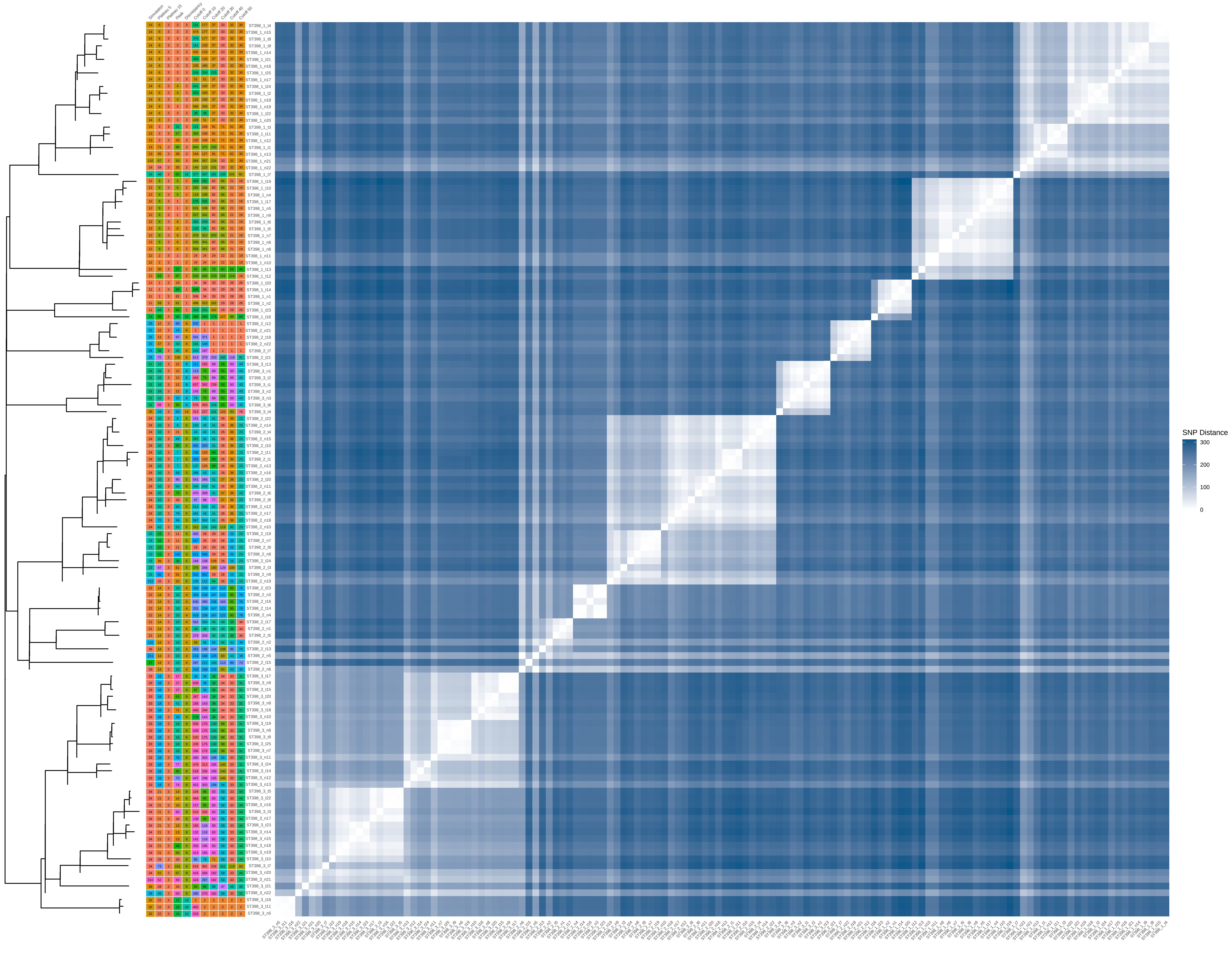

### ST398_100_iter_3.pdf

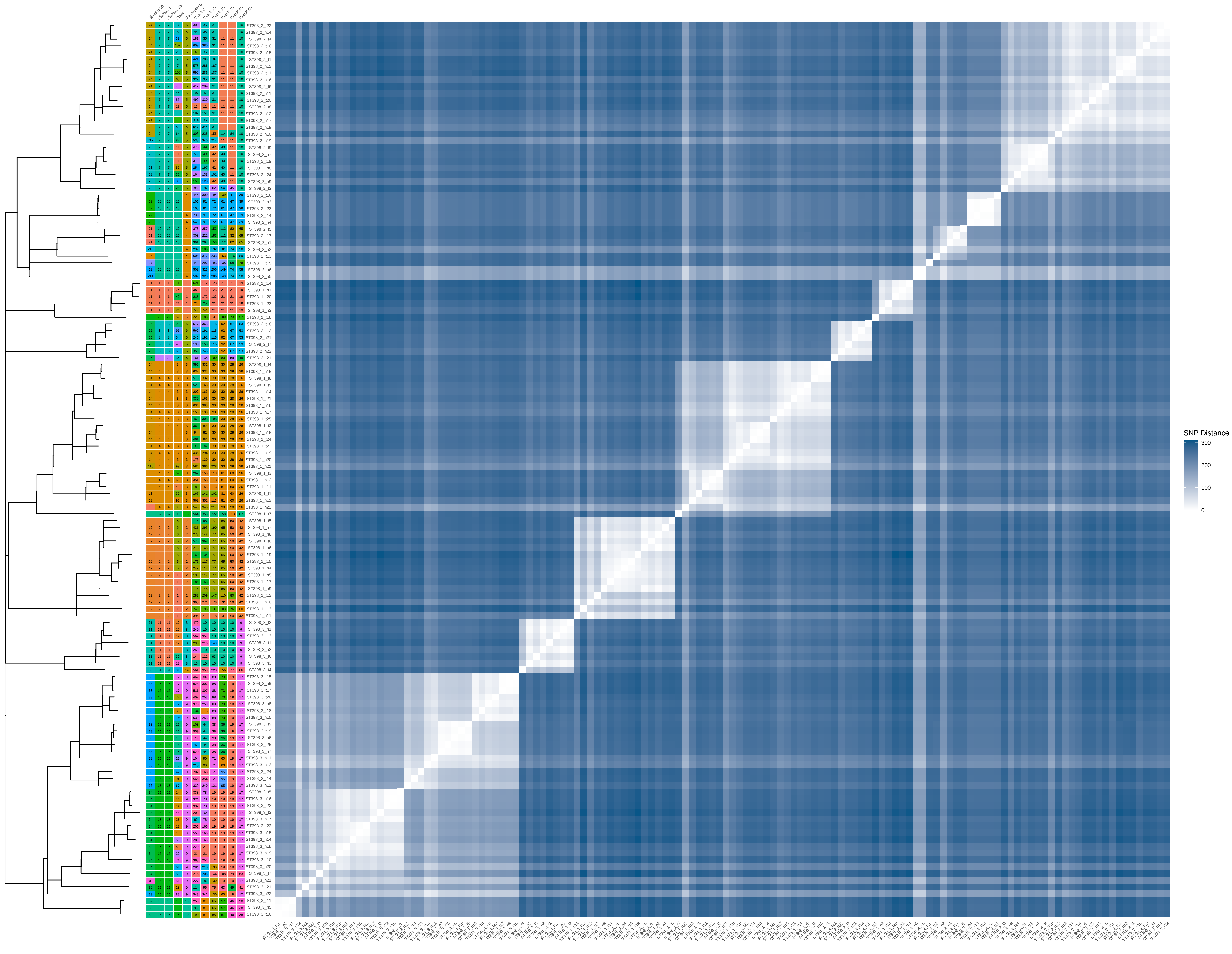

### ST398_100_iter_4.pdf

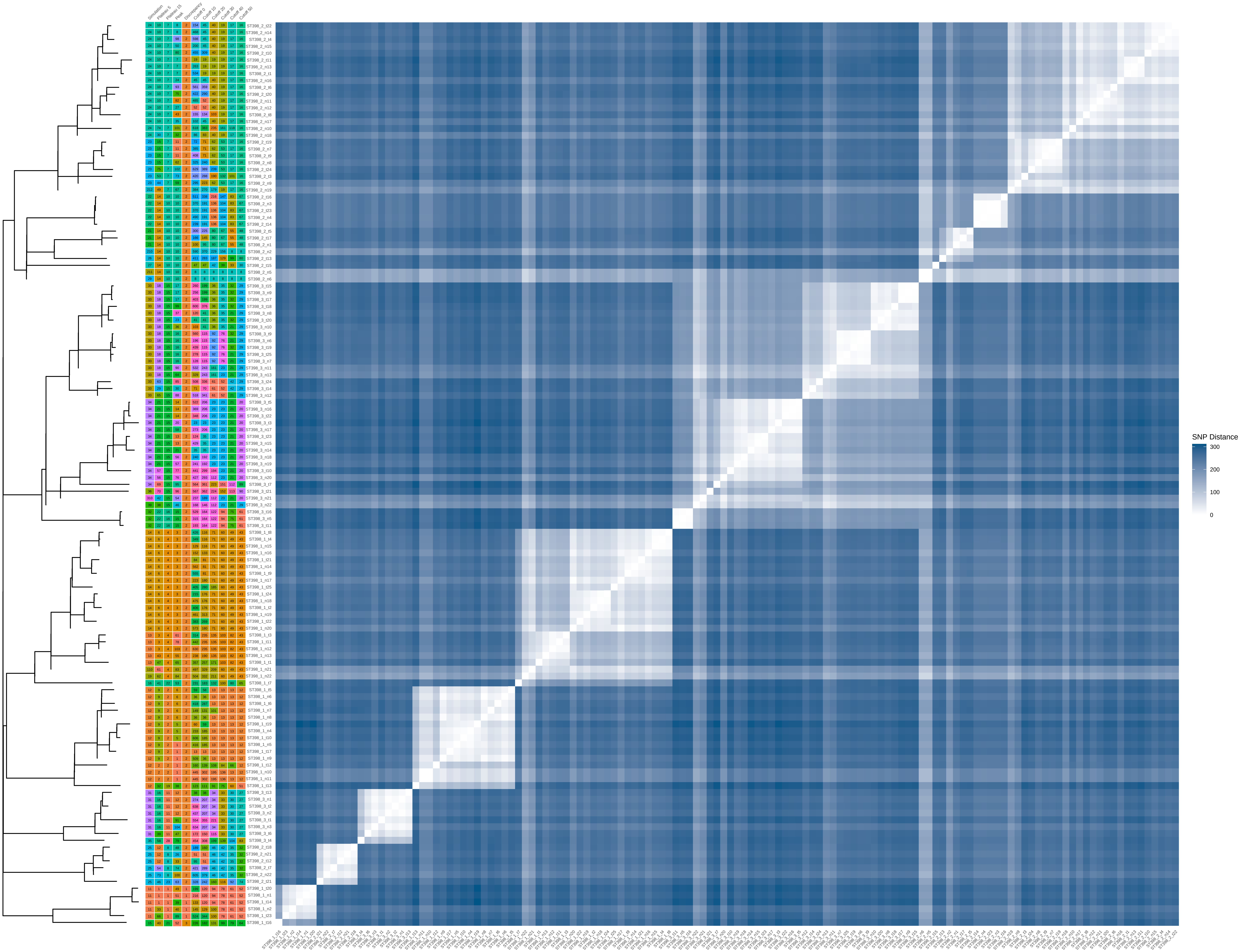

### ST398_100_iter_6.pdf

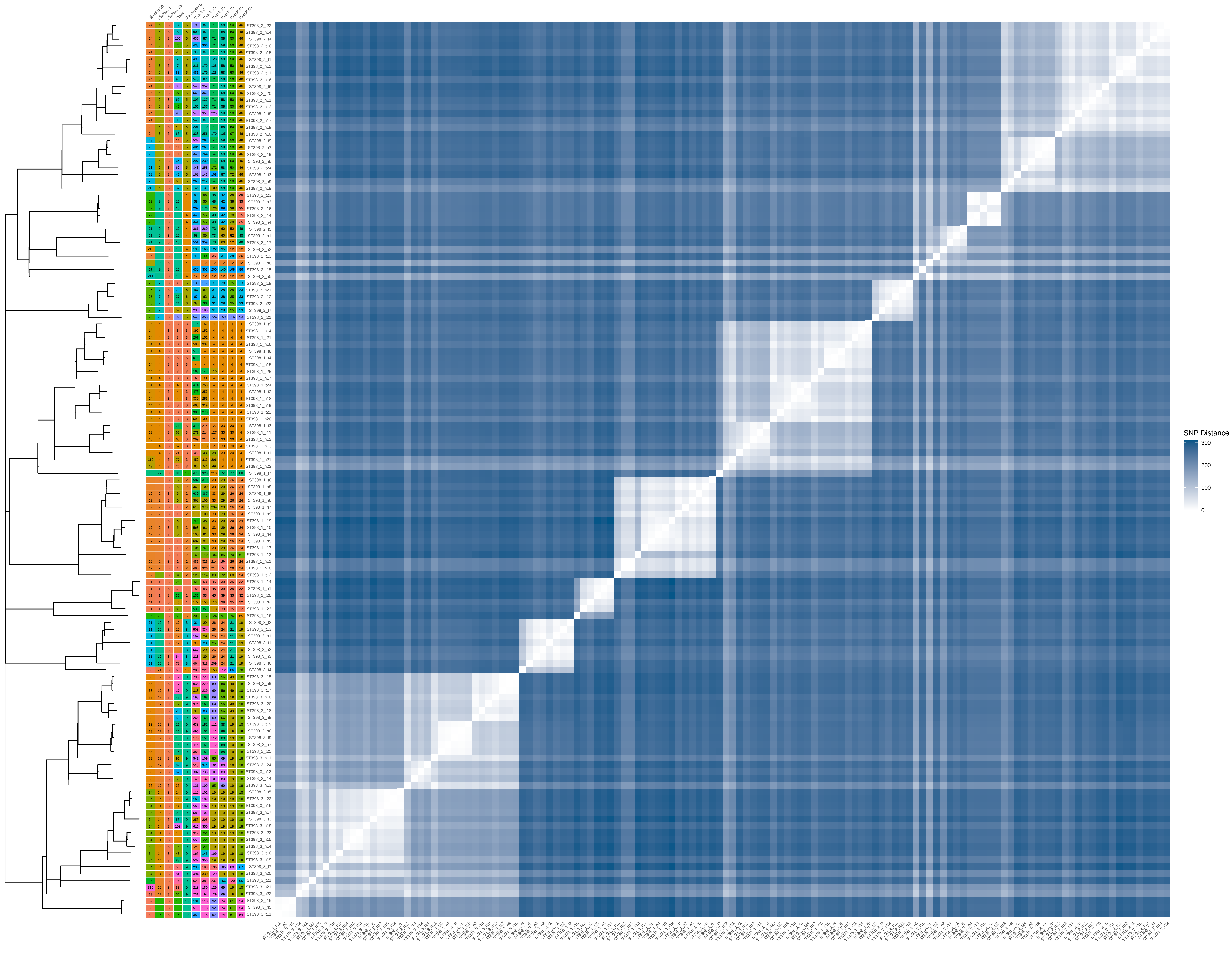

### ST398_100_iter_7.pdf

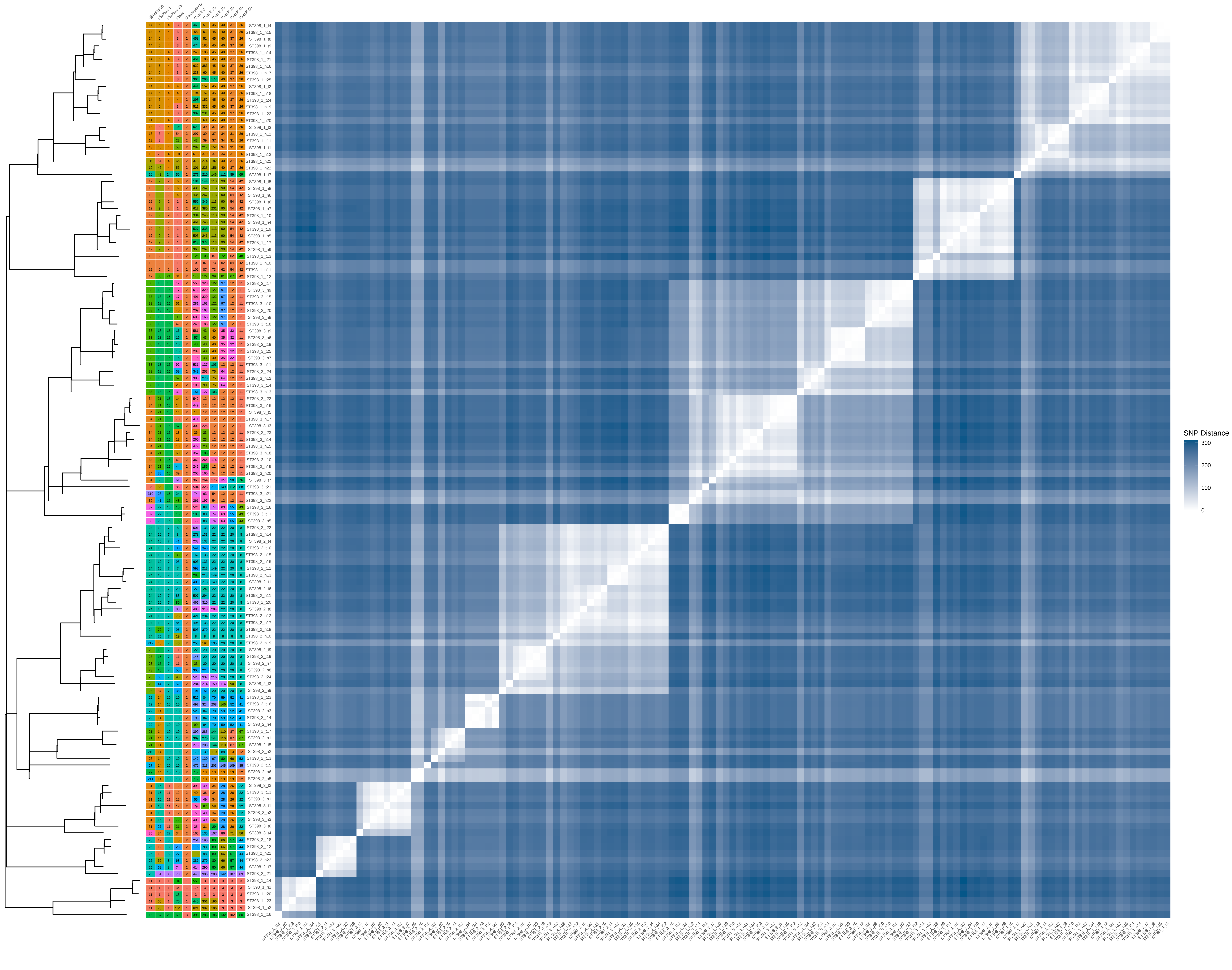
